# Implausible Effects of Psychological Interventions: Meta-Epidemiological Study and Development of a Simple Flagging Tool

**DOI:** 10.1101/2025.11.12.25340062

**Authors:** Mathias Harrer, Clara Miguel, Ian Hussey, Ioana A. Cristea, Wouter van Ballegooijen, Djordje Basic, Yingying Wang, Rory A. Pfund, Soledad Quero, Paula von Spreckelsen, Paula P. Schnurr, Annemieke van Straten, Toshi A. Furukawa, Davide Papola, Pim Cuijpers

## Abstract

In meta-analyses of psychological interventions, trials occasionally report effects that appear implausibly large. While such results are unlikely to reflect genuine treatment effects, this is rarely verifiable, and studies are often retained in the meta-analytic evidence. Existing guidance allows for highly questionable results to be discarded in evidence syntheses, but offers little direction on how to identify them in practice. Consequently, it remains unclear to what extent suspicious evidence has biased effect estimates in psychological intervention research. In this study, we develop a simple flagging tool to detect such trials, based on the (1) compatibility of their effect size with low risk of bias evidence, (2) achieved power, and (3) methodological rigor. We also examine specific characteristics of studies flagged by this tool, and the impact of their exclusion on pooled estimates and heterogeneity. In total, 2,881 effect sizes from 1,246 randomized trials were included from twelve living databases of psychological interventions for mental health problems. Overall, 5.3% of all effects (*n*=153 across 102 studies) were flagged. Reanalysis of 135 meta-analyses from a large-scale evaluation of psychological interventions showed that excluding flagged studies led to substantially lower effect estimates (reductions of up to 31.2%) and decreased between-study heterogeneity (up to 51.1%; indication-wide analyses). The flagging tool has been integrated into the open-source R package “metapsyTools”. We discuss potential explanations for the accumulation of improbable findings in the published literature, and how the application of our tool may strengthen quality control in meta-analytic research.

**Key Points:** *Question:* How can implausibly large effect sizes in psychological intervention trials be identified, and what impact do they have on meta-analytic evidence?

*Findings:* In this meta-epidemiological study, we developed a simple flagging tool based on effect size compatibility, statistical power, and methodological rigor. Applying it to 2,881 effect sizes from 12 living databases, we found that 5.3% of effects were flagged. Excluding flagged studies led to substantially lower pooled estimates (reductions of up to 31.2%) and decreased between-study heterogeneity (up to 51.1%).

*Meaning:* Implausible effects can substantially bias meta-analytic evidence. A simple flagging tool can help identify such effects and improve quality control in evidence syntheses. The tool has been integrated into an open-source R package for routine use.

## Introduction

Evidence syntheses remain a cornerstone of psychological research, particularly in evaluating psychological interventions, with hundreds of systematic reviews and meta-analyses published each year (Harrer, Miguel, van Ballegooijen, et al., 2025). Conclusions of these meta-analyses can strongly shape research priorities, guideline development, and professional practice. Consequently, inflated or biased effect estimates can have far-reaching and detrimental consequences for an entire research field.

A persistent challenge is that meta-analyses, by default, seek to include all eligible evidence on a research question; even though some of these studies may distort the result. Factors that lead to overestimated intervention effects have been extensively studied, from methodological flaws captured by risk-of-bias assessments (Miguel et al., 2025; Cuijpers et al., 2019), questionable research practices such as *P*-hacking, outcome switching, or HARKing (Andrade, 2021; John et al., 2012), to selective publication (van Aert et al., 2019). Paradoxically, the standard random-effects model, conventionally used to synthesize intervention effects, assigns greater weight to studies with inflated effect estimates than justified by their sample size alone. This can amplify upward biases and exaggerate between-study heterogeneity (Harrer et al., 2021, chap. 4.1; Lin et al., 2017). Conventional guidance, including the Cochrane handbook for health intervention trials (Higgins et al., 2024; chaps. 4.6.1, 8.2.1) and the Campbell Collaboration guidelines for social science reviews (Campbell Collaboration, 2020; chaps. 5.2.4.3 and 6.2) recommend addressing such concerns in sensitivity and ancillary analyses, but not by excluding studies from the primary synthesis based on the plausibility of the results.

Yet, there might be cases in which complete exclusion of a study from quantitative synthesis is indicated, even if it fulfills all eligibility criteria. Most obvious are studies that have been shown to be fabricated or fraudulent, although meta-analysts can usually only rely on published retraction notes and errata for this purpose. In recent years, there has also been greater attention on “zombie trials”: not necessarily proven fabrications, but studies so unreliable that their evidence becomes all but meaningless (Ioannidis, 2021); as well as shoddy research practices overall, even when there is no malicious intent (Clark et al., 2025). Tools that may help detect such suspicious trials are being tested (Mol et al., 2023; Wilkinson et al., 2024), but still require large investments of time and resources for each publication, especially since fabrication is often very difficult to “prove”.

This places meta-analysts in a difficult position: on one side, some eligible trials may report results that intuitively seem extreme, scarcely plausible, or outright suspicious; for instance, effect sizes far exceeding credible benchmarks (Hilgard, 2021). To illustrate, a trial might claim a between-group standardized mean difference (SMD) of 4, which is an effect comparable to individuals’ preference for chocolate over excrements (SMD=4.52; Balcetis & Dunning, 2010), and approximately ten times larger than the median across recent psychological research (Bogdan, 2025, Fig. 4b). Yet, the Metapsy living database on psychological treatments for depression (Cuijpers et al., 2025) alone includes nine randomized trials reporting such effects (SMD≥4). On the other side, methodological guidance holds that evidence syntheses should exclude eligible trials only in rare and exceptional cases (Higgins et al., 2024, chaps. 4.6.6, 4.6.1; Campbell Collaboration, 2020, chap. 6.2). The Cochrane handbook does state briefly that eligible trials may be excluded from meta-analyses if they are suspected to be “fraudulent”, or fatally flawed (chap. 4.6.6, chap. 8.2.1), but leaves open how this should be determined in practice.

Against this backdrop, we aim to provide psychological intervention researchers with a practical, empirically derived heuristic, which can be prespecified to exclude results too extreme and implausible to merit consideration in the meta-analysis – at least by default. As a simple flagging tool, this approach should not replace existing efforts to detect “zombie” trials or actual fraud, but serve as a method to discard results that clearly strain credulity from the main analysis, trigger further inspection of the trial, or compare newly published trials against empirical benchmarks. To be practical, it is strongly desirable that these flags can be computed using only information routinely included in meta-analytic databases anyway, without requiring additional data extraction steps. This may be particularly relevant for large-scale “living” databases in the field (Cuijpers et al., 2022), which can often include hundreds of trials.

The goal of this study is to develop and validate such a flagging tool for psychological intervention studies, combining information on (i) the compatibility of an effect with existing low-risk of bias evidence across psychological treatment research; (ii) the achieved power, using realistic reference values of the true effect size; and (iii) the overall methodological rigor of the study. Application of (i) and (ii) requires sensible thresholds to determine “extreme” values, which we derive from the Metapsy meta-analytic research domain for psychological treatments (MARD; Harrer, Miguel, van Ballegooijen, et al., 2025). Finally, we perform a meta-epidemiological analysis to characterize the studies flagged by our tool and assess how their exclusion changes previous estimates of psychological intervention effects and their heterogeneity.

## Methods

### Datasets

All development and validation data for this study were obtained from the “Metapsy” meta-analytic research domain (MARD) on psychological interventions (metapsy.org; Cuijpers et al., 2022; Harrer, Miguel, van Ballegooijen, et al., 2025). The Metapsy MARD provides comprehensive living databases of randomized trials across a wide range of indications and treatments. These databases are harmonized through a unified protocol (Harrer, Miguel, van Ballegooijen, et al., 2025) and have served as the basis for more than 100 published meta-analyses over the past 15 years (Cuijpers et al., 2023; see metapsy.org/published-articles for an overview). Detailed information on the search strategies, data extraction, and coding procedures for each database is available on its designated documentation webpage (docs.metapsy.org/databases).

For this study, effects of psychological interventions were included from twelve different databases, each focusing on a specific mental health problem: unipolar depression, panic disorder, social anxiety disorder (SAD), generalized anxiety disorder (GAD), specific phobias, posttraumatic stress disorder (PTSD), obsessive-compulsive disorder (OCD), borderline personality disorder (BPD), prolonged grief disorder, problem gambling, psychotic disorders (including schizophrenia/psychosis, schizophreniform disorder, schizoaffective disorder, delusional disorder), and suicidality. Methodological details for all of these databases, including the search strategy, are provided in the respective documentation entries (Basic, 2025; Cuijpers et al., 2025; de Ponti et al., 2025; National Center for PTSD, US Department of Veterans Affairs, 2025; Papola, 2025a, 2025b; Pfund, 2025; Quero et al., 2025; Setkowski et al., 2025; van Ballegooijen et al., 2025; Wang et al., 2025). Updated Preferred Reporting Items for Systematic reviews and Meta-Analyses (PRISMA; Page et al., 2021)-type study search and inclusion flowcharts are provided in Supplement S1.

For this study, we only selected databases in which psychological interventions are compared to treatment-as-usual (TAU), waitlists, or other inactive controls; existing Metapsy databases on head-to-head comparisons between two “bona fide” interventions (e.g., pharmacotherapy or other psychological treatment) were not considered. For the depression, SAD, GAD, panic disorder, specific phobias, PTSD, and problem gambling databases, the last available database update was January 1st, 2025. A more recent update was available for psychosis (March 1st, 2025), while only older updates could be used for OCD (September 1st, 2024), suicidal ideation (April 1st, 2023), prolonged grief, and borderline personality disorder (both January 1st, 2023). In all databases, risk of bias was assessed using Cochrane’s Risk of Bias (RoB) tool (Sterne et al., 2019) with two independent raters. For all databases except depression, the updated version (RoB-2) was applied. For the depression database, complete ratings were only available for the original RoB tool, with RoB-2 re-ratings currently ongoing (Miguel et al., 2025).

In all databases, small-sample bias-corrected standardized mean differences (SMD; Hedges’ g) and their sampling variances are calculated using the “metapsy-Tools” package (Harrer et al., 2022). Endpoint means and standard deviations are prioritized when calculating these outcomes measures (Harrer, Miguel, Luo, et al., 2025). If these data are unavailable, change scores are extracted instead, followed by response rates (converted to SMDs under the assumption of a logistic distribution; Chinn, 2000), and finally any other post-test statistic that can be transformed into an effect size (e.g., t or χ^2^ values, or regression coefficients).

### Development of the Flagging Tool

As part of this study, we aimed to devise a flagging tool suitable to identify extreme and/or barely credible results of psychological intervention trials, applicable using information routinely collected in meta-analytic databases. To this end, three risk indicators were considered: the (1) compatibility of an effect with existing low-risk-of-bias evidence across psychological treatment research; (2) achieved power, using realistic reference values of the true effect size; and (3) overall methodological rigor. These indicators were combined to ensure that evidence was not flagged solely because of unusually large effects (criterion 1). Instead, studies would only be discarded when meeting all three criteria. This would allow potential break-through findings to remain credible, provided they were supported by either sufficient power (criterion 2) or rigorous methodology (criterion 3).

Application of criteria (1) and (2) requires sensible empirical thresholds to determine “extreme” values, which first had to be derived from the collected database. To obtain a value beyond which the size of an effect would be considered “extreme” (i.e., incompatible with existing low-risk of bias evidence; criterion 1), a set of plausible parametric distributions (Pareto, exponential, Gamma, log-normal, Weibull, logistic, and Cauchy) was fitted to the empirical distribution of effect sizes in our dataset. The Pareto, exponential, Gamma, log-normal and Weibull distributions were selected because a right-skewed distribution was deemed plausible, with most effect sizes concentrated at the lower end of the distribution. The Cauchy distribution was selected due to its heavy-tailedness, which was hypothesized to be helpful in capturing extreme effects. Only effect sizes from studies with a low risk of bias rating were considered for this step. For each distribution, parameter values were optimized using maximum likelihood estimation, with the best-fitting distribution determined using the BIC and Anderson-Darling statistic. The Anderson-Darling statistic was prioritized over the Kolmogorov-Smirnov goodness of fit because it gives greater weight to the tails of a distribution (Ćmiel et al., 2020). Since the support for some of the candidate distributions is ℝ, absolute effect size values with a shift of 1×10^−5^ were used during the estimation step. To fulfill the i.i.d. assumption, multiple effects were pre-aggregated on a study level, assuming a constant sampling correlation of ρ=0.6.

The best-fitting distribution was then selected to determine an effect size cut-point indicating strong incompatibility with existing low-risk of bias evidence. As a measure of incompatibility, the chosen distributional form was used to derive the critical effect size θ that results in a Shannon information (S-value; Greenland, 2019) of 4.32:

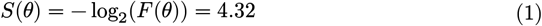

where F(θ) is the cumulative distribution function of the selected distribution. This threshold was considered appealing because the S-value expresses how “surprising” an observed effect size is relative to high-quality evidence, measured in bits of information. An S-value of 4.32 corresponds to the improbability of flipping four coins and obtaining heads each time, and coincides with the conventional significance level of p = 0.05. Where sufficient low–risk-of-bias evidence was available (≥20 effect sizes for a given indication), indication-specific thresholds were also estimated. This was the case for depression, PTSD, and psychosis trials.

As a second step, a risk indicator for lack of statistical power was derived (criterion 2). Once again, only low-risk of bias evidence was considered at this stage. We first estimated a meta-analytic reference effect size δ using an unrestricted weighted least squares (UWLS) estimator. Given α=0.05 and the desired power 1 − β, a trial with sampling variance v_i was then considered sufficiently powered if:

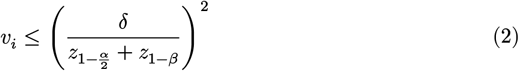

This formula was used to compute the achieved power of each trial. In line with prior work (de Vries et al., 2023), we expected most psychological intervention trials to be underpowered, and that the conventional threshold of 1 − β = 0.8 would result in nearly all trials being flagged. To improve specificity, we instead defined “severely underpowered” studies as those falling into the lowest empirical quartile of achieved power among low-risk of bias trials. It is important to note that these measures of achieved power were not intended to reproduce the a priori power calculations of individual trials, but rather to reflect how well a given study was powered to detect the meta-analytic reference effect size δ. Indication-specific reference values were also computed for depression, psychosis, and PTSD trials, given that sufficient study material was available (k ≥ 20).

For the third indicator (methodological rigor), risk of bias ratings available in the database were used. Trials not judged to be at “low risk” according to Cochrane’s RoB tool were flagged, with no further calculations required. Based on the empirically derived critical values, all effect sizes included in the dataset were subsequently rated on each of the three criteria (extreme effect size, severe lack of power, and risk of bias), and results were summed to yield a risk score ranging from 0 to 3. As part of the tool, we consider effect sizes to be “flagged” when all three risk indicators are present (equaling a score of 3).

### Validation of the Tool

To gauge the impact of the flagging tool on meta-analytic effect estimates and their heterogeneity, we first calculated a three-level correlated and hierarchical (CHE; Pustejovsky & Tipton, 2022) pooling model with cluster-robust variance estimation (CRVE) when using all available effect sizes in the database, and when excluding effects with one, two, or all three positive risk indicators. Studies including “flagged” effect sizes (defined as scoring “yes” on all 3 indicators) were also analyzed descriptively, and we examined potential differences to unflagged trials in terms of country of origin, world bank income region, sample size, indication, or publication year. As a final step, we assessed the “real-life” impact of applying the tool in practice. This was done by reanalyzing the results of Harrer et al. (2025), a large-scale meta-analysis across the same 12 indications as covered in the development set of the present study. For each of the indication-specific analyses, results were recalculated when excluding effects with one, two, or all three risk indicators, while recording percent changes in the available number of trials/effect sizes, pooled effect, and between-study heterogeneity variance τ^2^ compared to the “naïve” analysis. Additionally, we also examined changes when excluding each specific flag individually (extreme effect, strong lack of power, or risk of bias), and when repeating all these analyses in subgroups defined by the treatment format and comparator (e.g., cognitive-behavioral therapy versus waitlist, interpersonal therapy versus care as usual, etc.). As in the original investigation, CHE models with CRVE were used throughout, assuming a constant sampling correlation of ρ=0.6. Profile likelihood plots were inspected to ensure identifiability of the heterogeneity variance components.

### Reproducibility & Open-Source Software

A preregistered protocol for this study was made available via the Open Science Framework (osf.io/2bgsa). All analysis code and materials are openly accessible (Harrer, 2025), together with documentation on how to adapt the flagging tool to other databases or research fields. The flagging tool itself has been implemented in open-source software as part of the “metapsyTools” package (Harrer et al., 2022). Using the “flagEffectSizes” function (tools.metapsy.org/reference/flageffectsizes), users can select an appropriate reference distribution (e.g., overall, depression, PTSD, or psychosis trials), provide standardized mean differences and standard errors, and then automatically apply the flagging tool with corresponding diagnostics. If supplied to the main meta-analysis function of the package, flagged effect sizes (i.e., scoring positive on all three risk indicators) will be automatically discarded from the pooling models.

## Results

In total, 2,881 effect sizes from *K*=1,246 randomized trials were available for the present study.

References for these trials are provided in Supplement S2. A descriptive summary of the included studies can be found in Supplement S3. Most trials (*k*=504; 40.4%) focused on depression, followed by psychosis (*k*=175; 14%) and PTSD (*k*=102; 8.2%); while only *k*=27 trials (2.1%) were available for BPD. Across indications, trial sample sizes ranged between 8 and 1,140, with a median of 57. A substantial share of trials (40.1%; *k*=500) employed waitlist controls, and 22.6% (*k*=282) were judged to show low risk of bias.

### Characteristics of the Flagging Tool

In Supplement S4, we show an overlay between the empirical distribution of effect sizes in this study and the fitted distributions, along with their cumulative density functions. Based on the BIC and Anderson-Darling statistic, Gamma (*α*=1.227, *λ*=1.828) emerged as the most suitable approximation. Based on this distribution, SMD=1.87 serves as the critical threshold corresponding to an *S*-value of 4.32. Thus, all effect sizes beyond this value are deemed too extreme to be compatible with current low-risk of bias evidence. We also adapted these thresholds specifically for interventions in depression (*α*=1.495, *λ*=2.343; SMD=1.66), psychosis (*α*=1.249, *λ*=2.788; SMD=1.24), and PTSD (*α*=2.696, *λ*=2.026; SMD=2.88). Supplement S5 shows the estimated achieved power distribution across all effect sizes from low-risk of bias trials, assuming a target effect size of δ=0.46 (pooled UWLS estimate overall) as the reference. To detect δ with sufficient 80% power in an unadjusted *T*-test, a total of 152 individuals need to be randomized (assuming equal allocation and no dropout). As expected, most trials fell far below this threshold, with the first, second, and third quartile of achieved power at 32.4%, 52.6%, and 73.8%. Thus, as prespecified, 32.4% (the first quartile) was used as the threshold for a critically underpowered trial. Indication-specific critical values for severe lack of power were 41% for depression, 21.8% for psychosis, and 74.8% for PTSD.

### Characteristics of Flagged Trials

Figure 1 presents a funnel plot of all effect sizes in the database, and when excluding effects with all three, two, or just one risk indicator. Using all 2,881 available effect sizes, the pooled effect estimated using a random-effects CHE model was SMD=0.69 (95% CI: 0.66 to 0.73), and between-study heterogeneity estimated at τ^2^=0.306. Overall, 153 (5.3%) effect sizes were flagged (positive screen on all three indicators). When these effects were removed, the overall treatment effect decreased slightly (SMD=0.60), while between-study heterogeneity was almost halved (τ^2^=0.306→0.151). This pattern became even more pronounced when additionally excluding 998 effects with at least two positive indicators (SMD=0.51; τ^2^=0.112), and yet another 1,284 effects with at least one indicator (SMD=0.47; τ^2^=0.056). Visually, the plots also show a strong reduction in overall funnel plot asymmetry, with the exclusion of three-indicator effects (upper-right plot) having the greatest impact. In line with a reduction of small-study effects, standard errors became increasingly less predictive of the SMD, from β=3.910 (all effects), to 2.017 (3 indicators removed), 1.880 (at least two indicators), and 1.731 (at least one indicator).

**Figure 1:**
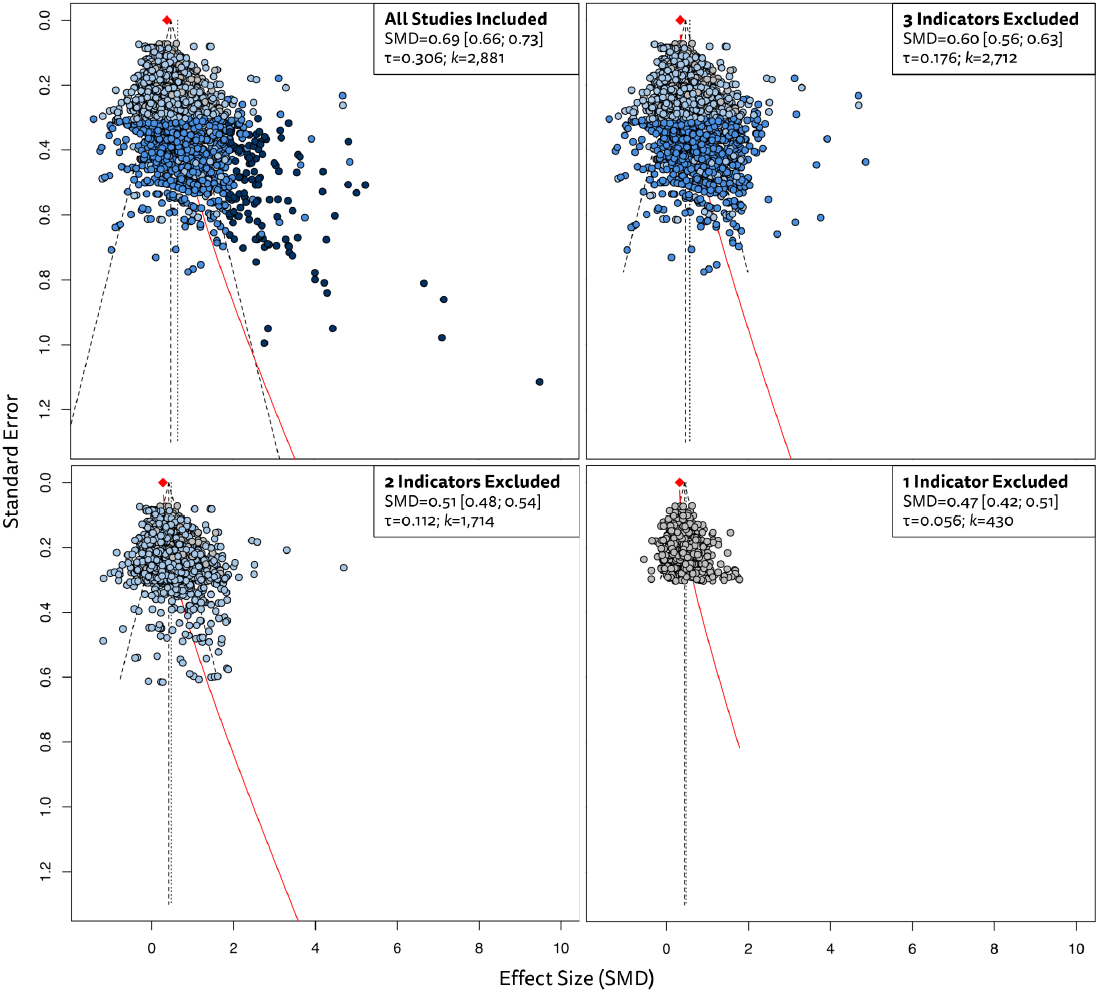
Funnel plots of all effect sizes in the database, and when excluding effects with all three, two, or just one risk indicator.

Characteristics of the 102 studies with flagged effect sizes (three positive indicators) are summarized in Supplement S9. Overall, reported effects in these studies ranged from SMD=1.88 to 15.41, with a median of 2.55. Flagged trials originated from all major world regions, with the United States (*k*=27; 26.47%), Iran (*k*=11; 10.8%), and Canada (*k*=9; 8.82%) leading the list. Overall, 68.6% (*k*=70) of all flagged trials came from high-income countries. This pattern reversed when we only considered trials published in the last ten years (2015-2025), with two-thirds (66.7%; *k*=26) of all flagged trials being conducted in low and middle-income countries (LMICs). Among these more recent flagged trials, most were published in Iran (*k*=9; 23.1%), followed by Nigeria (*k*=4; 10.2%). We did not find a difference in the median publication year between flagged and non-flagged trials (2014 for both). However, waitlists controls (70.6%; *k*=70) were overrepresented among flagged trials. In Supplement S10, we also show the share of newly published flagged and non-flagged trials over time (1973 to 2025). Poisson models indicated that, while the number of both unflagged and flagged trials increased yearly, this growth rate was smaller for flagged trials (β=-0.531; S.E. 0.153). This means that the predicted share of flagged trials has decreased over time.

### Reanalysis of Meta-Analytic Intervention Effects

Results of reanalyzing a recent meta-analysis across twelve mental health problems (Harrer et al., 2025; *K*=1,029) are presented in Table 1. Overall, excluding flagged effect sizes (3 risk indicators) led to modest reductions in the available evidence base, with a median of 7.1% of effects being discarded (range 1.1% to 18%). Much more evidence was excluded when removing effects with at least one or two risk indicators (range: 28.1% to 93.2%). For suicidality, SAD, specific phobias, OCD, and borderline personality disorder, not enough effect sizes (*k*≥3) were available for meta-analytic pooling after all comparisons with at least one flag were removed. Across all indications, removing flagged studies led to a considerable reduction in the pooled SMD (6.3% to 31.2%), and even greater decreases in the estimated between-study heterogeneity (6.8% to 51.1%). Similar patterns were also observed when excluding effects with at least one or two risk indicators, and when excluding specific indicators (extreme effect, strong lack of power, lack of methodological rigor) individually (see supplement S6).

**Table 1.** Changes in pooled effects and heterogeneity of psychological treatment after excluding flagged studies (reanalysis of Harrer et al., 2025).

| Condition/Analysis | $n_{\text{eff}}$ | $\Delta_n$ | SMD | 95%-CI | $\Delta_{\text{SMD}}$ | $\tau^2$ | $\Delta_{\tau^2}$ | 95%-PI | NNT |
| --- | --- | --- | --- | --- | --- | --- | --- | --- | --- |
| **Depressive Disorders** |  |  |  |  |  |  |  |  |  |
| All Effect Sizes Included | 1,246 | ref. | 0.73 | [0.67; 0.79] | ref. | 0.3917 | ref. | [-0.50; 1.96] | 4.04 |
| 3 Indicators Removed<sup>a</sup> | 1,163 | 6.7% | 0.62 | [0.58; 0.66] | 15.1% | 0.1853 | 31.2% | [-0.23; 1.46] | 4.92 |
| At Least 2 Indicators Removed | 662 | 46.9% | 0.53 | [0.49; 0.58] | 27.4% | 0.1517 | 37.8% | [-0.23; 1.30] | 5.82 |
| At Least 1 Indicator Removed | 268 | 78.5% | 0.45 | [0.40; 0.50] | 38.4% | 0.0614 | 60.4% | [-0.04; 0.94] | 7.12 |
| **Posttraumatic Stress Disorder** |  |  |  |  |  |  |  |  |  |
| All Effect Sizes Included | 131 | ref. | 1.18 | [1.01; 1.35] | ref. | 0.6177 | ref. | [-0.39; 2.74] | 2.79 |
| 3 Indicators Removed<sup>a</sup> | 124 | 5.3% | 1.06 | [0.92; 1.20] | 10.2% | 0.3761 | 22.0% | [-0.16; 2.28] | 3.21 |
| At Least 2 Indicators Removed | 90 | 31.3% | 1.03 | [0.86; 1.19] | 12.7% | 0.3918 | 20.4% | [-0.23; 2.28] | 3.35 |
| At Least 1 Indicator Removed | 23 | 82.4% | 1.00 | [0.69; 1.31] | 15.3% | 0.3487 | 24.9% | [-0.26; 2.26] | 3.45 |
| **Suicidal Ideation** |  |  |  |  |  |  |  |  |  |
| All Effect Sizes Included | 84 | ref. | 0.34 | [0.22; 0.45] | ref. | 0.1842 | ref. | [-0.52; 1.20] | 9.82 |
| 3 Indicators Removed<sup>a</sup> | 81 | 3.6% | 0.27 | [0.18; 0.36] | 20.6% | 0.0783 | 34.8% | [-0.30; 0.83] | 12.74 |
| At Least 2 Indicators Removed | 54 | 35.7% | 0.22 | [0.12; 0.32] | 35.3% | 0.0683 | 39.1% | [-0.31; 0.75] | 15.73 |
| At Least 1 Indicator Removed<sup>a</sup> | - | - | - | - | - | - | - | - | - |
| **Psychotic Disorders** |  |  |  |  |  |  |  |  |  |
| All Effect Sizes Included | 174 | ref. | 0.32 | [0.23; 0.40] | ref. | 0.0741 | ref. | [-0.23; 0.86] | 14.85 |
| 3 Indicators Removed<sup>a</sup> | 172 | 1.1% | 0.30 | [0.22; 0.38] | 6.3% | 0.0644 | 6.8% | [-0.21; 0.81] | 15.76 |
| At Least 2 Indicators Removed | 101 | 42.0% | 0.24 | [0.15; 0.34] | 25.0% | 0.0659 | 5.7% | [-0.28; 0.76] | 20.36 |
| At Least 1 Indicator Removed | 23 | 86.8% | 0.34 | [0.07; 0.61] | -6.3% | 0.0892 | -9.7% | [-0.32; 1.00] | 13.76 |
| **Panic Disorders** |  |  |  |  |  |  |  |  |  |
| All Effect Sizes Included | 88 | ref. | 0.83 | [0.68; 0.98] | ref. | 0.2841 | ref. | [-0.24; 1.90] | 3.83 |
| 3 Indicators Removed<sup>a</sup> | 80 | 9.1% | 0.75 | [0.62; 0.88] | 9.6% | 0.1805 | 20.3% | [-0.10; 1.61] | 4.32 |
| At Least 2 Indicators Removed | 29 | 67.0% | 0.67 | [0.47; 0.86] | 19.3% | 0.1635 | 24.1% | [-0.18; 1.52] | 5.02 |
| At Least 1 Indicator Removed | 6 | 93.2% | 0.44 | [-0.07; 0.94] | 47.0% | 0.1487 | 27.7% | [-0.66; 1.53] | 8.37 |
| **Social Anxiety Disorder** |  |  |  |  |  |  |  |  |  |
| All Effect Sizes Included | 279 | ref. | 0.95 | [0.80; 1.10] | ref. | 0.3460 | ref. | [-0.22; 2.12] | 3.56 |
| 3 Indicators Removed<sup>a</sup> | 258 | 7.5% | 0.83 | [0.73; 0.94] | 12.6% | 0.1234 | 40.3% | [0.14; 1.53] | 4.21 |
| At Least 2 Indicators Removed | 126 | 54.8% | 0.75 | [0.65; 0.86] | 21.1% | 0.0612 | 57.9% | [0.25; 1.25] | 4.81 |
| At Least 1 Indicator Removed<sup>a</sup> | - | - | - | - | - | - | - | - | - |
| **Generalized Anxiety Disorder** |  |  |  |  |  |  |  |  |  |
| All Effect Sizes Included | 118 | ref. | 0.86 | [0.69; 1.03] | ref. | 0.2672 | ref. | [-0.18; 1.90] | 3.66 |
| 3 Indicators Removed<sup>a</sup> | 108 | 8.5% | 0.75 | [0.62; 0.88] | 12.8% | 0.1455 | 26.2% | [-0.02; 1.51] | 4.35 |
| At Least 2 Indicators Removed | 57 | 51.7% | 0.71 | [0.53; 0.88] | 17.4% | 0.1460 | 26.1% | [-0.08; 1.49] | 4.66 |
| At Least 1 Indicator Removed | 10 | 91.5% | 0.69 | [0.25; 1.12] | 19.8% | 0.1023 | 38.1% | [-0.12; 1.49] | 4.83 |
| **Specific Phobias** |  |  |  |  |  |  |  |  |  |
| All Effect Sizes Included | 61 | ref. | 1.25 | [1.01; 1.49] | ref. | 0.4999 | ref. | [-0.18; 2.68] | 2.68 |
| 3 Indicators Removed<sup>a</sup> | 50 | 18.0% | 1.03 | [0.81; 1.25] | 17.6% | 0.2602 | 27.9% | [-0.01; 2.08] | 3.46 |
| At Least 2 Indicators Removed | 13 | 78.7% | 0.98 | [0.6; 1.35] | 21.6% | 0.2076 | 35.6% | [-0.08; 2.03] | 3.74 |
| At Least 1 Indicator Removed<sup>a</sup> | - | - | - | - | - | - | - | - | - |
| **Obsessive-Compulsive Disorder** |  |  |  |  |  |  |  |  |  |
| All Effect Sizes Included | 55 | ref. | 1.18 | [0.96; 1.39] | ref. | 0.2725 | ref. | [0.11; 2.24] | 3.71 |
| 3 Indicators Removed<sup>a</sup> | 50 | 9.1% | 1.10 | [0.93; 1.28] | 6.8% | 0.1542 | 24.8% | [0.29; 1.91] | 4.10 |
| At Least 2 Indicators Removed | 17 | 69.1% | 1.06 | [0.77; 1.35] | 10.2% | 0.1618 | 22.9% | [0.16; 1.96] | 4.35 |
| At Least 1 Indicator Removed<sup>a</sup> | - | - | - | - | - | - | - | - | - |
| **Prolonged Grief Disorder** |  |  |  |  |  |  |  |  |  |
| All Effect Sizes Included | 32 | ref. | 0.49 | [0.24; 0.73] | ref. | 0.3341 | ref. | [-0.71; 1.69] | 5.33 |
| 3 Indicators Removed<sup>a</sup> | 31 | 3.1% | 0.44 | [0.21; 0.66] | 10.2% | 0.2674 | 10.5% | [-0.64; 1.52] | 5.93 |
| At Least 2 Indicators Removed | 23 | 28.1% | 0.31 | [0.12; 0.51] | 36.7% | 0.1235 | 39.2% | [-0.44; 1.06] | 8.19 |
| At Least 1 Indicator Removed | 8 | 75.0% | 0.29 | [-0.08; 0.66] | 40.8% | 0.1485 | 33.3% | [-0.69; 1.27] | 8.77 |
| **Borderline Personality Disorder** |  |  |  |  |  |  |  |  |  |
| All Effect Sizes Included | 28 | ref. | 0.46 | [0.26; 0.67] | ref. | 0.1707 | ref. | [-0.41; 1.34] | 7.48 |
| 3 Indicators Removed<sup>a</sup> | 27 | 3.6% | 0.42 | [0.23; 0.61] | 8.7% | 0.1239 | 14.8% | [-0.33; 1.17] | 8.48 |
| At Least 2 Indicators Removed | 14 | 50.0% | 0.29 | [0.05; 0.53] | 37.0% | 0.1086 | 20.2% | [-0.46; 1.04] | 12.94 |
| At Least 1 Indicator Removed<sup>a</sup> | - | - | - | - | - | - | - | - | - |
| **Problem Gambling** |  |  |  |  |  |  |  |  |  |
| All Effect Sizes Included | 81 | ref. | 0.80 | [0.43; 1.17] | ref. | 0.8725 | ref. | [-1.09; 2.69] | 4.65 |
| 3 Indicators Removed<sup>a</sup> | 75 | 7.4% | 0.55 | [0.35; 0.76] | 31.2% | 0.2083 | 51.1% | [-0.38; 1.48] | 7.49 |
| At Least 2 Indicators Removed | 53 | 34.6% | 0.46 | [0.22; 0.70] | 42.5% | 0.2268 | 49.0% | [-0.52; 1.44] | 9.46 |
| At Least 1 Indicator Removed | 8 | 90.1% | 0.09 | [-0.46; 0.65] | 88.8% | 0.0000 | 100.0% | [-0.12; 0.31] | 57.38 |

Note. <sup>a</sup>Not enough effect sizes available for meta-analysis ( $k \geq 3$ ). <sup>b</sup>As part of the tool, effect sizes scoring positive on all three risk indicators are regarded as "flagged". $n[eff]$ = number of effect sizes; $\Delta[n]$ = percent difference in the number of available effect sizes compared to the "All Effect Sizes Included" analysis; SMD = standardized mean difference; $\Delta[SMD]$ = percent difference in the pooled effect size (SMD) compared to the "All Effect Sizes Included" analysis; $\tau^2$ = between-study heterogeneity variance; $\Delta[\tau^2]$ = percent difference in between-study heterogeneity variance compared to the "All Effect Sizes Included" analysis; PI = prediction interval; NNT = number needed to treat.

Figure 2 shows the change in effect size and heterogeneity when removing flagged effects across all possible subgroup-specific analyses (i.e., individual treatment formats, such as CBT for depression, compared to all available control groups, such as waitlists or care as usu-al). In total, 385 recalculated meta-analyses were performed, of which 272 (70.6%) resulted in a change in the pooled effect and/or heterogeneity. As shown in the figure, the majority of re-analyses lead to a reduction in both the pooled effect and heterogeneity (bottom-left quadrant). In 13% of all analyses (n=50), the estimated between-study heterogeneity was reduced by 100% to exactly zero. When using the flagging tool as intended (i.e., removing effects with three positive indicators only), all reanalyzed meta-analyses showed both lower effect estimates and between-study heterogeneity.

**Figure 2:**
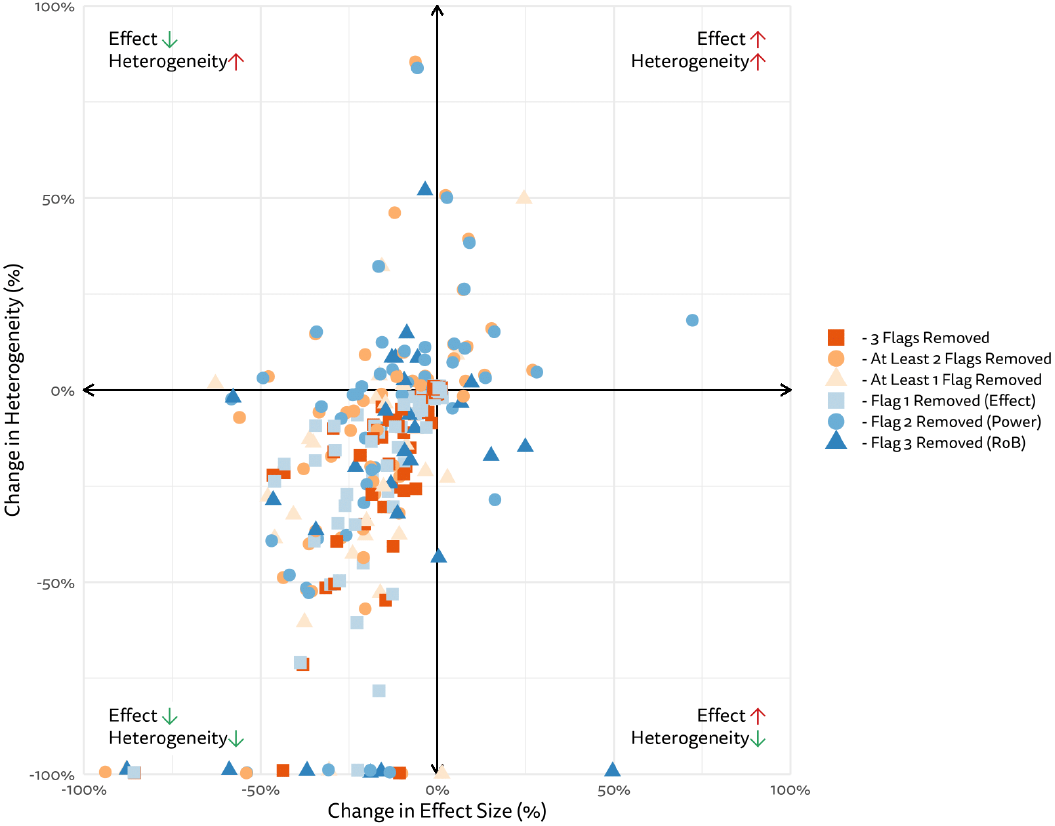
Change in effect size and heterogeneity when removing flagged effects across all possible subgroup-specific analyses.

## Discussion

In this study, we developed a simple tool to flag implausible effect sizes. Applying it to 2,881 effect sizes from 12 living databases, we found that about 5% of the available evidence would be flagged under its current calibration. Re-analysis of more than 300 meta-analytic effects further showed that excluding flagged studies often resulted in substantially lower pooled estimates and reduced between-study heterogeneity.

Our findings corroborate that extreme and implausible effects are not uncommon, affecting roughly one in twelve trials investigating psychological treatments (8.2%). We found that the number of such “problematic” studies published each year is increasing, but not faster than publication rates overall. This indicates that untrustworthy trials are not a new phenomenon, but that the scale of the problem has increased in the last decades as scientific output in general has surged (Pan et al., 2018; Harrer, Miguel, van Ballegooijen, et al., 2025). Flagged studies were not confined to any specific region: among earlier trials, nearly all originated from high-income countries, whereas in the last ten years, LMICs were overrepresented. Several explanations may account for this pattern. Intervention research has expanded considerably in LMICs (Harrer, Miguel, van Ballegooijen, et al., 2025), while groups in these areas may also have less experience with trial methodology, statistical evaluation, and reporting. Future studies could examine in greater detail how trial quality relates to local research capacities, including access to methodological expertise and infrastructural support (Dean et al., 2017).

The flagged effect sizes in our validation sample ranged from SMD=1.88 to 15.41. This is about four to 30 times as large as the reference effect we calculated for low risk of bias evidence (SMD=0.46), but also the median effects across psychological research more generally (Bogdan, 2025). To be flagged, studies had to report effects of this magnitude in combination with strongly insufficient sample sizes and low methodological rigor; extreme effects were still considered when they were found in studies with at least minimally adequate power, low risk of bias, or both. In this regard, we believe the tool strikes a sensible balance between filtering out published results that clearly strain credulity, without *a priori* ruling out the possibility of genuine “breakthrough” findings (Djulbegovic et al., 2025) – as long as they are reported in trials minimally suited to support such claims.

As outlined in the introduction, our tool is not intended to “prove” that a flagged effect is spurious, or to establish whether the result arises from reporting errors, flawed methodology, or even data fabrication. In some cases, a detailed investigation of a particular publication may clarify the source of an implausibly large effect, but this is far from guaranteed. Conversely, studies may report seemingly “plausible” results even when, in reality, questionable research practices were applied. Keeping in mind these limitations, the ever-increasing number of trial publications each year, and the fact that resources in research are scarce, we view our tool a pragmatic compromise. In meta-analysis projects, for example, use of the tool could be prespecified, providing clear-cut criteria that prevent highly implausible results from distorting the main quantitative synthesis. Flagged studies could then be included in a sensitivity analysis, but without “contaminating” the main results, which are typically the reported in the abstract. Within the Metapsy infrastructure, the flagging tool is fully integrated into the “metapsyTools” package, allowing users to include it as a preliminary analysis step when working with one of the openly accessible living databases, or user-supplied data. Since the tool operates exclusively on routinely collected information, it can be applied “out-of-the-box” without further extraction steps.

A more practical limitation is that the reference values for our tool were derived from a fairly specific field (i.e., psychological interventions for mental health). Possibly, thresholds suitable in this context may not always transfer to other domains, for example educational or occupational interventions. However, in principle, our approach is extensible to other fields if sufficiently large meta-analytic databases are available. We also saw that average effects differ between mental health problems as well, which may affect the calibration of the tool. To this end, we developed three indication-specific reference values for psychosis, depression, and PTSD, which can also serve as proxies for “low”, “moderate”, and “high-effects” fields. Finally, it is important to note that we exclusively focused on intervention effects compared to weak or inactive controls. Trials involving head-to-head comparisons between two “bona fide” treatments were excluded because (1) their average effects are smaller, making implausible results less likely to be detected alongside, e.g., waitlist-controlled trials; (2) fewer such trials are available; and (3) because conventional power calculations may not apply (e.g. in non-inferiority designs; see preregistration for further details).

In sum, our study indicates that a simple flagging tool for implausible effects could be a meaningful improvement in evidence syntheses and meta-research. Our findings suggest that only a small share of intervention trials will typically be flagged; yet their exclusion can substantially reduce bias in pooled estimates, and curb inflated between-study heterogeneity.

## Data Availability

Data and analysis code are available at: 10.5281/zenodo.17143121

## References

Andrade, C. (2021). HARKing, cherry-picking, p-hacking, fishing expeditions, and data dredging and mining as questionable research practices. The Journal of Clinical Psychiatry, 82(1), 25941.

Balcetis, E., & Dunning, D. (2010). Wishful Seeing: More Desired Objects Are Seen as Closer. Psychological Science, 21(1), 147–152. 10.1177/0956797609356283

Basic, D. (2025). Database of randomized clinical trials comparing psychological interventions for schizophrenia and psychosis with control conditions. Part of the Metapsy project (Version 23.1.4). 10.5281/zenodo.7782324

Bogdan, P. C. (2025). One Decade Into the Replication Crisis, How Have Psychological Results Changed? Advances in Methods and Practices in Psychological Science, 8(2), 25152459251323480. 10.1177/25152459251323480

Campbell Collaboration. (2020). Campbell systematic reviews: Policies and guidelines. 10.4073/cpg.2016.1

Chinn, S. (2000). A simple method for converting an odds ratio to effect size for use in meta-analysis. Statistics in Medicine, 19(22), 3127–3131.

Clark, A. M., Sousa, B. J., Ski, C. F., & Thompson, D. R. (2025). Honest yet unacceptable research practices: When research becomes a health risk. BMJ Open, 15(6), e097757.

Ćmiel, B., Inglot, T., & Ledwina, T. (2020). Intermediate efficiency of some weighted goodness-of-fit statistics. Journal of Nonparametric Statistics, 32(3), 667–703. 10.1080/10485252.2020.1789126

Cuijpers, P., Karyotaki, E., Reijnders, M., & Ebert, D. D. (2019). Was Eysenck right after all? A reassessment of the effects of psychotherapy for adult depression. Epidemiology and Psychiatric Sciences, 28(1), 21–30.

Cuijpers, P., Miguel, C., Harrer, M., Plessen, C. Y., Ciharova, M., Ebert, D., & Karyotaki, E. (2025). Database of depression psychotherapy trials with control conditions. Part of the Metapsy project (Version 24.0.2). Zenodo. 10.5281/zenodo.7254845

Cuijpers, P., Miguel, C., Harrer, M., Plessen, C. Y., Ciharova, M., Papola, D., Ebert, D., & Karyotaki, E. (2023). Psychological treatment of depression: A systematic overview of a ‘Meta-Analytic Research Domain.’ Journal of Affective Disorders, 335, 141–151.

Cuijpers, P., Miguel, C., Papola, D., Harrer, M., & Karyotaki, E. (2022). From living systematic reviews to meta-analytical research domains. Evidence-Based Mental Health, 25(4), 145–147.

de Ponti, N., Matbouriahi, M., Franco, P., Harrer, M., Miguel, C., Papola, D., Sicimoğlu, A., Cuijpers, P., & Karyotaki, E. (2025). Database of social anxiety disorder trials comparing psychological interventions with control conditions. Part of the Metapsy project (Version 24.0.1). 10.5281/zenodo.12726122

Dean, L., Gregorius, S., Bates, I., & Pulford, J. (2017). Advancing the science of health research capacity strengthening in low-income and middle-income countries: A scoping review of the published literature, 2000–2016. BMJ Open, 7(12), e018718. 10.1136/bmjopen-2017-018718

Djulbegovic, B., Hozo, I., Iskander, R., Parish, A. J., Kimmelman, J., & Ioannidis, J. P. (2025). There is no upper limit on the maximum effect that can be detected in randomized trials. Journal of Clinical Epidemiology, 111828.

Harrer, M. (2025). Extreme and Implausible Effect Sizes in Meta-Analyses of Psychological Treatments: Meta-Epidemiological Study and Development of a Simple Flagging Tool (Materials & Code) (Version 1.0.0). The Metapsy Collaboration. 10.5281/zenodo.17143121

Harrer, M., Cuijpers, P., A, F. T., & Ebert, D. D. (2021). Doing Meta-Analysis With R: A Hands-On Guide (1st ed.). Chapman & Hall/CRC Press.

Harrer, M., Kuper, P., Sprenger, A. A., & Cuijpers, P. (2022). metapsyTools: Several R Helper Functions For the “Metapsy” Database. tools.metapsy.org

Harrer, M., Miguel, C., Luo, Y., Ostinelli, E. G., Karyotaki, E., Leucht, S., Furukawa, T. A., & Cuijpers, P. (2025). Standardized effect sizes are far from “Standardized”: A primer and empirical illustration in depression psychotherapy meta-analyses. PLOS Mental Health, 2(7), e0000347.

Harrer, M., Miguel, C., van Ballegooijen, W., Ciharova, M., Plessen, C. Y., Kuper, P., Sprenger, A. A., Buntrock, C., Papola, D., Cristea, I. A., de Ponti, N., BašiĆ, Đ., Pauley, D., Driessen, E., Quero, S., Grimaldos, J., Buendía, S. F., Botella, C., Hamblen, J. L., … Cuijpers, P. (2025). Effectiveness of psychotherapy: Synthesis of a “meta-analytic research domain” across world regions and 12 mental health problems. Psychological Bulletin, 151(5), 600–667. 10.1037/bul0000465

Higgins, J. P. T., Chandler, J., Cumpston, M., Li, T., Page, M. J., & Welch, V. A. (2024). Cochrane handbook for systematic reviews of interventions version 6.4 (updated August 2024). Cochrane, 2024. www.cochrane.org/handbook

Hilgard, J. (2021). Maximal positive controls: A method for estimating the largest plausible effect size. Journal of Experimental Social Psychology, 93, 104082.

Ioannidis, J. P. A. (2021). Hundreds of thousands of zombie randomised trials circulate among us. Anaesthesia, 76(4), 444–447. 10.1111/anae.15297

John, L. K., Loewenstein, G., & Prelec, D. (2012). Measuring the Prevalence of Questionable Research Practices With Incentives for Truth Telling. Psychological Science, 23(5), 524–532. 10.1177/0956797611430953

Lin, L., Chu, H., & Hodges, J. S. (2017). Alternative Measures of Between-Study Heterogeneity in Meta-Analysis: Reducing the Impact of Outlying Studies. Biometrics, 73(1), 156–166. 10.1111/biom.12543

Miguel, C., Harrer, M., Karyotaki, E., Sakher, E., Sakata, M., Furukawa, T. A., & Cuijpers, P. (2025). Operationalization of Cochrane’s Risk of Bias 2 Tool (RoB 2) in the Context of Psychotherapy Trials (p. 2025.06.26.25330349). medRxiv. 10.1101/2025.06.26.25330349

Mol, B. W., Lai, S., Rahim, A., Bordewijk, E. M., Wang, R., van Eekelen, R., Gurrin, L. C., Thornton, J. G., van Wely, M., & Li, W. (2023). Checklist to assess Trustworthiness in RAndomised Controlled Trials (TRACT checklist): Concept proposal and pilot. Research Integrity and Peer Review, 8(1), 6. 10.1186/s41073-023-00130-8

National Center for PTSD, US Department of Veterans Affairs. (2025). Database of PTSD trials comparing psychological interventions with control conditions. Part of the Metapsy project (Version 23.0.3). 10.5281/zenodo.10027042

Page, M. J., McKenzie, J. E., Bossuyt, P. M., Boutron, I., Hoffmann, T. C., Mulrow, C. D., Shamseer, L., Tetzlaff, J. M., Akl, E. A., & Brennan, S. E. (2021). The PRISMA 2020 statement: An updated guideline for reporting systematic reviews. Bmj, 372. https://www.bmj.com/content/372/bmj.n71.short

Pan, R. K., Petersen, A. M., Pammolli, F., & Fortunato, S. (2018). The memory of science: Inflation, myopia, and the knowledge network. Journal of Informetrics, 12(3), 656–678. 10.1016/j.joi.2018.06.005

Papola, D. (2025a). Database of GAD trials comparing psychological interventions with control conditions. Part of the Metapsy project (Version 23.0.4). 10.5281/zenodo.10185216

Papola, D. (2025b). Database of panic disorder psychotherapy trials with control conditions. Part of the Metapsy project (Version 23.0.1). 10.5281/zenodo.7863722

Pfund, R. A. (2025). Database of psychological interventions for problem gambling and gambling disorder trials with control conditions. Part of the Metapsy project (Version 25.0.0). 10.5281/zenodo.8115993

Pustejovsky, J. E., & Tipton, E. (2022). Meta-analysis with robust variance estimation: Expanding the range of working models. Prevention Science, 23(3), 425–438.

Quero, S., Fernández-Buendia, S., Grimaldos, J., de la Coba-Cañizares, L., & Miguel, C. (2025). Database of prolonged grief trials comparing psychological interventions with control conditions. Part of the Metapsy project (Version 25.0.4). 10.5281/zenodo.15538811

Rücker, G., Schwarzer, G., Carpenter, J. R., Binder, H., & Schumacher, M. (2011). Treatment-effect estimates adjusted for small-study effects via a limit meta-analysis. Biostatistics, 12(1), 122–142.

Setkowski, K., Palantza, C., van Ballegooijen, W., Gilissen, R., Oud, M., Cristea, I., Noma, H., Furukawa, T., Arntz, A., van Balkom, A., & Cuijpers, P. (2025). Database of borderline personality disorder trials comparing psychological interventions with control conditions. Part of the Metapsy project (Version 23.0.1). 10.5281/zenodo.10185551

Sterne, J. A., SavoviĆ, J., Page, M. J., Elbers, R. G., Blencowe, N. S., Boutron, I., Cates, C. J., Cheng, H.-Y., Corbett, M. S., & Eldridge, S. M. (2019). RoB 2: A revised tool for assessing risk of bias in randomised trials. Bmj, 366.

Van Aert, R. C., Wicherts, J. M., & Van Assen, M. A. (2019). Publication bias examined in meta-analyses from psychology and medicine: A meta-meta-analysis. PloS One, 14(4), e0215052.

van Ballegooijen, W., Rawee, J., Palantza, C., Miguel, C., Harrer, M., & Cuijpers, P. (2025). Database of psychological interventions for suicide prevention trials with control conditions. Part of the Metapsy project (Version 24.0.1). 10.5281/zenodo.8364508

Wang, Y., Miguel, C., Ciharova, M., Amarnath, A., Lin, J., Zhao, R., Toffolo, M. B. J., Struijs, S. Y., de Wit, L. M., & Cuijpers, P. (2025). Database of OCD trials comparing psychological interventions with control conditions. Part of the Metapsy project (Version 24.0.1). 10.5281/zenodo.10471942

Wilkinson, J., Heal, C., Antoniou, G. A., Flemyng, E., Alfirevic, Z., Avenell, A., Barbour, G., Brown, N. J., Carlisle, J., & Clarke, M. (2024). Protocol for the development of a tool (INSPECT-SR) to identify problematic randomised controlled trials in systematic reviews of health interventions. BMJ Open, 14(3), e084164.

