## Supplement for "Implausible Effects of Psychological Interventions: Meta-Epidemiological Study and Development of a Simple Flagging Tool"

### S1. PRISMA flowcharts for all included databases

#### ***Depression***

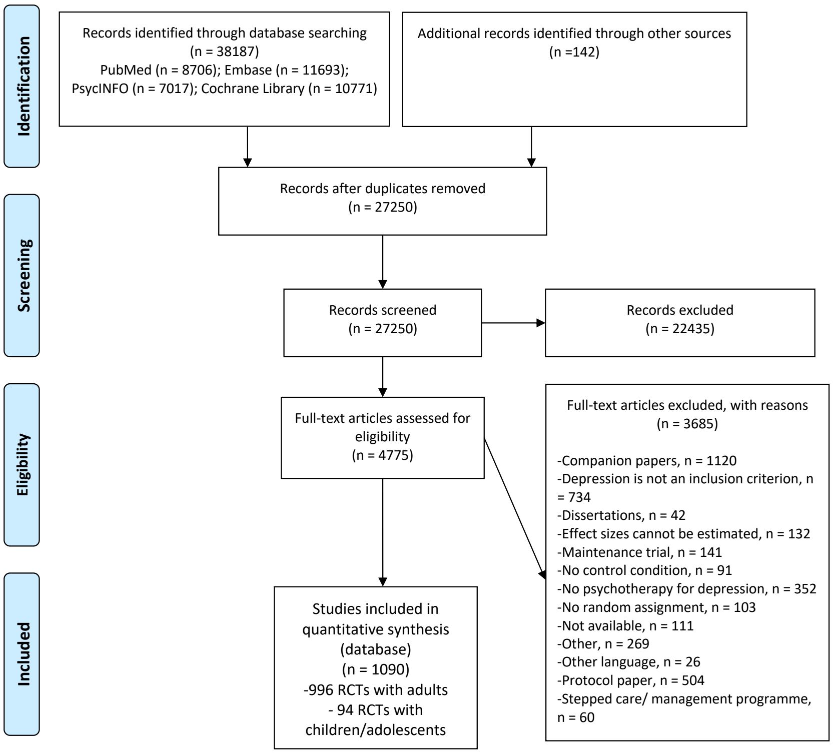

Search update: 2025-01-01

#### ***Anxiety Disorders***

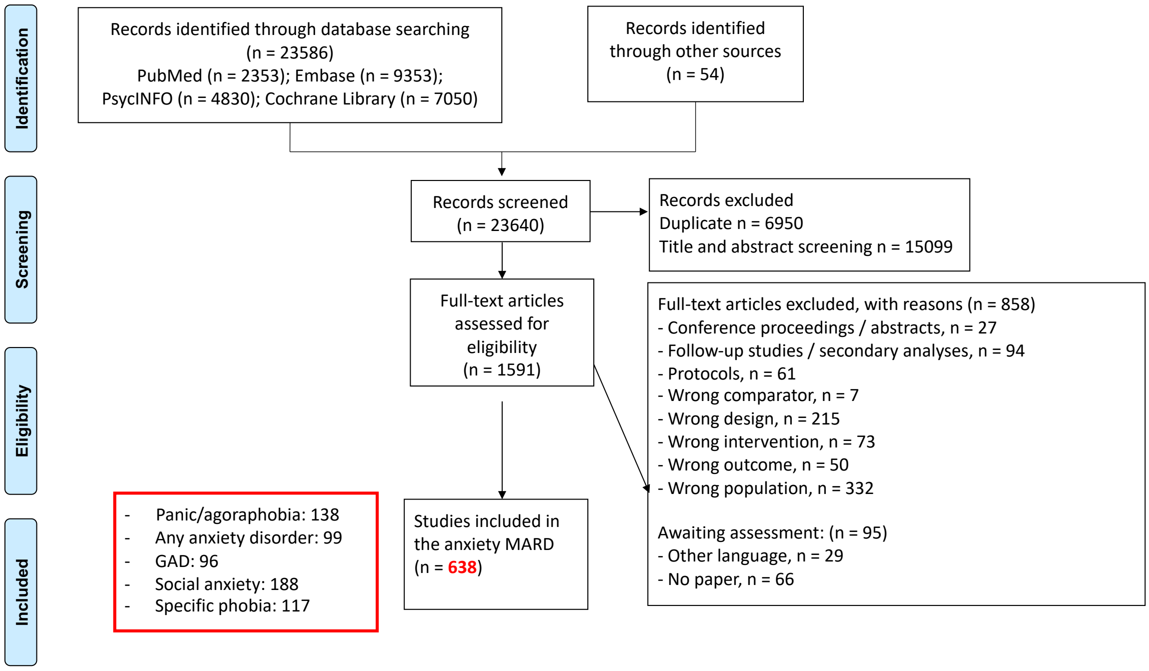

Search update: 2025-01-01

### ***Posttraumatic Stress Disorder***

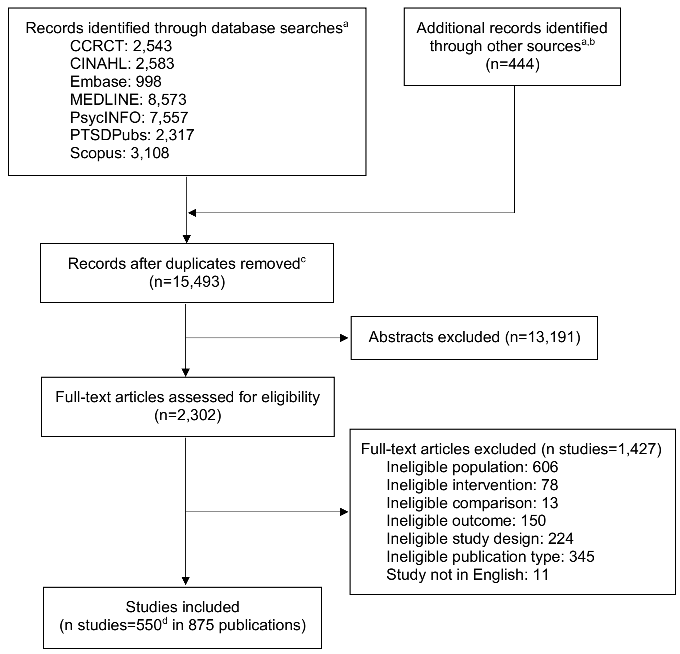

[a]Multiple update searches were performed with overlapping search dates; reported is number of unique records from each source across searches.
[b]Other sources include prior reports, reference lists of relevant articles, systematic reviews, etc.
[c]Number of unique records across sources.
[d]In this update report, there are 54 new trials.[21-74] There are 550 total studies in 548 records (Appendix B).

Search update: 2025-01-01

### ***Suicidal Ideation***

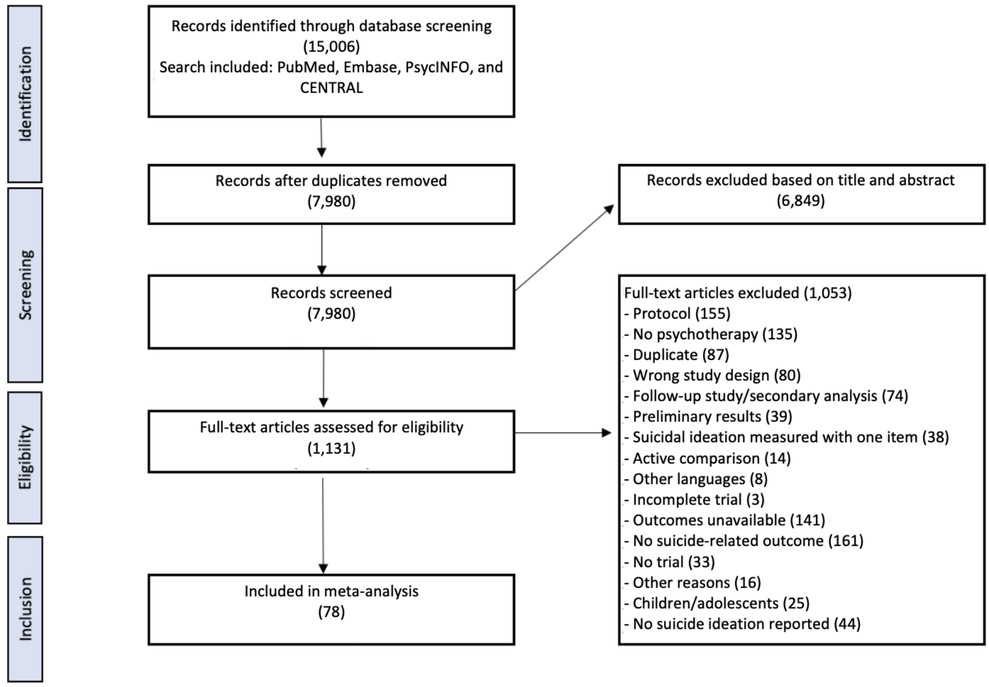

Search update: 2023-04-01

### *Psychosis*

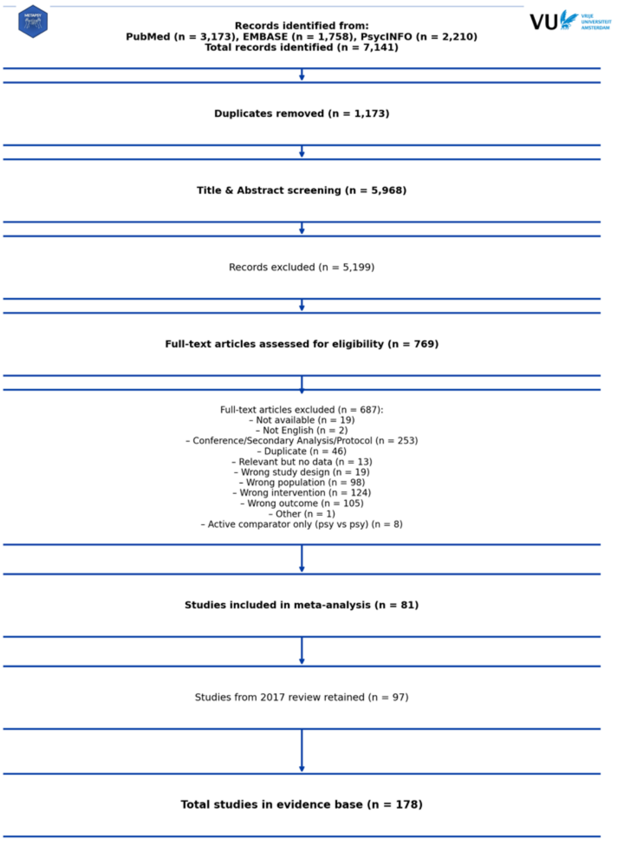

Search update: 2025-03-01

### *Obsessive-Compulsive Disorder*

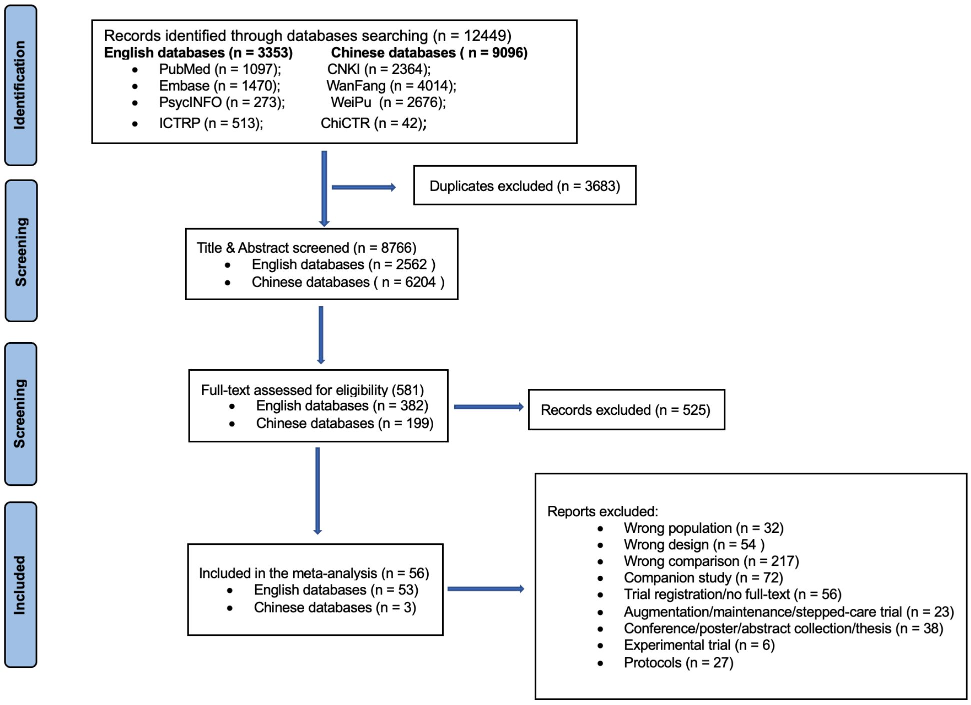

Search update: 2024-09-01

### ***Prolonged Grief Disorder***

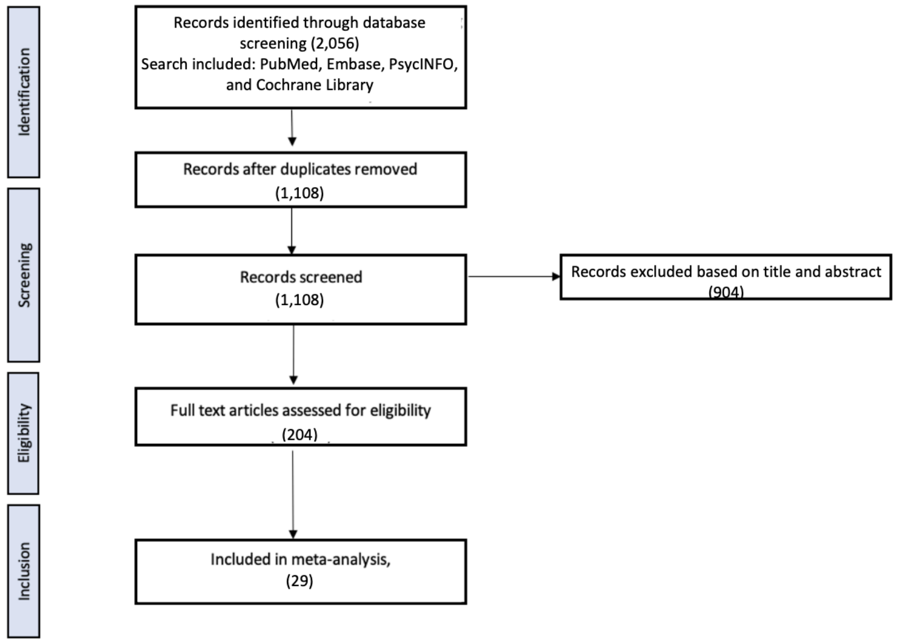

Search update: 2023-01-01

### ***Borderline Personality Disorder***

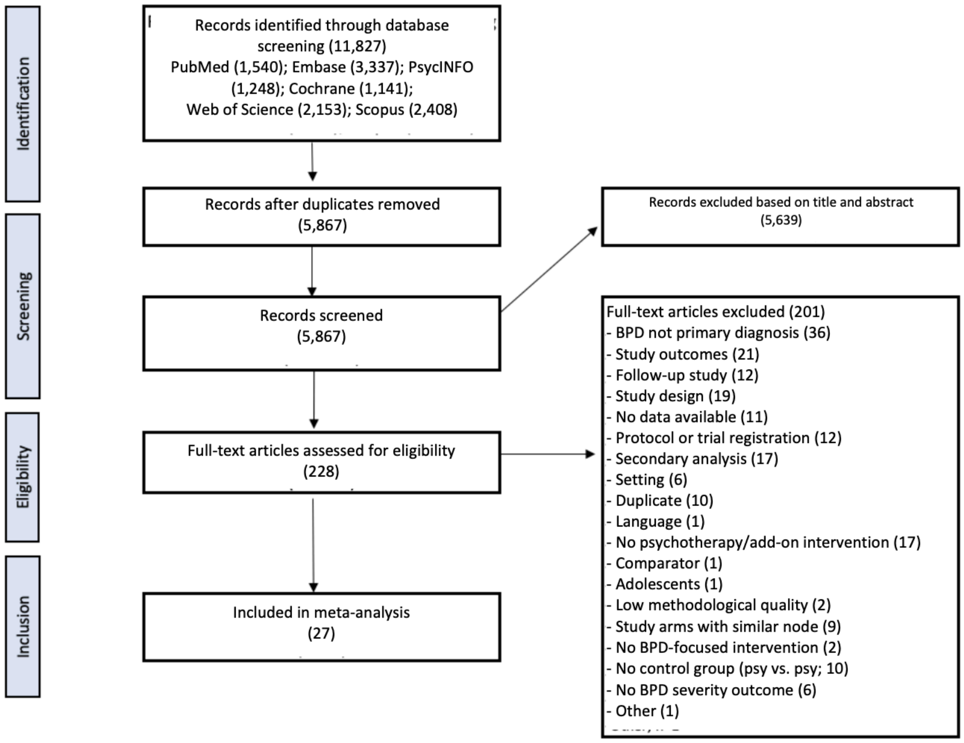

Search update: 2023-01-01

### Problem Gambling

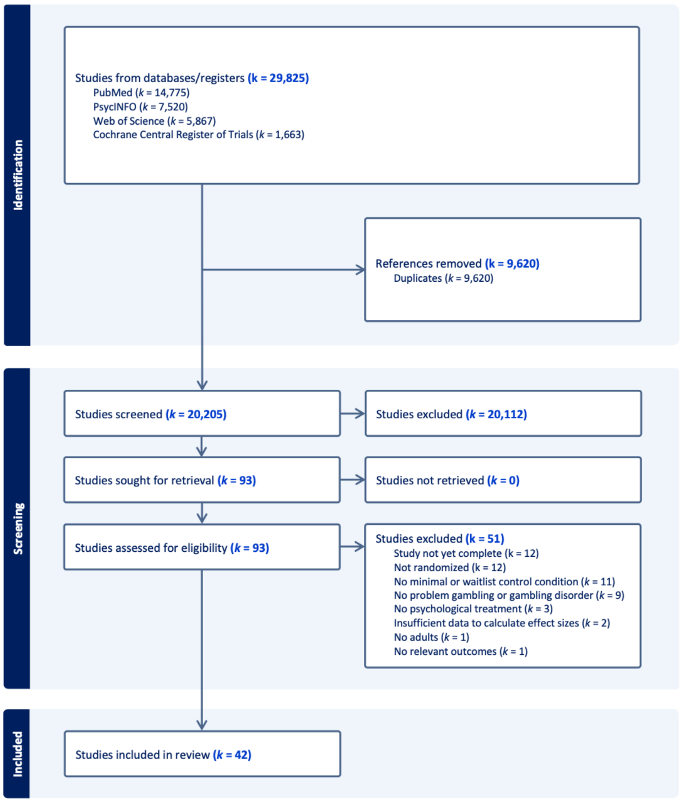

Search update: 2025-01-01

### S2. References of the included studies

Aagaard, J., Foldager, L., Makki, A., Hansen, V., & M¸ller-Nielsen, K. (2017). The efficacy of psychoeducation on recurrent depression: a randomized trial with a 2-year follow-up. Nord J Psychiatry, 71(3), 223-229.

Abas M, Nyamayaro P, Bere T, et al. Feasibility and acceptability of a task-shifted intervention to enhance adherence to HIV medication and improve depression in people living with HIV in Zimbabwe, a low income country in sub-Saharan Africa. AIDS and Behavior 2018; 22(1): 86-101.

Abbas, Q., Latif, S., Ayaz Habib, H., Shahzad, S., Sarwar, U., Shahzadi, M., . . . Washdev, W. (2023). Cognitive behavior therapy for diabetes distress, depression, health anxiety, quality of life and treatment adherence among patients with Type-II diabetes mellitus: A randomized control trial. BMC Psychiatry, 23. doi:10.1186/s12888-023-04546-w

Abbas, Q., Nisa, M., Khan, M. U., Anwar, N., Aljhani, S., Ramzan, Z., & Shahzadi, M. (2023). Brief cognitive behavior therapy for stigmatization, depression, quality of life, social support and adherence to treatment among patients with HIV/AIDS: a randomized control trial. BMC Psychiatry, 23(1), 539.

Abbott, M., Hodgins, D. C., Bellringer, M., Vandal, A. C., Palmer Du Preez, K., Landon, J., ... & Feigin, V. (2018). Brief telephone interventions for problem gambling: A randomized controlled trial. Addiction, 113(5), 883-895.

Abramowitz, J. S., Moore, E. L., Braddock, A. E., & Harrington, D. L. (2009). Self-help cognitive-behavioral therapy with minimal therapist contact for social phobia: a controlled trial. J Behav Ther Exp Psychiatry, 40(1), 98-105. https://doi.org/10.1016/j.jbtep.2008.04.004

Abujilban, S., Al-Omari, H., Issa, E., Alhamdan, A., Al-Nabulsi, L., Mrayan, L., . . . Kernohan, W. G. (2024). Effectiveness of Telephone-Based Interpersonal Psychotherapy on Antenatal Depressive Symptoms: A Prospective Randomized Controlled Trial in The Kingdom of Jordan. Journal of the American Psychiatric Nurses Association, 30(3), 635-645. doi:https://doi.org/10.1177/10783903231171595

Acarturk C, Konuk E, Cetinkaya M, et al. The efficacy of eye movement desensitization and reprocessing for post-traumatic stress disorder and depression among Syrian refugees: results of a randomized controlled trial. Psychol Med. 2016 Sep;46(12):2583-93. doi: 10.1017/S0033291716001070. PMID: 27353367.

Addis ME, Hatgis C, Krasnow AD, et al. Effectiveness of cognitive--behavioral treatment for panic disorder versus treatment as usual in a managed care setting. Journal of consulting and clinical psychology 2004;72(4):625-35.

Afonso, R., & Bueno, B. (2009). Efectos de un programa de reminiscencia sobre la sintomatología depresiva en una muestra de población mayor portuguesa. Revista Española de Geriatría y Gerontología, 44(6), 317-322.

Afrasiabifar, Ardashir & Hosseini, Nazafarin & Haghgoo, Amin. (2018). The Effects of Group Cognitive Behavior Therapy (GCBT) on Suicidal Thoughts in Patients with Major Depression. World Family Medicine Journal/Middle East Journal of Family Medicine. 16. 228-235. 10.5742/MEWFM.2018.93293.

Aghotor J, Pfueller U, Moritz S, Weisbrod M, Roesch-ely D. Metacognitive training for patients with schizophrenia (MCT): Feasibility and preliminary evidence for its efficacy. J Behav Ther Exp Psychiatry [Internet]. Elsevier Ltd; 2010;41(3):207, 11. Available from: http://dx.doi.org/10.1016/j.jbtep.2010.01.004

Aguilar-Raab C, Winter F, Warth M, Stoffel M, Moessner M, Hernández C, et al. A compassion-based treatment for couples with the female partner suffering from current depressive disorder: A randomized-controlled trial. Journal of Affective Disorders. 2023;342:127-38.

Ahmadi K, Hazrati M, Ahmadizadeh MJ, et al. REM desensitization as a new therapeutic method for post-traumatic stress disorder: a randomized controlled trial. Acta Med Indones. 2015 Apr;47(2):111-9. PMID: 26260553.

Ahmadpanah, M., Paghale, S. J., Bakhtyari, A., Kaikhavani, S., Aghaei, E., Nazaribadie, M., . . . Brand, S. (2016). Effects of psychotherapy in combination with pharmacotherapy, when compared to

pharmacotherapy only on blood pressure, depression, and anxiety in female patients with hypertension.
Ahmed N, Zavala GA, Siddiqui F, Aslam F, Keding A, Halmkan S, et al. A randomised controlled feasibility trial of Behavioural activation as a treatment for people with diabetes and depression: (DiaDeM feasibility trial). J Affect Disord. 2025;372:333-46.
Alavi, N., & Hirji, A. (2020). The Efficacy of PowerPoint-based CBT Delivered Through Email: Breaking the Barriers to Treat-ment for Generalized Anxiety Disorder. Journal of Psychiatric Practice, 26(2), 89-100. doi:10.1097/PRA.0000000000000455
Alavi, N., & Hirji, A. (2020). The Efficacy of PowerPoint-based CBT Delivered Through Email: Breaking the Barriers to Treatment for Generalized Anxiety Disorder. Journal of Psychiatric Practice, 26(2), 89-100. doi:10.1097/PRA.0000000000000455
Alcolado, G. M., & Radomsky, A. S. (2016). A novel cognitive intervention for compulsive checking: Targeting maladaptive beliefs about memory. Journal of Behavior Therapy and Experimental Psychiatry, 53, 75-83.
Aldahadha B, Al-Harthy H, Sulaiman S. The Efficacy of Eye Movement Desensitization Reprocessing in Resolving the Trauma Caused by the Road Accidents in the Sultanate of Oman. Journal of Instructional Psychology. 2012;39(3/4):146-58.
Alexopoulos GS, Raue PJ, McCulloch C, Kanellopoulos D, Seirup JK, Sirey JA, et al. Clinical case management versus case management with problem-solving therapy in low-income, disabled elders with major depression: A randomized clinical trial. American Journal of Geriatric Psychiatry. 2016;24(1):50-9.
Alghamdi M, Hunt NC, Thomas SA. The effectiveness of narrative exposure therapy with traumatised firefighters in Saudi Arabia: a randomized controlled study. Behav Res Ther. 2015 Mar;66:64-71. doi: 10.1016/j.brat.2015.01.008. PMID: 25701801.
Alhusen, J. L., Hayat, M. J., & Borg, L. (2021, Feb). A pilot study of a group-based perinatal depression intervention on reducing depressive symptoms and improving maternal-fetal attachment and maternal sensitivity. Arch Womens Ment Health, 24(1), 145-154. https://doi.org/10.1007/s00737-020-01032-0
Allart-Van Dam E, Hosman CMH, Hoogduin CAL, Schaap CPDR. The coping with depression course: Short-term outcomes and mediating effects of a randomized controlled trial in the treatment of subclinical depression. Behavior Therapy. 2003;34(3):381-96.
Alsheikh Ali ASS. Efficiency of Intervention Counseling Program on the Enhanced Psychological Well-being and Reduced Post-traumatic Stress Disorder Symptoms Among Syrian Women Refugee Survivors. Clin Pract Epidemiol Ment Health. 2020 Jul 30;16(Suppl-1):134-141. doi: 10.2174/1745017902016010134. PMID: 33029190; PMCID: PMC7536727.
Amani, B., Merza, D., Savoy, C., Streiner, D., Bieling, P., Ferro, M. A., & Van Lieshout, R. J. (2021, Nov 9). Peer-Delivered Cognitive-Behavioral Therapy for Postpartum Depression: A Randomized Controlled Trial. J Clin Psychiatry, 83(1). https://doi.org/10.4088/JCP.21m13928
Amano, M., Katayama, N., Umeda, S., Terasawa, Y., Tabuchi, H., Kikuchi, T., . . . Nakagawa, A. (2023). The effect of cognitive behavioral therapy on future thinking in patients with major depressive disorder: A randomized controlled trial. Frontiers in Psychiatry, 14. doi:10.3389/fpsyt.2023.997154
Amianto, F., Ferrero, A., Pierò, A., Cairo, E., Rocca, G., Simonelli, B., ... & Fassino, S. (2011). Supervised team management, with or without structured psychotherapy, in heavy users of a mental health service with borderline personality disorder: a two-year follow-up preliminary randomized study. BMC psychiatry, 11(1), 1-14.
Ammerman RT, Putnam FW, Altaye M, Stevens J, Teeters AR, Van Ginkel JB. A clinical trial of in-home CBT for depressed mothers in home visitation. Behavior Therapy. 2013;44(3):359-72.
Anderson, P. L., Price, M., Edwards, S. M., Obasaju, M. A., Schmertz, S. K., Zimand, E., & Calamaras, M. R. (2013). Virtual reality exposure therapy for social anxiety disorder: a randomized controlled trial. J Consult Clin Psychol, 81(5), 751-760. https://doi.org/10.1037/a0033559
Anderson, R. A., & Rees, C. S. (2007). Group versus individual cognitive-behavioural treatment for obsessive-compulsive disorder: a controlled trial. Behav Res Ther, 45(1), 123-137.
Andersson G, Bergström J, Holländare F, Carlbring P, Kaldo V, Ekselius L. Internet-based self-help for depression: Randomised controlled trial. British Journal of Psychiatry. 2005;187(5):456-61.

Andersson, E., Enander, J., Andrén, P., Hedman, E., Ljótsson, B., Hursti, T., . . . Rück, C. (2012). Internet-based cognitive behaviour therapy for obsessive-compulsive disorder: a randomized controlled trial. Psychol Med, 42(10), 2193-2203.

Andersson, G., Carlbring, P., Holmstrom, A., Sparthan, E., Furmark, T., Nilsson-Ihrfelt, E., Buhrman, M., & Ekselius, L. (2006). Internet-based self-help with therapist feedback and in vivo group exposure for social phobia: a randomized controlled trial. J Consult Clin Psychol, 74(4), 677-686. https://doi.org/10.1037/0022-006x.74.4.677

Andersson, G., Paxling, B., Roch-Norlund, P., Ostman, G., Norgren, A., Almlov, J., . . . Silverberg, F. (2012). Internet-based psychodynamic versus cognitive behavioral guided self-help for generalized anxiety disorder: a randomized controlled trial. Psychother Psychosom, 81(6), 344-355. doi:10.1159/000339371

Andersson, G., Paxling, B., Roch-Norlund, P., Ostman, G., Norgren, A., Almlov, J., . . . Silverberg, F. (2012). Internet-based psy-chodynamic versus cognitive behavioral guided self-help for generalized anxiety disorder: a randomized controlled trial. Psychother Psychosom, 81(6), 344-355. doi:10.1159/000339371

Andrade, A. S., Moreira, M., Sá, M., Pacheco, D., Almeida, V., & Rocha, J. C. (2017). Randomized controlled trial of a cognitive narrative crisis intervention for bereavement in primary healthcare. Behavioural and Cognitive Psychotherapy, 45(1), 85-90.

Andreasson, K., Krogh, J., Wenneberg, C., Jessen, H. K., Krakauer, K., Gluud, C., Thomsen, R. R., Randers, L., & Nordentoft, M. (2016). Effectiveness of dialectical behavior therapy versus collaborative assessment and management of suicidality treatment for reduction of self-harm in adults with borderline personality traits and disorder-a randomized observer-blinded clinical trial. Depression and anxiety, 33(6), 520–530. https://doi.org/10.1002/da.22472

Araya, R., Menezes, P. R., Claro, H. G., Brandt, L. R., Daley, K. L., Quayle, J., Diez-Canseco, F., Peters, T. J., Vera Cruz, D., Toyama, M., Aschar, S., Hidalgo-Padilla, L., Martins, H., Cavero, V., Rocha, T., Scotton, G., de Almeida Lopes, I. F., Begale, M., Mohr, D. C., & Miranda, J. J. (2021, May 11). Effect of a Digital Intervention on Depressive Symptoms in Patients With Comorbid Hypertension or Diabetes in Brazil and Peru: Two Randomized Clinical Trials. Jama, 325(18), 1852-1862. https://doi.org/10.1001/jama.2021.4348

Arean PA, Perri MG, Nezu AM, Schein RL, Christopher F, Joseph TX. Comparative effectiveness of social problem-solving therapy and reminiscence therapy as treatments for depression in older adults. Journal of Consulting and Clinical Psychology. 1993;61(6):1003-10.

Arjadi R, Nauta MH, Scholte WF, et al. Internet-based behavioural activation with lay counsellor support versus online minimal psychoeducation without support for treatment of depression: a randomised controlled trial in Indonesia. The lancet psychiatry 2018; 5(9): 707-16.

Arnevik, E., Wilberg, T., Urnes, O., Johansen, M., Monsen, J. T., & Karterud,S. (2009). Psychotherapy for personality disorders: short-term day hospital psychotherapy versus outpatient individual therapy - a randomized controlled study. European psychiatry : the journal of the Association of European Psychiatrists, 24(2), 71–78. https://doi.org/10.1016/j.eurpsy.2008.09.004

Arntz A, Tiesema M, Kindt M. Treatment of PTSD: a comparison of imaginal exposure with and without imagery rescripting. J Behav Ther Exp Psychiatry. 2007 Dec;38(4):345-70. doi: 10.1016/j.jbtep.2007.10.006. PMID: 18005935.

Asukai N, Saito A, Tsuruta N, et al. Efficacy of exposure therapy for Japanese patients with posttraumatic stress disorder due to mixed traumatic events: A randomized controlled study. J Trauma Stress. 2010 Dec;23(6):744-50. doi: 10.1002/jts.20589. PMID: 21171135.

Au, A., Nan, H., Sum, R., Ng, F., Kwong, A., & Wong, S. (2022). Cognitive behavioural therapy for adherence and sub-clinical depression in type 2 diabetes: a randomised controlled trial (abridged secondary publication). Hong Kong Med J, 28 Suppl 3(3), 21-23.

Ayar, D., & Sabanciogullari, S. (2021). The effect of a solution-oriented approach in depressive patients on social functioning levels and suicide probability. Perspectives in psychiatric care, 57(1), 235-245.

Ayen I, Hautzinger M. Cognitive behavior therapy for depression in menopausal women. A controlled, randomized treatment study. Zeitschrift fur Klinische Psychologie und Psychotherapie. 2004;33(4):290-9.

Babiy, Z., Layton, H., Savoy, C. D., Xie, F., Brown, J. S. L., Bieling, P. J., . . . Van Lieshout, R. J. (2024). One-Day Peer-Delivered Cognitive Behavioral Therapy-Based Workshops for Postpartum Depression: A Randomized Controlled Trial. Psychotherapy and Psychosomatics, 93(2), 129-140. doi:https://doi.org/10.1159/000536040

Baker AL, Kavanagh DJ, Kay-Lambkin FJ, Hunt SA, Lewin TJ, Carr VJ, et al. Randomized controlled trial of cognitive-behavioural therapy for coexisting depression and alcohol problems: short-term outcome. Addiction (Abingdon, England). 2010;105(1):87-99.

Bakhshani, N. M., Lashkaripour, K., & Sadjadi, S. A. (2007). Effectiveness of short term cognitive behavior therapy in patients with generalized anxiety disorder. Journal of Medical Sciences, 7(7), 1076-1081. doi:10.3923/jms.2007.1076.1081

Bakhshani, N. M., Lashkaripour, K., & Sadjadi, S. A. (2007). Effectiveness of short term cognitive behavior therapy in patients with generalized anxiety disorder. Journal of Medical Sciences, 7(7), 1076-1081. doi:10.3923/jms.2007.1076.1081

Bakker A, van Dyck R, Spinhoven P, et al. Paroxetine, clomipramine, and cognitive therapy in the treatment of panic disorder. The Journal of clinical psychiatry 1999;60(12):831-8.

Bannan, Noreen, (2010), Group-based problem-solving therapy in self- poisoning females: A pilot study. Counselling and Psychotherapy Research, 10 doi: 10.1080/14733140903337292.

Barber JP, Barrett MS, Gallop R, Rynn MA, Rickels K. Short-term dynamic psychotherapy versus pharmacotherapy for major depressive disorder: A randomized, placebo-controlled trial. Journal of Clinical Psychiatry. 2012;73(1):66-73.

Barbosa, V., Sá, M., & Carlos Rocha, J. (2014). Randomised controlled trial of a cognitive narrative intervention for complicated grief in widowhood. Aging & Mental Health, 18(3), 354-362.

Bark N, Revheim N, Huq F, Khalderov V, Watras Z, Medalia A. The impact of cognitive remediation on psychiatric symptoms of schizophrenia. Schizophr Res. 2003;63:229, 35.

Barlow DH, Craske MG, Cerny JA, Klosko JS. Behavioral treatment of panic disorder. Behavior Therapy 1989;20(2): 261–82.

Barlow DH, Gorman JM, Shear MK, et al. Cognitive-behavioral therapy, imipramine, or their combination for panic disorder: A randomized controlled trial. Jama 2000;283(19):2529-36.

Barlow, D. H., Rapee, R. M., & Brown, T. A. (1992). Behavioral treatment of generalized anxiety disorder. Behavior Therapy, 23(4), 551-570. doi:https://doi.org/10.1016/S0005-7894(05)80221-7

Barlow, D. H., Rapee, R. M., & Brown, T. A. (1992). Behavioral treatment of generalized anxiety disorder. Behavior Therapy, 23(4), 551-570. doi:https://doi.org/10.1016/S0005-7894(05)80221-7

Barnes, S. M., Borges, L. M., Smith, G. P., Walser, R. D., Forster, J. E., & Bahraini, N. H. (2021). Acceptance and commitment therapy to promote recovery from suicidal crises: A randomized controlled acceptability and feasibility trial of ACT for life. Journal of Contextual Behavioral Science, 20, 35-45.

Barnhofer T, Crane C, Hargus E, Amarasinghe M, Winder R, Williams JM. Mindfulness-based cognitive therapy as a treatment for chronic depression: A preliminary study. Behaviour Research and Therapy. 2009;47(5):366-73.

Barnhofer, T., Crane, C., Hargus, E., Amarasinghe, M., Winder, R., & Williams, J. M. (2009). Mindfulness-based cognitive therapy as a treatment for chronic depression: A preliminary study. Behaviour research and therapy, 47(5), 366–373. https://doi.org/10.1016/j.brat.2009.01.019

Barrett JE, Williams Jr JW, Oxman TE, Frank E, Katon W, Sullivan M, et al. Treatment of dysthymia and minor depression in primary care: A ramdomized trial in patients aged 18 to 59 years. Journal of Family Practice. 2001;50(5):405-12.

Barrowclough C, Haddock G, Lobban F, Jones S, Siddle RON, Roberts C, et al. Group cognitive-behavioural therapy for schizophrenia: Randomised controlled trial. Br J Psychiatry. 2006;(189):527, 32.

Basirat, Z., Kheirkhah, F., Faramarzi, M., Esmaelzadeh, S., Khafri, S., & Tajali, Z. (2022). Pharmacotherapy or Psychotherapy? Selective Treatment Depression in The Infertile Women with Recurrent Pregnancy Loss: A Triple-Arm Randomized Controlled Trial. International Journal of Fertility and Sterility, 16(3), 211-219. doi:10.22074/ijfs.2021.529258.1124

Basoglu M, Salcioglu E, Livanou M, et al. Single-session behavioral treatment of earthquake-related posttraumatic stress disorder: a randomized waiting list controlled trial. J Trauma Stress. 2005;18(1):1-11. doi: 10.1002/jts.20011. PMID: 16281190.

Basoglu M, Salcioglu E, Livanou M. A randomized controlled study of single-session behavioural treatment of earthquake-related post-traumatic stress disorder using an earthquake simulator. Psychological Medicine. 2007;37(2):203-13. doi: 10.1017/S0033291706009123. PMID: 17254365.

Bateman, A., & Fonagy, P. (2009). Randomized controlled trial of outpatient mentalization-based treatment versus structured clinical management for borderline personality disorder. American journal of Psychiatry, 166(12), 1355-1364.

Baumeister, H., Paganini, S., Sander, L. B., Lin, J., Schlicker, S., Terhorst, Y., Moshagen, M., Bengel, J., Lehr, D., & Ebert, D. D. (2021). Effectiveness of a Guided Internet- and Mobile-Based Intervention for Patients with Chronic Back Pain and Depression (WARD-BP): A Multicenter, Pragmatic Randomized Controlled Trial. Psychother Psychosom, 90(4), 255-268. https://doi.org/10.1159/000511881

Baumgartner, C., Schaub, M. P., Wenger, A., Malischnig, D., Augsburger, M., Lehr, D., . . . Haug, S. (2021). "Take Care of You" - Efficacy of integrated, minimal-guidance, internet-based self-help for reducing co-occurring alcohol misuse and depression symptoms in adults: Results of a three-arm randomized controlled trial. Drug Alcohol Depend, 225, 108806. doi:10.1016/j.drugalcdep.2021.108806

Beach SR, O'Leary KD. Treating depression in the context of marital discord: Outcome and predictors of response of marital therapy versus cognitive therapy. Behavior Therapy. 1992;23(4):507-28.

Beck AT, Sokol L, Clark DA, et al. A crossover study of focused cognitive therapy for panic disorder. The American journal of psychiatry 1992;149(6):778-83.

Beck JG, Coffey SF, Foy DW, et al. Group cognitive behavior therapy for chronic posttraumatic stress disorder: an initial randomized pilot study. Behav Ther. 2009 Mar;40(1):82-92. doi: 10.1016/j.beth.2008.01.003. PMID: 19187819.

Beck JG, Stanley MA, Baldwin LE, et al. Comparison of cognitive therapy and relaxation training for panic disorder. Journal of consulting and clinical psychology 1994;62(4):818-26.

Bedard M, Felteau M, Marshall S, Cullen N, Gibbons C, Dubois S, et al. Mindfulness-based cognitive therapy reduces symptoms of depression in people with a traumatic brain injury: Results from a randomized controlled trial. Journal of Head Trauma Rehabilitation. 2014;29(4):E13-E22.

Beeber LS, Holditch-Davis D, Perreira K, Schwartz TA, Lewis V, Blanchard H, et al. Short-term in-home intervention reduces depressive symptoms in Early Head Start Latina mothers of infants and toddlers. Research in Nursing and Health. 2010;33(1):60-76.

Beidel, D. C., Alfano, C. A., Kofler, M. J., Rao, P. A., Scharfstein, L., & Wong Sarver, N. (2014). The impact of social skills training for social anxiety disorder: a randomized controlled trial. J Anxiety Disord, 28(8), 908-918. https://doi.org/10.1016/j.janxdis.2014.09.016

Bellino, S., Rinaldi, C., & Bogetto, F. (2010). Adaptation of interpersonal psychotherapy to borderline personality disorder: a comparison of combined therapy and single pharmacotherapy. The Canadian Journal of Psychiatry, 55(2), 74-81.

Bendig, E., Bauereiß, N., Buntrock, C., Habibović, M., Ebert, D. D., & Baumeister, H. (2021). Lessons learned from an attempted randomized-controlled feasibility trial on "WIDeCAD" - An internet-based depression treatment for people living with coronary artery disease (CAD) [Article]. Internet Interventions, 24. https://doi.org/10.1016/j.invent.2021.100375

Bengtson AM, Filipowicz TR, Mphonda S, Udedi M, Kulisewa K, Meltzer-Brody S, et al. An Intervention to Improve Mental Health and HIV Care Engagement Among Perinatal Women in Malawi: A Pilot Randomized Controlled Trial. AIDS and behavior. 2023.

Bentley, K. H., Sauer-Zavala, S., Cassiello-Robbins, C. F., Conklin, L. R., Vento, S., & Homer, D. (2017). Treating Suicidal Thoughts and Behaviors Within an Emotional Disorders Framework: Acceptability and Feasibility of the Unified Protocol in an Inpatient Setting. Behavior modification, 41(4), 529–557. https://doi.org/10.1177/0145445516689661

Berger T, Hämmerli K, Gubser N, Andersson G, Caspar F. Internet-based treatment of depression: A randomized controlled trial comparing guided with unguided self-help. Cognitive Behaviour Therapy. 2011;40(4):251-66.

Berger, T., Hohl, E., & Caspar, F. (2009). Internet-based treatment for social phobia: a randomized controlled trial. J Clin Psychol, 65(10), 1021-1035. https://doi.org/10.1002/jclp.20603

Berman, M. I., Park, J., Kragenbrink, M. E., & Hegel, M. T. (2022). Accept Yourself! A Pilot Randomized Controlled Trial of a Self-Acceptance-Based Treatment for Large-Bodied Women With Depression. Behav Ther, 53(5), 913-926. doi:10.1016/j.beth.2022.03.002

Beutel ME, Weissflog G, Leuteritz K, Wiltink J, Haselbacher A, Ruckes C, et al. Efficacy of short-term psychodynamic psychotherapy (STPP) with depressed breast cancer patients: Results of a randomized controlled multicenter trial. Annals of Oncology. 2014;25(2):378-84.

Bilich, L. L., Deane, F. P., Phipps, A. B., Barisic, M., & Gould, G. (2008). Effectiveness of bibliotherapy self-help for depression with varying levels of telephone helpline support. Clin Psychol Psychother, 15(2), 61-74. doi:10.1002/cpp.562

Bisson JI, van Deursen R, Hannigan B, Kitchiner N, Barawi K, Jones K, Pickles T, Skipper J, Young C, Abbott LR, van Gelderen M, Nijdam MJ, Vermetten E. Randomized controlled trial of multi-modular motion-assisted memory desensitization and reconsolidation (3MDR) for male military veterans with treatment-resistant post-traumatic stress disorder. Acta Psychiatr Scand. 2020 Aug;142(2):141-151. doi: 10.1111/acps.13200. Epub 2020 Jun 28. PMID: 32495381.

Black DW, Wesner R, Bowers W, et al. A comparison of fluvoxamine, cognitive therapy, and placebo in the treatment of panic disorder. Archives of general psychiatry 1993;50(1):44-50.

Blanchard EB, Hickling EJ, Devineni T, et al. A controlled evaluation of cognitive behavioural therapy for posttraumatic stress in motor vehicle accident survivors. Behav Res Ther. 2003 Jan;41(1):79-96. PMID: 12488121.

Blanco, C., Heimberg, R. G., Schneier, F. R., Fresco, D. M., Chen, H., Turk, C. L., Vermes, D., Erwin, B. A., Schmidt, A. B., Juster, H. R., Campeas, R., & Liebowitz, M. R. (2010). A placebo-controlled trial of phenelzine, cognitive behavioral group therapy, and their combination for social anxiety disorder. Arch Gen Psychiatry, 67(3), 286-295. https://doi.org/10.1001/archgenpsychiatry.2010.11

Blomdahl, C., Guregård, S., Rusner, M., & Wijk, H. (2018). A manual-based phenomenological art therapy for individuals diagnosed with moderate to severe depression (PATd): A randomized controlled study. Psychiatric rehabilitation journal, 41(3), 169–182. https://doi.org/10.1037/prj0000300

Boele FW, Klein M, Verdonck-de Leeuw IM, et al. Internet-based guided self-help for glioma patients with depressive symptoms: a randomized controlled trial. Journal of neuro-oncology 2018; 137(1): 191-203.

Boele, F. W., et al. (2021). "The effects of SmartCare© on neuro-oncology family caregivers' distress: a randomized controlled trial." Supportive Care in Cancer.

Boeschoten, R. E., Dekker, J., Uitdehaag, B. M. J., Beekman, A. T. F., Hoogendoorn, A. W., Collette, E. H., . . . Van Oppen, P. (2017). Internet-based treatment for depression in multiple sclerosis: A randomized controlled trial. Multiple Sclerosis, 23(8), 1112-1122.

Bohlmeijer ET, Fledderus M, Rokx TA, Pieterse ME. Efficacy of an early intervention based on acceptance and commitment therapy for adults with depressive symptomatology: Evaluation in a randomized controlled trial. Behaviour research and therapy. 2011;49(1):62-7.

Bohus MJ, Dyer AS, Priebe K, et al. Dialectical behaviour therapy for post-traumatic stress disorder after childhood sexual abuse in patients with and without borderline personality disorder: a randomised controlled trial. Psychother Psychosom. 2013 Jun;82(4):221-33. doi: 10.1159/000348451. PMID: 23712109.

Bolton P, Bass J, Neugebauer R, Verdeli H, Clougherty KF, Wickramaratne P, et al. Group interpersonal psychotherapy for depression in rural Uganda: A randomized controlled trial. JAMA. 2003;289(23):3117-24.

Booth, R., & Rachman, S. (1992). The reduction of claustrophobia—I. Behaviour Research and Therapy, 30(3), 207-221. doi:https://doi.org/10.1016/0005-7967(92)90067-Q

Bornas, X., Tortella-Feliu, M., Llabres, J., & Fullana, M. (2001). Computer-Assisted Exposure Treatment for Flight Phobia: a Controlled Study. Psychotherapy Research - PSYCHOTHER RES, 11, 259-273. doi:10.1080/713663983

Botella, C. García-Palacios, A. Villa, H. Baños, R. M. Quero, S. Alcañiz, M. Riva, G. Virtual reality exposure in the treatment of panic disorder and agoraphobia: A controlled study. Clinical Psychology & Psychotherapy 2007. 14(4)164-75.

Botella, C., Gallego, M. J., Garcia-Palacios, A., Guillen, V., Baños, R. M., Quero, S., & Alcañiz, M. (2010). An Internet-based self-help treatment for fear of public speaking: a controlled trial. Cyberpsychol Behav Soc Netw, 13(4), 407-421. https://doi.org/10.1089/cyber.2009.0224
Bouchard, S., Dumoulin, S., Robillard, G., Guitard, T., Klinger, E., Forget, H., Loranger, C., & Roucaut, F. X. (2017). Virtual reality compared with in vivo exposure in the treatment of social anxiety disorder: a three-arm randomised controlled trial. Br J Psychiatry, 210(4), 276-283. https://doi.org/10.1192/bjp.bp.116.184234
Boudreault, C., Giroux, I., Jacques, C., Goulet, A., Simoneau, H., & Ladouceur, R. (2018). Efficacy of a self-help treatment for at-risk and pathological gamblers. Journal of Gambling Studies, 34(2), 561-580.
Bower, J. E., Partridge, A. H., Wolff, A. C., Thorner, E. D., Irwin, M. R., Joffe, H., Petersen, L., Crespi, C. M., & Ganz, P. A. (2021, Nov 1). Targeting Depressive Symptoms in Younger Breast Cancer Survivors: The Pathways to Wellness Randomized Controlled Trial of Mindfulness Meditation and Survivorship Education. J Clin Oncol, 39(31), 3473-3484. https://doi.org/10.1200/jco.21.00279
Bowman D, Scogin F, Lyrene B. The efficacy of self-examination therapy and cognitive bibliotherapy in the treatment of mild to moderate depression. Psychotherapy Research. 1995;5(2):131-40.
Bozzatello, P., & Bellino, S. (2020). Interpersonal psychotherapy as a single treatment for borderline personality disorder: a pilot randomized-controlled study. Frontiers in Psychiatry, 11, 578910.
Bradley GM, Couchman GM, Perlesz A, Nguyen AT, Singh B, Riess C. Multiple-family group treatment for families living with schizophrenia. Psychiatr Serv. 2006;57(4):521, 30.
Bradshaw RA, McDonald MJ, Grace R, et al. A randomized clinical trial of Observed and Experiential Integration (OEI): a simple, innovative intervention for affect regulation in clients with PTSD. Traumatology (Tallahass Fla). 2014 Sep;20(3):161-71. doi: 10.1037/h0099401.
Braga, D. T., Abramovitch, A., Fontenelle, L. F., Ferrão, Y. A., Gomes, J. B., Vivan, A. S., . . . Cordioli, A. V. (2016). Neuropsychological predictors of treatment response to cognitive behavioral group therapy in obsessive-compulsive disorder. Depress Anxiety, 33(9), 848-861.
Braun, L., Titzler, I., Terhorst, Y., Freund, J., Thielecke, J., Ebert, D. D., & Baumeister, H. (2021, Jan 1). Effectiveness of guided internet-based interventions in the indicated prevention of depression in green professions (PROD-A): Results of a pragmatic randomized controlled trial. J Affect Disord, 278, 658-671. https://doi.org/10.1016/j.jad.2020.09.066
Brenner, L. A., Forster, J. E., Hoffberg, A. S., Matarazzo, B. B., Hostetter, T. A., Signoracci, G., & Simpson, G. K. (2018). Window to Hope: A Randomized Controlled Trial of a Psychological Intervention for the Treatment of Hopelessness Among Veterans With Moderate to Severe Traumatic Brain Injury. The Journal of head trauma rehabilitation, 33(2), E64–E73. https://doi.org/10.1097/HTR.0000000000000351
Britton, P. C., Conner, K. R., Chapman, B. P., & Maisto, S. A. (2020). Motivational interviewing to address suicidal ideation: A randomized controlled trial in veterans. Suicide and Life-Threatening Behavior, 50(1), 233-248.
Brom D, Kleber RJ, Defares PB. Brief psychotherapy for posttraumatic stress disorders. J Consult Clin Psychol. 1989;57(5):607-12. PMID: 2571625.
Brown RA, Lewinsohn PM. A psychoeducational approach to the treatment of depression: Comparison of group, individual, and minimal contact procedures. Journal of Consulting and Clinical Psychology. 1984;52(5):774.
Brown, G. K., Ten Have, T., Henriques, G. R., Xie, S. X., Hollander, J. E., & Beck, A. T. (2005). Cognitive therapy for the prevention of suicide attempts: a randomized controlled trial. JAMA, 294(5), 563–570. https://doi.org/10.1001/jama.294.5.563
Bryan, C. J., Mintz, J., Clemans, T. A., Leeson, B., Burch, T. S., Williams, S. R., Maney, E., & Rudd, M. D. (2017). Effect of crisis response planning vs. contracts for safety on suicide risk in U.S. Army Soldiers: A randomized clinical trial. Journal of affective disorders, 212, 64–72. https://doi.org/10.1016/j.jad.2017.01.028
Bryant RA, Kenny L, Rawson N, et al. Efficacy of exposure-based cognitive behaviour therapy for post-traumatic stress disorder in emergency service personnel: a randomised clinical trial. Psychol Med. 2019 Jul;49(9):1565-73. doi: 10.1017/S0033291718002234. PMID: 30149825.

Buchholz, J. L., Blakey, S. M., Hellberg, S. N., Massing-Schaffer, M., Reuman, L., Ojalehto, H., . . . Abramowitz, J. S. (2022). Expectancy violation during exposure therapy: A pilot randomized controlled trial. Journal of Behavioral and Cognitive Therapy, 32(1), 13-24. doi:https://doi.org/10.1016/j.jbct.2021.12.004

Buck, H. G., Cairns, P., Emechebe, N., Hernandez, D. F., Mason, T. M., Bell, J., ... & Tofthagen, C. (2020). Accelerated resolution therapy: Randomized controlled trial of a complicated grief intervention. American Journal of Hospice and Palliative Medicine®, 37(10), 791-799.

Bücker, L., Bierbrodt, J., Hand, I., Wittekind, C., & Moritz, S. (2018). Effects of a depression-focused internet intervention in slot machine gamblers: a randomized controlled trial. PloS one, 13(6), e0198859.

Bücker, L., Gehlenborg, J., Moritz, S., & Westermann, S. (2021). A randomized controlled trial on a self-guided Internet-based intervention for gambling problems. Scientific Reports, 11(1), 13033.

Buhmann CB, Nordentoft M, Ekstrom M, et al. The effect of flexible cognitive-behavioural therapy and medical treatment, including antidepressants on post-traumatic stress disorder and depression in traumatised refugees: pragmatic randomised controlled clinical trial. Br J Psychiatry. 2016 Mar;208(3):252-9. doi: 10.1192/bjp.bp.114.150961. PMID: 26541687.

Buhrman M, Syk M, Burvall O, Hartig T, Gordh T, Andersson G. Individualized Guided Internet-delivered Cognitive Behaviour Therapy for Chronic Pain Patients with Comorbid Depression and Anxiety: A Randomized Controlled Trial. Clinical Journal of Pain 2014.

Buntrock C, Ebert D, Lehr D, Riper H, Smit F, Cuijpers P, et al. Effectiveness of a web-based cognitive behavioural intervention for subthreshold depression: Pragmatic randomised controlled trial. Psychotherapy and Psychosomatics. 2015;84(6):348-58.

Burns A, Banerjee S, Morris J, Woodward Y, Baldwin R, Proctor R, et al. Treatment and prevention of depression after surgery for hip fracture in older people: Randomized, controlled trials. Journal of the American Geriatrics Society. 2007;55(1):75-80.

Burns A, O'Mahen H, Baxter H, Bennert K, Wiles N, Ramchandani P, et al. A pilot randomised controlled trial of cognitive behavioural therapy for antenatal depression. BMC psychiatry. 2013;13:33.

Butler O, Willmund G, Gleich T, et al. Hippocampal gray matter increases following multimodal psychological treatment for combat-related post-traumatic stress disorder. Brain Behav. 2018 May;8(5):e00956. doi: 10.1002/brb3.956. PMID: 29761009.

Butler, G., Fennell, M., Robson, P., & Gelder, M. (1991). Comparison of behavior therapy and cognitive behavior therapy in the treatment of generalized anxiety disorder. J Consult Clin Psychol, 59(1), 167-175.

Butler, G., Fennell, M., Robson, P., & Gelder, M. (1991). Comparison of behavior therapy and cognitive behavior therapy in the treatment of generalized anxiety disorder. J Consult Clin Psychol, 59(1), 167-175.

Byrne LK, Peng D, McCabe M, Mellor D, Zhang J, Zhang T, et al. Does practice make perfect? Results from a Chinese feasibility study of cognitive remediation in schizophrenia. Neuropsychol Rehabilitation. 2013;23(4):580, 96.

Cai J, Zhu Y, Zhang W, Wang Y, Zhang C. Comprehensive family therapy: an effective approach for cognitive rehabilitation in schizophrenia. Neuropsychiatr Dis Treat. 2015;11:1247, 53.

Campos, D., Bretón-López, J., Botella, C., Mira, A., Castilla, D., Mor, S., . . . Quero, S. (2019). Efficacy of an internet-based exposure treatment for flying phobia (NO-FEAR Airlines) with and without therapist guidance: a randomized controlled trial. BMC Psychiatry, 19(1), 86. doi:10.1186/s12888-019-2060-4

Capafóns, J. I., Sosa, C. D., & Avero, P. (1997). La desensibilización sistemática en el tratamiento de la fobia a viajar en transporte áero. [Systematic desensitization in the treatment of fear of flying.]. Psicothema, 9, 17-25.

Capafóns, J. I., Sosa, C. D., & Avero, P. (1997). La desensibilización sistemática en el tratamiento de la fobia a viajar en transporte áero. [Systematic desensitization in the treatment of fear of flying.]. Psicothema, 9, 17-25.

Carl, J. R., Miller, C. B., Henry, A. L., Davis, M. L., Stott, R., Smits, J. A. J., . . . Espie, C. A. (2020). Efficacy of digital cognitive behavioral therapy for moderate-to-severe symptoms of generalized anxiety disorder: A randomized controlled trial. Depression and Anxiety. doi:10.1002/da.23079

Carl, J. R., Miller, C. B., Henry, A. L., Davis, M. L., Stott, R., Smits, J. A. J., . . . Espie, C. A. (2020). Efficacy of digital cognitive behavioral therapy for moderate-to-severe symptoms of generalized anxiety disorder: A randomized controlled trial. De-pression and Anxiety. doi:10.1002/da.23079

Carlbring P, Bohman S, Brunt S, et al. Remote treatment of panic disorder: a randomized trial of internet-based cognitive behavior therapy supplemented with telephone calls. The American journal of psychiatry 2006;163(12):2119-25.

Carlbring P, Hagglund M, Luthstrom A, Dahlin M, Kadowaki A, Vernmark K, et al. Internet-based behavioral activation and acceptance-based treatment for depression: A randomized controlled trial. Journal of Affective Disorders. 2013;148(2-3):331-7.

Carlbring P, Westling BE, Ljungstrand P, et al. Treatment of panic disorder via the Internet: A randomized trial of a self-help program. Behavior Therapy 2001;32(4):751-64.

Carlbring, P., & Smit, F. (2008). Randomized trial of internet-delivered self-help with telephone support for pathological gamblers. Journal of Consulting and Clinical Psychology, 76(6), 1090-1094.

Carlbring, P., Gunnarsdottir, M., Hedensjo, L., Andersson, G., Ekselius, L., & Furmark, T. (2007). Treatment of social phobia: randomised trial of internet-delivered cognitive-behavioural therapy with telephone support. Br J Psychiatry, 190, 123-128. https://doi.org/10.1192/bjp.bp.105.020107

Carlbring, P., Jonsson, J., Josephson, H., & Forsberg, L. (2010). Motivational interviewing versus cognitive behavioral group therapy in the treatment of problem and pathological gambling: A randomized controlled trial. Cognitive Behaviour Therapy, 39(2), 92-103.

Carlson JG, Chemtob CM, Rusnak K, et al. Eye Movement Desensitization and Reprocessing (EMDR) treatment for combat-related posttraumatic stress disorder. J Trauma Stress. 1998 Jan;11(1):3-24. doi: 10.1023/A:1024448814268. PMID: 9479673.

Carpenter KM, Smith JL, Aharonovich E, Nunes EV. Developing therapies for depression in drug dependence: Results of a stage 1 therapy study. American Journal of Drug and Alcohol Abuse. 2008;34(5):642-52.

Carr, A., Finnegan, L., Griffin, E., Cotter, P., & Hyland, A. (2017). A Randomized Controlled Trial of the Say Yes to Life (SYTL) Positive Psychology Group Psychotherapy Program for Depression: An Interim Report. Journal of Contemporary Psychotherapy, 47(3), 153-161.

Carta MG, Petretto D, Adamo S, Bhat KM, Lecca ME, Mura G, et al. Counseling in primary care improves depression and quality of life. Clinical Practice and Epidemiology in Mental Health. 2012;8.

Carter MM, Sbrocco T, Gore KL, et al. Cognitive-behavioral group therapy versus a wait-list control in the treatment of African American women with panic disorder. Cognitive therapy and research 2003;27(5):505-18.

Carter, G. L., Willcox, C. H., Lewin, T. J., Conrad, A. M., & Bendit, N. (2010). Hunter DBT project: randomized controlled trial of dialectical behaviour therapy in women with borderline personality disorder. Australian & New Zealand Journal of Psychiatry, 44(2), 162-173.

Casanas R, Catalan R, del Val JL, Real J, Valero S, Casas M. Effectiveness of a psycho-educational group program for major depression in primary care: A randomized controlled trial. BMC psychiatry. 2012;12:230.

Casey, L. M., Oei, T. P., Raylu, N., Horrigan, K., Day, J., Ireland, M., & Clough, B. A. (2017). Internet-based delivery of cognitive behaviour therapy compared to monitoring, feedback and support for problem gambling: a randomised controlled trial. Journal of gambling studies, 33(3), 993-1010.

Challacombe, F. L., Salkovskis, P. M., Woolgar, M., Wilkinson, E. L., Read, J., & Acheson, R. (2017). A pilot randomized controlled trial of time-intensive cognitive–behaviour therapy for postpartum obsessive–compulsive disorder: Effects on maternal symptoms, mother–infant interactions and attachment. Psychological Medicine, 47(8), 1478-1488. R

Chan AS, Wong QY, Sze SL, Kwong PP, Han YM, Cheung MC. A Chinese Chan-based mind-body intervention for patients with depression. Journal of Affective Disorders. 2012;142(1-3):283-9.

Chan MF, Ng SE, Tien A, Man Ho RC, Thayala J. A randomised controlled study to explore the effect of life story review on depression in older Chinese in Singapore. Health and Social Care in the Community. 2013;21(5):545-53.

Chan SW, Yip B, Tso S, Cheng B, Tam W. Evaluation of a psychoeducation program for Chinese clients with schizophrenia and their family caregivers. Patient Educ Couns. 2009;75:67, 76.

Chan, N. Y., Lam, S. P., Zhang, J., Chan, J. W. Y., Yu, M. M. W., Suh, S., ... & Li, S. X. (2022). Efficacy of email-delivered versus face-to-face group cognitive behavioral therapy for insomnia in youths: a randomized controlled trial. Journal of Adolescent Health, 70(5), 763-773.

Chanen, A. M., Betts, J. K., Jackson, H., Cotton, S. M., Gleeson, J., Davey, C. G., ... & McCutcheon, L. (2022). Effect of 3 forms of early intervention for young people with borderline personality disorder: The MOBY randomized clinical trial. JAMA psychiatry, 79(2), 109-119.

Chard KM. An evaluation of cognitive processing therapy for the treatment of posttraumatic stress disorder related to childhood sexual abuse. J Consult Clin Psychol. 2005 Oct;73(5):965-71. doi: 10.1037/0022-006X.73.5.965. PMID: 16287396.

Chen CH, Tseng YF, Chou FH, Wang SY. Effects of support group intervention in postnatally distressed women. A controlled study in Taiwan. Journal of psychosomatic research. 2000;49(6):395-9.

Chesney MA, Chambers DB, Taylor JM, Johnson LM, Folkman S. Coping effectiveness training for men living with HIV: Results from a randomized clinical trial testing a group-based intervention. Psychosomatic Medicine. 2003;65(6):1038-46.

Chiang KJ, Chen TH, Hsieh HT, Tsai JC, Ou KL, Chou KR. One-year follow-up of the effectiveness of cognitive behavioral group therapy for patients' depression: A randomized, single-blinded, controlled study. Scientific World Journal. 2015;2015:Article ID 373149.

Chien 2013a: Chien WT, Lee IYM. The mindfulness-based psychoeducation program for Chinese patients with schizophrenia. Psychiatr Serv. 2013;64(4).

Chien 2013b: Chien W-T, Leung S-F. A controlled trial of a needs-based, nurse-led psychoeducation programme for Chinese patients with first-onset mental disorders: 6 month follow up. Int J Nurs Pract. 2013;19:3, 13.

Chien WT, Thompson DR. Effects of a mindfulness-based psychoeducation programme for Chinese patients with schizophrenia: 2-year follow-up. Br J Psychiatry. 2014;205:52, 9.

Cho HJ, Kwon JH, Lee JJ. Antenatal cognitive-behavioral therapy for prevention of postpartum depression: A pilot study. Yonsei medical journal. 2008;49(4):553-62.

Choi I, Zou J, Titov N, Dear BF, Li S, Johnston L, et al. Culturally attuned Internet treatment for depression amongst Chinese Australians: A randomised controlled trial. Journal of Affective Disorders. 2012;136(3):459-68.

Choi NG, Marti CN, Wilson NL, et al. Effect of Telehealth Treatment by Lay Counselors vs by Clinicians on Depressive Symptoms Among Older Adults Who Are Homebound: A Randomized Clinical Trial. JAMA Netw Open. 2020;3(8):e2015648.

Choi, N. G., Hegel, M. T., Marti, C. N., Marinucci, M. L., Sirrianni, L., & Bruce, M. L. (2014). Telehealth problem-solving therapy for depressed low-income homebound older adults. American Journal of Geriatric Psychiatry, 22(3), 263-271.

Chowdhary N, Anand A, Dimidjian S, Shinde S, Weobong B, Balaji M, et al. The Healthy Activity Program lay counsellor delivered treatment for severe depression in India: Systematic development and randomised evaluation. British Journal of Psychiatry. 2016;208(4):381-8.

Choy JC, Lou VW. Effectiveness of the modified instrumental reminiscence intervention on psychological well-being among community-dwelling chinese older adults: A randomized controlled trial. American Journal of Geriatric Psychiatry. 2016;24(1):60-9.

Christ, C., van Schaik, D. J. F., Kikkert, M. J., de Waal, M. M., Dozeman, E., Hulstijn, H. L., . . . Dekker, J. J. M. (2024). Internet-based emotion regulation training aimed at reducing violent revictimization and depressive symptoms in victimized depressed patients: Results of a randomized controlled trial. Journal of Affective Disorders, 355, 95-103. doi:https://doi.org/10.1016/j.jad.2024.03.028

Ciuca AM, Berger T, Crişan LG, et al. Internet-based treatment for panic disorder: A three-arm randomized controlled trial comparing guided (via real-time video sessions) with unguided self-help treatment and a waitlist control. PAXPD study results. Journal of Anxiety Disorders 2018;56:43-55.

Cladder-Micus, M. B., Speckens, A. E. M., Vrijsen, J. N., A.R, T. D., Becker, E. S., & Spijker, J. (2018). Mindfulness-based cognitive therapy for patients with chronic, treatment-resistant depression: A pragmatic randomized controlled trial. Depress Anxiety, 35(10), 914-924.

Clark DM, Salkovskis PM, Hackmann A, et al. Brief cognitive therapy for panic disorder: a randomized controlled trial. Journal of consulting and clinical psychology 1999;67(4):583-9.

Clark R, Tluczek A, Brown R. A mother–infant therapy group model for postpartum depression. Infant Mental Health Journal. 2008;29(5):514-36.

Clark, D. M., Ehlers, A., Hackmann, A., McManus, F., Fennell, M., Grey, N., Waddington, L., & Wild, J. (2006). Cognitive therapy versus exposure and applied relaxation in social phobia: A randomized controlled trial. J Consult Clin Psychol, 74(3), 568-578. https://doi.org/10.1037/0022-006x.74.3.568

Clark, D., Wild, J., Warnock-Parkes, E., Stott, R., Grey, N., Thew, G., & Ehlers, A. (2023). More than doubling the clinical benefit of each hour of therapist time: A randomised controlled trial of internet cognitive therapy for social anxiety disorder. Psychol Med,53(11), 5022-5032. doi:10.1017/S0033291722002008

Cloitre M, Koenen KC, Cohen LR, et al. Skills training in affective and interpersonal regulation followed by exposure: a phase-based treatment for PTSD related to childhood abuse. J Consult Clin Psychol. 2002 Oct;70(5):1067-74. doi: 10.1037//0022-006X.70.5.1067. PMID: 12362957.

Cognitive and performance-based treatments for panic attacks in people with varying degrees of agoraphobic disability. Behaviour Research and Therapy 1996;34(3):253–64.

Cohen S, O'Leary KD, Foran H. A randomized clinical trial of a brief, problem-focused couple therapy for depression. Behavior Therapy. 2010;41(4):433-46.

Conner, K. R., Kearns, J. C., Esposito, E. C., Pizzarello, E., Wiegand, T. J., Britton, P. C., ... & Goldston, D. B. (2021). Pilot RCT of the Attempted Suicide Short Intervention Program (ASSIP) adapted for rapid delivery during hospitalization to adult suicide attempt patients with substance use problems. General hospital psychiatry, 72, 66-72.

Cooper PJ, Murray L, Wilson A, Romaniuk H. Controlled trial of the short- and long-term effect of psychological treatment of post-partum depression. I. Impact on maternal mood. The British Journal of Psychiatry. 2003;182(5):412-9.

Cooper, Z. W., Mowbray, O., Ali, M. K., & Johnson, L. C. M. (2024). Addressing depression and comorbid health conditions through solution-focused brief therapy in an integrated care setting: a randomized clinical trial. BMC Prim Care, 25(1), 313. doi:https://doi.org/10.1186/s12875-024-02561-8

Cordioli, A. V., Heldt, E., Bochi, D. B., Margis, R., De Sousa, M. B., Tonello, J. F., . . . Kapczinski, F. (2003). Cognitive-behavioral group therapy in obsessive-compulsive disorder: A randomized clinical trial. Psychotherapy and Psychosomatics, 72(4), 211-216.

Cottraux, J., Note, I. D., Boutitie, F., Milliery, M., Genouihlac, V., Yao, S. N., ... & Gueyffier, F. (2009). Cognitive therapy versus Rogerian supportive therapy in borderline personality disorder. Psychotherapy and psychosomatics, 78(5), 307-316.

Cragan, M. K., & Deffenbacher, J. L. (1984). Anxiety management training and relaxation as self-control in the treatment of generalized anxiety in medical outpatients. Journal of counseling psychology, 31(2), 123-131. Retrieved from https://www.cochranelibrary.com/central/doi/10.1002/central/CN-00216399/full

Cragan, M. K., & Deffenbacher, J. L. (1984). Anxiety management training and relaxation as self-control in the treatment of generalized anxiety in medical outpatients. Journal of counseling psychology, 31(2), 123-131. Retrieved from https://www.cochranelibrary.com/central/doi/10.1002/central/CN-00216399/full

Cramer H, Salisbury C, Conrad J, Eldred J, Araya R. Group cognitive behavioural therapy for women with depression: Pilot and feasibility study for a randomised controlled trial using mixed methods. BMC psychiatry. 2011;11:82.

Craske MG, Lang AJ, Aikins D, et al. Cognitive behavioral therapy for nocturnal panic. Behavior therapy 2005;36(1):43-54.

Craske MG, Maidenberg E, Bystritsky A. Brief cognitive-behavioral versus nondirective therapy for panic disorder. Journal of behavior therapy and experimental psychiatry 1995;26(2):113-20.

Craske, M. G., et al. (2011). "Disorder-specific impact of coordinated anxiety learning and management treatment for anxiety disorders in primary care." Arch Gen Psychiatry 68(4): 378-388.

Craske, M. G., Niles, A. N., Burklund, L. J., Wolitzky-Taylor, K. B., Vilardaga, J. C., Arch, J. J., Saxbe, D. E., & Lieberman, M. D. (2014). Randomized controlled trial of cognitive behavioral therapy and acceptance and commitment therapy for social phobia: outcomes and moderators. J Consult Clin Psychol, 82(6), 1034-1048. https://doi.org/10.1037/a0037212

Cuijpers, P., Heim, E., Abi Ramia, J., Burchert, S., Carswell, K., Cornelisz, I., . . . El Chammay, R. (2022). Effects of a WHO-guided digital health intervention for depression in Syrian refugees in Lebanon: A randomized controlled trial. PLoS Med, 19(6), e1004025. doi:10.1371/journal.pmed.1004025

Cuijpers, P., Heim, E., Ramia, J. A., Burchert, S., Carswell, K., Cornelisz, I., . . . El Chammay, R. (2022). Guided digital health intervention for depression in Lebanon: randomised trial. Evidence Based Mental Health, 25(e1), e34. doi:10.1136/ebmental-2021-300416

Cukor, D., Ver Halen, N., Asher, D. R., Coplan, J. D., Weedon, J., Wyka, K. E., . . . Kimmel, P. L. (2014). Psychosocial intervention improves depression, quality of life, and fluid adherence in hemodialysis. Journal of the American Society of Nephrology, 25(1), 196-206. doi:https://doi.org/10.1681/asn.2012111134

Cunningham, J. A., Godinho, A., & Hodgins, D. C. (2019). Pilot randomized controlled trial of an online intervention for problem gamblers. Addictive Behaviors Reports, 9, 100175.

Cunningham, J. A., Hodgins, D. C., Toneatto, T., & Murphy, M. (2012). A randomized controlled trial of a personalized feedback intervention for problem gamblers. PLoS One, 7(2), e31586.

Cunningham, J. A., Hodgins, D. C., Toneatto, T., Rai, A., & Cordingley, J. (2009). Pilot study of a personalized feedback intervention for problem gamblers. Behavior Therapy, 40(3), 219-224.

D'Amato T, Bation R, Cochet A, Jalenques I, Galland F, Giraud-Baro E, et al. A randomized, controlled trial of computer-assisted cognitive remediation for schizophrenia. Schizophr Res. 2011;125:284, 90.

Dahlin, M., Andersson, G., Magnusson, K., Johansson, T., Sjogren, J., Hakansson, A., . . . Carlbring, P. (2016). Internet-delivered acceptance-based behaviour therapy for generalized anxiety disorder: A randomized controlled trial. Behav Res Ther, 77, 86-95. doi:10.1016/j.brat.2015.12.007
Dahlin, M., Andersson, G., Magnusson, K., Johansson, T., Sjogren, J., Hakansson, A., . . . Carlbring, P. (2016). Internet-delivered acceptance-based behaviour therapy for generalized anxiety disorder: A randomized controlled trial. Behav Res Ther, 77, 86-95. doi:10.1016/j.brat.2015.12.007
Daneshvar, S., Shafiei, M., & Basharpoor, S. (2022). Compassion-focused therapy: Proof of concept trial on suicidal ideation and cognitive distortions in female survivors of intimate partner violence with PTSD. Journal of interpersonal violence, 37(11-12), NP9613-NP9634.

Davidson, J. R., Foa, E. B., Huppert, J. D., Keefe, F. J., Franklin, M. E., Compton, J. S., Zhao, N., Connor, K. M., Lynch, T. R., & Gadde, K. M. (2004). Fluoxetine, comprehensive cognitive behavioral therapy, and placebo in generalized social phobia. Arch Gen Psychiatry, 61(10), 1005-1013. https://doi.org/10.1001/archpsyc.61.10.1005

Davidson, K. M., Brown, T. M., James, V., Kirk, J., & Richardson, J. (2014). Manual-assisted cognitive therapy for self-harm in personality disorder and substance misuse: a feasibility trial. Psychiatric bulletin (2014), 38(3), 108– 111. https://doi.org/10.1192/pb.bp.113.043109

Davidson, K., Norrie, J., Tyrer, P., Gumley, A., Tata, P., Murray, H., & Palmer, S. (2006). The effectiveness of cognitive behavior therapy for borderline personality disorder: results from the borderline personality disorder study of cognitive therapy (BOSCOT) trial. Journal of personality disorders, 20(5), 450-465.

Davoudi M, Taheri AA, Foroughi AA, Ahmadi SM, Heshmati K. Effectiveness of acceptance and commitment therapy (ACT) on depression and sleep quality in painful diabetic neuropathy: a randomized clinical trial. Journal of Diabetes and Metabolic Disorders. 2020.

de Groot, M., de Keijser, J., Neeleman, J., Kerkhof, A., Nolen, W., & Burger, H. (2007). Cognitive behaviour therapy to prevent complicated grief among relatives and spouses bereaved by suicide: cluster randomised controlled trial. Bmj, 334(7601), 994.

de Groot, M., de Keijser, J., Neeleman, J., Kerkhof, A., Nolen, W., & Burger,H. (2007). Cognitive behaviour therapy to prevent complicated grief among relatives and spouses bereaved by suicide: cluster randomised controlled trial. BMJ (Clinical research ed.), 334(7601), 994. https://doi.org/10.1136/bmj.39161.457431.55

De Groot, M., Shubrook, J. H., Hornsby, W. G., Pillay, Y., Mather, K. J., Fitzpatrick, K., . . . Saha, C. (2019). Program ACTIVE II: Outcomes from a randomized, multistate community-based depression treatment for rural and urban adults with type 2 diabetes. Diabetes care, 42(7), 1185-1193. doi:10.2337/dc18-2400

De Jong M, Peeters F, Gard T, et al. A randomized controlled pilot study on mindfulness-based cognitive therapy for unipolar depression in patients with chronic pain. Journal of clinical psychiatry 2018; 79(1): 26-34.
De Jongh, A., Muris, P., Horst, G. T., Van Zuuren, F., Schoenmakers, N., & Makkes, P. (1995). One-session cognitive treatment of dental phobia: preparing dental phobics for treatment by restructuring negative cognitions. Behaviour Research and Therapy, 33(8), 947-954. doi:https://doi.org/10.1016/0005-7967(95)00027-U
Dekker RL, Moser DK, Peden AR, Lennie TA. Cognitive therapy improves three-month outcomes in hospitalized patients with heart failure. Journal of cardiac failure. 2012;18(1):10-20.
Demir, S., & Ercan, F. (2022). The effectiveness of cognitive behavioral therapy-based group counseling on depressive symptomatology, anxiety levels, automatic thoughts, and coping ways Turkish nursing students: A randomized controlled trial. Perspectives in psychiatric care. doi:10.1111/ppc.13073
Dennis CL, Grigoriadis S, Zupancic J, Kiss A, Ravitz P. Telephone-based nurse-delivered interpersonal psychotherapy for postpartum depression: nationwide randomised controlled trial. Br J Psychiatry. 2020;216(4):189-196.
DeRubeis RJ, Hollon SD, Amsterdam JD, Shelton RC, Young PR, Salomon RM, et al. Cognitive therapy vs medications in the treatment of moderate to severe depression. Archives of General Psychiatry. 2005;62(4):409-16.
Desautels, C., Savard, J., Ivers, H., Savard, M. H., & Caplette-Gingras, A. (2018). Treatment of depressive symptoms in patients with breast cancer: a randomized controlled trial comparing cognitive therapy and bright light therapy [Journal Article; Randomized Controlled Trial]. Health Psychology, 37(1), 1-13. https://doi.org/10.1037/hea0000539
Dickinson D, Ph D, Tenhula W, Ph D, Morris S, Ph D, et al. A randomized , controlled trial of computer-assisted cognitive remediation for schizophrenia. Am J Psychiatry. 2010;167:170, 80.
Dimidjian S, Hollon SD, Dobson KS, Schmaling KB, Kohlenberg RJ, Addis ME, et al. Randomized trial of behavioral activation, cognitive therapy, and antidepressant medication in the acute treatment of adults with major depression. Journal of Consulting and Clinical Psychology. 2006;74(4):658-70.
Dimidjian, S., Goodman, S., Sherwood, N., Simon, G., Ludman, E., Gallop, R., . . . Beck, A. (2017). A Pragmatic Randomized Clinical Trial of Behavioral Activation for Depressed Pregnant Women. Journal of Consulting and Clinical Psychology, 85(1), 26-36
Dindo, L. N., Recober, A., Calarge, C. A., Zimmerman, B. M., Weinrib, A., Marchman, J. N., & Turvey, C. (2019). One-Day Acceptance and Commitment Therapy Compared to Support for Depressed Migraine Patients: a Randomized Clinical Trial. Neurotherapeutics. doi:10.1007/s13311-019-00818-0
Dixon-Gordon, K. L., Chapman, A. L., & Turner, B. J. (2015). A preliminary pilot study comparing dialectical behavior therapy emotion regulation skills with interpersonal effectiveness skills and a control group treatment. Journal of Experimental Psychopathology, 6(4), 369-388.
Dobkin RD, Mann SL, Gara MA, Interian A, Rodriguez KM, Menza M. Telephone-based cognitive behavioral therapy for depression in Parkinson disease: A randomized controlled trial. Neurology. 2020;94(16):e1764-e1773.
Dobkin RD, Menza M, Allen LA, Gara MA, Mark MH, Tiu J, et al. Cognitive-behavioral therapy for depression in Parkinson's disease: A randomized, controlled trial. American Journal of Psychiatry. 2011;168(10):1066-74.
Dobkin, R. D., Mann, S. L., Weintraub, D., Rodriguez, K. M., Miller, R. B., St. Hill, L., King, A., Gara, M. A., & Interian, A. (2021). Innovating Parkinson's Care: A Randomized Controlled Trial of Telemedicine Depression Treatment [Article]. Movement disorders, 36(11), 2549-2558. https://doi.org/10.1002/mds.28548
Doering LV, Chen B, Cross Bodan R, Magsarili MC, Nyamathi A, Irwin MR. Early cognitive behavioral therapy for depression after cardiac surgery. Journal of Cardiovascular Nursing. 2013;28(4):370-9.
Doering, S., Hörz, S., Rentrop, M., Fischer-Kern, M., Schuster, P., Benecke, C., ... & Buchheim, P. (2010). Transference-focused psychotherapy v. treatment by community psychotherapists for borderline personality disorder: randomised controlled trial. The British Journal of Psychiatry, 196(5), 389-395.

Dominick, S. A., Irvine, A. B., Beauchamp, N., Seeley, J. R., Nolen-Hoeksema, S., Doka, K. J., & Bonanno, G. A. (2010). An internet tool to normalize grief. OMEGA-Journal of Death and Dying, 60(1), 71-87.

Dong, X., Sun, G., Zhan, J., Liu, F., Ma, S., Li, P., . . . Liu, Y. (2019). Telephone-based reminiscence therapy for colorectal cancer patients undergoing postoperative chemotherapy complicated with depression: a three-arm randomised controlled trial. Supportive care in cancer : official journal of the Multinational Association of Supportive Care in Cancer, 27(8), 2761-2769. doi:10.1007/s00520-018-4566-6

Dowling, N., Smith, D., & Thomas, T. (2007). A comparison of individual and group cognitive-behavioural treatment for female pathological gambling. Behaviour Research and Therapy, 45(9), 2192-2202.

Dowrick, C., Dunn, G., Ayuso-Mateos, J. L., Dalgard, O. S., Page, H., Lehtinen, V., ... Wilkinson, G. (2000). Problem solving treatment and group psychoeducation for depression: multicentre randomised controlled trial Outcomes of Depression International Network [ODIN] Group. Bmj, 321(7274), 1450-1454. Retrieved from http://onlinelibrary.wiley.com/o/cochrane/clcentral/articles/836/CN-00623836/frame.html

Duarte PS, Miyazaki MC, Blay SL, Sesso R. Cognitive-behavioral group therapy is an effective treatment for major depression in hemodialysis patients. Kidney international. 2009;76(4):414–21.

Ducasse, D., Dassa, D., Courtet, P., Brand-Arpon, V., Walter, A., Guillaume, S., Jaussent, I., & Olié, E. (2019). Gratitude diary for the management of suicidal inpatients: A randomized controlled trial. Depression and anxiety, 36(5), 400–411. https://doi.org/10.1002/da.22877

Duffy M, Gillespie K, Clark DM. Post-traumatic stress disorder in the context of terrorism and other civil conflict in Northern Ireland: randomised controlled trial. BMJ. 2007 Jun 2;334(7604):1147-50. doi: 10.1136/bmj.39021.846852.BE. PMID: 17495988.

Dugas, M. J., Brillon, P., Savard, P., Turcotte, J., Gaudet, A., Ladouceur, R., . . . Gervais, N. J. (2010). A randomized clinical trial of cognitive-behavioral therapy and applied relaxation for adults with generalized anxiety disorder. Behav Ther, 41(1), 46-58. doi:10.1016/j.beth.2008.12.004

Dugas, M. J., Brillon, P., Savard, P., Turcotte, J., Gaudet, A., Ladouceur, R., . . . Gervais, N. J. (2010). A randomized clinical trial of cognitive-behavioral therapy and applied relaxation for adults with generalized anxiety disorder. Behav Ther, 41(1), 46-58. doi:10.1016/j.beth.2008.12.004

Dugas, M. J., Ladouceur, R., Leger, E., Freeston, M. H., Langlois, F., Provencher, M. D., & Boisvert, J. M. (2003). Group cognitive-behavioral therapy for generalized anxiety disorder: treatment outcome and long-term follow-up. J Consult Clin Psychol, 71(4), 821-825.

Dugas, M. J., Ladouceur, R., Leger, E., Freeston, M. H., Langlois, F., Provencher, M. D., & Boisvert, J. M. (2003). Group cogni-tive-behavioral therapy for generalized anxiety disorder: treatment outcome and long-term follow-up. J Consult Clin Psy-chol, 71(4), 821-825.

Dugas, M. J., Sexton, K. A., Hebert, E. A., Bouchard, S., Gouin, J.-P., & Shafran, R. (2022). Behavioral experiments for intolerance of uncertainty: A randomized clinical trial for adults with generalized anxiety disorder. Behavior Therapy, 53(6), 1147-1160. doi:10.1016/j.beth.2022.05.003

Dugas, M. J., Sexton, K. A., Hebert, E. A., Bouchard, S., Gouin, J.-P., & Shafran, R. (2022). Behavioral experiments for intoler-ance of uncertainty: A randomized clinical trial for adults with generalized anxiety disorder. Behavior Therapy, 53(6), 1147-1160. doi:10.1016/j.beth.2022.05.003

Dunn NJ, Rehm LP, Schillaci J, Souchek J, Mehta P, Ashton CM, et al. A randomized trial of self-management and psychoeducational group therapies for comorbid chronic posttraumatic stress disorder and depressive disorder. Journal of traumatic stress. 2007;20(3):221-37.

Dunne RL, Kenardy JA, Sterling M. A randomized controlled trial of cognitive-behavioral therapy for the treatment of PTSD in the context of chronic whiplash. Clin J Pain. 2012 Nov-Dec;28(9):755-65. doi: 10.1097/AJP.0b013e318243e16b. PMID: 22209798.

Durham RC, Guthrie A, Morton RV, Reid DA, Treliving LR, Owler DF, et al. Tayside-Fife clinical trial of cognitive- behavioural therapy for medication-resistant psychotic symptoms; Results to 3-month follow-up. Br J Psychiatry. 2003;182:303, 12.

Dwight-Johnson M, Aisenberg E, Golinelli D, Hong S, O'Brien M, Ludman E. Telephone-based cognitive-behavioral therapy for Latino patients living in rural areas: A randomized pilot study. Psychiatric Services. 2011;62(8):936-42.

Ebert DD, Buntrock C, Lehr D, et al. Effectiveness of Web- and Mobile-Based Treatment of Subthreshold Depression With Adherence-Focused Guidance: a Single-Blind Randomized Controlled Trial. Behavior therapy 2018; 49(1): 71-83.
Ebert, D. D., Lehr, D., Boß, L., Riper, H., Cuijpers, P., Andersson, G., . . . Berking, M. (2014). Efficacy of an internet-based problem-solving training for teachers: results of a randomized controlled trial. Scandinavian journal of work, environment & health, 582-596.
Ede MO, Igbo JN, Eseadi C, et al. Effect of group cognitive behavioural therapy on depressive symptoms in a sample of college adolescents in Nigeria. Journal of Rational-Emotive & Cognitive-Behavior Therapy. 2020;38(3):306-318.
Efendi F, Indarwati R, Aurizki GE. Effect of trauma-focused cognitive behavior therapy on depression and the quality of life of the elderly in Indonesia. Work Older People. 2020;24(3):149-57. doi: 10.1108/WWOP-02-2020-0004.
Ehlers A, Clark DM, Hackmann A, et al. A randomized controlled trial of cognitive therapy, a self-help booklet, and repeated assessments as early interventions for posttraumatic stress disorder. Arch Gen Psychiatry. 2003;60(10):1024-32. doi: 10.1001/archpsyc.60.10.1024. PMID: 14557148.
Ehlers A, Clark DM, Hackmann A, McManus F, Fennell M. Cognitive therapy for post-traumatic stress disorder: development and evaluation. Behav Res Ther. 2005 Apr;43(4):413-31. doi: 10.1016/j.brat.2004.03.006. PMID: 15701354.
Ehlers A, Hackmann A, Grey N, et al. A randomized controlled trial of 7-day intensive and standard weekly cognitive therapy for PTSD and emotion-focused supportive therapy. Am J Psychiatry. 2014 Mar;171(3):294-304. doi: 10.1176/appi.ajp.2013.13040552. PMID: 24480899.
Eisma, M. C., Boelen, P. A., van den Bout, J., Stroebe, W., Schut, H. A., Lancee, J., & Stroebe, M. S. (2015). Internet-based exposure and behavioral activation for complicated grief and rumination: A randomized controlled trial. Behavior therapy, 46(6), 729-748.
Ekers D, Richards D, McMillan D, Bland JM, Gilbody S. Behavioural activation delivered by the non-specialist: Phase II randomised controlled trial. The British Journal of Psychiatry. 2011;198(1):66-72.
Ekkers W, Korrelboom K, Huijbrechts I, Smits N, Cuijpers P, Gaag M. Competitive memory training for treating depression and rumination in depressed older adults: A randomized controlled trial. Behaviour Research and Therapy. 2011;49(10):588-96.
El-Haj-Mohamad R, Böttche M, Vöhringer M, Specht F, Stammel N, Nesterko Y, et al. An internet-based cognitive behavioural intervention for adults with depression in Arabic-speaking countries: A randomized controlled trial. Stress Health. 2024;40(5):e3432.
El-Haj-Mohamad R, Stein J, Stammel N, Nesterko Y, Wagner B, Böttche M, et al. Efficacy of internet-based cognitive behavioral and interpersonal treatment for depression in Arabic speaking countries: A randomized controlled trial. J Affect Disord. 2025;368:573-83.
El-Monshed, A. H., Khonji, L. M., Altheeb, M., Saad, M. T. E., Elsheikh, M. A., Loutfy, A., . . . Zoromba, M. A. (2024). Does a program-based cognitive behavioral therapy affect insomnia and depression in menopausal women? A randomized controlled trial. Worldviews Evid Based Nurs, 21(2), 202-215. doi:https://doi.org/10.1111/wvn.12707
ElBarazi A, Badary OA, Elmazar MM, et al. Cognitive Processing Therapy versus medication for the treatment of comorbid substance use disorder and post-traumatic stress disorder in Egyptian patients (randomized clinical trial). Journal of Evidence-Based Psychotherapies. 2022;22(2):63-90. doi: 10.24193/jebp.2022.2.13.
Elkin I, Shea MT, Watkins JT, Imber SD, Sotsky SM, Collins JF, et al. National institute of mental health treatment of depression collaborative research program. General effectiveness of treatments. Archives of General Psychiatry. 1989;46(11):971-82; discussion 83.
Embling S. The effectiveness of cognitive behavioural therapy in depression. Nursing Standard. 2002;17(14-15):33-41.
Engel CC, Litz B, Magruder KM, et al. Delivery of self training and education for stressful situations (DESTRESS-PC): a randomized trial of nurse assisted online self-management for PTSD in primary care. Gen Hosp Psychiatry. 2015 Jul-Aug;37(4):323-8. doi: 10.1016/j.genhosppsych.2015.04.007. PMID: 25929985.
England M. Efficacy of cognitive nursing intervention for voice hearing. Perspect Psychiatr Care. 2007;43(2):69, 76.

Erickson, D. H., Janeck, A. S., & Tallman, K. (2007). A cognitive-behavioral group for patients with various anxiety disorders. Psychiatric Services, 58(9), 1205-1211.
Erickson, D. H., Janeck, A. S., & Tallman, K. (2007). A cognitive-behavioral group for patients with various anxiety disorders. Psychiatric Services, 58(9), 1205-1211.
Ertl, V., Pfeiffer, A., Schauer, E., Elbert, T., & Neuner, F. (2011). Community-implemented trauma therapy for former child soldiers in Northern Uganda: a randomized controlled trial. JAMA, 306(5), 503–512. https://doi.org/10.1001/jama.2011.1060

Eseadi C, Obidoa MA, Ogbuabor SE, Ikechukwu-Ilomuanya AB. Effects of Group-Focused Cognitive-Behavioral Coaching Program on Depressive Symptoms in a Sample of Inmates in a Nigerian Prison. International journal of offender therapy and comparative criminology 2018; 62(6): 1589-602.

Eseadi, C., Ilechukwu, L. C., Victor-Aigbodion, V., Sewagegn, A. A., & Amedu, A. N. (2022). Intervention for depression among undergraduate religious education students: A randomized controlled trial. Medicine (Baltimore), 101(41), e31034. doi:10.1097/md.0000000000031034

Eskin, M., Ertekin, K. & Demir, H. Efficacy of a Problem-Solving Therapy for Depression and Suicide Potential in Adolescents and Young Adults. Cogn Ther Res 32, 227–245 (2008). https://doi.org/10.1007/s10608-007-9172-8

Euteneuer, F., Dannehl, K., Del Rey, A., Engler, H., Schedlowski, M., & Rief, W. (2017). Immunological effects of behavioral activation with exercise in major depression: An exploratory randomized controlled trial. Translational Psychiatry, 7(5).

Euteneuer, F., Neuert, M., Salzmann, S., Fischer, S., Ehlert, U., & Rief, W. (2022). Does psychological treatment of major depression reduce cardiac risk biomarkers? An exploratory randomized controlled trial. Psychological medicine, 1-15. doi:10.1017/S0033291722000447

Ewais, T., et al. (2021). "Mindfulness based cognitive therapy for youth with inflammatory bowel disease and depression - Findings from a pilot randomised controlled trial." J Psychosom Res 149: 110594.

Eylem, O., van Straten, A., Bhui, K., & Kerkhof, A. J. (2015). Protocol: Reducing suicidal ideation among Turkish migrants in the Netherlands and in the UK: effectiveness of an online intervention. International review of psychiatry (Abingdon, England), 27(1), 72–81. https://doi.org/10.3109/09540261.2014.996121

Fals-Stewart, W., & Schafer, J. (1992). The treatment of substance abusers diagnosed with obsessive-compulsive disorder: an outcome study. J Subst Abuse Treat, 9(4), 365-370.

Falsetti SA, Resnick HS, Davis JL. Multiple channel exposure therapy for women with PTSD and comorbid panic attacks. Cogn Behav Ther. 2008;37(2):117-30. doi: 10.1080/16506070801969088. PMID: 18470742.

Fann JR, Bombardier CH, Vannoy S, Dyer J, Ludman E, Dikmen S, et al. Telephone and in-person cognitive behavioral therapy for major depression after traumatic brain injury: A randomized controlled trial. Journal of neurotrauma. 2015;32(1):45-57.

Faramarzi M, Alipor A, Esmaelzadeh S, Kheirkhah F, Poladi K, Pash H. Treatment of depression and anxiety in infertile women: Cognitive behavioral therapy versus fluoxetine. Journal of Affective Disorders. 2008;108(1-2):159-64.

Fardig R, Lewander T, Melin L, Folke F, Fredriksson A. A randomized controlled trial of the Illness Management and Recovery Program for persons with schizophrenia. Psychiatr Serv. 2011;62(6):606ì12.

Farrell, J. M., Shaw, I. A., & Webber, M. A. (2009). A schema-focused approach to group psychotherapy for outpatients with borderline personality disorder: a randomized controlled trial. Journal of behavior therapy and experimental psychiatry, 40(2), 317-328.

Farreny A, Aguado J, Ochoa S, Huerta-ramos E, Marsa F, Lopez-carrilero R, et al. REPYFLEC cognitive remediation group training in schizophrenia Looking for an integrative approach. Schizophr Res [Internet]. Elsevier B.V.; 2012;142(1, 3):137, 44. Available from: http://dx.doi.org/10.1016/j.schres.2012.08.035

Fecteau GW, Nicki RM. Cognitive behavioural treatment of post traumatic stress disorder after motor vehicle accident. Behav. Cogn. Psychother. 1999 Jul;27(3):201-14.

Feigenbaum, J. D., Fonagy, P., Pilling, S., Jones, A., Wildgoose, A., & Bebbington, P. E. (2012). A real-world study of the effectiveness of DBT in the UK National Health Service. British Journal of Clinical Psychology, 51(2), 121-141.

Fereidouni, Z., Behnammoghadam, M., Jahanfar, A., & Dehghan, A. (2019). The Effect of Eye Movement Desensitization and Reprocessing (EMDR) on the severity of suicidal thoughts in patients with major depressive disorder: a randomized controlled trial. Neuropsychiatric disease and treatment, 2459-2466.

Fereydouni, S., & Forstmeier, S. (2022). An Islamic Form of Logotherapy in the Treatment of Depression, Anxiety and Stress Symptoms in University Students in Iran. J Relig Health, 61(1), 139-157. doi:10.1007/s10943-021-01495-0

Fernandez-Gonzalo S, Turon M, Jodar M, Pousa E, Hernandez C, Garcia R, et al. A new computerized cognitive and social cognition training specifically designed for patients with schizophrenia/ schizoaffective disorder in early stages of illness : A pilot study. Psychiatry Res. Elsevier Ireland Ltd; 2015;228:501, 9.

Feske U, Goldstein AJ. Eye movement desensitization and reprocessing treatment for panic disorder: a controlled outcome and partial dismantling study. Journal of consulting and clinical psychology 1997;65(6):1026-35.

Fissler, M., Winnebeck, E., Schroeter, T. A., Gummbersbach, M., Huntenburg, J. M., Gärtner, M., & Barnhofer, T. (2017). Brief training in mindfulness may normalize a blunted error-related negativity in chronically depressed patients. Cognitive, Affective & Behavioral Neuroscience, 17(6), 1164-1175. doi:10.3758/s13415-017-0540-x

Fiszdon JM, Choi KH, Bell MD, Choi J, Silverstein SM. Cognitive remediation for individuals with psychosis : efficacy and mechanisms of treatment effects. Psychol Med. 2016;46:3275, 89.

Fledderus M, Bohlmeijer ET, Pieterse ME, Schreurs KM. Acceptance and commitment therapy as guided self-help for psychological distress and positive mental health: A randomized controlled trial. Psychological Medicine. 2012;42(3):485-95.

Floyd M, Scogin F, McKendree-Smith NL, Floyd DL, Rokke PD. Cognitive therapy for depression: A comparison of individual psychotherapy and bibliotherapy for depressed older adults. Behavior modification. 2004;28(2):297-318.

Flygare AL, Engström I, Hasselgren M, et al. Internet-based CBT for patients with depressive disorders in primary and psychiatric care: Is it effective and does comorbidity affect outcome? Internet Interventions. 2020;19.

Foa EB, Dancu CV, Hembree EA, et al. A comparison of exposure therapy, stress inoculation training, and their combination for reducing posttraumatic stress disorder in female assault victims. J Consult Clin Psychol. 1999;67(2):194. PMID: 10224729.

Foa EB, Hembree EA, Cahill SP, et al. Randomized trial of prolonged exposure for posttraumatic stress disorder with and without cognitive restructuring: outcome at academic and community clinics. J Consult Clin Psychol. 2005;73(5):953. doi: 10.1037/0022-006X.73.5.953. PMID: 16287395.

Foa EB, McLean CP, Zang Y, et al. Effect of prolonged exposure therapy delivered over 2 weeks vs 8 weeks vs present-centered therapy on PTSD symptom severity in military personnel: a randomized clinical trial. JAMA. 2018 Jan 23;319(4):354-64. doi: 10.1001/jama.2017.21242. PMID: 29362795.

Foa EB, Rothbaum BO, Riggs DS, et al. Treatment of posttraumatic stress disorder in rape victims: a comparison between cognitive-behavioral procedures and counseling. J Consult Clin Psychol. 1991 Oct;59(5):715-23. PMID: 1955605.

Foa, E. B., Blau, J. S., Prout, M., & Latimer, P. (1977). Is horror a necessary component of flooding (implosion)? Behaviour Research and Therapy, 15(5), 397-402. doi:https://doi.org/10.1016/0005-7967(77)90043-2

Foa, E. B., Liebowitz, M. R., Kozak, M. J., Davies, S., Campeas, R., Franklin, M. E., . . . Tu, X. (2005). Randomized, placebo-controlled trial of exposure and ritual prevention, clomipramine, and their combination in the treatment of obsessive-compulsive disorder. Am J Psychiatry, 162(1), 151-161.

Folke F, Parling T, Melin L. Acceptance and commitment therapy for depression: A preliminary randomized clinical trial for unemployed on long-term sick leave. Cognitive and Behavioral Practice. 2012;19(4):583-94.

Fonagy P, Lemma A, Target M, O'Keeffe S, Constantinou MP, Ventura Wurman T, et al. Dynamic interpersonal therapy for moderate to severe depression: a pilot randomized controlled and feasibility trial. Psychological medicine. 2019:1-10.

Fonagy P, Rost F, Carlyle JA, McPherson S, Thomas R, Pasco Fearon RM, et al. Pragmatic randomized controlled trial of long-term psychoanalytic psychotherapy for treatment-resistant depression: The Tavistock Adult Depression Study (TADS). World Psychiatry. 2015;14(3):312-21.

Fonzo GA, Goodkind MS, Oathes DJ, et al. PTSD psychotherapy outcome predicted by brain activation during emotional reactivity and regulation. Am J Psychiatry. 2017 Dec 1;174(12):1163-74. doi: 10.1176/appi.ajp.2017.16091072. PMID: 28715908.

Forand NR, Barnett JG, Strunk DR, Hindiyeh MU, Feinberg JE, Keefe JR. Efficacy of Guided iCBT for Depression and Mediation of Change by Cognitive Skill Acquisition. Behavior Therapy 2018; 49(2): 295-307.

Forman-Hoffman, V. L., Sihvonen, S., Wielgosz, J., Kuhn, E., Nelson, B. W., Peiper, N. C., & Gould, C. E. (2024). Therapist-supported digital mental health intervention for depressive symptoms: A randomized clinical trial. Journal of Affective Disorders, 349, 494-501. doi:https://doi.org/10.1016/j.jad.2024.01.057

Forsell E, Bendix M, Holländare F, et al. Internet delivered cognitive behavior therapy for antenatal depression: A randomised controlled trial. Journal of Affective Disorders 2017; 221: 56-64.

Frangou, E., Bertelli, G., Love, S., Mackean, M. J., Glasspool, R. M., Fotopoulou, C., Cook, A., Nicum, S., Lord, R., Ferguson, M., Roux, R. L., Martinez, M., Butcher, C., Hulbert-Williams, N., Howells, L., & Blagden, S. P. (2021). OVPSYCH2: A randomized controlled trial of psychological support versus standard of care following chemotherapy for ovarian cancer. Gynecol Oncol, 162(2), 431-439. https://doi.org/10.1016/j.ygyno.2021.05.024

Franklin CL, Cuccurullo LA, Walton JL, et al. Face to face but not in the same place: a pilot study of prolonged exposure therapy. J Trauma Dissociation. 2017 Jan-Feb;18(1):116-30. doi: 10.1080/15299732.2016.1205704. PMID: 27348462.

Freedland KE, Carney RM, Rich MW, Steinmeyer BC, Rubin EH. Cognitive behavior therapy for depression and self-care in heart failure patients: A randomized clinical trial. JAMA Internal Medicine. 2015;175(11):1773-82.

Freedland KE, Skala JA, Carney RM, Rubin EH, Lustman PJ, D·vila-Rom·n VG, et al. Treatment of depression after coronary artery bypass surgery: A randomized controlled trial. Archives of General Psychiatry. 2009;66(4):387-96.

Freeman 2015a: Freeman D, Dunn G, Startup H, Pugh K, Cordwell J, Mander H, et al. Effects of cognitive behaviour therapy for worry on persecutory delusions in patients with psychosis (WIT): a parallel, single-blind , randomised controlled trial with a mediation analysis. Lancet Psychiatry. 2015;2(April 2015):305, 13.

Freeman 2015b: Freeman D, Waite F, Startup H, Myers E, Lister R, Mcinerney J, et al. Efficacy of cognitive behavioural therapy for sleep improvement in patients with persistent delusions and hallucinations (BEST): a prospective , assessor-blind , randomised controlled pilot trial. Lancet Psychiatry. 2015;2:975, 83.

Freeman, D., Haselton, P., Freeman, J., Spanlang, B., Kishore, S., Albery, E., . . . Nickless, A. (2018). Automated psychological therapy using immersive virtual reality for treatment of fear of heights: a single-blind, parallel-group, randomised controlled trial. Lancet Psychiatry, 5(8), 625-632. doi:10.1016/s2215-0366(18)30226-8

Freeston, M. H., Ladouceur, R., Gagnon, F., Thibodeau, N., Rhéaume, J., Letarte, H., & Bujold, A. (1997). Cognitive-behavioral treatment of obsessive thoughts: a controlled study. J Consult Clin Psychol, 65(3), 405-413.

Fry P. Structured and unstructured reminiscence training and depression among the elderly. Clinical Gerontologist. 1983;1(3):15-37.

Fuhr, D. C., Weobong, B., Lazarus, A., Vanobberghen, F., Weiss, H. A., Singla, D. R., . . . Patel, V. (2019). Delivering the Thinking Healthy Programme for perinatal depression through peers: An individually randomised controlled trial in India. The Lancet Psychiatry, 6(2), 115-127. doi:10.1016/S2215-0366(18)30466-8

Funderburk, J. S., Pigeon, W. R., Shepardson, R. L., Wade, M., Acker, J., Fivecoat, H., Wray, L. O., & Maisto, S. A. (2021). Treating depressive symptoms among veterans in primary care: A multi-site RCT of brief behavioral activation. J Affect Disord, 283, 11-19. https://doi.org/10.1016/j.jad.2021.01.033

Furmark, T., Carlbring, P., Hedman, E., Sonnenstein, A., Clevberger, P., Bohman, B., Eriksson, A., Hallen, A., Frykman, M., Holmstrom, A., Sparthan, E., Tillfors, M., Ihrfelt, E. N., Spak, M., Eriksson, A., Ekselius, L., & Andersson, G. (2009). Guided and unguided self-help for social anxiety disorder: randomised controlled trial. Br J Psychiatry, 195(5), 440-447. https://doi.org/10.1192/bjp.bp.108.060996

Furukawa TA, Horikoshi M, Kawakami N, Kadota M, Sasaki M, Sekiya Y, et al. Telephone cognitive-behavioral therapy for subthreshold depression and presenteeism in workplace: A randomized controlled trial. PLoS One. 2012;7(4):e35330.

Garcia S, Fuentes I, RuiÃÅz JC, Gallach E, Roder V. Application of the IPT in a Spanish sample : Evaluation of the ‚ÄúSocial Perception Subprogramme.‚Äù Int J Psychol Psychol Ther. 2003;3(2):299, 310.

Garcia-Palacios, A., Hoffman, H., Carlin, A., Furness, T. A., 3rd, & Botella, C. (2002). Virtual reality in the treatment of spider phobia: a controlled study. Behav Res Ther, 40(9), 983-993. doi:10.1016/s0005-7967(01)00068-7

García-Peña C, Vázquez-Estupiñan F, Avalos-Pérez F, Jiménez LVR, Sánchez-Garcia S, Juárez-Cedillo T. Clinical effectiveness of group cognitive-behavioural therapy for depressed older people in primary care: A randomised controlled trial. Salud Mental. 2015;38(1):33-9.

Garcia, A., Yáñez, A. M., Bennasar-Veny, M., Navarro, C., Salva, J., Ibarra, O., . . . Garcia-Toro, M. (2023). Efficacy of an adjuvant non-face-to-face multimodal lifestyle modification program for patients with treatment-resistant major depression: A randomized controlled trial. Psychiatry Res, 319, 114975. doi:10.1016/j.psychres.2022.114975

García, J. A., Landa, V., Grandes, G., Pombo, H., & Mauriz, A. (2013). Effectiveness of "primary bereavement care" for widows: a cluster randomized controlled trial involving family physicians. Death Studies, 37(4), 287-310.

Garety 2008 (i) & (ii): Garety PA, Fowler DG, Freeman D, Bebbington P, Dunn G, Kuipers E. Cognitive , behavioural therapy and family intervention for relapse prevention and symptom reduction in psychosis : randomised controlled trial. Br J Psychiatry. 2008;192:412, 23.

Garety 2008 (i) & (ii): Garety PA, Fowler DG, Freeman D, Bebbington P, Dunn G, Kuipers E. Cognitive , behavioural therapy and family intervention for relapse prevention and symptom reduction in psychosis : randomised controlled trial. Br J Psychiatry. 2008;192:412, 23.

Gawrysiak M, Nicholas C, Hopko DR. Behavioral activation for moderately depressed university students: Randomized controlled trial. Journal of Counseling Psychology. 2009;56(3):468-75.

Gellis ZD, Bruce ML. Problem solving therapy for subthreshold depression in home healthcare patients with cardiovascular disease. The American Journal of Geriatric Psychiatry. 2010;18(6):464-74.

Gellis ZD, McGinty J, Tierney L, Jordan C, Burton J, Misener E. Randomized controlled trial of problem-solving therapy for minor depression in home care. Research on Social Work Practice. 2008;18(6):596-606.

Gensichen, J., et al. (2019). "Panic Disorder in Primary Care: The Effects of a Team-Based Intervention." 116(10): 159-166.

Geraedts AS, Kleiboer AM, Wiezer NM, van Mechelen W, Cuijpers P. Short-term effects of a web-based guided self-help intervention for employees with depressive symptoms: Randomized controlled trial. Journal of Medical Internet Research. 2014;16(5):e121.

Gersons BP, Carlier IV, Lamberts RD, et al. Randomized clinical trial of brief eclectic psychotherapy for police officers with posttraumatic stress disorder. J Trauma Stress. 2000 Apr;13(2):333-47. doi: 10.1023/A:1007793803627. PMID: 10838679..

Ghahramanlou-Holloway, M., Lee-Tauler, S. Y., LaCroix, J. M., Kauten, R., Perera, K., Chen, R., Weaver, J., & Soumoff, A. (2018). Dysfunctional personality disorder beliefs and lifetime suicide attempts among psychiatrically hospitalized military personnel. Comprehensive psychiatry, 82, 108–114. https://doi.org/10.1016/j.comppsych.2018.01.010

Ghorbani, V., Zanjani, Z., Omidi, A., & Sarvizadeh, M. (2021). Efficacy of acceptance and commitment therapy (ACT) on depression, pain acceptance, and psychological flexibility in married women with breast cancer: a pre- and post-test clinical trial. Trends Psychiatry Psychother, 43(2), 126-133. https://doi.org/10.47626/2237-6089-2020-0022

Gibbons MB, Thompson SM, Scott K, Schauble LA, Mooney T, Thompson D, et al. Supportive-expressive dynamic psychotherapy in the community mental health system: A pilot effectiveness trial for the treatment of depression. Psychotherapy (Chicago, Ill). 2012;49(3):303-16.

Gilbody, S., Littlewood, E., McMillan, D., Atha, L., Bailey, D., Baird, K., . . . Ekers, D. (2024). Behavioural activation to mitigate the psychological impacts of COVID-19 restrictions on older people in England and Wales (BASIL+): a pragmatic randomised controlled trial. The Lancet Healthy Longevity, 5(2), e97-e107. doi:https://doi.org/10.1016/s2666-7568(23)00238-6

GilSanz DG, Lorenzo MD, Seco RB, Rodriguez MA, Martinez IL, Calleja RS, et al. Efficacy of a social cognition training program for schizophrenic patients : A Pilot Study. Span J Psychol. 2009;12(1):184, 91.

Gitlin LN, Harris LF, McCoy MC, Chernett NL, Pizzi LT, Jutkowitz E, et al. A home-based intervention to reduce depressive symptoms and improve quality of life in older African Americans: A randomized trial. Annals of Internal Medicine. 2013;159(4):243-52.

Gloster AT, Sonntag R, Hoyer J, et al. Treating treatment-resistant patients with panic disorder and agoraphobia using psychotherapy: A randomized controlled switching trial. Psychotherapy and Psychosomatics 2015;84(2):100-09.

Gloster AT, Wittchen HU, Einsle F, et al. Psychological treatment for panic disorder with agoraphobia: a randomized controlled trial to examine the role of therapist-guided exposure in situ in CBT. Journal of consulting and clinical psychology 2011;79(3):406-20.

Gold SM, Friede T, Meyer B, Moss-Morris R, Hudson J, Asseyer S, et al. Internet-delivered cognitive behavioural therapy programme to reduce depressive symptoms in patients with multiple sclerosis: a multicentre, randomised, controlled, phase 3 trial. The Lancet Digital Health. 2023;5(10):e668-e78.

Goldin, P. R., Morrison, A., Jazaieri, H., Brozovich, F., Heimberg, R., & Gross, J. J. (2016). Group CBT versus MBSR for social anxiety disorder: A randomized controlled trial. J Consult Clin Psychol, 84(5), 427-437. https://doi.org/10.1037/ccp0000092

Goldin, P. R., Ziv, M., Jazaieri, H., Werner, K., Kraemer, H., Heimberg, R. G., & Gross, J. J. (2012). Cognitive reappraisal self-efficacy mediates the effects of individual cognitive-behavioral therapy for social anxiety disorder. J Consult Clin Psychol, 80(6), 1034-1040. https://doi.org/10.1037/a0028555

Goldstein AJ, De Beurs E, Chambless DL, Wilson KA. EMDR for panic disorder with agoraphobia: comparison with waiting list and credible attention-placebo control conditions. Journal of Consulting and Clinical Psychology 2000;68(6):947–56.

Gomes, J. B., Cordioli, A. V., Bortoncello, C. F., Braga, D. T., Gonçalves, F., & Heldt, E. (2016). Impact of cognitive-behavioral group therapy for obsessive-compulsive disorder on family accommodation: A randomized clinical trial. Psychiatry Res, 246, 70-76.

Goodkin, K., Blaney, N. T., Feaster, D. J., Baldewicz, T., Burkhalter, J. E., & Leeds, B. (1999). A randomized controlled clinical trial of a bereavement support group intervention in human immunodeficiency virus type 1–seropositive and–seronegative homosexual men. Archives of General Psychiatry, 56(1), 52-59.

Goodman JH, Prager J, Goldstein R, Freeman M. Perinatal Dyadic Psychotherapy for postpartum depression: A randomized controlled pilot trial. Archives of Women's Mental Health. 2015;18(3):493-506.

Goodman, M., Sullivan, S. R., Spears, A. P., Crasta, D., Mitchell, E. L., Stanley, B., ... & Glynn, S. (2022). A pilot randomized control trial of a dyadic safety planning intervention: Safe actions for families to encourage recovery. Couple and Family Psychology: Research and Practice, 11(1), 42.

Gould RA, Clum GA, Shapiro D. The use of bibliotherapy in the treatment of panic: A preliminary investigation. Behavior Therapy 1993;24(2):241-52.

Grant, J. E., Donahue, C. B., Odlaug, B. L., Kim, S. W., Miller, M. J., & Petry, N. M. (2009). Imaginal desensitisation plus motivational interviewing for pathological gambling: randomised controlled trial. The British Journal of Psychiatry, 195(3), 266-267.

Gray R, Budden-Potts D, Bourke F. Reconsolidation of traumatic memories for PTSD: a randomized controlled trial of 74 male veterans. Psychother Res. 2019 29(5): 621-639. DOI: 10.1080/10503307.2017.1408973

Greenberg J, Datta T, Shapero BG, Sevinc G, Mischoulon D, Lazar SW. Compassionate hearts protect against wandering minds: Self-compassion moderates the effect of mind-wandering on depression. Spirituality in Clinical Practice 2018; 5(3): 155-69.

Gregory, R. J., Chlebowski, S., Kang, D., Remen, A. L., Soderberg, M. G., Stepkovitch, J., & Virk, S. (2008). A controlled trial of psychodynamic psychotherapy for co-occurring borderline personality disorder and alcohol use disorder. Psychotherapy: Theory, Research, Practice, Training, 45(1), 28.

Greist, J. H., Marks, I. M., Baer, L., Kobak, K. A., Wenzel, K. W., Hirsch, M. J., . . . Clary, C. M. (2002). Behavior therapy for obsessive-compulsive disorder guided by a computer or by a clinician compared with relaxation as a control. J Clin Psychiatry, 63(2), 138-145.

Griegel LE. Breathing retraining in panic disorder: physiological mechanisms or perceived controllability. Dissertation Abstracts International: Section B: The Sciences and Engineering 1995;55(9):4120.

Grote NK, Swartz HA, Geibel SL, Zuckoff A, Houck PR, Frank E. A randomized controlled trial of culturally relevant, brief interpersonal psychotherapy for perinatal depression. Psychiatric Services. 2009;60(3):313-21.

Gruber, K., Moran, P. J., Roth, W. T., & Taylor, C. B. (2001). Computer-assisted cognitive behavioral group therapy for social phobia. Behavior Therapy, 32(1), 155-165. https://www.cochranelibrary.com/central/doi/10.1002/central/CN-00395686/full

Gujjar, K. R., van Wijk, A., Kumar, R., & de Jongh, A. (2019). Efficacy of virtual reality exposure therapy for the treatment of dental phobia in adults: A randomized controlled trial. J Anxiety Disord, 62, 100-108. doi:10.1016/j.janxdis.2018.12.001

Gumley A, O'Grady M, McNay L, Reilly J, Power K, Norrie J. Early intervention for relapse in schizophrenia : results of a 12-month randomized controlled trial of cognitive behavioural therapy. Psychol Med. 2003;33:419, 31.

Guo X, Zhai J, Liu Z, Fang M, Wang B, Wang C, et al. Antipsychotic medication alone versus combined with psychosocial intervention on outcomes of early stage schizophrenia: a randomized, one-year study. Arch Gen Psychiatry. 2010;67(9):895, 904.

Gureje, O., Oladeji, B. D., Kola, L., Bello, T., Ayinde, O., Faregh, N., . . . Zelkowitz, P. (2022). Effect of intervention delivered by frontline maternal care providers to improve outcome and parenting skills among adolescents with perinatal depression in Nigeria (the RAPiD study): A cluster randomized controlled trial. J Affect Disord, 312, 169-176. doi:10.1016/j.jad.2022.06.032

Guthrie, E., Kapur, N., Mackway-Jones, K., Chew-Graham, C., Moorey, J., Mendel, E., Marino-Francis, F., Sanderson, S., Turpin, C., Boddy, G., & Tomenson, B. (2001). Randomised controlled trial of brief psychological intervention after deliberate self poisoning. BMJ (Clinical research ed.), 323(7305), 135–138. https://doi.org/10.1136/bmj.323.7305.135

Haakana, R., Rosenström, T., Parkkinen, L., Tuomisto, M. T., & Isometsä, E. (2024). Effectiveness of an add-on brief group behavioral activation treatment for depression in psychiatric care: a randomized clinical trial. Frontiers in Psychiatry, 15. doi:https://doi.org/10.3389/fpsyt.2024.1284363

Haddock, G., Pratt, D., Gooding, P. A., Peters, S., Emsley, R., Evans, E., Kelly, J., Huggett, C., Munro, A., Harris, K., Davies, L., & Awenat, Y. (2019). Feasibility and acceptability of suicide prevention therapy on acute psychiatric wards: randomised controlled trial. BJPsych open, 5(1), e14. https://doi.org/10.1192/bjo.2018.85

Hagen, R., Hjemdal, O., Solem, S., Kennair, L. E. O., Nordahl, H. M., Fisher, P., & Wells, A. (2017). Metacognitive therapy for depression in adults: A waiting list randomized controlled trial with six months follow-up. Frontiers in Psychology, 8.

Hahm, H. C., Zhou, L., Lee, C., Maru, M., Petersen, J. M., & Kolaczyk, E. D. (2019). Feasibility, preliminary efficacy, and safety of a randomized clinical trial for Asian Women's Action for Resilience and Empowerment (AWARE) intervention. The American journal of orthopsychiatry, 89(4), 462–474. https://doi.org/10.1037/ort0000383

Hallford DJ, Mellor D. Autobiographical memory-based intervention for depressive symptoms in young adults: A randomized controlled trial of cognitive-reminiscence therapy. Psychotherapy and Psychosomatics. 2016;85(4):246-9.

Hallgren M, Kraepelien M, Öjehagen A, Lindefors N, Zeebari Z, Kaldo V, et al. Physical exercise and internet-based cognitive-behavioural therapy in the treatment of depression: Randomised controlled trial. British Journal of Psychiatry. 2015;207(3):227-34.

Hamamci Z. Integrating psychodrama and cognitive behavioral therapy to treat moderate depression. Arts in Psychotherapy. 2006;33(3):199-207.
Hamdan-Mansour AM, Puskar K, Bandak AG. Effectiveness of cognitive-behavioral therapy on depressive symptomatology, stress and coping strategies among Jordanian university students. Issues in mental health nursing. 2009;30(3):188-96.
Haringsma R, Engels G, Cuijpers P, Spinhoven P. Effectiveness of the Coping With Depression (CWD) course for older adults provided by the community-based mental health care system in the Netherlands: A randomized controlled field trial. International Psychogeriatrics. 2006;18(02):307-25.
Harley R, Sprich S, Safren S, Jacobo M, Fava M. Adaptation of dialectical behavior therapy skills training group for treatment-resistant depression. Journal of Nervous and Mental Disease. 2008;196(2):136-43.
Harrer, M., Apolinário-Hagen, J., Fritsche, L., Salewski, C., Zarski, A. C., Lehr, D., Baumeister, H., Cuijpers, P., & Ebert, D. D. (2021). Effect of an internet- and app-based stress intervention compared to online psychoeducation in university students with depressive symptoms: Results of a randomized controlled trial [Article]. Internet Interventions, 24. https://doi.org/10.1016/j.invent.2021.100374
Harris, N., & Mazmanian, D. (2016). Cognitive behavioural group therapy for problem gamblers who gamble over the internet: A controlled study. Journal of Gambling Issues, 33, 170-188.
Hashemi, Z., Eyni, S., & Ebadi, M. (2022). Effectiveness of Acceptance and Commitment Therapy in Depression and Anxiety in People with Substance Use Disorder. Iranian Journal of Psychiatry and Behavioral Sciences, 16(1). doi:10.5812/ijpbs.110135
Hassiotis A, Serfaty M, Azam K, Strydom A, Blizard R, Romeo R, et al. Manualised individual cognitive behavioural therapy for mood disorders in people with mild to moderate intellectual disability: A feasibility randomised controlled trial. Journal of Affective Disorders. 2013;151(1):186-95.
Hatcher, S., Sharon, C., Parag, V., & Collins, N. (2011). Problem-solving therapy for people who present to hospital with self-harm: Zelen randomised controlled trial. The British journal of psychiatry : the journal of mental science, 199(4), 310–316. https://doi.org/10.1192/bjp.bp.110.090126
Haukebø, K., Skaret, E., Ost, L. G., Raadal, M., Berg, E., Sundberg, H., & Kvale, G. (2008). One- vs. five-session treatment of dental phobia: a randomized controlled study. J Behav Ther Exp Psychiatry, 39(3), 381-390. doi:10.1016/j.jbtep.2007.09.006
Hautzinger M, Welz S. Kognitive Verhaltenstherapie bei Depressionen im Alter: Ergebnisse einer kontrollierten Vergleichsstudie unter ambulanten Bedingungen an Depressionen mittleren Schweregrads. = Cognitive behavioral therapy for depressed older outpatients: A controlled, randomized trial. Zeitschrift für Gerontologie und Geriatrie. 2004;37(6):427-35.
Hayati, M. S., Shams, J., Meibodi, S. S., Shafighi, A. H., Shattell, M., & Ghadirian, F. (2024). The effects of a telenursing scheduled intervention of brief behavioral activation therapy on depression and anxiety symptoms of patients with mixed depression and anxiety disorder: A randomized controlled trial. Arch Psychiatr Nurs, 52, 39-44. doi:https://doi.org/10.1016/j.apnu.2024.07.014
Hayman PM, Cope CS. Effects of assertion training on depression. Journal of clinical psychology. 1980;36(2):534-43.
Hazen AL,Walker JR, Eldridge GD. Anxiety sensitivity and treatment outcome in panic disorder. Anxiety 1996;2(1):34–9.
He, H. L., Zhang, M., Gu, C. Z., Xue, R. R., Liu, H. X., Gao, C. F., & Duan, H. F. (2019). Effect of Cognitive Behavioral Therapy on Improving the Cognitive Function in Major and Minor Depression. The Journal of nervous and mental disease, 207(4), 232-238.
He, L., Han, W., & Shi, Z. (2021). The Effects of Mindfulness-Based Stress Reduction on Negative Self-Representations in Social Anxiety Disorder-A Randomized Wait-List Controlled Trial. Front Psychiatry, 12, 582333. https://doi.org/10.3389/fpsyt.2021.582333
Heading, K., Kirkby, K. C., Martin, F., Daniels, B. A., Gilroy, L. J., & Menzies, R. G. (2001). Controlled Comparison of Single-session Treatments for Spider Phobia: Live Graded Exposure Alone versus Computer-aided Vicarious Exposure. Behaviour Change, 18(2), 103-113. doi:10.1375/bech.18.2.103
Heckman TG, Sikkema KJ, Hansen N, Kochman A, Heh V, Neufeld S, et al. A randomized clinical trial of a coping improvement group intervention for HIV-infected older adults. Journal of behavioral medicine. 2011;34(2):102-11.

Heckman, T. G., Heckman, B. D., Anderson, T., Lovejoy, T. I., Markowitz, J. C., Shen, Y., & Sutton, M. (2017). Tele-interpersonal psychotherapy acutely reduces depressive symptoms in depressed HIV-infected rural persons: A randomized clinical trial. Behavioral Medicine, 43(4), 285-295.
Heckman, T. G., Heckman, B. D., Anderson, T., Lovejoy, T. I., Mohr, D., Sutton, M., . . . Gau, J.-T. (2013). Supportive-expressive and coping group teletherapies for HIV-infected older adults: a randomized clinical trial. AIDS Behav, 17(9), 3034-3044.
Hegerl U, Hautzinger M, Mergl R, Kohnen R, Sch¸tze M, Scheunemann W, et al. Effects of pharmacotherapy and psychotherapy in depressed primary-care patients: A randomized, controlled trial including a patients' choice arm. International Journal of Neuropsychopharmacology. 2010;13(1):31-44.
Heim, E., Ramia, J. A., Hana, R. A., Burchert, S., Carswell, K., Cornelisz, I., . . . van't Hof, E. (2021). Step-by-step: Feasibility randomised controlled trial of a mobile-based intervention for depression among populations affected by adversity in Lebanon. Internet Interventions, 24, 100380. doi:https://doi.org/10.1016/j.invent.2021.100380
Heimberg, R. G., Liebowitz, M. R., Hope, D. A., Schneier, F. R., Holt, C. S., Welkowitz, L. A., Juster, H. R., Campeas, R., Bruch, M. A., Cloitre, M., Fallon, B., & Klein, D. F. (1998). Cognitive behavioral group therapy vs phenelzine therapy for social phobia: 12-week outcome. Arch Gen Psychiatry, 55(12), 1133-1141. https://doi.org/10.1001/archpsyc.55.12.1133
Hekmat, H. (1973). Systematic versus semantic desensitization and implosive therapy: A comparative study. Journal of Consulting and Clinical Psychology, 40, 202-209. doi:10.1037/h0034552
Hemanny, C., Carvalho, C., Maia, N., Reis, D., Botelho, A. C., Bonavides, D., . . . De Oliveira, I. R. (2019). Efficacy of trial-based cognitive therapy, behavioral activation and treatment as usual in the treatment of major depressive disorder: Preliminary findings from a randomized clinical trial. CNS Spectrums. doi:10.1017/S1092852919001457
Hendriks GJ, Keijsers GP, Kampman M, et al. A randomized controlled study of paroxetine and cognitive-behavioural therapy for late-life panic disorder. Acta psychiatrica Scandinavica 2010;122(1):11-9.
Herbst, N., Voderholzer, U., Thiel, N., Schaub, R., Knaevelsrud, C., Stracke, S., . . . Külz, A. K. (2014). No talking, just writing! Efficacy of an Internet-based cognitive behavioral therapy with exposure and response prevention in obsessive compulsive disorder. Psychother Psychosom, 83(3), 165-175.
Hermanns N, Schmitt A, Gahr A, Herder C, Nowotny B, Roden M, et al. The effect of a diabetes-specific cognitive behavioral treatment program (DIAMOS) for patients with diabetes and subclinical depression: Results of a randomized controlled trial. Diabetes care. 2015;38(4):551-60.
Herpertz, S. C., Matzke, B., Hillmann, K., Neukel, C., Mancke, F., Jaentsch, B., ... & Dempfle, A. (2021). A mechanism-based group-psychotherapy approach to aggressive behaviour in borderline personality disorder: findings from a cluster-randomised controlled trial. BJPsych open, 7(1), e17.
Herrmann-Lingen C, Beutel ME, Bosbach A, Deter HC, Fritzsche K, Hellmich M, et al. A stepwise psychotherapy intervention for reducing risk in coronary artery disease (SPIRR-CAD): Results of an observer-blinded, multicenter, randomized trial in depressed patients with coronary artery disease. Psychosomatic Medicine. 2016;78(6):704-15.
Heshmati R, Wienicke FJ, Driessen E. The effects of intensive short-term dynamic psychotherapy on depressive symptoms, negative affect, and emotional repression in single treatment-resistant depression: A randomized controlled trial. Psychotherapy. 2023;60(4):497-511.
Hilden, H. M., Rosenström, T., Karila, I., Elokorpi, A., Torpo, M., Arajärvi, R., & Isometsä, E. (2021). Effectiveness of brief schema group therapy for borderline personality disorder symptoms: a randomized pilot study. Nordic journal of psychiatry, 75(3), 176-185.
Himle, J. A., Bybee, D., Steinberger, E., Laviolette, W. T., Weaver, A., Vlnka, S., Golenberg, Z., Levine, D. S., Heimberg, R. G., & O'Donnell, L. A. (2014). Work-related CBT versus vocational services as usual for unemployed persons with social anxiety disorder: A randomized controlled pilot trial. Behav Res Ther, 63, 169-176. https://doi.org/10.1016/j.brat.2014.10.005
Hinton DE, Hofmann SG, Pollack MH, et al. Mechanisms of efficacy of CBT for Cambodian refugees with PTSD: improvement in emotion regulation and orthostatic blood pressure response. CNS Neurosci Ther. 2009 Fall;15(3):255-63. doi: 10.1111/j.1755-5949.2009.00100.x. PMID: 19691545.
Hodgins, D. C., Currie, S. R., & el-Guebaly, N. (2001). Motivational enhancement and self-help treatments for problem gambling. Journal of Consulting and Clinical Psychology, 69(1), 50-57.

Hodgins, D. C., Currie, S. R., Currie, G., & Fick, G. H. (2009). Randomized trial of brief motivational treatments for pathological gamblers: More is not necessarily better. Journal of Consulting and Clinical Psychology, 77(5), 950-960.

Hogberg G, Pagani M, Sundin O, et al. On treatment with eye movement desensitization and reprocessing of chronic post-traumatic stress disorder in public transportation workers--a randomized controlled trial. Nord J Psychiatry. 2007;61(1):54-61. doi: 10.1080/08039480601129408. PMID: 17365790.

Hoifodt RS, Lillevoll KR, Griffiths KM, et al. The clinical effectiveness of web-based cognitive behavioral therapy with face-to-face therapist support for depressed primary care patients: randomized controlled trial. Journal of medical Internet research 2013; 15(8): e153.

Holden JM, Sagovsky R, Cox JL. Counselling in a general practice setting: Controlled study of heath visitor intervention in treatment of postnatal depression. British Medical Journal. 1989;298(6668):223-6.

Hollifield M, Sinclair-Lian N, Warner TD, et al. Acupuncture for posttraumatic stress disorder: a randomized controlled pilot trial. J Nerv Ment Dis. 2007 Jun;195(6):504-13. doi: 10.1097/NMD.0b013e31803044f8. PMID: 17568299.

Honey KL, Bennett P, Morgan M. A brief psycho-educational group intervention for postnatal depression. British Journal of Clinical Psychology. 2002;41(4):405-9.

Hope, D. A., Heimberg, R. G., & Bruch, M. A. (1995). Dismantling cognitive-behavioral group therapy for social phobia. Behav Res Ther, 33(6), 637-650. https://doi.org/10.1016/0005-7967(95)00013-n

Horrell L, Goldsmith KA, Tylee AT, Schmidt UH, Murphy CL, Bonin E-M, et al. One-day cognitive–behavioural therapy self-confidence workshops for people with depression: Randomised controlled trial. The British Journal of Psychiatry. 2014;204(3):222-33.

Hou Y, Hu P, Zhang Y, Lu Q, Wang D, Yin L, et al. Cognitive behavioral therapy in combination with systemic family therapy improves mild to moderate postpartum depression. Revista Brasileira de Psiquiatria. 2014;36(1):47-52.

Howard, R., Cort, E., Rawlinson, C., Wiegand, M., Downey, A., Lawrence, V., . . . Gould, R. (2024). Adapted problem adaptation therapy for depression in mild to moderate Alzheimer's disease dementia: A randomized controlled trial. Alzheimers Dement, 20(4), 2990-2999. doi:https://doi.org/10.1002/alz.13766

Hoyer, J., Beesdo, K., Gloster, A. T., Runge, J., Hofler, M., & Becker, E. S. (2009). Worry exposure versus applied relaxation in the treatment of generalized anxiety disorder. Psychother Psychosom, 78(2), 106-115. doi:10.1159/000201936

Hoyer, J., Beesdo, K., Gloster, A. T., Runge, J., Hofler, M., & Becker, E. S. (2009). Worry exposure versus applied relaxation in the treatment of generalized anxiety disorder. Psychother Psychosom, 78(2), 106-115. doi:10.1159/000201936

Hsiao, F.-H., Lai, Y.-M., Chen, Y.-T., Yang, T.-T., Liao, S.-C., Ho, R. T., Ng, S.-M., Chan, C. L., & Jow, G.-M. (2014). Efficacy of psychotherapy on diurnal cortisol patterns and suicidal ideation in adjustment disorder with depressed mood. General hospital psychiatry, 36(2), 214-219. http://ac.els-cdn.com/S0163834313003113/1-s2.0-S0163834313003113-main.pdf?_tid=685fbc62-cf4e-11e5-b7e9-00000aab0f6c&acdnat=1455037317_a8c762fea010c7ac31f2152769dad94c

Huang, C.-Y., Lai, H.-L., Chen, C.-I., Lu, Y.-C., Li, S.-C., Wang, L.-W., & Su, Y. (2016). Effects of motivational enhancement therapy plus cognitive behaviour therapy on depressive symptoms and health-related quality of life in adults with type II diabetes mellitus: A randomised controlled trial. Quality of Life Research: An International Journal of Quality of Life Aspects of Treatment, Care & Rehabilitation, 25(5), 1275-1283.

Huh, K., Layton, H., Savoy, C. D., Ferro, M. A., Bieling, P. J., Hicks, A., & Van Lieshout, R. J. (2023). Online Public Health Nurse-Delivered Group Cognitive Behavioral Therapy for Postpartum Depression: A Randomized Controlled Trial During the COVID-19 Pandemic. J Clin Psychiatry, 84(5).

Hui, C., & Zhihui, Y. (2017). Group cognitive behavioral therapy targeting intolerance of uncertainty: a randomized trial for older Chinese adults with generalized anxiety disorder. Aging Ment Health, 21(12), 1294-1302. doi:10.1080/13607863.2016.1222349

Hui, C., & Zhihui, Y. (2017). Group cognitive behavioral therapy targeting intolerance of uncertainty: a randomized trial for older Chinese adults with generalized anxiety disorder. Aging Ment Health, 21(12), 1294-1302. doi:10.1080/13607863.2016.1222349

Hum, K. M., Chan, C. J., Gane, J., Conway, L., McAndrews, M. P., & Smith, M. L. (2019). Do distance-delivery group interventions improve depression in people with epilepsy? Epilepsy & Behavior, 98, 153-160. doi:10.1016/j.yebeh.2019.06.037

Hummel, J., Weisbrod, C., Boesch, L., Himpler, K., Hauer, K., Hautzinger, M., . . . Kopf, D. (2017). AIDE–Acute Illness and Depression in Elderly Patients. Cognitive Behavioral Group Psychotherapy in Geriatric Patients With Comorbid Depression: A Randomized, Controlled Trial. Journal of the american medical directors association, 18(4), 341-349.

Hunter SB, Watkins KE, Hepner KA, Paddock SM, Ewing BA, Osilla KC, et al. Treating depression and substance use: A randomized controlled trial. Journal of substance abuse treatment. 2012;43(2):137-51.

Hurtado-Santiago, S., Guzmán-Parra, J., Mayoral, F., & Bersabé, R. M. (2022). Iconic Therapy for the reduction of borderline personality disorder symptoms among suicidal youth: a preliminary study. BMC psychiatry, 22(1), 1-11.

Husain N, Lunat F, Lovell K, Miah J, Chew-Graham CA, Bee P, et al. Efficacy of a culturally adapted, cognitive behavioural therapy-based intervention for postnatal depression in British south Asian women (ROSHNI-2): a multicentre, randomised controlled trial. Lancet. 2024;404(10461):1430-43.

Husain N, Lunat F, Lovell K, Sharma D, Zaidi N, Bokhari A, et al. Exploratory RCT of a group psychological intervention for postnatal depression in British mothers of South Asian origin - ROSHNI-D. Acta Psychol (Amst). 2023;238:103974.

Husain, N., Afsar, S., Ara, J., Fayyaz, H., Rahman, R. U., Tomenson, B., Hamirani, M., Chaudhry, N., Fatima, B., Husain, M., Naeem, F., & Chaudhry,B. (2014). Brief psychological intervention after self-harm: randomised controlled trial from Pakistan. The British journal of psychiatry : the journal of mental science, 204(6), 462–470. https://doi.org/10.1192/bjp.bp.113.138370

Husain, N., Kiran, T., Fatima, B., Chaudhry, I. B., Husain, M., Shah, S., Bassett, P., Cohen, N., Jafri, F., Naeem, S., Zadeh, Z., Roberts, C., Rahman, A., Naeem, F., Husain, M. I., & Chaudhry, N. (2021). An integrated parenting intervention for maternal depression and child development in a low-resource setting: Cluster randomized controlled trial [Article]. Depress Anxiety, 38(9), 925-939. https://doi.org/10.1002/da.23169

Husain, N., Kiran, T., Shah, S., Rahman, A., Raza Ur, R., Saeed, Q., Naeem, S., Bassett, P., Husain, M., Haq, S. U., Jaffery, F., Cohen, N., Naeem, F., & Chaudhry, N. (2021). Efficacy of learning through play plus intervention to reduce maternal depression in women with malnourished children: A randomized controlled trial from Pakistan(☆). J Affect Disord, 278, 78-84. https://doi.org/10.1016/j.jad.2020.09.001

Husain, N., Zulqernain, F., Carter, L.-A., Chaudhry, I., Fatima, B., Kiran, T., . . . Rahman, A. (2017). Treatment of maternal depression in urban slums of Karachi, Pakistan: a randomized controlled trial (RCT) of an integrated maternal psychological and early child development intervention. Asian journal of psychiatry, 29, 63-70.

Interian, A., Chesin, M. S., Stanley, B., Latorre, M., Hill, L. M. S., Miller, R. B., ... & Kline, A. (2021). Mindfulness-based cognitive therapy for preventing suicide in military veterans: a randomized clinical trial. The Journal of Clinical Psychiatry, 82(5), 36479.

Ito LM, de Araujo LA, Tess VL, et al. Self-exposure therapy for panic disorder with agoraphobia: randomised controlled study of external v. interoceptive self-exposure. The British journal of psychiatry : the journal of mental science 2001;178:331-6.

Ivanova, E., Lindner, P., Ly, K. H., Dahlin, M., Vernmark, K., Andersson, G., & Carlbring, P. (2016). Guided and unguided Acceptance and Commitment Therapy for social anxiety disorder and/or panic disorder provided via the Internet and a smartphone application: A randomized controlled trial. J Anxiety Disord, 44, 27-35. https://doi.org/10.1016/j.janxdis.2016.09.012

Ivarsson D, Blom M, Hesser H, et al. Guided internet-delivered cognitive behavior therapy for post-traumatic stress disorder: a randomized controlled trial. Internet Interv. 2014 March;1(1):33-40. doi: 10.1016/j.invent.2014.03.002.

Jacob N, Neuner F, Maedl A, et al. Dissemination of psychotherapy for trauma spectrum disorders in postconflict settings: a randomized controlled trial in Rwanda. Psychother Psychosom. 2014 Nov;83(6):354-63. doi: 10.1159/000365114. PMID: 25323203.

Jain S, Ortigo K, Gimeno J, Baldor DA, Weiss BJ, Cloitre M. A Randomized Controlled Trial of Brief Skills Training in Affective and Interpersonal Regulation (STAIR) for Veterans in Primary Care. J Trauma Stress. 2020 Aug;33(4):401-409. doi: 10.1002/jts.22523. Epub 2020 Jun 7. PMID: 32506563.

Jalali, F., Hasani, A., Hashemi, S. F., Kimiaei, S. A., & Babaei, A. (2019). Cognitive Group Therapy Based on Schema-Focused Approach for Reducing Depression in Prisoners Living With HIV. International journal of offender therapy and comparative criminology, 63(2), 276-288. doi:10.1177/0306624X18784185

Jamison C, Scogin F. The outcome of cognitive bibliotherapy with depressed adults. Journal of Consulting and Clinical Psychology. 1995;63(4):644-50.

Jamshidi, F., Rajabi, S., & Dehghani, Y. (2021). How to heal their psychological wounds? effectiveness of EMDR therapy on post-traumatic stress symptoms, mind-wandering and suicidal ideation in Iranian child abuse victims. Counselling and Psychotherapy Research, 21(2), 412-421.

Janssen NP, Lucassen P, Huibers MJH, Ekers D, Broekman T, Bosmans JE, et al. Behavioural Activation versus Treatment as Usual for Depressed Older Adults in Primary Care: A Pragmatic Cluster-Randomised Controlled Trial. Psychotherapy and Psychosomatics. 2023.

Jarero I, Schnaider S, Givaudan M. Randomized controlled trial: Provision of EMDR protocol for recent critical incidents and ongoing traumatic stress to first responders. Journal of EMDR Practice and Research. 2019;13(2):100-10. doi: 10.1891/1933-3196.13.2.100.

Jarrett RB, Schaffer M, McIntire D, Witt-Browder A, Kraft D, Risser RC. Treatment of atypical depression with cognitive therapy or phenelzine: A double-blind, placebo-controlled trial. Archives of General Psychiatry. 1999;56(5):431-7.

Jelinek L, Hauschildt M, Wittekind CE, Schneider BC, Kriston L, Moritz S. Efficacy of metacognitive training for depression: A randomized controlled trial. Psychotherapy and Psychosomatics. 2016;85(4):231-4.

Jenner JA, Nienhuis FJ, Wiersma D, van de Willige G. Hallucination focused integrative treatment : A randomized controlled trial. Schizophr Bull. 2004;30(1):133, 46.

Jensen JA. An investigation of Eye Movement Desensitization Reprocessing (EMD/R) as a treatment for posttraumatic stress disorder (PTSD) symptoms of Vietnam combat veterans. Behav Ther. 1994 Spring;25(2):311-25. doi: 10.1016/S0005-7894(05)80290-4.

Jensen, A. M., & Ramasamy, A. (2009). Treating spider phobia using Neuro Emotional Technique: findings from a pilot study. J Altern Complement Med, 15(12), 1363-1374. doi:10.1089/acm.2008.0595

Jesse DE, Gaynes BN, Feldhousen EB, Newton ER, Bunch S, Hollon SD. Performance of a culturally tailored cognitive-behavioral intervention integrated in a public health setting to reduce risk of antepartum depression: A randomized controlled trial. Journal of Midwifery and Women's Health. 2015;60(5):578-92.

Jessup, S. C., Tomarken, A., Viar-Paxton, M. A., & Olatunji, B. O. (2020). Effects of repeated exposure to fearful and disgusting stimuli on fear renewal in blood-injection-injury phobia. J Anxiety Disord, 74, 102272. doi:10.1016/j.janxdis.2020.102272

Jiang L, Wang ZZ, Qiu LR, Wan GB, Lin Y, Wei Z. Psychological intervention for postpartum depression. Journal of Huazhong University of Science and Technology: Medical sciences. 2014;34(3):437-42.

Jiang, M. Y. W., Upton, E., & Newby, J. M. (2020). A randomised wait-list controlled pilot trial of one-session virtual reality exposure therapy for blood-injection-injury phobias. J Affect Disord, 276, 636-645. doi:10.1016/j.jad.2020.07.076

Jobes, D. A., Comtois, K. A., Gutierrez, P. M., Brenner, L. A., Huh, D., Chalker, S. A., Ruhe, G., Kerbrat, A. H., Atkins, D. C., Jennings, K., Crumlish, J., Corona, C. D., Connor, S. O., Hendricks, K. E., Schembari, B., Singer, B., & Crow, B. (2017). A Randomized Controlled Trial of the Collaborative Assessment and Management of Suicidality versus Enhanced Care as Usual With Suicidal Soldiers. Psychiatry, 80(4), 339–356. https://doi.org/10.1080/00332747.2017.1354607

Johannsen, M., Schlander, C., Farver-Vestergaard, I., Lundorff, M., Wellnitz, K. B., Komischke-Konnerup, K. B., & O'Connor, M. (2022). Group-based compassion-focused therapy for prolonged grief symptoms in adults–Results from a randomized controlled trial. Psychiatry Research, 314, 114683.

Johansson P, Westas M, Andersson G, Alehagen U, Broström A, Jaarsma T, Mourad G, Lundgren J An Internet-Based Cognitive Behavioral Therapy Program Adapted to Patients With Cardiovascular Disease and Depression: Randomized Controlled Trial JMIR Ment Health 2019;6(10):e14648

Johansson R, Ekbladh S, Hebert A, Lindström M, Möller S, Petitt E, et al. Psychodynamic guided self-help for adult depression through the internet: A randomised controlled trial. PloS One. 2012;7(5):e38021.

Johansson R, Sjöberg E, Sjögren M, Johnsson E, Carlbring P, Andersson T, et al. Tailored vs. standardized internet-based cognitive behavior therapy for depression and comorbid symptoms: A randomized controlled trial. PloS One. 2012;7(5):e36905.

Johansson, O., Bjärehed, J., Andersson, G., Carlbring, P., & Lundh, L. G. (2019). Effectiveness of guided internet-delivered cognitive behavior therapy for depression in routine psychiatry: A randomized controlled trial. Internet Interventions, 17. doi:10.1016/j.invent.2019.100247

Johansson, R., Hesslow, T., Ljotsson, B., Jansson, A., Jonsson, L., Fardig, S., Karlsson, J., Hesser, H., Frederick, R. J., Lilliengren, P., Carlbring, P., & Andersson, G. (2017). Internet-based affect-focused psychodynamic therapy for social anxiety disorder: A randomized controlled trial with 2-year follow-up. Psychotherapy (Chic), 54(4), 351-360. https://doi.org/10.1037/pst0000147

Johnson DM, Zlotnick C, Perez SK. Cognitive behavioral treatment of PTSD in residents of battered women's shelters: results of a randomized clinical trial. J Consult Clin Psychol. 2011 Aug;79(4):542-51. doi: 10.1037/a0023822. PMID: 21787052.

Johnson JE, Zlotnick C. Pilot study of treatment for major depression among women prisoners with substance use disorder. Journal of Psychiatric Research. 2012;46(9):1174-83.

Johnson, J. E., Stout, R. L., Miller, T. R., Zlotnick, C., Cerbo, L. A., Andrade,J. T., Nargiso, J., Bonner, J., & Wiltsey-Stirman, S. (2019). Randomized cost- effectiveness trial of group interpersonal psychotherapy (IPT) for prisoners with major depression. Journal of consulting and clinical psychology, 87(4), 392–406. https://doi.org/10.1037/ccp0000379

Johnson, J. E., Stout, R. L., Miller, T. R., Zlotnick, C., Cerbo, L. A., Andrade, J. T., . . . Wiltsey-Stirman, S. (2019). Randomized cost-effectiveness trial of group interpersonal psychotherapy (IPT) for prisoners with major depression. Journal of consulting and clinical psychology, 87(4), 392-406. doi:10.1037/ccp0000379

Joling KJ, Hout HP, van't Veer-Tazelaar PJ, Horst HE, Cuijpers P, Ven PM, et al. How effective is bibliotherapy for very old adults with subthreshold depression? A randomized controlled trial. American Journal of Geriatric Psychiatry. 2011;19(3):256-65.

Jonas, B., Leuschner, F., Eiling, A., Schoelen, C., Soellner, R., & Tossmann, P. (2020). Web-based intervention and email-counseling for problem gamblers: Results of a randomized controlled trial. Journal of Gambling Studies, 36, 1341-1358.

Jones, S. L., Hadjistavropoulos, H. D., & Soucy, J. N. (2016). A randomized controlled trial of guided internet-delivered cognitive behaviour therapy for older adults with generalized anxiety. J Anxiety Disord, 37, 1-9. doi:10.1016/j.janxdis.2015.10.006

Jones, S. L., Hadjistavropoulos, H. D., & Soucy, J. N. (2016). A randomized controlled trial of guided internet-delivered cognitive behaviour therapy for older adults with generalized anxiety. J Anxiety Disord, 37, 1-9. doi:10.1016/j.janxdis.2015.10.006

Jordans, M. J. D., Luitel, N. P., Garman, E., Kohrt, B. A., Rathod, S. D., Shrestha, P., . . . Patel, V. (2019). Effectiveness of psychological treatments for depression and alcohol use disorder delivered by community-based counsellors: Two pragmatic randomised controlled trials within primary healthcare in Nepal. British Journal of Psychiatry, 215(2), 485-493. doi:10.1192/bjp.2018.300

Jorgensen R, Licht RW, Lysaker PH, Munk-Jorgensen P, Buck KD, Jensen SOW, et al. Effects on cognitive and clinical insight with the use of Guided Self-Determination in outpatients with schizophrenia : A randomized open trial. Eur Psychiatry. 2015;30:655, 63.

Kaiser, J., Nagl, M., Hoffmann, R., Linde, K., & Kersting, A. (2022). Therapist-assisted web-based intervention for prolonged grief disorder after cancer bereavement: Randomized controlled trial. JMIR mental health, 9(2), e27642.

Käll A, Bäck M, Fahlroth O, Ekeflod E, Lundberg A, Viberg N, et al. Internet-based therapist-supported interpersonal psychotherapy for depression: A randomized controlled trial. J Affect Disord. 2025;369:188-94.

Kamga, H., McCusker, J., Yaffe, M., Sewitch, M., Sussman, T., Strumpf, E., . . . Freeman, E. (2017). Self-care tools to treat depressive symptoms in patients with age-related eye disease: a randomized controlled clinical trial. Clinical & experimental ophthalmology, 45(4), 371-378.

Kampmann, I. L., Emmelkamp, P. M., Hartanto, D., Brinkman, W. P., Zijlstra, B. J., & Morina, N. (2016). Exposure to virtual social interactions in the treatment of social anxiety disorder: A randomized controlled trial. Behav Res Ther, 77, 147-156. https://doi.org/10.1016/j.brat.2015.12.016

Kang R, Wu Y, Li Z, Jiang J, Gao Q, Yu Y, et al. Effect of community-based social skills training and Tai-Chi exercise on outcomes in patients with chronic schizophrenia : A randomized , one-year study. Psychopathology. 2016;49:345, 55.

Kanter JW, Santiago-Rivera AL, Santos MM, Nagy G, López M, Hurtado GD, et al. A randomized hybrid efficacy and effectiveness trial of behavioral activation for latinos with depression. Behavior Therapy. 2015;46(2):177-92.

Kantrowitz JT, Sharif Z, Medalia A, Keefe RSE, Harvey P, Bruder G, et al. A multicenter, rater-blinded, randomised controlled study of auditory processing-focused cognitive remediation combined with open-label Lurasidone in patients with schizphrenia and schizoaffective disorder. J Clin Psychiatry. 2016;77(6):799, 806.

Karatzias T, Brown M, Taggart L, et al. A mixed-methods, randomized controlled feasibility trial of Eye Movement Desensitization and Reprocessing (EMDR) plus Standard Care (SC) versus SC alone for DSM-5 Posttraumatic Stress Disorder (PTSD) in adults with intellectual disabilities. J Appl Res Intellect Disabil. 2019 Jul;32(4):806-18. doi: 10.1111/jar.12570. PMID: 30714684.

Kashdan, T. B., Adams, L., Read, J., & Hawk, L., Jr. (2012). Can a one-hour session of exposure treatment modulate startle response and reduce spider fears? Psychiatry Res, 196(1), 79-82. doi:10.1016/j.psychres.2011.12.002

Kaslow, N. J., Leiner, A. S., Reviere, S., Jackson, E., Bethea, K., Bhaju, J., Rhodes, M., Gantt, M. J., Senter, H., & Thompson, M. P. (2010). Suicidal, abused African American women's response to a culturally informed intervention. Journal of consulting and clinical psychology, 78(4), 449–458. https://doi.org/10.1037/a0019692

Kay-Lambkin, F. J., Baker, A. L., Lewin, T. J., & Carr, V. J. (2009). Computer-based psychological treatment for comorbid depression and problematic alcohol and/or cannabis use: A randomized controlled trial of clinical efficacy. Addiction, 104(3), 378-388. doi:10.1111/j.1360-0443.2008.02444.x

Keane TM, Fairbank JA, Caddell JM, et al. Implosive (flooding) therapy reduces symptoms of PTSD in Vietnam combat veterans. Behav Ther. 1989 Spring;20(2):245-60. doi: 10.1016/S0005-7894(89)80072-3.

Keefe RSE, Vinogradov S, Medalia A, Peter F, Caroff SN, Souza DCD, et al. Feasibility and pilot efficacy results from the multi-site Cognitive Remediation in the Schizophrenia Trials Network (CRSTN) Study. J Clin Psychiatry. 2012;73(7):1016, 22.

Keeley RD, Brody DS, Engel M, Burke BL, Nordstrom K, Moralez E, et al. Motivational interviewing improves depression outcome in primary care: A cluster randomized trial. Journal of Consulting and Clinical Psychology. 2016;84(11):993-1007.

Kelly JA, Murphy DA, Bahr GR, Kalichman SC, Morgan MG, Stevenson LY, et al. Outcome of cognitive-behavioral and support group brief therapies for depressed, HIV-infected persons. American Journal of Psychiatry. 1993;150(11):1679-86.

Kemmeren, L. L., van Schaik, A., Draisma, S., Kleiboer, A., Riper, H., & Smit, J. H. (2023). Effectiveness of Blended Cognitive Behavioral Therapy Versus Treatment as Usual for Depression in Routine Specialized Mental Healthcare: E-COMPARED Trial in the Netherlands. Cognitive Therapy and Research. doi:10.1007/s10608-023-10363-y

Kenardy JA, Dow MG, Johnston DW, et al. A comparison of delivery methods of cognitive-behavioral therapy for panic disorder: an international multicenter trial. Journal of consulting and clinical psychology 2003;71(6):1068-75.

Kent M, Davis MC, Stark SL, et al. A resilience-oriented treatment for posttraumatic stress disorder: results of a preliminary randomized clinical trial. J Trauma Stress. 2011 Oct;24(5):591-5. doi: 10.1002/jts.20685. PMID: 21898603.

Kenter, R. M. F., Cuijpers, P., Beekman, A., & van Straten, A. (2016). Effectiveness of a Web-based guided self-help intervention for outpatients with a depressive disorder: Short-term results from a randomized controlled trial. Journal of medical Internet research, 18(3).

Kessler D, Lewis G, Kaur S, Wiles N, King M, Weich S, et al. Therapist-delivered Internet psychotherapy for depression in primary care: A randomised controlled trial. Lancet. 2009;374(9690):628-34.

Khazraee, H., Bakhtiari, M., Kianimoghadam, A. S., & Ghorbanikhah, E. (2023). The Effectiveness of Mindful Hypnotherapy on Depression, Self-Compassion, and Psychological Inflexibility in Females with Major Depressive Disorder: A Single-Blind, Randomized Clinical Trial. Int J Clin Exp Hypn, 71(1), 63-78. doi:10.1080/00207144.2022.2160257

Khodarahimi, S. (2009). Satiation therapy and exposure response prevention in the treatment of obsessive compulsive disorder. Journal of Contemporary Psychotherapy, 39(3), 203-207.

Khoramnia, S., Bavafa, A., Jaberghaderi, N., Parvizifard, A., Foroughi, A., Ahmadi, M., & Amiri, S. (2020). The effectiveness of acceptance and commitment therapy for social anxiety disorder: A randomized clinical trial. Trends in Psychiatry and Psychotherapy, 42(1), 30-38. https://doi.org/10.1590/2237-6089-2019-0003

Khoshbooii, R., Hassan, S. A., Deylami, N., Muhamad, R., Engku Kamarudin, E. M., & Alareqe, N. A. (2021). Effects of Group and Individual Culturally Adapted Cognitive Behavioral Therapy on Depression and Sexual Satisfaction among Perimenopausal Women. Int J Environ Res Public Health, 18(14). https://doi.org/10.3390/ijerph18147711

Kim D, Choi J, Kim SH, Oh DH, Park S, Lee SH. A pilot study of brief Eye Movement Desensitization and Reprocessing(EMDR) for treatment of acute phase schizophrenia. Korean J Biol Psychiatry. 2010;17(2):94, 102.

Kim YH, Choi KS, Han K, Kim HW. A psychological intervention programme for patients with breast cancer under chemotherapy and at a high risk of depression: a randomised clinical trial. Journal of clinical nursing 2018; 27(3-4): 572-81.

Kim, H., Kim, B. H., Kim, M. K., Eom, H., & Kim, J. J. (2022). Alteration of resting-state functional connectivity network properties in patients with social anxiety disorder after virtual reality-based self-training [Article]. Frontiers in Psychiatry, 13. https://doi.org/10.3389/fpsyt.2022.959696

King M, Sibbald B, Ward E, Bower P, Lloyd M, Gabbay M, et al. Randomised controlled trial of non-directive counselling, cognitive-behaviour therapy and usual general practitioner care in the management of depression as well as mixed anxiety and depression in primary care. Health Technology Assessment. 2000;4(19):1-83.

Kip KE, Rosenzweig L, Hernandez DF, et al. Randomized controlled trial of Accelerated Resolution Therapy (ART) for symptoms of combat-related post-traumatic stress disorder (PTSD). Mil Med. 2013 Dec;178(12):1298-309. doi: 10.7205/MILMED-D-13-00298. PMID: 24306011.

Kirsch, I., Tennen, H., Wickless, C., Saccone, A. J., & Cody, S. (1983). The role of expectancy in fear reduction. Behavior Therapy, 14(4), 520-533. doi:https://doi.org/10.1016/S0005-7894(83)80075-6

Kivi M, Eriksson MCM, Hange D, Petersson E-L, Vernmark K, Johansson B, et al. Internet-based therapy for mild to moderate depression in Swedish primary care: Short term results from the PRIM-NET randomized controlled trial. Cognitive Behaviour Therapy. 2014;43(4):289-98.

Klein B, Richards JC, Austin DW. Efficacy of internet therapy for panic disorder. Journal of behavior therapy and experimental psychiatry 2006;37(3):213-38.

Klosko JS, Barlow DH, Tassinari R, Cerny JA. A comparison of alprazolam and behavior therapy in treatment of panic disorder. Journal of Consulting and Clinical Psychology 1990; 58(1):77–84.

Knaevelsrud C, Böttche M, Pietrzak RH, et al. Efficacy and feasibility of a therapist-guided internet-based intervention for older persons with childhood traumatization: a randomized controlled trial. Am J Geriatr Psychiatry. 2017 Aug;25(8):878-88. doi: 10.1016/j.jagp.2017.02.024. PMID: 28365000.

Knaevelsrud C, Brand J, Lange A, et al. Web-based psychotherapy for posttraumatic stress disorder in war-traumatized Arab patients: randomized controlled trial. J Med Internet Res. 2015 Mar 20;17(3):e71. doi: 10.2196/jmir.3582. PMID: 25799024.

Kobayashi, Y., Kanie, A., Nakagawa, A., Takebayashi, Y., Shinmei, I., Nakayama, N., . . . Mimura, M. (2020). An evaluation of family-based treatment for OCD in Japan: a pilot randomized controlled trial. Frontiers in Psychiatry, 10, 932.

Koch T, Ehring T, Liedl A. Effectiveness of a transdiagnostic group intervention to enhance emotion regulation in young Afghan refugees: A pilot randomized controlled study. Behav Res Ther. 2020 Jun 27;132:103689. doi: 10.1016/j.brat.2020.103689. Epub ahead of print. PMID: 32688046.

Kocovski, N. L., Fleming, J. E., Hawley, L. L., Huta, V., & Antony, M. M. (2013). Mindfulness and acceptance-based group therapy versus traditional cognitive behavioral group therapy for social anxiety disorder: a randomized controlled trial. Behav Res Ther, 51(12), 889-898. https://doi.org/10.1016/j.brat.2013.10.007

Koelen, J., Klein, A., Wolters, N., Bol, E., De Koning, L., Roetink, S., ... & Wiers, R. (2024). Web-Based, Human-Guided, or Computer-Guided Transdiagnostic Cognitive Behavioral Therapy in University Students With Anxiety and Depression: Randomized Controlled Trial. JMIR Mental Health, 11, e50503.

Koochaki M, Mahmoodi Z, Esmaelzadeh–Saeieh S, et al. Effects of cognitive-behavioral counseling on posttraumatic stress disorder in mothers with infants hospitalized at neonatal intensive care units: a randomized controlled trial. Iran J Psychiatry Behav Sci. 2018;12(4):e65159. doi: 10.5812/ijpbs.65159.

Koons, C. R., Robins, C. J., Tweed, J. L., Lynch, T. R., Gonzalez, A. M., Morse, J. Q., ... & Bastian, L. A. (2001). Efficacy of dialectical behavior therapy in women veterans with borderline personality disorder. Behavior therapy, 32(2), 371-390.

Koons, Cedar & Robins, Clive & Tweed, J. & Lynch, Thomas & Gonzalez, Alicia & Morse, Jennifer & Bishop, G. & Butterfield, Marian. (2001). Efficacy of dialectical behavior therapy in women veterans with borderline Personality disorder. Behavior Therapy. 32. 371-390. 10.1016/S0005-7894(01)80009-5.

Korrelboom K, Maarsingh M, Huijbrechts I. Competitive memory training (COMET) for treating low self-esteem in patients with depressive disorders: A randomized clinical trial. Depression and anxiety. 2012;29(2):102-10.

Korte J, Bohlmeijer ET, Cappeliez P, Smit F, Westerhof GJ. Life review therapy for older adults with moderate depressive symptomatology: A pragmatic randomized controlled trial. Psychological Medicine. 2012;42(6):1163-73.

Koszycki, D., Thake, J., Mavounza, C., Daoust, J. P., Taljaard, M., & Bradwejn, J. (2016). Preliminary Investigation of a Mindfulness-Based Intervention for Social Anxiety Disorder That Integrates Compassion Meditation and Mindful Exposure. J Altern Complement Med, 22(5), 363-374. https://doi.org/10.1089/acm.2015.0108

Kovac, S. H., & Range, L. M. (2000). Writing projects: Lessening undergraduates' unique suicidal bereavement. Suicide and Life-Threatening Behavior, 30(1), 50-60.

Krakow BJ, Hollifield M, Schrader R, et al. A controlled study of imagery rehearsal for chronic nightmares in sexual assault survivors with PTSD: a preliminary report. Journal of Traumatic Stress. 2000 Oct;13(4):589-609. doi: 10.1023/A:1007854015481. PMID: 11109233.

Kramer, J., Conijn, B., Oijevaar, P., & Riper, H. (2014). Effectiveness of a web-based solution-focused brief chat treatment for depressed adolescents and young adults: randomized controlled trial. J Med Internet Res, 16(5), e141. doi:10.2196/jmir.3261

Krämer, L. V., Grünzig, S. D., Baumeister, H., Ebert, D. D., & Bengel, J. (2021). Effectiveness of a Guided Web-Based Intervention to Reduce Depressive Symptoms before Outpatient Psychotherapy: A Pragmatic Randomized Controlled Trial. Psychother Psychosom, 90(4), 233-242. https://doi.org/10.1159/000515625

Kredlow, M. A., Szuhany, K. L., Lo, S., Xie, H., Gottlieb, J. D., Rosenberg, S. D., & Mueser, K. T. (2017). Cognitive behavioral therapy for posttraumatic stress disorder in individuals with severe mental illness and borderline personality disorder. Psychiatry Research, 249, 86-93.

Krijn, M., Emmelkamp, P. M., Biemond, R., de Wilde de Ligny, C., Schuemie, M. J., & van der Mast, C. A. (2004). Treatment of acrophobia in virtual reality: the role of immersion and presence. Behav Res Ther, 42(2), 229-239. doi:10.1016/s0005-7967(03)00139-6

Krupnick JL, Green BL, Amdur RL, et al. An internet-based writing intervention for PTSD in veterans: a feasibility and pilot effectiveness trial. Psychol Trauma. 2017 Jul;9(4):461-70. doi: 10.1037/tra0000176. PMID: 27607767.

Krupnick JL, Green BL, Stockton P, et al. Group interpersonal psychotherapy for low-income women with posttraumatic stress disorder. Psychother Res. 2008 Sep;18(5):497-507. doi: 10.1080/10503300802183678. PMID: 18816001.

Kubany ES, Hill EE, Owens JA, et al. Cognitive trauma therapy for battered women with PTSD (CTT-BW). J Consult Clin Psychol. 2004 Feb;72(1):3-18. doi: 10.1037/0022-006x.72.1.3. PMID: 14756610.

Kuipers E, Garety PA, Fowler D, Dunn G, Bebbington P, Freeman D, et al. London-East Anglia randomised controlled trial of cognitive-behavioural therapy for psychosis. Br J Psychiatry. 1997;171:319, 27.

Kumar D, Zia M, Haq U, Dubey I, Dotivala KN, Siddiqui SV, et al. Effect of meta-cognitive training in the reduction of _positive symptoms in schizophrenia. Eur J Psychother Couns. 2010;12(2):149, 58.

Kyrios, M., Ahern, C., Fassnacht, D. B., Nedeljkovic, M., Moulding, R., & Meyer, D. (2018). Therapist-Assisted Internet-Based Cognitive Behavioral Therapy Versus Progressive Relaxation in Obsessive-Compulsive Disorder: Randomized Controlled Trial. J Med Internet Res, 20(8), e242.

LaBrie, R. A., Peller, A. J., LaPlante, D. A., Bernhard, B., Harper, A., Schrier, T., & Shaffer, H. J. (2012). A brief self-help toolkit intervention for gambling problems: a randomized multisite trial. American Journal of Orthopsychiatry, 82(2), 278-289.

LaCroix, J. M., Perera, K. U., Neely, L. L., Grammer, G., Weaver, J., & Ghahramanlou-Holloway, M. (2018). Pilot trial of post-admission cognitive therapy: Inpatient program for suicide prevention. Psychological services, 15(3), 279–288. https://doi.org/10.1037/ser0000224 100.Smits, M. L., Feenstra, D. J., Eeren, H. V., Bales, D. L., Laurenssen, E.,

Ladouceur, R., Dugas, M. J., Freeston, M. H., Leger, E., Gagnon, F., & Thibodeau, N. (2000). Efficacy of a cognitive-behavioral treatment for generalized anxiety disorder: evaluation in a controlled clinical trial. J Consult Clin Psychol, 68(6), 957-964.

Ladouceur, R., Dugas, M. J., Freeston, M. H., Leger, E., Gagnon, F., & Thibodeau, N. (2000). Efficacy of a cognitive-behavioral treatment for generalized anxiety disorder: evaluation in a controlled clinical trial. J Consult Clin Psychol, 68(6), 957-964.

Ladouceur, R., Sylvain, C., Boutin, C., Lachance, S., Doucet, C., Leblond, J., & Jacques, C. (2001). Cognitive treatment of pathological gambling. The Journal of Nervous and Mental Disease, 189(11), 774-780.

Ladouceur, R., Sylvain, C., Boutin, C., Lachance, S., Doucet, C., & Leblond, J. (2003). Group therapy for pathological gamblers: A cognitive approach. Behaviour Research and Therapy, 41(5), 587-596.

Laidlaw K, Davidson K, Toner H, Jackson G, Clark S, Law J, et al. A randomised controlled trial of cognitive behaviour therapy vs treatment as usual in the treatment of mild to moderate late life depression. International Journal of Geriatric Psychiatry. 2008;23(8):843-50.

Lamers F, Jonkers CC, Bosma H, Kempen GI, Meijer JA, Penninx BW, et al. A minimal psychological intervention in chronically ill elderly patients with depression: A randomized trial. Psychotherapy and Psychosomatics. 2010;79(4):217-26.

Lamers SMA, Bohlmeijer ET, Korte J, Westerhof GJ. The efficacy of life-review as online-guided self-help for adults: A randomized trial. The Journals of Gerontology: Series B: Psychological Sciences and Social Sciences 2015; 70B(1): 24-34.

Landreville P, Bissonnette L. Effects of cognitive bibliotherapy for depressed older adults with a disability. Clinical Gerontologist. 1997;17(4):35-55.

Lappalainen P, Langrial S, Oinas-Kukkonen H, Tolvanen A, Lappalainen R. Web-based acceptance and commitment therapy for depressive symptoms with minimal support: A randomized controlled trial. Behavior modification. 2015;39(6):805-34.

Larcombe NA, Wilson PH. An evaluation of cognitive-behaviour therapy for depression in patients with multiple sclerosis. The British Journal of Psychiatry. 1984;145:366-71.

Larimer, M. E., Neighbors, C., Lostutter, T. W., Whiteside, U., Cronce, J. M., Kaysen, D., & Walker, D. D. (2012). Brief motivational feedback and cognitive behavioral interventions for prevention of disordered gambling: A randomized clinical trial. Addiction, 107(6), 1148-1158.

Latif M, Husain MI, Gul M, et al. Culturally adapted trauma-focused CBT-based guided self-help (CatCBT GSH) for female victims of domestic violence in Pakistan: feasibility randomized controlled trial. Behav. 2021 Jan;49(1):50-61. doi: 10.1017/S1352465820000685. PMID: 32993831

Launes, G., Hagen, K., Sunde, T., Öst, L.-G., Klovning, I., Laukvik, I.-L., . . . Hansen, B. (2019). A randomized controlled trial of concentrated ERP, self-help and waiting list for obsessive-compulsive disorder: the Bergen 4-day treatment. Frontiers in Psychology, 10, 2500.

Laurenssen, E. M., Luyten, P., Kikkert, M. J., Westra, D., Peen, J., Soons, M. B., ... & Dekker, J. J. (2018). Day hospital mentalization-based treatment v. specialist treatment as usual in patients with borderline personality disorder: randomized controlled trial. Psychological medicine, 48(15), 2522-2529.

Leclerc C, Lesage AD, Ricard N, Lecomte T, Cyr M. Assessment of a new rehabilitative coping skills module for persons with schizophrenia. Am J Orthopsychiatry. 2000;70(3):380, 8.

Ledley, D. R., Heimberg, R. G., Hope, D. A., Hayes, S. A., Zaider, T. I., Dyke, M. V., Turk, C. L., Kraus, C., & Fresco, D. M. (2009). Efficacy of a manualized and workbook-driven individual treatment for social anxiety disorder. Behav Ther, 40(4), 414-424. https://doi.org/10.1016/j.beth.2008.12.001

Lee, B. K., & Awosoga, O. (2015). Congruence couple therapy for pathological gambling: A pilot randomized controlled trial. Journal of Gambling Studies, 31, 1047-1068.

Lee, E., Han, Y., Cha, Y. J., Oh, J. H., Hwang, N. R., Seo, H. J., & Choi, K. H. (2021). Community-Based Multi-Site Randomized Controlled Trial of Behavioral Activation for Patients with Depressive Disorders [Article in Press]. Community mental health journal. https://doi.org/10.1007/s10597-021-00828-3

Lee, W K. Effectiveness of computerized cognitive rehabilitation training on symptomatological, neuropsychological and work function in patients with schizophrenia. Asia-Pacific Psychiatry, 2013; 5: 90, 100.

Lehavot K, Millard SP, Thomas RM, Yantsides K, Upham M, Beckman K, Hamilton AB, Sadler A, Litz B, Simpson T. A randomized trial of an online, coach-assisted self-management PTSD intervention tailored for women veterans. J Consult Clin Psychol. 2021 Feb;89(2):134-142. doi: 10.1037/ccp0000556. PMID: 33705169; PMCID: PMC8238393.

Leichsenring, F., Salzer, S., Beutel, M. E., Herpertz, S., Hiller, W., Hoyer, J., Huesing, J., Joraschky, P., Nolting, B., Poehlmann, K., Ritter, V., Stangier, U., Strauss, B., Stuhldreher, N., Tefikow, S., Teismann, T., Willutzki, U., Wiltink, J., & Leibing, E. (2013). Psychodynamic therapy and cognitive-behavioral therapy in social anxiety disorder: a multicenter randomized controlled trial. Am J Psychiatry, 170(7), 759-767. https://doi.org/10.1176/appi.ajp.2013.12081125

Lemma A, Fonagy P. Feasibility study of a psychodynamic online group intervention for depression. Psychoanalytic Psychology. 2013;30(3):367-80.

Lenferink, L. I., de Keijser, J., Smid, G. E., & Boelen, P. A. (2020). Cognitive therapy and EMDR for reducing psychopathology in bereaved people after the MH17 plane crash: Findings from a randomized controlled trial. Traumatology, 26(4), 427.

Lenze, S., & Potts, M. (2017). Brief Interpersonal Psychotherapy for depression during pregnancy in a low-income population: a randomized controlled trial. Journal of Affective Disorders, 210, 151-157.

Leppänen, V., Hakko, H., Sintonen, H., & Lindeman, S. (2016). Comparing effectiveness of treatments for borderline personality disorder in communal mental health care: The Oulu BPD Study. Community mental health journal, 52, 216-227.

Lerner D, Adler DA, Rogers WH, Chang H, Greenhill A, Cymerman E, et al. A randomized clinical trial of a telephone depression intervention to reduce employee presenteeism and absenteeism. Psychiatric Services. 2015;66(6):570-7.

Leung S, Lee A, Wong D, Wong C, Leung K, Chiang V, et al. A brief group intervention using a cognitive-behavioural approach to reduce postnatal depressive symptoms: A randomised controlled trial. Hong Kong Medical Journal. 2016;22(1 Supplement 2).

Leung SS, Lee AM, Chiang VC, Lam SK, Kuen YW, Wong DF. Culturally sensitive, preventive antenatal group cognitive-behavioural therapy for Chinese women with depression. Internationan Journal of Nursing Practice. 2013;19(Supp 1):28-37.

Leutgeb, V., Schäfer, A., & Schienle, A. (2009). An event-related potential study on exposure therapy for patients suffering from spider phobia. Biol Psychol, 82(3), 293-300. doi:10.1016/j.biopsycho.2009.09.003

Levy Berg, A., Sandell, R., & Sandahl, C. (2009). Affect-focused body psychotherapy in patients with generalized anxiety disorder: Evaluation of an integrative method. Journal of Psychotherapy Integration, 19, 67-85. doi:10.1037/a0015324
Levy Berg, A., Sandell, R., & Sandahl, C. (2009). Affect-focused body psychotherapy in patients with generalized anxiety disorder: Evaluation of an integrative method. Journal of Psychotherapy Integration, 19, 67-85. doi:10.1037/a0015324
Lewis CE, Farewell D, Groves V, et al. Internet-based guided self-help for posttraumatic stress disorder (PTSD): randomized controlled trial. Depress Anxiety. 2017 Jun;34(6):555-65. doi: 10.1002/da.22645. PMID: 28557299.
Lewis S, Tarrier N, Haddock G, Bentall R, Kinderman P, Kingdon D, et al. Randomised controlled trial of cognitive-behavioural therapy in early schizophrenia : acute-phase outcomes. Br J Psychiatry. 2002;181 (suppl:s91-98.
Lexis MA, Jansen NW, Huibers MJ, Amelsvoort LG, Berkouwer A, Tjin ATG, et al. Prevention of long-term sickness absence and major depression in high-risk employees: A randomised controlled trial. Occupational and Environmental Medicine. 2011;68(6):400-7.
Liang, L., Feng, L., Zheng, X., Wu, Y., Zhang, C., & Li, J. (2021). Effect of dialectical behavior group therapy on the anxiety and depression of medical students under the normalization of epidemic prevention and control for the COVID-19 epidemic: a randomized study. Ann Palliat Med, 10(10), 10591-10599. https://doi.org/10.21037/apm-21-2466
Liberman RP, Kopelowicz A. Training skills for illness self-management in the rehabilitation of schizophrenia . A family-assisted program for Latinos in California. Salud Ment. 2009;31:93, 105.
Liberman, R. P., & Eckman, T. (1981). Behavior therapy vs insight-oriented therapy for repeated suicide attempters. Archives of general psychiatry, 38(10), 1126–1130. https://doi.org/10.1001/archpsyc.1981.01780350060007
Lidren DM, Watkins PL, Gould RA, et al. A comparison of bibliotherapy and group therapy in the treatment of panic disorder. Journal of consulting and clinical psychology 1994;62(4):865-9.
Lin, C. J., Huang, Y. H., Huang, K. Y., Wu, S. I., Chang, Y. H., Yeh, H. M., ... & Liu, S. I. (2020). A randomized controlled trial of transcultural validation of group-based psychosocial intervention for patients with bipolar disorder. Psychiatry Research, 290, 113139.
Lin, T. J., Ko, H. C., Wu, J. Y., Oei, T. P., Lane, H. Y., & Chen, C. H. (2019). The Effectiveness of Dialectical Behavior Therapy Skills Training Group vs. Cognitive Therapy Group on Reducing Depression and Suicide Attempts for Borderline Personality Disorder in Taiwan. Archives of suicide research : official journal of the International Academy for Suicide Research, 23(1), 82– 99. https://doi.org/10.1080/13811118.2018.1436104
Lincoln TM, Ziegler M, Mehl S, Kesting M, Lullmann E, Westermann S, et al. Moving from efficacy to effectiveness in Cognitive Behavioral Therapy for Psychosis : A randomized clinical practice trial. J Consult Clin Psychol. 2012;80(4):674, 86.
Lindauer RJ, Gersons BP, van Meijel EP, et al. Effects of brief eclectic psychotherapy in patients with posttraumatic stress disorder: randomized clinical trial. Journal of traumatic stress. 2005 Jun;18(3):205-12. doi: 10.1002/jts.20029. PMID: 16281214.
Linde JA, Simon GE, Ludman EJ, Ichikawa LE, Operskalski BH, Arterburn D, et al. A randomized controlled trial of behavioral weight loss treatment versus combined weight loss/depression treatment among women with comorbid obesity and depression. Annals of Behavioral Medicine. 2011;41(1):119-30.
Linden, M., Zubraegel, D., Baer, T., Franke, U., & Schlattmann, P. (2005). Efficacy of cognitive behaviour therapy in generalized anxiety disorders. Results of a controlled clinical trial (Berlin CBT-GAD Study). Psychother Psychosom, 74(1), 36-42. doi:10.1159/000082025
Linden, M., Zubraegel, D., Baer, T., Franke, U., & Schlattmann, P. (2005). Efficacy of cognitive behaviour therapy in generalized anxiety disorders. Results of a controlled clinical trial (Berlin CBT-GAD Study). Psychother Psychosom, 74(1), 36-42. doi:10.1159/000082025
Lindsay, M., Crino, R., & Andrews, G. (1997). Controlled trial of exposure and response prevention in obsessive-compulsive disorder. Br J Psychiatry, 171, 135-139.

Lindsay, W. R., Gamsu, C. V., McLaughlin, E., Hood, E. M., & Espie, C. A. (1987). A controlled trial of treatments for generalized anxiety. British journal of clinical psychology, 26 ( Pt 1), 3-15. Retrieved from https://www.cochranelibrary.com/central/doi/10.1002/central/CN-00210383/full

Lindsay, W. R., Gamsu, C. V., McLaughlin, E., Hood, E. M., & Espie, C. A. (1987). A controlled trial of treatments for generalized anxiety. British journal of clinical psychology, 26 ( Pt 1), 3-15. Retrieved from https://www.cochranelibrary.com/central/doi/10.1002/central/CN-00210383/full

Linehan, M. M., Comtois, K. A., Murray, A. M., Brown, M. Z., Gallop, R. J., Heard, H. L., Korslund, K. E., Tutek, D. A., Reynolds, S. K., & Lindenboim, N. (2006). Two-year randomized controlled trial and follow-up of dialectical behavior therapy vs therapy by experts for suicidal behaviors and borderline personality disorder. Archives of general psychiatry, 63(7), 757–766. https://doi.org/10.1001/archpsyc.63.7.757

Linehan, M. M., Comtois, K. A., Murray, A. M., Brown, M. Z., Gallop, R. J., Heard, H. L., ... & Lindenboim, N. (2006). Two-year randomized controlled trial and follow-up of dialectical behavior therapy vs therapy by experts for suicidal behaviors and borderline personality disorder. Archives of general psychiatry, 63(7), 757-766.

Litz, B. T., Schorr, Y., Delaney, E., Au, T., Papa, A., Fox, A. B., ... & Prigerson, H. G. (2014). A randomized controlled trial of an internet-based therapist-assisted indicated preventive intervention for prolonged grief disorder. Behaviour research and therapy, 61, 23-34.

Liu ET-H, Chen W-L, Li Y-H, Wang CH, Mok TJ, Huang HS. Exploring the efficacy of cognitive bibliotherapy and a potential mechanism of change in the treatment of depressive symptoms among the Chinese: A randomized controlled trial. Cognitive Therapy and Research. 2009;33(5):449-61.

Liu W, Yuan J, Wu Y, Xu L, Wang X, Meng J, et al. A randomized controlled trial of mindfulness-based cognitive therapy for major depressive disorder in undergraduate students: Dose- response effect, inflammatory markers and BDNF. Psychiatry Research. 2024;331.

Liu YE, Lv J, Sun FZ, Liang JJ, Zhang YY, Chen J, et al. Effectiveness of group acceptance and commitment therapy in treating depression for acute stroke patients. Brain Behav. 2023;13(12):e3260.

Liu, H., & Yang, Y. (2021). Effects of a psychological nursing intervention on prevention of anxiety and depression in the postpartum period: a randomized controlled trial [Article]. Annals of General Psychiatry, 20(1). https://doi.org/10.1186/s12991-020-00320-4

Lloyd-Williams M, Shiels C, Ellis J, et al. Pilot randomised controlled trial of focused narrative intervention for moderate to severe depression in palliative care patients: DISCERN trial. Palliative medicine 2018; 32(1): 206-15.

Loerch B, Graf-Morgenstern M, Hautzinger M, et al. Randomised placebo-controlled trial of moclobemide, cognitive-behavioural therapy and their combination in panic disorder with agoraphobia. The British journal of psychiatry : the journal of mental science 1999;174:205-12.

Lök, N., Bademli, K., & Selçuk-Tosun, A. (2019). The effect of reminiscence therapy on cognitive functions, depression, and quality of life in Alzheimer patients: randomized controlled trial. International Journal of Geriatric Psychiatry, 34(1), 47-53. doi:10.1002/gps.4980

LoParo, D., Mack, S. A., Patterson, B., Negi, L. T., & Kaslow, N. J. (2018). The efficacy of cognitively-based compassion training for African American suicide attempters. Mindfulness, 9(6), 1941–1954. https://doi.org/10.1007/s12671-018-0940-1

López-Luengo B, Muela-MartiÃÅnez JA. Preliminary study of a rehabilitation program based on attentional processes to treat auditory hallucinations. Cogn Neuropsychiatry. Taylor & Francis; 2016;21(4):315, 34.

López-Luengo B, Muela-MartiÃÅnez JA. Preliminary study of a rehabilitation program based on attentional processes to treat auditory hallucinations. Cogn Neuropsychiatry. Taylor & Francis; 2016;21(4):315, 34.

Lorian, C. N., Titov, N., & Grisham, J. R. (2012). Changes in risk-taking over the course of an internet-delivered cognitive behavioral therapy treatment for generalized anxiety disorder. J Anxiety Disord, 26(1), 140-149. doi:10.1016/j.janxdis.2011.10.003

Lorian, C. N., Titov, N., & Grisham, J. R. (2012). Changes in risk-taking over the course of an internet-delivered cognitive behav-ioral therapy treatment for generalized anxiety disorder. J Anxiety Disord, 26(1), 140-149. doi:10.1016/j.janxdis.2011.10.003

Losada A, Márquez-González M, Romero-Moreno R, Mausbach BT, López J, Fernández-Fernández V, et al. Cognitive–behavioral therapy (CBT) versus acceptance and commitment therapy (ACT) for dementia family caregivers with significant depressive symptoms: Results of a randomized clinical trial. Journal of Consulting and Clinical Psychology. 2015;83(4):760-72.

Lovell K, Bower P, Richards D, Barkham Mi, Sibbald B, Roberts C, et al. Developing guided self-help for depression using the medical research council complex interventions framework: A description of the modelling phase and results of an exploratory randomised controlled trial. BMC psychiatry. 2008;8.

Lund, C., Schneider, M., Garman, E. C., Davies, T., Munodawafa, M., Honikman, S., . . . Susser, E. (2019). Task-sharing of psychological treatment for antenatal depression in Khayelitsha, South Africa: Effects on antenatal and postnatal outcomes in an individual randomised controlled trial. Behaviour Research and. doi:10.1016/j.brat.2019.103466

Lundgren JG, Dahlstrom O, Andersson G, Jaarsma T, Karner Kohler A, Johansson P. The effect of guided web-based cognitive behavioral therapy on patients with depressive symptoms and heart failure: A pilot randomized controlled trial. Journal of Medical Internet Research. 2016;18(8):e194.

Luquiens, A., Tanguy, M. L., Lagadec, M., Benyamina, A., Aubin, H. J., & Reynaud, M. (2016). The efficacy of three modalities of Internet-based psychotherapy for non–treatment-seeking online problem gamblers: A randomized controlled trial. Journal of Medical Internet Research, 18(2), e36.

Lustman PJ, Griffith LS, Freedland KE, Kissel SS, Clouse RE. Cognitive behavior therapy for depression in type 2 diabetes mellitus. A randomized, controlled trial. Annals of Internal Medicine. 1998;129(8):613-21.

Lynch D, Tamburrino M, Nagel R, Smith MK. Telephone-based treatment for family practice patients with mild depression. Psychological reports. 2004;94(3 Pt 1):785-92.

Lynch DJ, Tamburrino MB, Nagel R. Telephone counseling for patients with minor depression: Preliminary findings in a family practice setting. The Journal of family practice. 1997;44(3):293-8.

Lynch, T. R., Hempel, R. J., Whalley, B., Byford, S., Chamba, R., Clarke, P., . . . Russell, I. T. (2019). Refractory depression - mechanisms and efficacy of radically open dialectical behaviour therapy (RefraMED): findings of a randomised trial on benefits and harms. The British journal of psychiatry : the journal of mental science, 1-9. doi:10.1192/bjp.2019.53

MacLean S, Corsi DJ, Litchfield S, et al. Coach-facilitated web-based therapy compared with information about web-based resources in patients referred to secondary mental health care for depression: Randomized controlled trial. Journal of Medical Internet Research. 2020;22(6).

MacPherson H, Richmond S, Bland M, Brealey S, Gabe R, Hopton A, et al. Acupuncture and counselling for depression in primary care: A randomised controlled trial. PLoS medicine. 2013;10(9):e1001518.

Maguen S, Burkman KM, Madden E, et al. Impact of killing in war: a randomized, controlled pilot trial. J Clin Psychol. 2017 Sep;73(9):997-1012. doi: 10.1002/jclp.22471. PMID: 28294318.

Mahmoodi, M., Bakhtiyari, M., Masjedi Arani, A., Mohammadi, A., & Saberi Isfeedvajani, M. (2021). The comparison between CBT focused on perfectionism and CBT focused on emotion regulation for individuals with depression and anxiety disorders and dysfunctional perfectionism: A randomized controlled trial. Behavioural and Cognitive Psychotherapy, 49(4), 454-471. https://doi.org/10.1017/S1352465820000909

Maina G, Forner F, Bogetto F. Randomized controlled trial comparing brief dynamic and supportive therapy with waiting list condition in minor depressive disorders. Psychotherapy and Psychosomatics. 2005;74(1):43-50.

Majdara, E., Rahimmian, I., Talepassand, S., & Gregory, R. J. (2019). A randomized trial of dynamic deconstructive psychotherapy in Iran for borderline personality disorder. Journal of the American Psychoanalytic Association, 67(5), NP1-NP7.

Majidzadeh S, Mirghafourvand M, Farvareshi M, Yavarikia P. The effect of cognitive behavioral therapy on depression and anxiety of women with polycystic ovary syndrome: a randomized controlled trial. BMC Psychiatry. 2023;23(1):332.

Malouff JM, Lanyon RI, Schutte NS. Effectiveness of a brief group RET treatment for divorce-related dysphoria. Journal of Rational-Emotive and Cognitive-Behavior Therapy. 1988;6(3):162-71.

Mancebo, M. C., Yip, A. G., Boisseau, C. L., Rasmussen, S. A., & Zlotnick, C. (2021). Behavioral Therapy Teams for Obsessive-Compulsive Disorder: Lessons Learned From a Pilot Randomized Trial in a Community Mental Health Center. Behav Ther, 52(5), 1296-1309.

Mansour, N., Labib, N., Khalil, M., & Esmat, S. (2022). Brief Cognitive Behavioral Therapy for Patients with Comorbid Depression and Type 2 Diabetes in an Urban Primary Care Facility: Randomized Controlled Trial. Open Access Macedonian Journal of Medical Sciences, 10, 60-67. doi:10.3889/oamjms.2022.7883

Marasinghe, R. B., Edirippulige, S., Kavanagh, D., Smith, A., & Jiffry, M. T. (2012). Effect of mobile phone-based psychotherapy in suicide prevention: a randomized controlled trial in Sri Lanka. Journal of telemedicine and telecare, 18(3), 151–155. https://doi.org/10.1258/jtt.2012.SFT107

Marasinghe, R. B., Edirippulige, S., Kavanagh, D., Smith, A., & Jiffry, M. T. (2012). Effect of mobile phone-based psychotherapy in suicide prevention: a randomized controlled trial in Sri Lanka. Journal of telemedicine and telecare, 18(3), 151–155. https://doi.org/10.1258/jtt.2012.SFT107

Marceaux, J. C., & Melville, C. L. (2011). Twelve-step facilitated versus mapping-enhanced cognitive-behavioral therapy for pathological gambling: A controlled study. Journal of Gambling Studies, 27(1), 171-190.

Marchand A, Coutu MF, Dupuis G, et al. Treatment of panic disorder with agoraphobia: randomized placebo-controlled trial of four psychosocial treatments combined with imipramine or placebo. Cognitive behaviour therapy 2008;37(3):146-59.

Margolies SO, Rybarczyk B, Vrana SR, et al. Efficacy of a cognitive-behavioral treatment for insomnia and nightmares in Afghanistan and Iraq veterans with PTSD. Journal of Clinical Psychology. 2013 Oct;69(10):1026-42. doi: 10.1002/jclp.21970. PMID: 23629959.

Martens, M. P., Arterberry, B. J., Takamatsu, S. K., Masters, J., & Dude, K. (2015). The efficacy of a personalized feedback-only intervention for at-risk college gamblers. Journal of Consulting and Clinical Psychology, 83(3), 494–499.

Martin PR, Aiello R, Gilson K, Meadows G, Milgrom J, Reece J. Cognitive behavior therapy for comorbid migraine and/or tension-type headache and major depressive disorder: An exploratory randomized controlled trial. Behaviour Research and Therapy. 2015;73:8-18.

Martini B, Rosso G, Chiodelli DF, et al. Brief dynamic therapy combined with pharmacotherapy in the treatment of panic disorder with concurrent depressive symptoms. Clinical Neuropsychiatry 2011;8(3):204-11.

Mathur, S., Sharma, M. P., Balachander, S., Kandavel, T., & Reddy, Y. C. J. (2021). A randomized controlled trial of mindfulness-based cognitive therapy vs stress management training for obsessive-compulsive disorder. Journal of Affective Disorders, 282, 58-68.

Matsumoto, K., Hamatani, S., Makino, T., Takahashi, J., Suzuki, F., Ida, T., . . . Omori, I. M. (2022). Guided internet-based cognitive behavioral therapy for obsessive-compulsive disorder: A multicenter randomized controlled trial in Japan. Internet interventions, 28, 100515.

Matsuzaka, C., Wainberg, M., Norcini, P. A., Hoffmann, E., Coimbra, B., Braga, R., . . . Mello, M. (2017). Task shifting interpersonal counseling for depression: a pragmatic randomized controlled trial in primary care. BMC Psychiatry, 17(1)

Mattick, R. P., Peters, L., & Clarke, J. C. (1989). Exposure and cognitive restructuring for social phobia: A controlled study. Behav Ther, 20(1), 3-23. https://doi.org/10.1016/S0005-7894(89)80115-7

Mattick, R. P., Peters, L., & Clarke, J. C. (1989). Exposure and cognitive restructuring for social phobia: A controlled study. Behav Ther, 20(1), 3-23. https://doi.org/10.1016/S0005-7894(89)80115-7

Mawson, D., Marks, I. M., Ramm, L., & Stern, R. S. (1981). Guided mourning for morbid grief: A controlled study. The British Journal of Psychiatry, 138(3), 185-193.

McAfee, N. W., Martens, M. P., Herring, T. E., Takamatsu, S. K., & Foss, J. M. (2020). The efficacy of personalized feedback interventions delivered via smartphone among at-risk college student gamblers. Journal of Gambling Issues, 45.

McClay CA, Collins K, Matthews L, Haig C, McConnachie A, Morrison J, et al. A community-based pilot randomised controlled study of life skills classes for individuals with low mood and depression. BMC psychiatry. 2015;15:17.

McCusker, J., Jones, J. M., Li, M., Faria, R., Yaffe, M. J., Lambert, S. D., Ciampi, A., Belzile, E., & de Raad, M. (2021). CanDirect: Effectiveness of a Telephone-Supported Depression Self-Care Intervention for Cancer Survivors. J Clin Oncol, 39(10), 1150-1161. https://doi.org/10.1200/jco.20.01802

McDonagh A, Friedman MJ, McHugo GJ, et al. Randomized trial of cognitive-behavioral therapy for chronic posttraumatic stress disorder in adult female survivors of childhood sexual abuse. J Consult Clin Psychol. 2005 Jun;73(3):515-24. doi: 10.1037/0022-006X.73.3.515. PMID: 15982149.
McGeary DD, Resick PA, Penzien DB, et al. Cognitive behavioral therapy for veterans with comorbid posttraumatic headache and posttraumatic stress disorder symptoms: a randomized clinical trial. JAMA Neurol. 2022;79(8):746-57. doi: 10.1001/jamaneurol.2022.1567. PMID: 35759281.
McGuinness, B., Finucane, N., & Roberts, A. (2015). A hospice-based bereavement support group using creative arts: an exploratory study. Illness, Crisis & Loss, 23(4), 323-342.
McIndoo CC, File AA, Preddy T, Clark CG, Hopko DR. Mindfulness-based therapy and behavioral activation: A randomized controlled trial with depressed college students. Behaviour Research and Therapy. 2016;77:118-28.
McKee MD, Zayas LH, Fletcher J, Boyd RC, Nam SH. Results of an intervention to reduce perinatal depression among low-income minority women in community primary care. Journal of Social Service Research. 2006;32(4):63-81.
McLay RN, Wood DP, Webb-Murphy JA, et al. A randomized, controlled trial of virtual reality-graded exposure therapy for post-traumatic stress disorder in active duty service members with combat-related post-traumatic stress disorder. Cyberpsychol Behav Soc Netw. 2011 Apr;14(4):223-9. doi: 10.1089/cyber.2011.0003. PMID: 21332375.
McMain, S. F., Guimond, T., Barnhart, R., Habinski, L., & Streiner, D. L. (2017). A randomized trial of brief dialectical behaviour therapy skills training in suicidal patients suffering from borderline disorder. Acta Psychiatrica Scandinavica, 135(2), 138-148.
Mennen, F. E., Palmer Molina, A., Monro, W. L., Duan, L., Stuart, S., & Sosna, T. (2021). Effectiveness of an Interpersonal Psychotherapy (IPT) Group Depression Treatment for Head Start Mothers: A Cluster-Randomized Controlled Trial. J Affect Disord, 280(Pt B), 39-48. https://doi.org/10.1016/j.jad.2020.11.074
Mennin, D. S., Fresco, D. M., O'Toole, M. S., & Heimberg, R. G. (2018). A randomized controlled trial of emotion regulation therapy for generalized anxiety disorder with and without co-occurring depression. Journal of Consulting and Clinical Psychology, 86(3), 268-281. doi:10.1037/ccp0000289
Mennin, D. S., Fresco, D. M., O'Toole, M. S., & Heimberg, R. G. (2018). A randomized controlled trial of emotion regulation therapy for generalized anxiety disorder with and without co-occurring depression. Journal of Consulting and Clinical Psy-chology, 86(3), 268-281. doi:10.1037/ccp0000289
Merza D, Amani B, Savoy C, Babiy Z, Bieling PJ, Streiner DL, et al. Online peer-delivered group cognitive-behavioral therapy for postpartum depression: A randomized controlled trial. Acta Psychiatrica Scandinavica. 2023.
Meulenbeek P, Willemse G, Smit F, et al. Early intervention in panic: pragmatic randomised controlled trial. The British journal of psychiatry 2010;196(4):326-31.
Meuret AE, Wilhelm FH, Ritz T, Roth WT. Feedback of end-tidal pCO2 as a therapeutic approach for panic disorder. Journal of Psychiatric Research 2008;42(7):560–8.
Meyerbroeker K, Morina N, Kerkhof GA, et al. Virtual reality exposure therapy does not provide any additional value in agoraphobic patients: a randomized controlled trial. Psychotherapy and psychosomatics 2013;82(3):170-6.
Michalak J, Schultze M, Heidenreich T, Schramm E. A randomized controlled trial on the efficacy of mindfulness-based cognitive therapy and a group version of cognitive behavioral analysis system of psychotherapy for chronically depressed patients. Journal of Consulting and Clinical Psychology. 2015;83(5):951-63.
Milgrom J, Danaher BG, Gemmill AW, Holt C, Holt CJ, Seeley JR, et al. Internet cognitive behavioral therapy for women with postnatal depression: A randomized controlled trial of MumMoodBooster. Journal of Medical Internet Research. 2016;18(3):e54.
Milgrom J, Holt C, Holt CJ, Ross J, Ericksen J, Gemmill AW. Feasibility study and pilot randomised trial of an antenatal depression treatment with infant follow-up. Archives of Women's Mental Health. 2015;18(5):717-30.
Milgrom J, Holt CJ, Gemmill AW, Ericksen J, Leigh B, Buist A, et al. Treating postnatal depressive symptoms in primary care: A randomised controlled trial of GP management, with and without adjunctive counselling. BMC psychiatry. 2011;11:95.

Milgrom J, Negri LM, Gemmill AW, McNeil M, Martin PR. A randomized controlled trial of psychological interventions for postnatal depression. British Journal of Clinical Psychology. 2005;44(4):529-42.
Milgrom, J., Danaher, B. G., Seeley, J. R., Holt, C. J., Holt, C., Ericksen, J., Tyler, M. S., Gau, J. M., & Gemmill, A. W. (2021). Internet and Face-to-face Cognitive Behavioral Therapy for Postnatal Depression Compared With Treatment as Usual: Randomized Controlled Trial of MumMoodBooster. J Med Internet Res, 23(12), e17185. https://doi.org/10.2196/17185
Minns, S., Levihn-Coon, A., Carl, E., Smits, J. A. J., Miller, W., Howard, D., . . . Powers, M. B. (2018). Immersive 3D exposure-based treatment for spider fear: A randomized controlled trial. J Anxiety Disord, 58, 1-7. doi:10.1016/j.janxdis.2018.05.006
Miranda J, Chung JY, Green BL, Krupnick J, Siddique J, Revicki DA, et al. Treating depression in predominantly low-income young minority women: A randomized controlled trial. Jama. 2003;290(1):57-65.
Miyahira SD, Folen RA, Hoffman HG, et al. The effectiveness of VR exposure therapy for PTSD in returning warfighters. Stud Health Technol Inform. 2012;181:128-32. PMID: 22954842.
Mohlman, J., Gorenstein, E. E., Kleber, M., De Jesus, M., Gorman, J. M., & Papp, L. A. (2003). Standard and enhanced cognitive-behavior therapy for late-life generalized anxiety disorder: Two pilot investigations. American Journal of Geriatric Psychiatry, 11(1), 24-32. doi:10.1097/00019442-200301000-00005
Mohlman, J., Gorenstein, E. E., Kleber, M., De Jesus, M., Gorman, J. M., & Papp, L. A. (2003). Standard and enhanced cognitive-behavior therapy for late-life generalized anxiety disorder: Two pilot investigations. American Journal of Geriatric Psychia-try, 11(1), 24-32. doi:10.1097/00019442-200301000-00005
Mohr DC, Carmody T, Erickson L, Jin L, Leader J. Telephone-administered cognitive behavioral therapy for veterans served by community-based outpatient clinics. Journal of Consulting and Clinical Psychology. 2011;79(2):261-5.
Mohr DC, Duffecy J, Ho J, Kwasny M, Cai X, Burns MN, et al. A randomized controlled trial evaluating a manualized TeleCoaching protocol for improving adherence to a web-based intervention for the treatment of depression. PLoS One. 2013;8(8):e70086.
Mohr DC, Likosky W, Bertagnolli A, Goodkin DE, Van Der Wende J, Dwyer P, et al. Telephone-administered cognitive-behavioral therapy for the treatment of depressive symptoms in multiple sclerosis. Journal of Consulting and Clinical Psychology. 2000;68(2):356-61.
Moldovan R, Cobeanu O, David D. Cognitive bibliotherapy for mild depressive symptomatology: Randomized clinical trial of efficacy and mechanisms of change. Clinical Psychology and Psychotherapy. 2013;20(6):482-93.
Monson CM, Fredman SJ, Macdonald A, et al. Effect of cognitive-behavioral couple therapy for PTSD: a randomized controlled trial. JAMA. 2012 Aug 15;308(7):700-9. doi: 10.1001/jama.2012.9307. PMID: 22893167.
Monson CM, Schnurr PP, Resick PA, et al. Cognitive processing therapy for veterans with military-related posttraumatic stress disorder. J Consult Clin Psychol. 2006 Oct;74(5):898-907. doi: 10.1037/0022-006X.74.5.898. PMID: 17032094.
Montero-Marín, J., Araya, R., Pérez-Yus, M. C., Mayoral, F., Gili, M., Botella, C., . . . López-Del-Hoyo, Y. (2016). An internet-based intervention for depression in primary Care in Spain: a randomized controlled trial. Journal of medical Internet research, 18(8), e231.
Montesó-Curto, P., García-Martínez, M., Gómez-Martínez, C., Ferré-Almo, S., Panisello-Chavarria, M. L., Genís, S. R., Mateu Gil, M. L., Cubí Guillén, M. T., Colás, L. S., Usach, T. S., Herrero, A. S., & Ferré-Grau, C. (2015).
Moon, J. R., Huh, J., Song, J., Kang, I. S., Park, S. W., Chang, S. A., . . . Han, J. S. (2021). The effects of rational emotive behavior therapy for depressive symptoms in adults with congenital heart disease. Heart Lung, 50(6), 906-913. doi:10.1016/j.hrtlng.2021.07.011
Morath J, Gola H, Sommershof A, et al. The effect of trauma-focused therapy on the altered T cell distribution in individuals with PTSD: evidence from a randomized controlled trial. J Psychiatr Res. 2014 Jul;54:1-10. doi: 10.1016/j.jpsychires.2014.03.016. PMID: 24726027.
Morley, K. C., Sitharthan, G., Haber, P. S., Tucker, P., & Sitharthan, T. (2014). The efficacy of an opportunistic cognitive behavioral intervention package (OCB) on substance use and comorbid

suicide risk: a multisite randomized controlled trial. Journal of consulting and clinical psychology, 82(1), 130–140. https://doi.org/10.1037/a0035310
Morrison AP, Turkington D, Pyle M, Spencer H, Brabban A, Dunn G, et al. Cognitive therapy for people with schizophrenia spectrum disorders not taking antipsychotic drugs : a single- blind randomised controlled trial. Lancet [Internet]. Elsevier Ltd; 2014: 383(9926):1395, 403. Available from: http://dx.doi.org/10.1016/S0140-6736(13)62246-1
Mortberg, E., Clark, D. M., Sundin, O., & Aberg Wistedt, A. (2007). Intensive group cognitive treatment and individual cognitive therapy vs. treatment as usual in social phobia: a randomized controlled trial. Acta Psychiatr Scand, 115(2), 142-154. https://doi.org/10.1111/j.1600-0447.2006.00839.x
Mortberg, E., Karlsson, A., Fyring, C., & Sundin, O. (2006). Intensive cognitive-behavioral group treatment (CBGT) of social phobia: a randomized controlled study. J Anxiety Disord, 20(5), 646-660. https://doi.org/10.1016/j.janxdis.2005.07.005
Moses, A. N., & Hollandsworth, J. G. (1985). Relative effectiveness of education alone versus stress inoculation training in the treatment of dental phobia. Behavior Therapy, 16(5), 531-537. doi:https://doi.org/10.1016/S0005-7894(85)80031-9
Mossey JM, Knott KA, Higgins M, Talerico K. Effectiveness of a psychosocial intervention, interpersonal counseling, for subdysthymic depression in medically ill elderly. The journals of gerontology Series A, Biological sciences and medical sciences. 1996;51(4):M172-8.
Mueller-Weinitschke C, Bengel J, Baumeister H, Krämer LV. Effects of a Web-Based Behavioral Activation Intervention on Depressive Symptoms, Activation, Motivation, and Volition: Results of a Randomized Controlled Trial. Psychotherapy and Psychosomatics. 2023.
Mueser KT, Rosenberg SD, Xie HY, et al. A randomized controlled trial of cognitive-behavioral treatment for posttraumatic stress disorder in severe mental illness. J Consult Clin Psychol. 2008 Apr;76(2):259-71. doi: 10.1037/0022-006X.76.2.259. PMID: 18377122.
Mukhtar, F., Oei, T. P., & Yaacob, M. (2011). Effectiveness of group cognitive behaviour therapy augmentation in reducing negative cognitions in the treatment of depression in Malaysia. ASEAN Journal of Psychiatry, 12(1), 50-65.
Mulcahy R, Reay RE, Wilkinson RB, Owen C. A randomised control trial for the effectiveness of group interpersonal psychotherapy for postnatal depression. Archives of Women's Mental Health. 2010;13(2):125-39.
Mulkens, S., Bögels, S. M., de Jong, P. J., & Louwers, J. (2001). Fear of blushing: effects of task concentration training versus exposure in vivo on fear and physiology. J Anxiety Disord, 15(5), 413-432. https://doi.org/10.1016/s0887-6185(01)00073-1
Muris, P., & Merckelbach, H. (1997). Treating Spider Phobics with Eye Movement Desensitization and Reprocessing: A Controlled Study. Behavioural and Cognitive Psychotherapy, 25, 39-50. doi:10.1017/S1352465800015381
Musa, Z. A., Soh, K. L., Mukhtar, F., Soh, K. Y., Oladele, T. O., & Soh, K. G. (2021). Effectiveness of mindfulness-based cognitive therapy among depressed individuals with disabilities in Nigeria: A randomized controlled trial. Psychiatry Res, 296, 113680. doi:10.1016/j.psychres.2020.113680
Mynors-Wallis L, Gath D, Lloyd-Thomas A, Tomlinson D. Randomised controlled trial comparing problem solving treatment with amitriptyline and placebo for major depression in primary care. BMJ. 1995;310(6977):441-5.
Myrseth, H., Litlerè, I., Støylen, I. J., & Pallesen, S. (2009). A controlled study of the effect of cognitive–behavioural group therapy for pathological gamblers. Nordic Journal of Psychiatry, 63(1), 22-31.
Nadort, E., Schouten, R. W., Boeschoten, R. E., Smets, Y., Chandie Shaw, P., Vleming, L. J., . . . Siegert, C. E. H. (2022). Internet-based treatment for depressive symptoms in hemodialysis patients: A cluster randomized controlled trial. Gen Hosp Psychiatry, 75, 46-53. doi:10.1016/j.genhosppsych.2022.01.008
Naeem F, Gul M, Irfan M, Munshi T, Asif A, Rashid S, et al. Brief Culturally adapted CBT (CaCBT) for depression: A randomized controlled trial from Pakistan. Journal of Affective Disorders. 2015;177:101-7.
Naeem F, Johal R, Mckenna C, Rathod S, Ayub M, Lecomte T, et al. Cognitive behavior therapy for psychosis based Guided Self-help (CBTp-GSH) delivered by frontline mental health professionals :

Results of a feasibility study. Schizophr Res [Internet]. Elsevier B.V.; 2016;173(1, 2):69, 74. Available from: http://dx.doi.org/10.1016/j.schres.2016.03.003
Naeem F, Saeed S, Irfan M, Kiran T, Mehmood N, Gul M, et al. Brief culturally adapted CBT for psychosis (CaCBTp): A randomized controlled trial from a low income country. Schizophr Res. 2015;164:143, 8.
Naeem F, Sarhandi I, Gul M, Khalid M, Aslam M, Anbrin A. A multicentre randomised controlled trial of a carer supervised culturally adapted cbt (cacbt) based self-help for depression in pakistan. Journal of Affective Disorders. 2013;156:224-7.
Nakagawa, A., Mitsuda, D., Sado, M., Abe, T., Fujisawa, D., Kikuchi, T., . . . Ono, Y. (2017). Effectiveness of supplementary cognitive-behavioral therapy for pharmacotherapy-resistant depression: A randomized controlled trial. Journal of clinical psychiatry, 78(8), 1126-1135
Nakao, S., Nakagawa, A., Oguchi, Y., Mitsuda, D., Kato, N., Nakagawa, Y., Tamura, N., Kudo, Y., Abe, T., Hiyama, M., Iwashita, S., Ono, Y., & Mimura, M. Web-Based Cognitive Behavioral Therapy Blended With Face-to-Face Sessions for Major Depression: Randomized Controlled Trial. J Med Internet Res, 20(9), e10743.
Nakimuli-Mpungu E, Musisi S, Wamala K, et al. Effectiveness and cost-effectiveness of group support psychotherapy delivered by trained lay health workers for depression treatment among people with HIV in Uganda: a cluster-randomised trial. Lancet Glob Health. 2020;8(3):e387-e398.
Nakimuli-Mpungu E, Wamala K, Okello J, Alderman S, Odokonyero R, Mojtabai R, et al. Group support psychotherapy for depression treatment in people with HIV/AIDS in northern Uganda: A single-centre randomised controlled trial. The Lancet HIV. 2015;2(5):e190-e9.
Nam, I. (2016). Complicated grief treatment for older adults: The critical role of a supportive person. Psychiatry Research, 244, 97-102.
Nam, I. (2017). Restoration-focused coping reduces complicated grief among older adults: A randomized controlled study. The European Journal of Psychiatry, 31(3), 93-98.
Nasrin, F., Rimes, K., Reinecke, A., Rinck, M., & Barnhofer, T. (2017). Effects of Brief Behavioural Activation on Approach and Avoidance Tendencies in Acute Depression: preliminary Findings. Behavioural and Cognitive Psychotherapy, 45(1), 58-72.
Neugebauer R, Kline J, Markowitz JC, Bleiberg KL, Baxi L, Rosing MA, et al. Pilot randomized controlled trial of interpersonal counseling for subsyndromal depression following miscarriage. Journal of Clinical Psychiatry. 2006;67(8):1299-304.
Neuner F, Kurreck S, Ruf M, et al. Can asylum-seekers with posttraumatic stress disorder be successfully treated? A randomized controlled pilot study. Cogn Behav Ther. 2010;39(2):81-91. doi: 10.1080/16506070903121042. PMID: 19816834.
Newby JM, Lang T, Werner-Seidler A, Holmes E, Moulds ML. Alleviating distressing intrusive memories in depression: A comparison between computerised cognitive bias modification and cognitive behavioural education. Behaviour Research and Therapy. 2014;56:60-7.
Newby, J. M., et al. (2013). "Internet cognitive behavioural therapy for mixed anxiety and depression: a randomized controlled trial and evidence of effectiveness in primary care." Psychol Med 43(12): 2635-2648.
Newby, J., Robins, L., Wilhelm, K., Smith, J., Fletcher, T., Gillis, I., . . . Andrews, G. (2017). Web-Based Cognitive Behavior Therapy for Depression in People With Diabetes Mellitus: a Randomized Controlled Trial. Journal of medical Internet research, 19(5), e157.
Newman, M. G., Hofmann, S. G., Trabert, W., Roth, W. T., & Taylor, C. B. (1994). Does behavioral treatment of social phobia lead to cognitive changes? Behav Ther, 25(3), 503-517. https://doi.org/https://doi.org/10.1016/S0005-7894(05)80160-1
Newman, M. G., Jacobson, N. C., Rackoff, G. N., Bell, M. J., & Taylor, C. B. (2020). A randomized controlled trial of a smartphone-based application for the treatment of anxiety. Psychother Res, 1-12. doi:10.1080/10503307.2020.1790688
Newman, M. G., Jacobson, N. C., Rackoff, G. N., Bell, M. J., & Taylor, C. B. (2020). A randomized controlled trial of a smartphone-based application for the treatment of anxiety. Psychother Res, 1-12. doi:10.1080/10503307.2020.1790688
Nezu AM, Perri MG. Social problem-solving therapy for unipolar depression: An initial dismantling investigation. Journal of Consulting and Clinical Psychology. 1989;57(3):408-13.

Nezu AM. Efficacy of a social problem-solving therapy approach for unipolar depression. Journal of Consulting and Clinical Psychology. 1986;54(2):196-202.

Ng SE, Tien A, Thayala JN, Ho RC, Chan MF. The effect of life story review on depression of older community-dwelling Chinese adults in Singapore: A preliminary result. International Journal of Geriatric Psychiatry. 2013;28(3):328-30.

Ngai FW, Wong P-C, Leung KY, Chau PH, Chung KF. The effect of telephone-based cognitive-behavioral therapy on postnatal depression: A randomized controlled trial. Psychotherapy and Psychosomatics. 2015;84(5):294-303.

Niedermoser DW, Kalak N, Kiyhankhadiv A, et al. Workplace-Related Interpersonal Group Psychotherapy to Improve Life at Work in Individuals With Major Depressive Disorders: a Randomized Interventional Pilot Study. Frontiers in psychiatry. 2020;11.

Nilsson, J. E., Lundh, L. G., & Viborg, G. (2012). Imagery rescripting of early memories in social anxiety disorder: an experimental study. Behav Res Ther, 50(6), 387-392. https://doi.org/10.1016/j.brat.2012.03.004

Ninomiya, A., et al. (2019). "Effectiveness of mindfulness-based cognitive therapy in patients with anxiety disorders in secondary-care settings: a randomized controlled trial." Psychiatry and clinical neurosciences.

Nobis S, Lehr D, Ebert DD, Baumeister H, Snoek F, Riper H, et al. Efficacy of a web-based intervention with mobile phone support in treating depressive symptoms in adults with type 1 and type 2 diabetes: A randomized controlled trial. Diabetes care. 2015;38(5):776-83.

Nollett CL, Bray N, Bunce C, Casten RJ, Edwards RT, Hegel MT, et al. Depression in Visual Impairment Trial (DEPVIT): A randomized clinical trial of depression treatments in people with low vision. Investigative Ophthalmology and Visual Science. 2016;57(10):4247-54.

Noone, D., Payne, J., Stott, J., Aguirre, E., Patel-Palfreman, M. M., Stoner, C., . . . Spector, A. (2022). The Feasibility of a Mindfulness Intervention for Depression in People with Mild Dementia: A Pilot Randomized Controlled Trial. Clinical gerontologist, 1-13. doi:10.1080/07317115.2022.2094741

Nordahl, H. M., Borkovec, T. D., Hagen, R., Kennair, L. E. O., Hjemdal, O., Solem, S., . . . Wells, A. (2018). Metacognitive therapy versus cognitive-behavioural therapy in adults with generalised anxiety disorder. BJPsych Open, 4(5), 393-400. doi:10.1192/bjo.2018.54

Nordahl, H. M., Borkovec, T. D., Hagen, R., Kennair, L. E. O., Hjemdal, O., Solem, S., . . . Wells, A. (2018). Metacognitive ther-apy versus cognitive-behavioural therapy in adults with generalised anxiety disorder. BJPsych Open, 4(5), 393-400. doi:10.1192/bjo.2018.54

Northwood AK, Vukovich MM, Beckman A, et al. Intensive psychotherapy and case management for Karen refugees with major depression in primary care: a pragmatic randomized control trial. BMC Fam Pract. 2020;21(1):17.

Nyström, M., Stenling, A., Sjöström, E., Neely, G., Lindner, P., HassmÈn, P., . . . Carlbring, P. (2017). Behavioral activation versus physical activity via the internet: a randomized controlled trial. Journal of Affective Disorders, 215, 85-93.

O'Connor, K. P., Aardema, F., Robillard, S., Guay, S., Pélissier, M. C., Todorov, C., . . . Doucet, P. (2006). Cognitive behaviour therapy and medication in the treatment of obsessive-compulsive disorder. Acta Psychiatr Scand, 113(5), 408-419.

O'Connor, K., Todorov, C., Robillard, S., Borgeat, F., & Brault, M. (1999). Cognitive-behaviour therapy and medication in the treatment of obsessive-compulsive disorder: a controlled study. The Canadian Journal of Psychiatry, 44(1), 64-71.

O'Connor, M., Nikoletti, S., Kristjanson, L. J., Loh, R., & Willcock, B. (2003). Writing therapy for the bereaved: Evaluation of an intervention. Journal of palliative medicine, 6(2), 195-204.

O'Hara MW, Stuart S, Gorman LL, Wenzel A. Efficacy of interpersonal psychotherapy for postpartum depression. Archives of General Psychiatry. 2000;57(11):1039-45.

O'Mahen H, Himle JA, Fedock G, Henshaw E, Flynn H. A pilot randomized controlled trial of cognitive behavioral therapy for perinatal depression adapted for women with low incomes. Depression and anxiety. 2013;30(7):679-87.

O'Neil A, Taylor B, Sanderson K, Cyril S, Chan B, Hawkes AL, et al. Efficacy and feasibility of a tele-health intervention for acute coronary syndrome patients with depression: Results of the "MoodCare" randomized controlled trial. Annals of Behavioral Medicine. 2014;48(2):163-74.

O'Toole, M. S., Arendt, M. B., & Pedersen, C. M. (2019). Testing an App- Assisted Treatment for Suicide Prevention in a Randomized Controlled Trial: Effects on Suicide Risk and Depression. Behavior therapy, 50(2), 421–429. https://doi.org/10.1016/j.beth.2018.07.007

Oehler C, Görges F, Rogalla M, Rummel-Kluge C, Hegerl U. Efficacy of a Guided Web-Based Self-Management Intervention for Depression or Dysthymia: Randomized Controlled Trial With a 12-Month Follow-Up Using an Active Control Condition. J Med Internet Res. 2020;22(7):e15361.

Oei, T. P. S., Raylu, N., & Lai, W. W. (2018). Effectiveness of a self help cognitive behavioural treatment program for problem gamblers: a randomised controlled trial. Journal of Gambling Studies, 34(2), 581-595.

Oei, T. P., Raylu, N., & Casey, L. M. (2010). Effectiveness of group and individual formats of a combined motivational interviewing and cognitive behavioral treatment program for problem gambling: A randomized controlled trial. Behavioural and Cognitive Psychotherapy, 38(2), 233-238.

Okeke NM, Onah BN, Ekwealor NE, Ekwueme SC, Ezugwu JO, Edeh EN, et al. Effect of a randomized group intervention for depression among Nigerian pre-service adult education teachers. Medicine (Baltimore). 2023;102(27):e34159.

Olivares-Olivares, P. J., Olivares, J., Macia, D., Macia, A., & Montesinos, L. (2016). Community versus clinical cognitive-behavioral intervention in young-adult Spanish population with generalized social phobia. Terapia psicologica, 34(1), 23-30. https://doi.org/10.4067/S0718-48082016000100003

Olukolade, O., & Osinowo, H. (2017). Efficacy of Cognitive Rehabilitation Therapy on Poststroke Depression among Survivors of First Stroke Attack in Ibadan, Nigeria. Behavioural Neurology, 2017

Omidi A, Mohammadkhani P, Mohammadi A, Zargar F. Comparing mindfulness based cognitive therapy and traditional cognitive behavior therapy with treatments as usual on reduction of major depressive disorder symptoms. Iranian Red Crescent Medical Journal. 2013;15(2):142-6.

Omiya H, Yamashita K, Miyata T, Hatakeyama Y, Miyajima M, Yambe K, et al. Pilot study of the effects of cognitive remediation therapy using the frontal/executive program for treating chronic schizophrenia. Open Psychol J. 2016;9:121, 8.

Onuigbo, L. N., Eseadi, C., Ebifa, S., Ugwu, U. C., Onyishi, C. N., & Oyeoku, E. K. (2019). Effect of rational emotive behavior therapy program on depressive symptoms among university students with blindness in Nigeria. Journal of Rational-Emotive & Cognitive-Behavior Therapy, 37(1), 17-38. doi:10.1007/s10942-018-0297-3

Oosterbaan, D., Balkom, A., Spinhoven, P., Oppen, P., & van Dyck, R. (2001). Cognitive Therapy Versus Moclobemide in Social Phobia: A Controlled Study. Clinical psychology & psychotherapy, 8, 263-273. https://doi.org/10.1002/cpp.291

Oromendia P, Orrego J, Bonillo A, et al. Internet-based self-help treatment for panic disorder: a randomized controlled trial comparing mandatory versus optional complementary psychological support. Cognitive behaviour therapy 2016;45(4):270-86.

Orvati Aziz, M., Mehrinejad, S. A., Hashemian, K., & Paivastegar, M. (2020). Integrative therapy (short-term psychodynamic psychotherapy & cognitive-behavioral therapy) and cognitive-behavioral therapy in the treatment of generalized anxiety disorder: A randomized controlled trial. Complement Ther Clin Pract, 39, 101122. doi:10.1016/j.ctcp.2020.101122

Orvati Aziz, M., Mehrinejad, S. A., Hashemian, K., & Paivastegar, M. (2020). Integrative therapy (short-term psychodynamic psychotherapy & cognitive-behavioral therapy) and cognitive-behavioral therapy in the treatment of generalized anxiety disorder: A randomized controlled trial. Complement Ther Clin Pract, 39, 101122. doi:10.1016/j.ctcp.2020.101122

Ost LG, Thulin U, Ramnero J. Cognitive behavior therapy vs exposure in vivo in the treatment of panic disorder with agoraphobia (corrected from agrophobia). Behaviour research and therapy 2004;42(10):1105-27.

Öst, L. G., Alm, T., Brandberg, M., & Breitholtz, E. (2001). One vs five sessions of exposure and five sessions of cognitive therapy in the treatment of claustrophobia. Behav Res Ther, 39(2), 167-183. doi:10.1016/s0005-7967(99)00176-x

Öst, L.-G., Johansson, J., & Jerremalm, A. (1982). Individual response patterns and the effects of different behavioral methods in the treatment of claustrophobia. Behaviour Research and Therapy, 20(5), 445-460. doi:https://doi.org/10.1016/0005-7967(82)90066-3

Otu MS, Ebizie EN, Otu FM, Eseadi C. Efficacy of a counselling video blog intervention from YouTube on depression reduction among primary school teachers. Psychological Studies. 2023;68(3):335-41.

Pace TM, Dixon DN. Changes in depressive self-schemata and depressive symptoms following cognitive therapy. Journal of Counseling Psychology. 1993;40(3):288.

Pagoto S, Schneider KL, Whited MC, Oleski JL, Merriam P, Appelhans B, et al. Randomized controlled trial of behavioral treatment for comorbid obesity and depression in women: The Be Active Trial. International Journal of Obesity. 2013;37(11):1427-34.

Pallavicini, F., Algeri, D., Repetto, C., Gorini, A., & Riva, G. (2009). Biofeedback, virtual reality and mobile phones in the treatment of generalized anxiety disorder (gad): A phase-2 controlled clinical trial. Journal of Cyber Therapy and Rehabilitation, 2(4), 315-327. Retrieved from http://www.embase.com/search/results?subaction=viewrecord&from=export&id=L362154058

Pallavicini, F., Algeri, D., Repetto, C., Gorini, A., & Riva, G. (2009). Biofeedback, virtual reality and mobile phones in the treat-ment of generalized anxiety disorder (gad): A phase-2 controlled clinical trial. Journal of Cyber Therapy and Rehabilitation, 2(4), 315-327. Retrieved from http://www.embase.com/search/results?subaction=viewrecord&from=export&id=L362154058

Pan, W. L., Lin, L. C., Kuo, L. Y., Chiu, M. J., & Ling, P. Y. (2023). Effects of a prenatal mindfulness program on longitudinal changes in stress, anxiety, depression, and mother-infant bonding of women with a tendency to perinatal mood and anxiety disorder: a randomized controlled trial. BMC Pregnancy Childbirth, 23(1), 547.

Patel, V., Weobong, B., Weiss, H., Anand, A., Bhat, B., Katti, B., . . . Fairburn, C. (2017). The Healthy Activity Program (HAP), a lay counsellor-delivered brief psychological treatment for severe depression, in primary care in India: a randomised controlled trial. Lancet (london, england), 389(10065), 176-185.

Patsiokas, A. T., & Clum, G. A. (1985). Effects of psychotherapeutic strategies in the treatment of suicide attempters. Psychotherapy: Theory, Research, Practice, Training, 22(2), 281–290. https://doi.org/10.1037/h0085507

Pecheur DR, Edwards KJ. A comparison of secular and religious versions of cognitive therapy with depressed Christian college students. Journal of Psychology and Theology. 1984.

Peden AR, Hall LA, Rayens MK, Beebe LL. Reducing negative thinking and depressive symptoms in college women. Journal of Nursing Scholarship. 2000;32(2):145-51.

Pellas, J., Renner, F., Ji, J. L., & Damberg, M. (2022). Telephone-based behavioral activation with mental imagery for depression: A pilot randomized clinical trial in isolated older adults during the Covid-19 pandemic. Int J Geriatr Psychiatry, 37(1). doi:10.1002/gps.5646

Penckofer SM, Ferrans C, Mumby P, Byrn M, Emanuele MA, Harrison PR, et al. A psychoeducational intervention (SWEEP) for depressed women with diabetes. Annals of Behavioral Medicine. 2012;44(2):192-206.

Pendleton, M. G., & Higgins, R. L. (1983). A comparison of negative practice and systematic desensitization in the treatment of acrophobia. J Behav Ther Exp Psychiatry, 14(4), 317-323. doi:10.1016/0005-7916(83)90074-5

Perini S, Titov N, Andrews G. Clinician-assisted Internet-based treatment is effective for depression: Randomized controlled trial. Australian and New Zealand Journal of Psychiatry. 2009;43(6):571-8.

Peters E, Landau S, Mccrone P, Cooke M, Fisher P, Steel C, et al. A randomised controlled trial of cognitive behaviour therapy for psychosis in a routine clinical service. Acta Psychiatr Scand. 2010;122:302, 18.

Petersen I, Hanass Hancock J, Bhana A, Govender K. A group-based counselling intervention for depression comorbid with HIV/AIDS using a task shifting approach in South Africa: A randomized controlled pilot study. Journal of Affective Disorders. 2014;158:78-84.

Petry, N. M., Ammerman, Y., Bohl, J., Doersch, A., Gay, H., Kadden, R., ... & Steinberg, K. (2006). Cognitive-behavioral therapy for pathological gamblers. Journal of Consulting and Clinical Psychology, 74(3), 555-567.

Petry, N. M., Rash, C. J., & Alessi, S. M. (2016). A randomized controlled trial of brief interventions for problem gambling in substance abuse treatment patients. Journal of Consulting and Clinical Psychology, 84(10), 874-886.

Petry, N. M., Weinstock, J., Ledgerwood, D. M., & Morasco, B. (2008). A randomized trial of brief interventions for problem and pathological gamblers. Journal of Consulting and Clinical Psychology, 76(2), 318-328.

Petry, N. M., Weinstock, J., Morasco, B. J., & Ledgerwood, D. M. (2009). Brief motivational interventions for college student problem gamblers. Addiction, 104(9), 1569-1578.

Petterson K, Cesare S. Panic disorder: a cognitive behavioural approach to treatment. Counselling Psychology Quarterly 1996;9(2):191–201.

Pfeiffer, P. N., King, C., Ilgen, M., Ganoczy, D., Clive, R., Garlick, J., Abraham, K., Kim, H. M., Vega, E., Ahmedani, B., & Valenstein, M. (2019). Development and pilot study of a suicide prevention intervention delivered by peer support specialists. Psychological services, 16(3), 360–371. https://doi.org/10.1037/ser0000257

Philips, B., Wennberg, P., Konradsson, P., & Franck, J. (2018). Mentalization-based treatment for concurrent borderline personality disorder and substance use disorder: a randomized controlled feasibility study. European Addiction Research, 24(1), 1-8.

Pibernik-Okanovic M, Begic D, Ajdukovic D, Andrijasevic N, Metelko Z. Psychoeducation versus treatment as usual in diabetic patients with subthreshold depression: Preliminary results of a randomized controlled trial. Trials. 2009;10:78.

Pibernik-Okanović M, Hermanns N, Ajduković D, Kos J, Prašek M, Šekerija M, Lovrenčić MV. Does treatment of subsyndromal depression improve depression-related and diabetes-related outcomes? A randomised controlled comparison of psychoeducation, physical exercise and enhanced treatment as usual. Trials. 2015 Jul 15;16:305.

Piers, R. J., Farchione, T. J., Wong, B., Rosellini, A. J., & Cronin-Golomb, A. (2023). Telehealth Transdiagnostic Cognitive Behavioral Therapy for Depression in Parkinson's Disease: A Pilot Randomized Controlled Trial. Movement Disorders Clinical Practice, 10(1), 79-85. doi:10.1002/mdc3.13587

Pigeon, W. R., Funderburk, J. S., Cross, W., Bishop, T. M., & Crean, H. F. (2019). Brief CBT for insomnia delivered in primary care to patients endorsing suicidal ideation: a proof-of-concept randomized clinical trial. Translational behavioral medicine, 9(6), 1169–1177. https://doi.org/10.1093/tbm/ibz108

Pihlaja S, Lahti J, Lipsanen JO, et al. Scheduled Telephone Support for Internet Cognitive Behavioral Therapy for Depression in Patients at Risk for Dropout: Pragmatic Randomized Controlled Trial. J Med Internet Res. 2020;22(7):e15732.

Pinniger, R., Brown, R. F., Thorsteinsson, E. B., & McKinley, P. (2012). Argentine tango dance compared to mindfulness meditation and a waiting-list control: A randomised trial for treating depression. Complement Ther Med, 20(6), 377-384. doi:10.1016/j.ctim.2012.07.003

Pishyar, R., Harris, L. M., & Menzies, R. G. (2008). Responsiveness of measures of attentional bias to clinical change in social phobia. Cogn Emot, 22, 1209-1227. https://doi.org/10.1080/02699930701686008

Pistorello, J., Fruzzetti, A. E., Maclane, C., Gallop, R., & Iverson, K. M. (2012). Dialectical behavior therapy (DBT) applied to college students: a randomized clinical trial. Journal of consulting and clinical psychology, 80(6), 982–994. https://doi.org/10.1037/a0029096

Pistorello, J., Fruzzetti, A. E., MacLane, C., Gallop, R., & Iverson, K. M. (2012). Dialectical behavior therapy (DBT) applied to college students: a randomized clinical trial. Journal of consulting and clinical psychology, 80(6), 982.

Pistorello, J., Jobes, D. A., Gallop, R., Compton, S. N., Locey, N. S., Au, J. S., Noose, S. K., Walloch, J. C., Johnson, J., Young, M., Dickens, Y., Chatham, P., & Jeffcoat, T. (2021). A Randomized Controlled Trial of the Collaborative Assessment and Management of Suicidality (CAMS) Versus Treatment as Usual (TAU) for Suicidal College Students. Archives of suicide research : official journal of the International Academy for Suicide Research, 25(4), 765–789. https://doi.org/10.1080/13811118.2020.1749742

Pitti CT, Penate W, de la Fuente J, et al. The combined use of virtual reality exposure in the treatment of agoraphobia. Actas espanolas de psiquiatria 2015;43(4):133-41.

Poleshuck EL, Gamble SA, Bellenger K, Lu N, Tu X, Sorensen S, et al. Randomized controlled trial of interpersonal psychotherapy versus enhanced treatment as usual for women with co-occurring depression and pelvic pain. Journal of psychosomatic research. 2014;77(4):264-72.

Pot AM, Bohlmeijer ET, Onrust S, Melenhorst A-S, Veerbeek M, De Vries W. The impact of life review on depression in older adults: A randomized controlled trial. International Psychogeriatrics. 2010;22(4):572-81.

Pots WT, Fledderus M, Meulenbeek PA, ten Klooster PM, Schreurs KM, Bohlmeijer ET. Acceptance and commitment therapy as a web-based intervention for depressive symptoms: Randomised controlled trial. The British Journal of Psychiatry. 2016;208(1):69-77.

Pots WTM, Meulenbeek PAM, Veehof MM, Klungers J, Bohlmeijer ET. The efficacy of mindfulness-based cognitive therapy as a public mental health intervention for adults with mild to moderate depressive symptomatology: A randomized controlled trial. PLoS One. 2014;9(10).

Pott, S. L., Kellett, S., Green, S., Daughters, S., & Delgadillo, J. (2022). Behavioral activation for depression delivered by drug and alcohol treatment workers: A pilot randomized controlled trial. J Subst Abuse Treat, 139, 108769. doi:10.1016/j.jsat.2022.108769

Power K, McGoldrick T, Brown K, et al. A controlled comparison of Eye Movement Desensitization and Reprocessing versus exposure plus cognitive restructuring versus waiting list in the treatment of post-traumatic stress disorder. Clin Psychol Psychother. 2002 Sep/Oct;9(5):299-318. doi: 10.1002/cpp.341.

Power MJ, Freeman C. A randomized controlled trial of IPT versus CBT in primary care: With some cautionary notes about handling missing values in clinical trials. Clinical Psychology and Psychotherapy. 2012;19(2):159-69.

Power, K. G., Jerrom, D. W. A., Simpson, R. J., Mitchell, M. J., & Swanson, V. (1989). A controlled comparison of Cognitive-Behaviour Therapy, Diazepam and Placebo in the management of generalized anxiety. Behavioural Psychotherapy, 17(1), 1-14. Retrieved from http://www.embase.com/search/results?subaction=viewrecord&from=export&id=L19077017

Power, K. G., Jerrom, D. W. A., Simpson, R. J., Mitchell, M. J., & Swanson, V. (1989). A controlled comparison of Cognitive-Behaviour Therapy, Diazepam and Placebo in the management of generalized anxiety. Behavioural Psychotherapy, 17(1), 1-14. Retrieved from http://www.embase.com/search/results?subaction=viewrecord&from=export&id=L19077017

Power, K. G., Simpson, R. J., Swanson, V., & Wallace, L. A. (1990). Controlled comparison of pharmacological and psychological treatment of generalized anxiety disorder in primary care. Br J Gen Pract, 40(336), 289-294. Retrieved from https://bjgp.org/content/bjgp/40/336/289.full.pdf

Power, K. G., Simpson, R. J., Swanson, V., & Wallace, L. A. (1990). Controlled comparison of pharmacological and psychological treatment of generalized anxiety disorder in primary care. Br J Gen Pract, 40(336), 289-294. Retrieved from https://bjgp.org/content/bjgp/40/336/289.full.pdf

Powers, A., Lathan, E. C., Dixon, H. D., Mekawi, Y., Hinrichs, R., Carter, S., . . . Kaslow, N. J. (2023). Primary care-based mindfulness intervention for posttraumatic stress disorder and depression symptoms among Black adults: A pilot feasibility and acceptability randomized controlled trial. Psychol Trauma, 15(5), 858-867.

Powers, M. B., Smits, J. A., & Telch, M. J. (2004). Disentangling the effects of safety-behavior utilization and safety-behavior availability during exposure-based treatment: a placebo-controlled trial. J Consult Clin Psychol, 72(3), 448-454. doi:10.1037/0022-006x.72.3.448

Prabhu S, George LS, Guruvare S, Noronha JA, Jose TT, Nayak BS, et al. Effectiveness of psychosocial education program on postnatal depression, stress, and perceived maternal parenting self-efficacy among pregnant women in South India. Patient Education and Counseling. 2025;130.

Pratt, D., Tarrier, N., Dunn, G., Awenat, Y., Shaw, J., Ulph, F., & Gooding, P. (2015). Cognitive-behavioural suicide prevention for male prisoners: a pilot randomized controlled trial. Psychological medicine, 45(16), 3441–3451. https://doi.org/10.1017/S0033291715001348

Prendergast J, Austin MP. Early childhood nurse-delivered cognitive behavioural counselling for post-natal depression. Australasian Psychiatry. 2001;9(3):255-9.

Preschl B, Maercker A, Wagner B, Forstmeier S, Banos RM, Alcaniz M, et al. Life-review therapy with computer supplements for depression in the elderly: A randomized controlled trial. Aging and Mental Health. 2012;16(8):964-74.

Price, M., & Anderson, P. L. (2011). The impact of cognitive behavioral therapy on post event processing among those with social anxiety disorder. Behav Res Ther, 49(2), 132-137. https://doi.org/10.1016/j.brat.2010.11.006

Priebe, S., Bhatti, N., Barnicot, K., Bremner, S., Gaglia, A., Katsakou, C., ... & Zinkler, M. (2012). Effectiveness and cost-effectiveness of dialectical behaviour therapy for self-harming patients with personality disorder: a pragmatic randomised controlled trial. Psychotherapy and psychosomatics, 81(6), 356-365.

Propst LR, Ostrom R, Watkins P, Dean T, Mashburn D. Comparative efficacy of religious and nonreligious cognitive-behavioral therapy for the treatment of clinical depression in religious individuals. Journal of Consulting and Clinical Psychology. 1992;60(1):94-103.

Psaros, C., Stanton, A. M., Raggio, G. A., Mosery, N., Goodman, G. R., Briggs, E. S., . . . Safren, S. A. (2022). Optimizing PMTCT Adherence by Treating Depression in Perinatal Women with HIV in South Africa: A Pilot Randomized Controlled Trial. International journal of behavioral medicine. doi:10.1007/s12529-022-10071-z

Psarraki, E. E., Bacopoulou, F., Panagoulias, E., Michou, M., Pelekasis, P., Artemiadis, A., . . . Darviri, C. (2021). The effects of Pythagorean Self-Awareness Intervention on patients with major depressive disorder: A pilot randomized controlled trial. J Psychiatr Res, 138, 326-334. doi:10.1016/j.jpsychires.2021.03.067

Puckering C, McIntosh E, Hickey A, Longford J. Mellow Babies: A group intervention for infants and mothers experiencing postnatal depression. Counselling Psychology Review. 2010;25(1):28-38.

Pugh, N. E., Hadjistavropoulos, H. D., & Dirkse, D. (2016). A Randomised Controlled Trial of Therapist-Assisted, Internet-Delivered Cognitive Behavior Therapy for Women with Maternal Depression. PLoS ONE, 11(3), e0149186.

Qiu J, Chen W, Gao X, Xu Y, Tong H, Yang M, et al. A randomized controlled trial of group cognitive behavioral therapy for Chinese breast cancer patients with major depression. Journal of psychosomatic obstetrics and gynaecology. 2013;34(2):60-7.

Raabe S, Ehring T, Marquenie L, et al. Imagery rescripting as a stand-alone treatment for posttraumatic stress disorder related to childhood abuse: a randomized controlled trial. J Behav Ther Exp Psychiatry. 2022;77:101769. doi: 10.1016/j.jbtep.2022.101769. PMID: 36113906.

Raevuori, A., Vahlberg, T., Korhonen, T., Hilgert, O., Aittakumpu-Hyden, R., & Forman-Hoffman, V. (2021). A therapist-guided smartphone app for major depression in young adults: A randomized clinical trial. J Affect Disord, 286, 228-238. doi:10.1016/j.jad.2021.02.007

Rahman A, Malik A, Sikander S, Roberts C, Creed F. Cognitive behaviour therapy-based intervention by community health workers for mothers with depression and their infants in rural Pakistan: A cluster-randomised controlled trial. Lancet. 2008;372(9642):902-9.

Rahmani, F., Abbass, A., Hemmati, A., Ghaffari, N., & Rezaei Mirghaed, S. (2020). Challenging the role of challenge in intensive short-term dynamic psychotherapy for social anxiety disorder: A randomized controlled trial. J Clin Psychol, 76(12), 2123-2132. https://doi.org/10.1002/jclp.22993

Raji Lahiji, M., Sajadian, A., Haghighat, S., Zarrati, M., Dareini, H., Raji Lahiji, M., & Razmpoosh, E. (2022). Effectiveness of logotherapy and nutrition counseling on psychological status, quality of life, and dietary intake among breast cancer survivors with depressive disorder: a randomized clinical trial. Supportive Care in Cancer. doi:10.1007/s00520-022-07237-6

Rakitzi S, Georgila P, Efthimiou K, Mueller DR. Efficacy and feasibility of the Integrated Psychological Therapy for outpatients with schizophrenia in Greece : Final results of a RCT. Psychiatry Res [Internet]. Elsevier; 2016;242:137, 43. Available from: http://dx.doi.org/10.1016/j.psychres.2016.05.039

Range, M., Stacey H. Kovac, Michelle S. Marion, L. (2000). Does writing about the bereavement lessen grief following sudden, unintentional death?. Death studies, 24(2), 115-134.

Ransom D, Heckman TG, Anderson T, Garske J, Holroyd K, Basta T. Telephone-delivered, interpersonal psychotherapy for HIV-infected rural persons with depression: A pilot trial. Psychiatric Services. 2008;59(8):871-7.

Rapee, R. M., Abbott, M. J., Baillie, A. J., & Gaston, J. E. (2007). Treatment of social phobia through pure self-help and therapist-augmented self-help. Br J Psychiatry, 191, 246-252. https://doi.org/10.1192/bjp.bp.106.028167

Rathod S, Phiri P, Harris S, Underwood C, Thagadur M, Padmanabi U, et al. Cognitive behaviour therapy for psychosis can be adapted for minority ethnic groups : A randomised controlled trial. Schizophr Res [Internet]. Elsevier B.V.; 2013;143(2, 3):319, 26. Available from: http://dx.doi.org/10.1016/j.schres.2012.11.007

Raue, P. J., Sirey, J. A., Dawson, A., Berman, J., & Bruce, M. L. (2019). Lay-delivered behavioral activation for depressed senior center clients: Pilot RCT. International journal of geriatric psychiatry, 34(11), 1715-1723. doi:10.1002/gps.5186

Raya-Tena, A., Fernández-San-Martin, M. I., Martin-Royo, J., Casañas, R., Sauch-Valmaña, G., Cols-Sagarra, C., . . . Jiménez-Herrera, M. F. (2021). Effectiveness of a Psychoeducational Group Intervention Carried Out by Nurses for Patients with Depression and Physical Comorbidity in Primary Care: Randomized Clinical Trial. Int J Environ Res Public Health, 18(6). doi:10.3390/ijerph18062948

Rector NA, Seeman M V, Segal Z V. Cognitive therapy for schizophrenia : a preliminary randomized controlled trial. Schizophr Res. 2003;63:1, 11.

Reger GM, Koenen-Woods P, Zetocha K, et al. Randomized controlled trial of prolonged exposure using imaginal exposure vs. virtual reality exposure in active duty soldiers with deployment-related posttraumatic stress disorder (PTSD). J Consult Clin. Psychol. 2016 Nov;84(11):946-59. doi: 10.1037/ccp0000134. PMID: 27606699.

Rehm LP, Kornblith SJ, O'Hara MW, Lamparski DM, Romano JM, Volkin JI. An evaluation of major components in a self-control therapy program for depression. Behavior modification. 1981;5(4):459-89.

Reimer, S. G., & Moscovitch, D. A. (2015). The impact of imagery rescripting on memory appraisals and core beliefs in social anxiety disorder. Behav Res Ther, 75, 48-59. https://doi.org/10.1016/j.brat.2015.10.007

Reinecke A, Waldenmaier L, Cooper MJ, Harmer CJ. Changes in automatic threat processing precede and predict clinical changes with exposure-based cognitive-behavior therapy for panic disorder.

Reins, J. A., Boß, L., Lehr, D., Berking, M., & Ebert, D. D. (2019). The more I got, the less I need? Efficacy of Internet-based guided self-help compared to online psychoeducation for major depressive disorder. Journal of, 246, 695-705. doi:10.1016/j.jad.2018.12.065

Reneses, B., Galián, M., Serrano, R., Figuera, D., Fernandez del Moral, A., López-Ibor, J. J., ... & Trujillo, M. (2013). A new time limited psychotherapy for BPD: preliminary results of a randomized and controlled trial. Actas Espanolas de Psiquiatria, 41(3).

Resick PA, Nishith P, Weaver TL, et al. A comparison of cognitive-processing therapy with prolonged exposure and a waiting condition for the treatment of chronic posttraumatic stress disorder in female rape victims. J Consult Clin Psychol. 2002 Aug;70(4):867-79. PMID: 12182270.

Richards D, Timulak L, O'Brien E, Hayes C, Vigano N, Sharry J, et al. A randomized controlled trial of an internet-delivered treatment: Its potential as a low-intensity community intervention for adults with symptoms of depression. Behaviour Research and Therapy. 2015;75:20-31.

Richards JC, Klein B, Austin DW. Internet cognitive behavioural therapy for panic disorder: does the inclusion of stress management information improve end-state functioning? Clinical psychologist 2006;10(1):2-15.

Richards SH, Dickens C, Anderson R, et al. Assessing the effectiveness of Enhanced Psychological Care for patients with depressive symptoms attending cardiac rehabilitation compared with treatment as usual (CADENCE): a pilot cluster randomised controlled trial. Trials 2018; 19(1).

Richards, D., Timulak, L., Rashleigh, C., McLoughlin, O., Colla, A., Joyce, C., . . . Anderson-Gibbons, M. (2016). Effectiveness of an internet-delivered intervention for generalized anxiety disorder in routine care: A randomised controlled trial in a student population. Internet Interventions, 6, 80-88. doi:10.1016/j.invent.2016.10.003

Richards, D., Timulak, L., Rashleigh, C., McLoughlin, O., Colla, A., Joyce, C., . . . Anderson-Gibbons, M. (2016). Effectiveness of an internet-delivered intervention for generalized anxiety disorder in routine care: A randomised controlled trial in a student population. Internet Interventions, 6, 80-88. doi:10.1016/j.invent.2016.10.003

Riddle-Walker, L., Veale, D., Chapman, C., Ogle, F., Rosko, D., Najmi, S., . . . Hicks, T. (2016). Cognitive behaviour therapy for specific phobia of vomiting (Emetophobia): A pilot randomized controlled trial. J Anxiety Disord, 43, 14-22. doi:10.1016/j.janxdis.2016.07.005

Rief W, Bleichhardt G, Dannehl K, Euteneuer F, Wambach K. Comparing the Efficacy of CBASP with Two Versions of CBT for Depression in a Routine Care Center: a Randomized Clinical Trial. Psychotherapy and psychosomatics 2018;

Ritvo, P., et al. (2021). "Online Mindfulness-Based Cognitive Behavioral Therapy Intervention for Youth With Major Depressive Disorders: Randomized Controlled Trial." J Med Internet Res 23(3): e24380.

Rizvi SJ, Zaretsky A, Schaffer A, Levitt A. Is immediate adjunctive CBT more beneficial than delayed CBT in treating depression?: A Pilot Study. Journal of Psychiatric Practice. 2015;21(2):107-13.

Roberts DL, Combs DR, Willoughby M, Mintz J, Gibson C, Rupp B, et al. A randomized , controlled trial of Social Cognition and Interaction Training (SCIT) for outpatients with schizophrenia spectrum disorders. Br J Clin Psychol. 2014;53:281, 98.

Robillard, G., Bouchard, S., Dumoulin, S., Guitard, T., & Klinger, E. (2010). Using virtual humans to alleviate social anxiety: preliminary report from a comparative outcome study. Stud Health Technol Inform, 154, 57-60. https://doi.org/10.3233/978-1-60750-561-7-57

Robinson, E., Titov, N., Andrews, G., McIntyre, K., Schwencke, G., & Solley, K. (2010). Internet treatment for generalized anxiety disorder: a randomized controlled trial comparing clinician vs. technician assistance. PLoS One, 5(6), e10942. doi:10.1371/journal.pone.0010942

Robinson, E., Titov, N., Andrews, G., McIntyre, K., Schwencke, G., & Solley, K. (2010). Internet treatment for generalized anxie-ty disorder: a randomized controlled trial comparing clinician vs. technician assistance. PLoS One, 5(6), e10942. doi:10.1371/journal.pone.0010942

Roemer, L., Orsillo, S. M., & Salters-Pedneault, K. (2008). Efficacy of an acceptance-based behavior therapy for generalized anxiety disorder: evaluation in a randomized controlled trial. J Consult Clin Psychol, 76(6), 1083-1089. doi:10.1037/a0012720

Roemer, L., Orsillo, S. M., & Salters-Pedneault, K. (2008). Efficacy of an acceptance-based behavior therapy for generalized anxiety disorder: evaluation in a randomized controlled trial. J Consult Clin Psychol, 76(6), 1083-1089. doi:10.1037/a0012720

Rogiers, R., Baeken, C., Van den Abbeele, D., Watkins, E. R., Remue, J., Colman, R., . . . Lemmens, G. M. D. (2021). Group Intervention 'Drop it!' Decreases Repetitive Negative Thinking in Major Depressive Disorder and/or Generalized Anxiety Disorder: A Randomised Controlled Study. Cognitive Therapy and Research. doi:10.1007/s10608-021-10240-6

Rogiers, R., Baeken, C., Van den Abbeele, D., Watkins, E. R., Remue, J., Colman, R., . . . Lemmens, G. M. D. (2021). Group Inter-vention 'Drop it!' Decreases Repetitive Negative Thinking in Major Depressive Disorder and/or Generalized Anxiety Dis-order: A Randomised Controlled Study. Cognitive Therapy and Research. doi:10.1007/s10608-021-10240-6

Rohan KJ, Roecklein KA, Lindsey KT, Johnson LG, Lippy RD, Lacy TJ, et al. A randomized controlled trial of cognitive-behavioral therapy, light therapy, and their combination for seasonal affective disorder. Journal of Consulting and Clinical Psychology. 2007;75(3):489-500.

Rohde P, Stice E, Shaw H, Gau JM. Pilot trial of a dissonance-based cognitive-behavioral group depression prevention with college students. Behaviour Research and Therapy. 2016;82:21-7.

Rohde, P., Stice, E., Shaw, H., & Gau, J. M. (2014). Cognitive-behavioral group depression prevention compared to bibliotherapy and brochure control: Nonsignificant effects in pilot effectiveness trial with college students. Behaviour Research and Therapy, 55(1), 48-53.

Rohricht F, Papadopoulos N, Priebe S. An exploratory randomized controlled trial of body psychotherapy for patients with chronic depression. Journal of Affective Disorders. 2013;151(1):85-91.

Rolvien, L., Buddeberg, L., Gehlenborg, J., Borsutzky, S., & Moritz, S. (2024). A Self-Guided Internet-Based Intervention for the Reduction of Gambling Symptoms: A Randomized Clinical Trial. JAMA Network Open, 7(6), e2417282-e2417282.

Rosen, L. A., Weinstock, J., & Peter, S. C. (2020). A Randomized Clinical Trial Exploring Gambling Attitudes, Barriers to Treatment, and Efficacy of a Brief Motivational Intervention Among Ex-Offenders with Disordered Gambling. Journal of Forensic Sciences, 65(5), 1646-1655.

Rosner, R., Pfoh, G., Kotoučová, M., & Hagl, M. (2014). Efficacy of an outpatient treatment for prolonged grief disorder: A randomized controlled clinical trial. Journal of Affective Disorders, 167, 56-63.

Ross M, Scott M. An evaluation of the effectiveness of individual and group cognitive therapy in the treatment of depressed patients in an inner city health centre. The Journal of the Royal College of General Practitioners. 1985;35(274):239-42.

Rothbaum BO, Astin MC, Marsteller F. Prolonged exposure versus Eye Movement Desensitization and Reprocessing (EMDR) for PTSD rape victims. J Trauma Stress. 2005 Dec;18(6):607-16. doi: 10.1002/jts.20069. PMID: 16382428.

Rothbaum BO. A controlled study of Eye Movement Desensitization and Reprocessing in the treatment of posttraumatic stress disordered sexual assault victims. Bull Menninger Clin. 1997 Summer;61(3):317-34. PMID: 9260344.

Rothbaum, B. O., Anderson, P., Zimand, E., Hodges, L., Lang, D., & Wilson, J. (2006). Virtual reality exposure therapy and standard (in vivo) exposure therapy in the treatment of fear of flying. Behav Ther, 37(1), 80-90. doi:10.1016/j.beth.2005.04.004

Rothbaum, B. O., Hodges, L. F., Kooper, R., Opdyke, D., Williford, J. S., & North, M. (1995). Effectiveness of computer-generated (virtual reality) graded exposure in the treatment of acrophobia. Am J Psychiatry, 152(4), 626-628. doi:10.1176/ajp.152.4.626

Rothbaum, B. O., Hodges, L., Smith, S., Lee, J. H., & Price, L. (2000). A controlled study of virtual reality exposure therapy for the fear of flying. J Consult Clin Psychol, 68(6), 1020-1026. doi:10.1037//0022-006x.68.6.1020

Roy-Byrne PP, Craske MG, Stein MB, et al. A randomized effectiveness trial of cognitive-behavioral therapy and medication for primary care panic disorder. Archives of general psychiatry 2005;62(3):290-8.

Roy-Byrne, P., Craske, M. G., Sullivan, G., Rose, R. D., Edlund, M. J., Lang, A. J., ... & Stein, M. B. (2010). Delivery of evidence-based treatment for multiple anxiety disorders in primary care: a randomized controlled trial. Jama, 303(19), 1921-1928.

Roy-Byrne, P., Craske, M. G., Sullivan, G., Rose, R. D., Edlund, M. J., Lang, A. J., ... & Stein, M. B. (2010). Delivery of evidence-based treatment for multiple anxiety disorders in primary care: a randomized controlled trial. Jama, 303(19), 1921-1928.

Rudd, M. D., Bryan, C. J., Wertenberger, E. G., Peterson, A. L., Young- McCaughan, S., Mintz, J., Williams, S. R., Arne, K. A., Breitbach, J., Delano, K., Wilkinson, E., & Bruce, T. O. (2015). Brief cognitive-behavioral therapy effects on post-treatment suicide attempts in a military sample: results of a randomized clinical trial with 2-year follow-up. The American journal of psychiatry, 172(5), 441–449. https://doi.org/10.1176/appi.ajp.2014.14070843

Rudd, M. D., Rajab, M. H., Orman, D. T., Joiner, T., Stulman, D. A., & Dixon, W. (1996). Effectiveness of an outpatient intervention targeting suicidal young adults: preliminary results. Journal of consulting and clinical psychology, 64(1), 179–190. https://doi.org/10.1037//0022-006x.64.1.179

Rude SS. Relative benefits of assertion or cognitive self-control treatment for depression as a function of proficiency in each domain. Journal of Consulting and Clinical Psychology. 1986;54(3):390-4.

Rupp, C., Jürgens, C., Doebler, P., Andor, F., & Buhlmann, U. (2019). A randomized waitlist-controlled trial comparing detached mindfulness and cognitive restructuring in obsessive-compulsive disorder. PLoS One, 14(3), e0213895.

Rus-Calafell M, GutieÃÅrrez-Maldonado J, Ortega-Bravo M, Ribas-SabateÃÅ J, Caqueo-UriÃÅzar A. A brief cognitive , behavioural social skills training for stabilised outpatients with schizophrenia : A preliminary study. Schizophr Res [Internet]. Elsevier B.V.; 2013;143(2, 3):327, 36. Available from: http://dx.doi.org/10.1016/j.schres.2012.11.014

Russell, A., Gaunt, D. M., Cooper, K., Barton, S., Horwood, J., Kessler, D., . . . et al. (2019). The feasibility of low-intensity psychological therapy for depression co-occurring with autism in adults: the Autism Depression Trial (ADEPT) â€" a pilot randomised controlled trial. Autism. doi:10.1177/1362361319889272

Russman Block, S., Norman, L. J., Zhang, X., Mannella, K. A., Yang, H., Angstadt, M., . . . Fitzgerald, K. D. (2023). Resting-State Connectivity and Response to Psychotherapy Treatment in Adolescents and Adults With OCD: A Randomized Clinical Trial. Am J Psychiatry, 180(1), 89-99. doi:10.1176/appi.ajp.21111173

Ruwaard J, Broeksteeg J, Schrieken B, et al. Web-based therapist-assisted cognitive behavioral treatment of panic symptoms: a randomized controlled trial with a three-year follow-up. Journal of anxiety disorders 2010;24(4):387-96.

Ruwaard J, Schrieken B, Schrijver M, Broeksteeg J, Dekker J, Vermeulen H, et al. Standardized web-based cognitive behavioural therapy of mild to moderate depression: A randomized controlled trial with a long-term follow-up. Cognitive Behaviour Therapy. 2009;38(4):206-21.

Ruzickova, T., Carson, J., Murphy, S., & Harmer, C. (2021). P.230 Effects of online behavioural activation on depression during covid-19. European neuropsychopharmacology, 44, S36-S37. doi:10.1016/j.euroneuro.2021.01.059

Ryberg, W., Zahl, P. H., Diep, L. M., Landrø, N. I., & Fosse, R. (2019). Managing suicidality within specialized care: A randomized controlled trial. Journal of affective disorders, 249, 112–120. https://doi.org/10.1016/j.jad.2019.02.022

Sadler, P., McLaren, S., Klein, B., Harvey, J., & Jenkins, M. (2018). Cognitive behavior therapy for older adults with insomnia and depression: a randomized controlled trial in community mental health services. Sleep, 41(8) (no pagination). doi:10.1093/sleep/zsy106

Safren SA, Bedoya CA, O'Cleirigh C, Biello KB, Pinkston MM, Stein MD, et al. Cognitive behavioural therapy for adherence and depression in patients with HIV: A three-arm randomised controlled trial. The Lancet HIV. 2016;3(11):e529-e38.

Safren SA, Gonzalez JS, Wexler DJ, Psaros C, Delahanty LM, Blashill AJ, et al. A randomized controlled trial of cognitive behavioral therapy for adherence and depression (CBT-AD) in patients with uncontrolled type 2 diabetes. Diabetes care. 2014;37(3):625-33.

Safren SA, O'Cleirigh C, Tan JY, Raminani SR, Reilly LC, Otto MW, et al. A randomized controlled trial of cognitive behavioral therapy for adherence and depression (CBT-AD) in HIV-infected individuals. Health Psychology. 2009;28(1):1-10.

Safren, S. A., O'Cleirigh, C., Andersen, L. S., Magidson, J. F., Lee, J. S., Bainter, S. A., . . . Joska, J. A. (2021). Treating depression and improving adherence in HIV care with task-shared cognitive behavioural therapy in Khayelitsha, South Africa: a randomized controlled trial. J Int AIDS Soc, 24(10), e25823. doi:10.1002/jia2.25823

Saisanan Na Ayudhaya, W., Pityaratstian, N., & Jiamjarasrangsi, W. (2020). Effectiveness of Behavioral Activation in Treating Thai Older Adults with Subthreshold Depression Residing in the Community. Clin Interv Aging, 15, 2363-2374. doi:10.2147/cia.S274262

Salaberria, K., & Echeburua, E. (1998). Long-term outcome of cognitive therapy's contribution to self-exposure in vivo to the treatment of generalized social phobia. Behav Modif, 22(3), 262-284. https://doi.org/10.1177/01454455980223003

Salamanca-Sanabria A, Richards D, Timulak L, et al. A culturally adapted cognitive behavioral internet-delivered intervention for depressive symptoms: Randomized controlled trial. JMIR Mental Health. 2020;7(1).

Saloheimo HP, Markowitz J, Saloheimo TH, Laitinen JJ, Sundell J, Huttunen MO, et al. Psychotherapy effectiveness for major depression: A randomized trial in a Finnish community. BMC psychiatry. 2016;16:131.

Samaraweera, S., Sivayogan, S., Sumathipala, A., Bhugra, D., & Siribaddana,S. (2007). RCT of Cognitive Behaviour Therapy in active suicidal ideation-as feasibility study in Sri Lanka. The European Journal of Psychiatry, 21(3), 175–178. https://doi.org/10.4321/S0213-61632007000300001

Sanabria-Mazo JP, Colomer-Carbonell A, Borràs X, Castaño-Asins JR, McCracken LM, Montero-Marin J, et al. Efficacy of Videoconference Group Acceptance and Commitment Therapy (ACT) and Behavioral Activation Therapy for Depression (BATD) for Chronic Low Back Pain (CLBP) Plus Comorbid Depressive Symptoms: A Randomized Controlled Trial (IMPACT Study). J Pain. 2023;24(8):1522-40.

Sánchez P, PenÃÉa J, Bengoetxea E, Ojeda N, ElizagaÃÅrate E, Ezcurra J, et al. Improvements in negative symptoms and functional outcome after a new generation cognitive remediation program: A randomized controlled trial. Schizophr Bull. 2014;40(3):707, 15.

Sander LB, Paganini S, Terhorst Y, et al. Effectiveness of a Guided Web-Based Self-help Intervention to Prevent Depression in Patients with Persistent Back Pain: The PROD-BP Randomized Clinical Trial. JAMA Psychiatry. 2020;77(10):1001-1011.

Sanz Cruces, J. M., García Cuenca, I. M., Lacomba-Trejo, L., Cuquerella Adell, M. Á., Cano Navarro, I., Ferrandis Cortés, M., Jordá Carreres, E., & Carbajo Álvarez, E. (2018). Group Therapy for Patients with Adjustment Disorder in Primary Care. The Spanish journal of psychology, 21, E50. https://doi.org/10.1017/sjp.2018.51
Savard J, Simard S, Giguere I, Ivers H, Morin CM, Maunsell E, et al. Randomized clinical trial on cognitive therapy for depression in women with metastatic breast cancer: Psychological and immunological effects. Palliative and Supportive Care. 2006;4(3):219-37.
Savari, Y., Mohagheghi, H., & Petrocchi, N. (2021). A preliminary investigation on the effectiveness of compassionate mind training for students with major depressive disorder: A randomized controlled trial. Mindfulness, 12(5), 1159-1172. doi:10.1007/s12671-020-01584-3
Scazufca, M., Nakamura, C. A., Seward, N., Moreno-Agostino, D., van de Ven, P., Hollingworth, W., . . . Araya, R. (2022). A task-shared, collaborative care psychosocial intervention for improving depressive symptomatology among older adults in a socioeconomically deprived area of Brazil (PROACTIVE): a pragmatic, two-arm, parallel-group, cluster-randomised controlled trial. Lancet Healthy Longev, 3(10), e690-e702. doi:10.1016/s2666-7568(22)00194-5
Schlicker S, Baumeister H, Buntrock C, et al. A web- And mobile-based intervention for comorbid, recurrent depression in patients with chronic back pain on sick leave (get.back): Pilot randomized controlled trial on feasibility, user satisfaction, and effectiveness. JMIR Mental Health. 2020;7(4).
Schmidt MM, Miller WR. Amount of therapist contact and outcome in a multidimensional depression treatment program. Acta Psychiatrica Scandinavica. 1983;67(5):319-32.
Schmidt NB, Staab JP, Trakowski JH, Jr., et al. Efficacy of a brief psychosocial treatment for panic disorder in an active duty sample: implications for military readiness. Military medicine 1997;162(2):123-9.
Schmidt NB, Trakowski JH, Staab JP. Extinction of panicogenic effects of a 35% CO2 challenge in patients with panic disorder. Journal of Abnormal Psychology 1997; 106(4):630–8.
Schramm E, Mack S, Thiel N, Jenkner C, Elsaesser M, Fangmeier T. Interpersonal psychotherapy vs treatment as usual for major depression related to work stress: A pilot randomized controlled study. Frontiers in Psychiatry. 2020;11.
Schrank B, Brownell T, Jakaite Z, Larkin C, Pesola F, Riches S, Tylee A, Slade M. Evaluation of a _positive psychotherapy group intervention for people with psychosis: pilot randomised controlled trial. Epid Psychiatric Sci. 2016; 25: 235, 246.
Schulberg HC, Block MR, Madonia MJ, Scott CP, Rodriguez E, Imber SD, et al. Treating major depression in primary care practice. Eight-month clinical outcomes. Archives of General Psychiatry. 1996;53(10):913-9.
Schulz, A., Stolz, T., Vincent, A., Krieger, T., Andersson, G., & Berger, T. (2016). A sorrow shared is a sorrow halved? A three-arm randomized controlled trial comparing internet-based clinician-guided individual versus group treatment for social anxiety disorder. Behav Res Ther, 84, 14-26. https://doi.org/10.1016/j.brat.2016.07.001
Schuster, R., Leitner, I., Carlbring, P., & Laireiter, A.-R. (2017). Exploring blended group interventions for depression: randomised controlled feasibility study of a blended computer- and multimedia-supported psychoeducational group intervention for adults with depressive symptoms. Internet Interventions, 8, 63-71.
Schweden, T. L. K., Pittig, A., Brauer, D., Klumbies, E., Kirschbaum, C., & Hoyer, J. (2016). Reduction of depersonalization during social stress through cognitive therapy for social anxiety disorder: A randomized controlled trial. J Anxiety Disord, 43, 99-105. https://doi.org/10.1016/j.janxdis.2016.09.005
Scogin F, Hamblin D, Beutler L. Bibliotherapy for depressed older adults: A self-help alternative. The Gerontologist. 1987;27(3):383-7.
Scogin F, Jamison C, Gochneaur K. Comparative efficacy of cognitive and behavioral bibliotherapy for mildly and moderately depressed older adults. Journal of Consulting and Clinical Psychology. 1989;57(3):403-7.
Scogin F, Lichstein K, DiNapoli EA, et al. Effects of integrated telehealth-delivered cognitive-behavioral therapy for depression and insomnia in rural older adults. Journal of psychotherapy integration 2018; 28(3): 292-309.

Scott AI, Freeman CP. Edinburgh primary care depression study: Treatment outcome, patient satisfaction, and cost after 16 weeks. BMJ. 1992;304(6831):883-7.
Scott C, Tacchi MJ, Jones R, Scott J. Acute and one-year outcome of a randomised controlled trial of brief cognitive therapy for major depressive disorder in primary care. The British Journal of Psychiatry. 1997;171:131-4.
Scott MJ, Stradling SG. Group cognitive therapy for depression produces clinically significant reliable change in community-based settings. Behavioural Psychotherapy. 1990;18(01):1-19.
Sedghy, Z., Yoosefi, N., & Navidian, A. (2020). The effect of motivational interviewing-based training on the rate of using mental health services and intensity of suicidal ideation in individuals with suicide attempt admitted to the emergency department. Journal of education and health promotion, 9.
Segal, Z. V., Dimidjian, S., Beck, A., Boggs, J. M., Vanderkruik, R., Metcalf, C. A., . . . Levy, J. (2020). Outcomes of Online Mindfulness-Based Cognitive Therapy for Patients with Residual Depressive Symptoms: A Randomized Clinical Trial. JAMA Psychiatry, 77(6), 563-573. doi:10.1001/jamapsychiatry.2019.4693
Segre LS, Brock RL, O'Hara MW. Depression treatment for impoverished mothers by point-of-care providers: A randomized controlled trial. Journal of Consulting and Clinical Psychology. 2015;83(2):314-24.
Selmi PM, Klein MH, Greist JH, Sorrell SP, Erdman HP. Computer-administered cognitive-behavioral therapy for depression. American Journal of Psychiatry. 1990;147(1):51-6.
Sensky T, Turkington D, Kingdon D, Scott JL, Scott J, Siddle R, et al. A randomized controlled trial of cognitive-behavioral therapy for persistent symptoms in schizophrenia resistant to medication. Arch Gen Psychiatry. 2000;57:165, 72.
Serfaty MA, Haworth D, Blanchard M, Buszewicz M, Murad S, King M. Clinical effectiveness of individual cognitive behavioral therapy for depressed older people in primary care: A randomized controlled trial. Archives of General Psychiatry. 2009;66(12):1332-40.
Serfaty, M., King, M., Nazareth, I., Moorey, S., Aspden, T., Tookman, A., . . . Jones, L. (2019). Manualised cognitive and behavioural therapy in treating depression in advanced cancer: the CanTalk RCT. Health Technology Assessment, 23(19), 1-106. doi:10.3310/hta23190
Serrano JP, Latorre JM, Gatz M, Montanes J. Life review therapy using autobiographical retrieval practice for older adults with depressive symptomatology. Psychology and Aging. 2004;19(2):270-7.
Serrano Selva JP, Latorre Postigo JM, Ros Segura L, Navarro Bravo B, Aguilar Corcoles MJ, Nieto Lopez M, et al. Life review therapy using autobiographical retrieval practice for older adults with clinical depression. Psicothema. 2012;24(2):224-9.
Shan, Q., Xinxin, S., Zhijuan, X., Rongjing, D., & Minjie, Z. (2022). Effects of Cognitive Behavior Therapy on Depression, Illness Perception, and Quality of Life in Atrial Fibrillation Patients. Frontiers in Psychiatry, 13. doi:10.3389/fpsyt.2022.830363
Shapiro E, Laub B. Early EMDR intervention following a community critical incident: a randomized clinical trial. Journal of EMDR Practice and Research. 2015 Feb;9(1):17-27. doi: 10.1891/1933-3196.9.1.17.
Sharp DM, Power KG, Simpson RJ, et al. Global measures of outcome in a controlled comparison of pharmacological and psychological treatment of panic disorder and agoraphobia in primary care. The British journal of general practice 1997;47(416):150-5.
Sharp DM, Power KG, Swanson V. A comparison of the efficacy and acceptability of group versus individual cognitive behaviour therapy in the treatment of panic disorder and agoraphobia in primary care. Clinical Psychology and Psychotherapy 2004;11(2):73-82.
Shaw BF. Comparison of cognitive therapy and behavior therapy in the treatment of depression. Journal of Consulting and Clinical Psychology. 1977;45(4):543.
Shaygan, M., Sheybani Negad, S., & Motazedian, S. (2022). The effect of combined sertraline and positive psychotherapy on hopelessness and suicidal ideation among patients with major depressive disorder: a randomized controlled trial. The Journal of Positive Psychology, 17(5), 655-664.
Shear MK, Houck P, Greeno C, et al. Emotion-focused psychotherapy for patients with panic disorder. The American journal of psychiatry 2001;158(12):1993-8.
Shear MK, Pilkonis PA, Cloitre M, et al. Cognitive behavioral treatment compared with nonprescriptive treatment of panic disorder. Archives of general psychiatry 1994;51(5):395-401.

Shear, M. K., Reynolds, C. F., Simon, N. M., Zisook, S., Wang, Y., Mauro, C., ... & Skritskaya, N. (2016). Optimizing treatment of complicated grief: A randomized clinical trial. JAMA psychiatry, 73(7), 685-694.

Sheaves, B., Holmes, E. A., Rek, S., Taylor, K. M., Nickless, A., Waite, F., Germain, A., Espie, C. A., Harrison, P. J., Foster, R., & Freeman, D. (2019). Cognitive Behavioural Therapy for Nightmares for Patients with Persecutory Delusions (Nites): An Assessor-Blind, Pilot Randomized Controlled Trial. Canadian journal of psychiatry. Revue canadienne de psychiatrie, 64(10), 686–696. https://doi.org/10.1177/0706743719847422

Sheeber LB, Seeley JR, Feil EG, Davis B, Sorensen E, Kosty DB, et al. Development and pilot evaluation of an Internet-facilitated cognitive-behavioral intervention for maternal depression. Journal of Consulting and Clinical Psychology. 2012;80(5):739-49.

Sheeber, L., Feil, E., Seeley, J., Leve, C., Gau, J., Davis, B., . . . Allan, S. (2017). Mom-net: evaluation of an internet-facilitated cognitive behavioral intervention for low-income depressed mothers. Journal of Consulting and Clinical Psychology, 85(4), 355-366.

Shih, V. W. Y., Chan, W. C., Tai, O. K., Wong, H. L., Cheng, C. P. W., & Wong, C. S. M. (2021). Mindfulness-Based Cognitive Therapy for Late-Life Depression: a Randomised Controlled Trial. East Asian Arch Psychiatry, 31(2), 27-35. doi:10.12809/eaap2075

Sikander, S., Ahmad, I., Atif, N., Zaidi, A., Vanobberghen, F., Weiss, H. A., . . . Rahman, A. (2019). Delivering the Thinking Healthy Programme for perinatal depression through volunteer peers: a cluster randomised controlled trial in Pakistan. The Lancet Psychiatry, 6(2), 128-139. doi:10.1016/S2215-0366(18)30467-X

Sikkema, K. J., Hansen, N. B., Kochman, A., Tate, D. C., & Difranceisco, W. (2004). Outcomes from a randomized controlled trial of a group intervention for HIV positive men and women coping with AIDS-related loss and bereavement. Death studies, 28(3), 187-209.

Silfvernagel K, Carlbring P, Kabo J, et al. Individually tailored internet-based treatment for young adults and adults with panic attacks: randomized controlled trial. Journal of medical Internet research 2012;14(3):e65.

Simoni JM, Wiebe JS, Sauceda JA, Huh D, Sanchez G, Longoria V, et al. A preliminary RCT of CBT-AD for adherence and depression among HIV-positive Latinos on the U.S.-Mexico border: The Nuevo Dia study. AIDS and behavior. 2013;17(8):2816-29.

Simpson S, Corney R, Beecham J. A randomized controlled trial to evaluate the effectiveness and cost-effectiveness of psychodynamic counselling for general practice patients with chronic depression. Psychological Medicine. 2003;33(2):229-39.

Simpson TL, Kaysen DL, Fleming CB, et al. Cognitive processing therapy or relapse prevention for comorbid posttraumatic stress disorder and alcohol use disorder: a randomized clinical trial. PLoS One. 2022;17(11):e0276111. doi: 10.1371/journal.pone.0276111. PMID: 36445895.

Simpson, G. K., Tate, R. L., Whiting, D. L., & Cotter, R. E. (2011). Suicide prevention after traumatic brain injury: a randomized controlled trial of a program for the psychological treatment of hopelessness. The Journal of head trauma rehabilitation, 26(4), 290–300. Simon, G. E., Beck, A., Rossom, R., Richards, J., Kirlin, B., King, D., Shulman, L., Ludman, E. J., Penfold, R., Shortreed, S. M., & Whiteside, U. (2016). Population-based outreach versus care as usual to prevent suicide attempt: study protocol for a randomized controlled trial. Trials, 17(1), 452. https://doi.org/10.1186/s13063-016-1566-z159.https://doi.org/10.1097/HTR.0b013e3182225250

Simson U, Nawarotzky U, Friese G, Porck W, Schottenfeld-Naor Y, Hahn S, et al. Psychotherapy intervention to reduce depressive symptoms in patients with diabetic foot syndrome. Diabetic Medicine. 2008;25(2):206-12.

Sinniah, A., Oei, T., Maniam, T., & Subramaniam, P. (2017). Positive effects of Individual Cognitive Behavior Therapy for patients with unipolar mood disorders with suicidal ideation in Malaysia: A randomised controlled trial. Psychiatry research, 254, 179–189. https://doi.org/10.1016/j.psychres.2017.04.026

Sinniah, A., Oei, T., Maniam, T., & Subramaniam, P. (2017). Positive effects of Individual Cognitive Behavior Therapy for patients with unipolar mood disorders with suicidal ideation in Malaysia: a randomised controlled trial. Psychiatry research, 254, 179-189.

Sirey, J. A., Solomonov, N., Guillod, A., Zanotti, P., Lee, J., Soliman, M., & Alexopoulos, G. S. (2021). PROTECT: a novel psychotherapy for late-life depression in elder abuse victims. Int Psychogeriatr, 33(5), 521-525. doi:10.1017/s1041610221000430

Slee, N., Garnefski, N., van der Leeden, R., Arensman, E., & Spinhoven, P. (2008). Cognitive-behavioural intervention for self-harm: randomised controlled trial. The British journal of psychiatry : the journal of mental science, 192(3), 202–211. https://doi.org/10.1192/bjp.bp.107.037564

Slesnick, N., Zhang, J., Feng, X., Wu, Q., Walsh, L., & Granello, D. H. (2020). Cognitive therapy for suicide prevention: A randomized pilot with suicidal youth experiencing homelessness. Cognitive Therapy and Research, 44, 402-411.

Sloan DM, Marx BP, Bovin MJ, et al. Written exposure as an intervention for PTSD: a randomized clinical trial with motor vehicle accident survivors. Behav Res Ther. 2012 Oct;50(10):627-35. doi: 10.1016/j.brat.2012.07.001. PMID: 22863540.

Sloane R, Staples F, Schneider L. Interpersonal therapy versus nortriptyline for depression in the elderly. Clinical and Pharmacological Studies in Psychiatric Disorders. 1985:344-6.

Smit A, Kluiter H, Conradi HJ, Meer K, Tiemens BG, Jenner JA, et al. Short-term effects of enhanced treatment for depression in primary care: Results from a randomized controlled trial. Psychological Medicine. 2006;36(1):15-26.

Smith, J., Newby, J. M., Burston, N., Murphy, M. J., Michael, S., Mackenzie, A., . . . Andrews, G. (2017). Help from home for depression: A randomised controlled trial comparing internet-delivered cognitive behaviour therapy with bibliotherapy for depression. Internet Interventions, 9, 25-37.

So, R., Furukawa, T. A., Matsushita, S., Baba, T., Matsuzaki, T., Furuno, S., ... & Higuchi, S. (2020). Unguided chatbot-delivered cognitive behavioural intervention for problem gamblers through messaging app: a randomised controlled trial. Journal of Gambling Studies, 36(4), 1391-1407.

Solis, E. C., Carlier, I. V. E., Kamminga, N. G. A., Giltay, E. J., & van Hemert, A. M. (2024). The clinical effectiveness of a self-management intervention for patients with persistent depressive disorder and their partners/caregivers: results from a multicenter, pragmatic randomized controlled trial. Trials, 25(1). doi:https://doi.org/10.1186/s13063-024-08033-9

Songprakun W, McCann TV. Evaluation of a cognitive behavioural self-help manual for reducing depression: A randomized controlled trial. Journal of psychiatric and mental health nursing. 2012;19(7):647-53.

Spek V, Nyklicek I, Smits N, Cuijpers P, Riper H, Keyzer J, et al. Internet-based cognitive behavioural therapy for subthreshold depression in people over 50 years old: A randomized controlled clinical trial. Psychological Medicine. 2007;37(12):1797-806.

Spence J, Titov N, Dear BF, et al. Randomized controlled trial of internet-delivered cognitive behavioral therapy for posttraumatic stress disorder. Depress Anxiety. 2011 Jul;28(7):541-50. doi: 10.1002/da.20835. PMID: 21721073.

Spinelli MG, Endicott J, Leon AC, Goetz RR, Kalish RB, Brustman LE, et al. A controlled clinical treatment trial of interpersonal psychotherapy for depressed pregnant women at 3 New York City sites. Journal of Clinical Psychiatry. 2013;74(4):393-9.

Spinelli MG, Endicott J. Controlled clinical trial of interpersonal psychotherapy versus parenting education program for depressed pregnant women. American Journal of Psychiatry. 2003;160(3):555-62.

Springer, T., Lohr, N. E., Buchtel, H. A., & Silk, K. R. (1996). A preliminary report of short-term cognitive-behavioral group therapy for inpatients with personality disorders. The Journal of psychotherapy practice and research, 5(1), 57–71.

Spruill, T. M., Friedman, D., Diaz, L., Butler, M. J., Goldfeld, K. S., O'Kula, S., . . . Devinsky, O. (2021). Telephone-based depression self-management in Hispanic adults with epilepsy: a pilot randomized controlled trial. Transl Behav Med, 11(7), 1451-1460. doi:10.1093/tbm/ibab045

Stangier, U., Heidenreich, T., Peitz, M., Lauterbach, W., & Clark, D. M. (2003). Cognitive therapy for social phobia: individual versus group treatment. Behav Res Ther, 41(9), 991-1007. https://doi.org/10.1016/s0005-7967(02)00176-6

Stangier, U., Schramm, E., Heidenreich, T., Berger, M., & Clark, D. M. (2011). Cognitive therapy vs interpersonal psychotherapy in social anxiety disorder: a randomized controlled trial. Arch Gen Psychiatry, 68(7), 692-700. https://doi.org/10.1001/archgenpsychiatry.2011.67

Stanley, M. A., Beck, J. G., Novy, D. M., Averill, P. M., Swann, A. C., & Diefenbach, G. J. (2003). Cognitive-behavioral treatment of late-life generalized anxiety disorder. Journal of Consulting and Clinical Psychology, 71(2), 309-319. Retrieved from https://www.cochranelibrary.com/central/doi/10.1002/central/CN-00431425/full
Stanley, M. A., Beck, J. G., Novy, D. M., Averill, P. M., Swann, A. C., & Diefenbach, G. J. (2003). Cognitive-behavioral treatment of late-life generalized anxiety disorder. Journal of Consulting and Clinical Psychology, 71(2), 309-319. Retrieved from https://www.cochranelibrary.com/central/doi/10.1002/central/CN-00431425/full
Steel, C., Korrelboom, K., Baksh, M. F., Kingdon, D., Simon, J., Wykes, T., ... & van der Gaag, M. (2020). Positive memory training for the treatment of depression in schizophrenia: A randomised controlled trial. Behaviour Research and Therapy, 135, 103734.

Steinert C, Bumke PJ, Hollekamp RL, et al. Treating post-traumatic stress disorder by resource activation in Cambodia. World Psychiatry. 2016 Jun;15(2):183-5. doi: 10.1002/wps.20303. PMID: 27265714.

Stiles-Shields, C., Montague, E., Kwasny, M. J., & Mohr, D. C. (2019). Behavioral and cognitive intervention strategies delivered via coached apps for depression: Pilot trial. Psychological services, 16(2), 233-238. doi:10.1037/ser0000261

Stolz, T., Schulz, A., Krieger, T., Vincent, A., Urech, A., Moser, C., Westermann, S., & Berger, T. (2018). A mobile app for social anxiety disorder: A three-arm randomized controlled trial comparing mobile and PC-based guided self-help interventions. J Consult Clin Psychol, 86(6), 493-504. https://doi.org/10.1037/ccp0000301

Strauss C, Hayward M, Chadwick P. Group person-based cognitive therapy for chronic depression: A pilot randomized controlled trial. British Journal of Clinical Psychology. 2012;51(3):345-50.

Strong V, Waters R, Hibberd C, Murray G, Wall L, Walker J, et al. Management of depression for people with cancer (SMaRT oncology 1): A randomised trial. Lancet. 2008;372(9632):40-8.

Sugg HVR, Richards DA, Frost J. Morita Therapy for depression (Morita Trial): a pilot randomised controlled trial. BMJ open 2018; 8(8).

Sun, Q., Xu, H., Zhang, W., Zhou, Y., & Lv, Y. (2022). Behavioral Activation Therapy for Subthreshold Depression in Stroke Patients: An Exploratory Randomized Controlled Trial. Neuropsychiatric Disease and Treatment, 18, 2795-2805. doi:10.2147/NDT.S392403

Supiano, K. P., & Luptak, M. (2014). Complicated grief in older adults: A randomized controlled trial of complicated grief group therapy. The Gerontologist, 54(5), 840-856.

Swartz HA, Frank E, Zuckoff A, Cyranowski JM, Houck PR, Cheng Y, et al. Brief interpersonal psychotherapy for depressed mothers whose children are receiving psychiatric treatment. American Journal of Psychiatry. 2008;165(9):1155-62.

Swinson RP, Fergus KD, Cox BJ, et al. Efficacy of telephone-administered behavioral therapy for panic disorder with agoraphobia. Behaviour research and therapy 1995;33(4):465-9.

Swinson RP, Soulios C, Cox BJ, et al. Brief treatment of emergency room patients with panic attacks. The American journal of psychiatry 1992;149(7):944-6.

Sylvain, C., Ladouceur, R., & Boisvert, J. M. (1997). Cognitive and behavioral treatment of pathological gambling: a controlled study. Journal of consulting and Clinical Psychology, 65(5), 727-732.

Szumska I, Gola M, Rusanowska M, et al. Mindfulness-based cognitive therapy reduces clinical symptoms, but do not change frontal alpha asymmetry in people with major depression disorder. International Journal of Neuroscience. 2020.

Takagaki K, Okamoto Y, Jinnin R, Mori A, Nishiyama Y, Yamamura T, et al. Behavioral activation for late adolescents with subthreshold depression: A randomized controlled trial. European Child and Adolescent Psychiatry. 2016;25(11):1171-82.

Talbot NL, Chaudron LH, Ward EA, Duberstein PR, Conwell Y, O'Hara MW, et al. A randomized effectiveness trial of interpersonal psychotherapy for depressed women with sexual abuse histories. Psychiatric Services. 2011;62(4):374-80.

Tan S, Zou Y, Wykes T, Reeder C, Zhu X, Yang F, et al. Group cognitive remediation therapy for chronic schizophrenia : A randomized controlled trial. Neurosci Lett [Internet]. Elsevier Ireland Ltd; 2016;626:106, 11. Available from: http://dx.doi.org/10.1016/j.neulet.2015.08.036

Tao J, Zeng Q, Liang J, Zhou A, Yin X, Xu A. Effects of cognitive rehabilitation training on schizophrenia : 2 years of follow-up. Int J Clin Exp Med. 2015;8(9):16089, 94.

Tarrier N, Kelly J, Maqsood S, Snelson N, Maxwell J, Law H, et al. The cognitive behavioural prevention of suicide in psychosis : A clinical trial. Schizophr Res [Internet]. Elsevier B.V.; 2014;156(2, 3):204, 10. Available from: http://dx.doi.org/10.1016/j.schres.2014.04.029

Tarrier, N., Kelly, J., Maqsood, S., Snelson, N., Maxwell, J., Law, H., Dunn, G., & Gooding, P. (2014). The cognitive behavioural prevention of suicide in psychosis: a clinical trial. Schizophrenia research, 156(2-3), 204–210. https://doi.org/10.1016/j.schres.2014.04.029

Tas C, Danaci AE, Cubukcuoglu Z, BruÃàne M. Impact of family involvement on social cognition training in clinically stable outpatients with schizophrenia ,Äî A randomized pilot study. Psychiatry Res [Internet]. Elsevier Ireland Ltd; 2012;195:32, 8. Available from: http://dx.doi.org/10.1016/j.psychres.2011.07.031

Taylor CB, Conrad A, Wilhelm FH, Strachowski D, Khaylis A, Neri E, et al. Does improving mood in depressed patients alter factors that may affect cardiovascular disease risk? Journal of Psychiatric Research. 2009;43(16):1246-52.

Taylor FG, Marshall WL. Experimental analysis of a cognitive-behavioral therapy for depression. Cognitive Therapy and Research. 1977;1(1):59-72.

Teasdale JD, Fennell MJ, Hibbert GA, Amies PL. Cognitive therapy for major depressive disorder in primary care. The British Journal of Psychiatry. 1984;144:400-6.

Teichman Y, Bar-el Z, Shor H, Sirota P, Elizur A. A comparison of two modalities of cognitive therapy (individual and marital) in treating depression. Psychiatry. 1995;58(2):136-48.

Telch MJ, Lucas JA, Schmidt NB, et al. Group cognitive-behavioral treatment of panic disorder. Behaviour research and therapy 1993;31(3):279-87.

Tellez, M., Potter, C. M., Kinner, D. G., Jensen, D., Waldron, E., Heimberg, R. G., . . . Ismail, A. I. (2015). Computerized Tool to Manage Dental Anxiety: A Randomized Clinical Trial. J Dent Res, 94(9 Suppl), 174s-180s. doi:10.1177/0022034515598134

Teri L, Logsdon RG, Uomoto J, McCurry SM. Behavioral treatment of depression in dementia patients: A controlled clinical trial. The journals of gerontology Series B, Psychological sciences and social sciences. 1997;52(4):P159-66.

Thew, G. R., Kwok, A. P. L., Lissillour Chan, M. H., Powell, C., Wild, J., Leung, P. W. L., & Clark, D. M. (2022). Internet-delivered cognitive therapy for social anxiety disorder in Hong Kong: A randomized controlled trial. Internet Interv, 28, 100539. https://doi.org/10.1016/j.invent.2022.100539

Thielecke, J., Buntrock, C., Titzler, I., Braun, L., Freund, J., Berking, M., . . . Ebert, D. D. (2024). Telephone coaching for the prevention of depression in farmers: Results from a pragmatic randomized controlled trial. J Telemed Telecare, 30(6), 918-930. doi:https://doi.org/10.1177/1357633x221106027

Thomas, S. A., Drummond, A. E., Lincoln, N. B., Palmer, R. L., das Nair, R., Latimer, N. R., . . . Topcu, G. (2019). Behavioural activation therapy for post-stroke depression: the BEADS feasibility RCT. Health Technol Assess, 23(47), 1-176. doi:10.3310/hta23470

Tighe, J., Shand, F., Ridani, R., Mackinnon, A., De La Mata, N., & Christensen, H. (2017). Ibobbly mobile health intervention for suicide prevention in Australian Indigenous youth: a pilot randomised controlled trial. BMJ open, 7(1), e013518. https://doi.org/10.1136/bmjopen-2016-013518 174.Turner, R. M. (2000). Naturalistic evaluation of dialectical behavior therapy-oriented treatment for borderline personality disorder. Cognitive and Behavioral Practice, 7(4), 413–419. https://doi.org/10.1016/S1077-7229(00)80052-8

Titov N, Andrews G, Davies M, McIntyre K, Robinson E, Solley K. Internet treatment for depression: A randomized controlled trial comparing clinician vs. technician assistance. PLoS One. 2010;5(6):e10939.

Titov N, Dear BF, Ali S, Zou JB, Lorian CN, Johnston L, et al. Clinical and cost-effectiveness of therapist-guided internet-delivered cognitive behavior therapy for older adults with symptoms of depression: A randomized controlled trial. Behavior Therapy. 2015;46(2):193-205.

Titov, N., Andrews, G., & Schwencke, G. (2008b). Shyness 2: treating social phobia online: replication and extension. Aust N Z J Psychiatry, 42(7), 595-605. https://doi.org/10.1080/00048670802119820

Titov, N., Andrews, G., Choi, I., Schwencke, G., & Mahoney, A. (2008c). Shyness 3: randomized controlled trial of guided versus unguided Internet-based CBT for social phobia. Aust N Z J Psychiatry, 42(12), 1030-1040. https://doi.org/10.1080/00048670802512107

Titov, N., Andrews, G., Johnston, L., Robinson, E., & Spence, J. (2010). Transdiagnostic Internet treatment for anxiety disorders: A randomized controlled trial. Behaviour research and therapy, 48(9), 890-899.
Titov, N., Andrews, G., Johnston, L., Robinson, E., & Spence, J. (2010). Transdiagnostic Internet treatment for anxiety disorders: A randomized controlled trial. Behaviour research and therapy, 48(9), 890-899.
Titov, N., Andrews, G., Robinson, E., Schwencke, G., Johnston, L., Solley, K., & Choi, I. (2009). Clinician-assisted Internet-based treatment is effective for generalized anxiety disorder: Randomized controlled trial. Australian and New Zealand Journal of Psychiatry, 43(10), 905-912. doi:10.1080/00048670903179269
Titov, N., Andrews, G., Robinson, E., Schwencke, G., Johnston, L., Solley, K., & Choi, I. (2009). Clinician-assisted Internet-based treatment is effective for generalized anxiety disorder: Randomized controlled trial. Australian and New Zealand Journal of Psychiatry, 43(10), 905-912. doi:10.1080/00048670903179269
Titov, N., Andrews, G., Schwencke, G., Drobny, J., & Einstein, D. (2008a). Shyness 1: distance treatment of social phobia over the Internet. Aust N Z J Psychiatry, 42(7), 585-594. https://doi.org/10.1080/00048670802119762

Tobin, K., Davey-Rothwell, M. A., Nonyane, B. A. S., Knowlton, A., Wissow, L., & Latkin, C. A. (2017). RCT of an integrated CBT-HIV intervention on depressive symptoms and HIV risk.

Tomasino, K., Lattie, E., Ho, J., Palac, H., Kaiser, S., & Mohr, D. (2017). Harnessing Peer Support in an Online Intervention for Older Adults with Depression. American Journal of Geriatric Psychiatry, 25(10), 1109-1119.

Toneatto, T., Pillai, S., & Courtice, E. L. (2014). Mindfulness-enhanced cognitive behavior therapy for problem gambling: A controlled pilot study. International Journal of Mental Health and Addiction, 12(2), 197-205.

Tong, P., Bu, P., Yang, Y., Dong, L., Sun, T., & Shi, Y. (2019). Group cognitive behavioural therapy can reduce stigma and improve treatment compliance in major depressive disorder patients. Early intervention in psychiatry. doi:10.1111/eip.12841

Tønning, M. L., et al. (2021). "The effect of smartphone-based monitoring and treatment on the rate and duration of psychiatric readmission in patients with unipolar depressive disorder: The RADMIS randomized controlled trial." J Affect Disord 282: 354-363.

Tovote KA, Fleer J, Snippe E, Peeters A, Emmelkamp PMG, Sanderman R, et al. Individual mindfulness-based cognitive therapy and cognitive behavior therapy for treating depressive symptoms in patients with diabetes: Results of a randomized controlled trial. Diabetes care. 2014;37(9):2427-34.

Town, J., Abbass, A., Stride, C., & Bernier, D. (2017). A randomised controlled trial of Intensive Short-Term Dynamic Psychotherapy for treatment resistant depression: the Halifax Depression Study. Journal of Affective Disorders, 214, 15-25.

Treml, J., Nagl, M., Linde, K., Kündiger, C., Peterhänsel, C., & Kersting, A. (2021). Efficacy of an Internet-based cognitive-behavioural grief therapy for people bereaved by suicide: a randomized controlled trial. European journal of psychotraumatology, 12(1), 1926650.

Trevillion K, Ryan EG, Pickles A, et al. An exploratory parallel-group randomised controlled trial of antenatal Guided Self-Help (plus usual care) versus usual care alone for pregnant women with depression: DAWN trial. Journal of Affective Disorders. 2020;261:187-197.

Tulbure BT, Andersson G, Salagean N, Pearce M, Koenig HG. Religious versus Conventional Internet-based Cognitive Behavioral Therapy for Depression. J Relig Health 2018; 57(5): 1634-48.

Turkington D, Kingdon D, Turner T. Effectiveness of a brief cognitive-behavioural therapy intervention in the treatment of schizophrenia. Br J Psychiatry. 2002;180:523, 7.

Turner A, Hambridge J, Baker A, Bowman J, McElduff P. Randomised controlled trial of group cognitive behaviour therapy versus brief intervention for depression in cardiac patients. Australian and New Zealand Journal of Psychiatry. 2013;47(3):235-43.

Turner RW, Ward MF, Turner DJ. Behavioral treatment for depression: An evaluation of therapeutic components. Journal of clinical psychology. 1979;35(1):166-75.

Turner, R. M. (2000). Naturalistic evaluation of dialectical behavior therapyoriented treatment for borderline personality disorder. Cognitive and Behavioral Practice, 7(4), 413–419. https://doi.org/10.1016/S1077- 7229(00)80052-8
Turner, R. M. (2000). Naturalistic evaluation of dialectical behavior therapy-oriented treatment for borderline personality disorder. Cognitive and Behavioral Practice, 7(4), 413-419.
Turner, S. M., Beidel, D. C., & Jacob, R. G. (1994). Social phobia: a comparison of behavior therapy and atenolol. J Consult Clin Psychol, 62(2), 350-358. https://doi.org/10.1037//0022-006x.62.2.350
Twohig, M. P., Hayes, S. C., Plumb, J. C., Pruitt, L. D., Collins, A. B., Hazlett-Stevens, H., & Woidneck, M. R. (2010). A randomized clinical trial of acceptance and commitment therapy versus progressive relaxation training for obsessive-compulsive disorder. J Consult Clin Psychol, 78(5), 705-716.
Tylee DS, Gray R, Glatt SJ, et al. Evaluation of the reconsolidation of traumatic memories protocol for the treatment of PTSD: a randomized, wait-list-controlled trial. J Mil Veteran Fam Health. 2017;3(1):21-33. doi: 10.3138/jmvfh.4120.
Tyson GM, Range LM. Gestalt dialogues as a treatment for mild depression: Time works just as well. Journal of clinical psychology. 1987;43(2):227-31.
Ulmer CS, Edinger JD, Calhoun PS. A multi-component cognitive-behavioral intervention for sleep disturbance in veterans with PTSD: a pilot study. J Clin Sleep Med. 2011 Feb 15;7(1):57-68. PMID: 21344046.
Unlu Ince B, Cuijpers P, t Hof E, Ballegooijen W, Christensen H, Riper H. Internet-based, culturally sensitive, problem-solving therapy for Turkish migrants with depression: Randomized controlled trial. Journal of Medical Internet Research. 2013;15(10):e227.
Valencia M, Fresan A, JuaÃÅrez F, Escamilla R, Saracco R. The beneficial effects of combining pharmacological and psychosocial treatment on remission and functional outcome in outpatients with schizophrenia. J Psychiatr Res [Internet]. Elsevier Ltd; 2013;47(12):1886, 92. Available from: http://dx.doi.org/10.1016/j.jpsychires.2013.09.006
Valencia M, Juarez F, Ortega H. Integrated treatment to achieve functional recovery for first- episode psychosis. Schizophr Res Treat, 2012; Article ID 962371
Valencia M, Rascon M L, Juarez F, Escamilla R, Saracco R, Liberman R P. Application in Mexico of psychosocial rehabilitation with schizophrenia patients. Psychiatry. 2010; 73(3): 248, 263.
Valencia M, Rascon ML, Juarez F, Murow E. A psychosocial skills training approach in Mexican out-patients with schizophrenia. Psychol Med. 2007;37:1393, 402.
van Balkom, A. J., de Haan, E., van Oppen, P., Spinhoven, P., Hoogduin, K. A., & van Dyck, R. (1998). Cognitive and behavioral therapies alone versus in combination with fluvoxamine in the treatment of obsessive compulsive disorder. The Journal of nervous and mental disease, 186(8), 492-499.
van Ballegooijen W, Riper H, Klein B, et al. An Internet-based guided self-help intervention for panic symptoms: randomized controlled trial. Journal of medical internet research 2013;15(7):e154.
van Bastelaar KM, Pouwer F, Cuijpers P, Riper H, Snoek FJ. Web-based depression treatment for type 1 and type 2 diabetic patients: A randomized, controlled trial. Diabetes care. 2011;34(2):320-5.
Van Beek, W. Future thinking in suicidal patients, Ede: Print Service Ede B.V. , 2013, p226.
van den Berg DP, de Bont PA, van der Vleugel BM, et al. Prolonged exposure vs Eye Movement Desensitization and Reprocessing vs waiting list for posttraumatic stress disorder in patients with a psychotic disorder: a randomized clinical trial. JAMA Psychiatry. 2015 Mar;72(3):259-67. doi: 10.1001/jamapsychiatry.2014.2637. PMID: 25607833.
van Denderen M, de Keijser J, Stewart R, et al. Treating complicated grief and posttraumatic stress in homicidally bereaved individuals: a randomized controlled trial. Clin Psychol Psychother. 2018 Feb 26(25):497-508. doi: 10.1002/cpp.2183. PMID: 29479767.
van der Heiden, C., Muris, P., & van der Molen, H. T. (2012). Randomized controlled trial on the effectiveness of metacognitive therapy and intolerance-of-uncertainty therapy for generalized anxiety disorder. Behav Res Ther, 50(2), 100-109. doi:10.1016/j.brat.2011.12.005
van der Heiden, C., Muris, P., & van der Molen, H. T. (2012). Randomized controlled trial on the effectiveness of metacognitive therapy and intolerance-of-uncertainty therapy for generalized anxiety disorder. Behav Res Ther, 50(2), 100-109. doi:10.1016/j.brat.2011.12.005

van der Houwen, K., Schut, H., van den Bout, J., Stroebe, M., & Stroebe, W. (2010). The efficacy of a brief internet-based self-help intervention for the bereaved. Behaviour research and therapy, 48(5), 359-367.

Van der Kolk BA, Spinazzola J, Blaustein ME, et al. A randomized clinical trial of Eye Movement Desensitization and Reprocessing (EMDR), fluoxetine, and pill placebo in the treatment of posttraumatic stress disorder: treatment effects and long-term maintenance. J Clin Psychiatry. 2007 Jan;68(1):37-46. doi: 10.4088/JCP.v68n0105. PMID: 17284128.

van der Zanden, R., Kramer, J., Gerrits, R., & Cuijpers, P. (2012). Effectiveness of an online group course for depression in adolescents and young adults: a randomized trial. Journal of medical Internet research, 14(3), e86.

Van Gerwen, L. J., Spinhoven, P., & Van Dyck, R. (2006). Behavioral and cognitive group treatment for fear of flying: a randomized controlled trial. J Behav Ther Exp Psychiatry, 37(4), 358-371. doi:10.1016/j.jbtep.2006.05.002

Van Horne, B. S., Nong, Y. H., Cain, C. M., Sampson, M., Greeley, C. S., & Puryear, L. (2022). A promising new model of care for postpartum depression: A randomised controlled trial of a brief home visitation program conducted in Houston, Texas, USA. Health Soc Care Community, 30(5), e2203-e2213. doi:10.1111/hsc.13658

Van Lieshout RJ, Layton H, Savoy CD, Xie F, Brown JSL, Huh K, et al. In-person 1-day cognitive behavioral therapy-based workshops for postpartum depression: A randomized controlled trial. Psychological Medicine. 2023;53(14):6888-98.

Van Lieshout, R. J., Layton, H., Savoy, C. D., Brown, J. S. L., Ferro, M. A., Streiner, D. L., . . . Hanna, S. (2021). Effect of Online 1-Day Cognitive Behavioral Therapy-Based Workshops Plus Usual Care vs Usual Care Alone for Postpartum Depression: A Randomized Clinical Trial. JAMA Psychiatry, 78(11), 1200-1207. doi:10.1001/jamapsychiatry.2021.2488

Van Lieshout, R. J., Layton, H., Savoy, C. D., Haber, E., Feller, A., Biscaro, A., . . . Ferro, M. A. (2022). Public Health Nurse-delivered Group Cognitive Behavioural Therapy for Postpartum Depression: A Randomized Controlled Trial. Can J Psychiatry, 67(6), 432-440. doi:10.1177/07067437221074426

van Luenen, S., Garnefski, N., Spinhoven, P., & Kraaij, V. (2018). Guided internet-based intervention for people with HIV and depressive symptoms: a randomised controlled trial in the Netherlands. The lancet HIV, 5(9), e488-e497.

van Schaik A, van Marwijk H, Adèr H, van Dyck R, de Haan M, Penninx B, et al. Interpersonal psychotherapy for elderly patients in primary care. The American Journal of Geriatric Psychiatry. 2006;14(9):777-86.

Varela-Moreno, E., Anarte-Ortiz, M. T., Jodar-Sanchez, F., Garcia-Palacios, A., Monreal-Bartolomé, A., Gili, M., . . . Mayoral-Cleries, F. (2024). Economic Evaluation of a Web Application Implemented in Primary Care for the Treatment of Depression in Patients With Type 2 Diabetes Mellitus: Multicenter Randomized Controlled Trial. JMIR mHealth and uHealth, 12, e55483. doi:https://doi.org/10.2196/55483

Vázquez FL, López L, Torres Á J, et al. Analysis of the Components of a Cognitive-Behavioral Intervention for the prevention of Depression Administered via Conference Call to Nonprofessional Caregivers: A Randomized Controlled Trial. Int J Environ Res Public Health. 2020;17(6).

Vázquez González FL, Otero Otero P, Torres Iglesias A, Hermida Garcìa E, Blanco Seoane V, Dìaz Fernández O. A brief problem-solving indicated-prevention intervention for prevention of depression in nonprofessional caregivers. Psicothema. 2013;25(1):87-92.

Vázquez, F. L., Blanco, V., Hita, I., Torres, Á. J., Otero, P., Páramo, M., & Salmerón, M. (2023). Efficacy of a Cognitive Behavioral Intervention for the Prevention of Depression in Nonprofessional Caregivers Administered through a Smartphone App: A Randomized Controlled Trial. Journal of Clinical Medicine, 12(18).

Vázquez, F. L., Torres, Á., Otero, P., Blanco, V., Díaz, O., & Estévez, L. E. (2017). Analysis of the components of a cognitive-behavioral intervention administered via conference call for preventing depression among non-professional caregivers: A pilot study. Aging Ment Health, 21(9), 938-946.

Velligan DI, Tai S, Roberts DL, Maples-Aguilar N, Brown M, Mintz J, et al. A randomized controlled trial comparing cognitive behavior therapy , cognitive adaptation training , their combination and treatment as usual in chronic schizophrenia. Schizophr Bull. 2015;41(3):597, 603.

Vera M, Reyes-Rabanillo ML, Juarbe D, et al. Prolonged exposure for the treatment of Spanish-speaking Puerto Ricans with posttraumatic stress disorder: a feasibility study. BMC Res. Notes. 2011 Oct 17;4:415. doi: 10.1186/1756-0500-4-415. PMID: 22005187.

Vera, M., Obén, A., Juarbe, D., Hernández, N., & Pérez-Pedrogo, C. (2021). Randomized pilot trial of cognitive-behavioral therapy and acceptance-based behavioral therapy in the treatment of Spanish-speaking Latino primary care patients with generalized anxiety disorder. Journal of Behavioral and Cognitive Therapy, 31(2), 91-103. doi:10.1016/j.jbct.2020.11.007

Vera, M., Obén, A., Juarbe, D., Hernández, N., & Pérez-Pedrogo, C. (2021). Randomized pilot trial of cognitive-behavioral thera-py and acceptance-based behavioral therapy in the treatment of Spanish-speaking Latino primary care patients with gener-alized anxiety disorder. Journal of Behavioral and Cognitive Therapy, 31(2), 91-103. doi:10.1016/j.jbct.2020.11.007

Verduyn C, Barrowclough C, Roberts J, Tarrier N, Harrington R. Maternal depression and child behaviour problems: Randomised placebo-controlled trial of a cognitive-behavioural group intervention. British Journal of Psychiatry. 2003;183(OCT.):342-8.

Verheul, R., Van Den Bosch, L. M., Koeter, M. W., De Ridder, M. A., Stijnen, T., & Van Den Brink, W. (2003). Dialectical behaviour therapy for women with borderline personality disorder: 12-month, randomised clinical trial in The Netherlands. The British journal of psychiatry, 182(2), 135-140.

Vernmark K, Lenndin J, Bjärehed J, Carlsson M, Karlsson J, Oberg J, et al. Internet administered guided self-help versus individualized e-mail therapy: A randomized trial of two versions of CBT for major depression. Behaviour Research and Therapy. 2010;48(5):368-76.

Victor-Aigbodion, V., Eseadi, C., Ardi, Z., Sewagegn, A. A., Ololo, K., Abonor, L. B., . . . Effanga, O. A. (2023). Effectiveness of rational emotive behavior therapy in reducing depression among undergraduate medical students. Medicine (Baltimore), 102(4), e32724. doi:10.1097/md.0000000000032724

Vigod, S. N., Slyfield Cook, G., Macdonald, K., Hussain-Shamsy, N., Brown, H. K., de Oliveira, C., . . . Dennis, C.-L. (2021). Mother matters: Pilot randomized wait-list controlled trial of an online therapist-facilitated discussion board and support group for postpartum depression symptoms. Depress Anxiety, 38(8), 816-825. doi:10.1002/da.23163

Vita 2011a: Vita A, De Peri L, Barlati S, Cacciani P, Deste G, Poli R, Agrimi E, Cesana B M, Sacchetti E. Effectiveness of different modalities of cognitive remediation on symptomatological, neuropsychological, and functional outcome domains in schizophrenia: a prospective study in a real-world setting. Schizoph Res; 2011; 133, 223, 231.

Vita 2011b: Vita A, Peri L De, Barlati S, Cacciani P, Cisima M, Deste G, et al. Psychopathologic , neuropsychological and functional outcome measures during cognitive rehabilitation in schizophrenia : A prospective controlled study in a real-world setting. Eur Psychiatry. 2011;26:276, 83.

Vitriol VG, Ballesteros ST, Florenzano RU, Weil KP, Benadof DF. Evaluation of an outpatient intervention for women with severe depression and a history of childhood trauma. Psychiatric Services. 2009;60(7):936-42.

Vogel, P. A., Solem, S., Hagen, K., Moen, E. M., Launes, G., Håland, Å. T., . . . Himle, J. A. (2014). A pilot randomized controlled trial of videoconference-assisted treatment for obsessive-compulsive disorder. Behaviour Research and Therapy, 63, 162-168.

Wagner AW, Jakupcak M, Kowalski HM, et al. Behavioral activation as a treatment for posttraumatic stress disorder among returning veterans: a randomized trial. Psychiatr Serv. 2019 Oct 1;70(10):867-73. doi: 10.1176/appi.ps.201800572. PMID: 31337325.

Wagner AW, Zatzick DF, Ghesquiere A, et al. Behavioral activation as an early intervention for posttraumatic stress disorder and depression among physically injured trauma survivors. Cogn Behav Pract. 2007 Nov;14(4):341-9. doi: 10.1016/j.cbpra.2006.05.002.

Wagner, B., Grafiadeli, R., Schäfer, T., & Hofmann, L. (2022). Efficacy of an online-group intervention after suicide bereavement: A randomized controlled trial. Internet Interventions, 28, 100542.

Wagner, B., Knaevelsrud, C., & Maercker, A. (2006). Internet-based cognitive-behavioral therapy for complicated grief: a randomized controlled trial. Death studies, 30(5), 429-453.

Walker, T., Shaw, J., Turpin, C., Reid, C., & Abel, K. (2017). The WORSHIP II study: a pilot of psychodynamic interpersonal therapy with women offenders who self-harm. The Journal of Forensic Psychiatry & Psychology, 28(2), 158-171.

Wall, H., Magnusson, K., Hellner, C., Andersson, G., Jayaram-Lindström, N., & Rosendahl, I. (2023). The evaluation of a brief ICBT program with therapist support for individuals with gambling problems in the context of a gambling helpline: a randomized pilot trial. Pilot and Feasibility Studies, 9(1), 26.
Wang L-Q, Chien WT, Yip LK, Karatzias T. A randomized controlled trial of a mindfulness- based intervention program for people with schizophrenia : 6-month follow-up. Neuropsychiatr Dis Treat. 2016;12:3097, 110.
Wang, H., Zhao, Q., Mu, W., Rodriguez, M., Qian, M., & Berger, T. (2020). The Effect of Shame on Patients With Social Anxiety Disorder in Internet-Based Cognitive Behavioral Therapy: Randomized Controlled Trial. JMIR Ment Health, 7(7), e15797. https://doi.org/10.2196/15797
Ward-Ciesielski, E. F., Tidik, J. A., Edwards, A. J., & Linehan, M. M. (2017).Comparing brief interventions for suicidal individuals not engaged in treatment: A randomized clinical trial. Journal of affective disorders, 222, 153–161. https://doi.org/10.1016/j.jad.2017.07.011
Warmerdam L, Straten A, Twisk J, Riper H, Cuijpers P. Internet-based treatment for adults with depressive symptoms: Randomized controlled trial. Journal of Medical Internet Research. 2008;10(4):e44.
Watkins ER, Taylor RS, Byng R, Baeyens C, Read R, Pearson K, et al. Guided self-help concreteness training as an intervention for major depression in primary care: A Phase II randomized controlled trial. Psychological Medicine. 2012;42(7):1359-71.
Watt LM, Cappeliez P. Integrative and instrumental reminiscence therapies for depression in older adults: Intervention strategies and treatment effectiveness. Aging and Mental Health. 2000;4(2):166-77.
Wei, S., Liu, L., Bi, B., Li, H., Hou, J., Tan, S., Chen, X., Chen, W., Jia, X.,Dong, G., Qin, X., & Liu, Y. (2013). An intervention and follow-up study following a suicide attempt in the emergency departments of four general hospitals in Shenyang, China. Crisis, 34(2), 107–115. https://doi.org/10.1027/0227-5910/a000181
Weinberg, I., Gunderson, J. G., Hennen, J., & Cutter, C. J., Jr (2006). Manual assisted cognitive treatment for deliberate self-harm in borderline personality disorder patients. Journal of personality disorders, 20(5), 482–492. https://doi.org/10.1521/pedi.2006.20.5.482
Weissman MM, Prusoff BA, Dimascio A, Neu C, Goklaney M, Klerman GL. The efficacy of drugs and psychotherapy in the treatment of acute depressive episodes. American Journal of Psychiatry. 1979;136(4b):555-8.
Wells A, Colbear JS. Treating posttraumatic stress disorder with metacognitive therapy: a preliminary controlled trial. J Clin Psychol. 2012 Apr;68(4):373-81. doi: 10.1002/jclp.20871. PMID: 24469928.
Wells A, Walton D, Lovell K, et al. Metacognitive therapy versus prolonged exposure in adults with chronic post-traumatic stress disorder. Cognit Ther Res. 2015 February;39(1):70-80. doi: 10.1007/s10608-014-9636-6.
Westerhof, G. J., Lamers, S. M. A., Postel, M. G., & Bohlmeijer, E. T. (2019). Online Therapy for Depressive Symptoms: An Evaluation of Counselor-Led and Peer-Supported Life Review Therapy. The Gerontologist, 59(1), 135-146. doi:10.1093/geront/gnx140
Wetherell, J. L., Gatz, M., & Craske, M. G. (2003). Treatment of generalized anxiety disorder in older adults. J Consult Clin Psychol, 71(1), 31-40.
Wetherell, J. L., Gatz, M., & Craske, M. G. (2003). Treatment of generalized anxiety disorder in older adults. J Consult Clin Psychol, 71(1), 31-40.
Whittal, M. L., Woody, S. R., McLean, P. D., Rachman, S. J., & Robichaud, M. (2010). Treatment of obsessions: a randomized controlled trial. Behav Res Ther, 48(4), 295-303.
Wiborg IM, Dahl AA. Does brief dynamic psychotherapy reduce the relapse rate of panic disorder? Archives of general psychiatry 1996;53(8):689-94.
Wickberg B, Hwang CP. Counselling of postnatal depression: A controlled study on a population based Swedish sample. Journal of Affective Disorders. 1996;39(3):209-16.
Wiechers M, Strupf M, Bajbouj M, Böge K, Karnouk C, Goerigk S, et al. Empowerment group therapy for refugees with affective disorders: results of a multicenter randomized controlled trial. Eur Psychiatry. 2023;66(1):e64.
Wiersma JE, Schaik DJF, Hoogendorn AW, Dekker JJ, Van HL, Schoevers RA, et al. The effectiveness of the cognitive behavioral analysis system of psychotherapy for chronic depression: A randomized controlled trial. Psychotherapy and Psychosomatics. 2014;83(5):263-9.

Wiklund I, Mohlkert P, Edman G. Evaluation of a brief cognitive intervention in patients with signs of postnatal depression: A randomized controlled trial. Acta obstetricia et gynecologica Scandinavica. 2010;89(8):1100-4.

Wilhelm, S., Steketee, G., Fama, J. M., Buhlmann, U., Teachman, B. A., & Golan, E. (2009). Modular cognitive therapy for obsessive-compulsive disorder: A wait-list controlled trial. Journal of Cognitive Psychotherapy, 23(4), 294-305.

Wilks, C. R., Lungu, A., Ang, S. Y., Matsumiya, B., Yin, Q., & Linehan, M.M. (2018). A randomized controlled trial of an Internet delivered dialectical behavior therapy skills training for suicidal and heavy episodic drinkers. Journal of affective disorders, 232, 219–228. https://doi.org/10.1016/j.jad.2018.02.053

Williams AD, Blackwell SE, Mackenzie A, Holmes EA, Andrews G. Combining imagination and reason in the treatment of depression: A randomized controlled trial of internet-based cognitive-bias modification and internet-CBT for depression. Journal of Consulting and Clinical Psychology. 2013;81(5):793-9.

Williams C, McClay CA, Matthews L, et al. Community-based group guided self-help intervention for low mood and stress: Randomised controlled trial. British Journal of Psychiatry 2018; 212(2): 88-95.

Williams C, Wilson P, Morrison J, McMahon A, Andrew W, Allan L, et al. Guided self-help cognitive behavioural therapy for depression in primary care: A randomised controlled trial. PLoS One. 2013;8(1):e52735.

Williams JW, Barrett J, Oxman T, Frank E, Katon W, Sullivan M, et al. Treatment of dysthymia and minor depression in primary care: A randomized controlled trial in older adults. JAMA. 2000;284(12):1519-26.

Williams, S. L., Turner, S. M., & Peer, D. F. (1985). Guided mastery and performance desensitization treatments for severe acrophobia. Journal of Consulting and Clinical Psychology, 53, 237-247. doi:10.1037/0022-006X.53.2.237

Wilson PH, Goldin JC, Charbonneau-Powis M. Comparative efficacy of behavioral and cognitive treatments of depression. Cognitive Therapy and Research. 1983;7(2):111-24.

Wims E, Titov N, Andrews G, et al. Clinician-assisted Internet-based treatment is effective for panic: A randomized controlled trial. The Australian and New Zealand journal of psychiatry 2010;44(7):599-607.

Winnebeck, E., Fissler, M., Gärtner, M., Chadwick, P., & Barnhofer, T. (2017). Brief training in mindfulness meditation reduces symptoms in patients with a chronic or recurrent lifetime history of depression: A randomized controlled study. Behaviour Research and Therapy, 99, 124-130. doi:10.1016/j.brat.2017.10.005

Wittouck, C., Van Autreve, S., Portzky, G., & van Heeringen, K. (2014). A CBT-based psychoeducational intervention for suicide survivors. Crisis.

Wolitzky, K. B., & Telch, M. J. (2009). Augmenting in vivo exposure with fear antagonistic actions: a preliminary test. Behav Ther, 40(1), 57-71. doi:10.1016/j.beth.2007.12.006

Wollburg E, Roth WT, Kim S. Effects of breathing training on voluntary hypo- and hyperventilation in patients with panic disorder and episodic anxiety. Applied Psychophysiology & Biofeedback 2011;36:81–91.

Wollersheim JP, Wilson GL. Group treatment of unipolar depression: A comparison of coping, supportive, bibliotherapy, and delayed treatment groups. Professional Psychology: Research and Practice. 1991;22(6):496.

Wong DF. Cognitive and health-related outcomes of group cognitive behavioural treatment for people with depressive symptoms in Hong Kong: Randomized wait-list control study. Australian and New Zealand Journal of Psychiatry. 2008;42(8):702-11.

Wong DF. Cognitive behavioral treatment groups for people with chronic depression in Hong Kong: A randomized wait-list control design. Depression and anxiety. 2008;25(2):142-8.

Wong SYS, Sun YY, Chan ATY, et al. Treating Subthreshold Depression in Primary Care: a Randomized Controlled Trial of Behavioral Activation With Mindfulness. Annals of family medicine 2018; 16(2): 111-9.

Wong, D. F. K., Chung, C. L. P., Wu, J., Tang, J., & Lau, P. (2015). A preliminary study of an integrated and culturally attuned cognitive behavioral group treatment for Chinese problem gamblers in Hong Kong. Journal of Gambling Studies, 31(3), 1015-1027.

Wong, S. Y., Yip, B. H., Mak, W. W., Mercer, S., Cheung, E. Y., Ling, C. Y., . . . Ma, H. S. (2016). Mindfulness-based cognitive therapy v. group psychoeducation for people with generalised anxiety disorder: randomised controlled trial. Br J Psychiatry, 209(1), 68-75. doi:10.1192/bjp.bp.115.166124

Wong, S. Y., Yip, B. H., Mak, W. W., Mercer, S., Cheung, E. Y., Ling, C. Y., . . . Ma, H. S. (2016). Mindfulness-based cognitive therapy v. group psychoeducation for people with generalised anxiety disorder: randomised controlled trial. Br J Psychiatry, 209(1), 68-75. doi:10.1192/bjp.bp.115.166124

Woodward, R., & Jones, R. B. (1980). Cognitive restructuring treatment: a controlled trial with anxious patients. Behaviour Research and Therapy, 18(5), 401-407. Retrieved from https://www.cochranelibrary.com/central/doi/10.1002/central/CN-00178686/full

Woodward, R., & Jones, R. B. (1980). Cognitive restructuring treatment: a controlled trial with anxious patients. Behaviour Re-search and Therapy, 18(5), 401-407. Retrieved from https://www.cochranelibrary.com/central/doi/10.1002/central/CN-00178686/full

Wootton, B. M., Dear, B. F., Johnston, L., Terides, M. D., & Titov, N. (2013). Remote treatment of obsessive-compulsive disorder: A randomized controlled trial. Journal of Obsessive-Compulsive and Related Disorders, 2(4), 375-384.

Wright JH, Wright AS, Albano AM, Basco MR, Goldsmith LJ, Raffield T, et al. Computer-assisted cognitive therapy for depression: Maintaining efficacy while reducing therapist time. American Journal of Psychiatry. 2005;162(6):1158-64.

Wright, J. H., Owen, J., Eells, T. D., Antle, B., Bishop, L. B., Girdler, R., . . . Ali, S. (2022). Effect of Computer-Assisted Cognitive Behavior Therapy vs Usual Care on Depression Among Adults in Primary Care: A Randomized Clinical Trial. JAMA Netw Open, 5(2), e2146716. doi:10.1001/jamanetworkopen.2021.46716

Wu, R., Zhong, S. Y., Wang, G. H., Wu, M. Y., Xu, J. F., Zhu, H., ... & Jiang, C. L. (2023). The effect of brief mindfulness meditation on suicidal ideation, stress and sleep quality. Archives of suicide research, 27(2), 215-230.

Wulfert, E., Wemm, S. E., & Broussard, J. D. (2025). Betting on change: An analysis of cognitive motivational behavior therapy versus referral to gamblers anonymous for gambling disorder. Psychology of Addictive Behaviors.

Wuthrich VM, Rapee RM, Kangas M, Perini S. Randomized controlled trial of group cognitive behavioral therapy compared to a discussion group for co-morbid anxiety and depression in older adults. Psychological Medicine. 2016;46(4):785-95.

Wuthrich VM, Rapee RM. Randomised controlled trial of group cognitive behavioural therapy for comorbid anxiety and depression in older adults. Behaviour Research and Therapy. 2013;51(12):779-86.

Wykes T, Reeder C, Landau S, Everitt B, Knapp M, Patel A, Romeo R. Cognitive remediation therapy in schizophrenia. Br J Psychiatry, 2007; 190, 421, 427.

Xiang, X., Kayser, J., Turner, S., Ash, S., & Himle, J. A. (2024). Layperson-Supported, Web-Delivered Cognitive Behavioral Therapy for Depression in Older Adults: Randomized Controlled Trial. Journal of Medical Internet Research, 26(1). doi:https://doi.org/10.2196/53001

Xie, J., He, G., Ding, S., Pan, C., Zhang, X., Zhou, J., & Iennaco, J. D. (2019). A randomized study on the effect of modified behavioral activation treatment for depressive symptoms in rural left-behind elderly. Psychotherapy research : journal of the Society for Psychotherapy Research, 29(3), 372-382. doi:10.1080/10503307.2017.1364444

Yan S, Zhong S, Lyu S, Lai S, Zhang Y, Luo Y, et al. A randomized trial of virtual reality eye movement desensitization and reprocessing therapy for major depressive disorder with childhood trauma: A 3-month follow-up study. Psychological Trauma: Theory, Research, Practice, and Policy. 2025.

Yang X, Zhao J, Chen Y, Zu S, Zhao J. Comprehensive self-control training benefits depressed college students: a six-month randomized controlled intervention trial. Journal of affective disorders 2018; 226: 251-60.

Ye, H. (2017). Impact of mindfulness-based stress reduction (MBSR) on students' social anxiety: A randomized controlled trial [Article]. NeuroQuantology, 15(4), 101-106. https://doi.org/10.14704/nq.2017.15.4.1134

Yehuda R, Pratchett LC, Elmes MW, et al. Glucocorticoid-related predictors and correlates of post-traumatic stress disorder treatment response in combat veterans. Interface Focus. 2014 Oct 6;4(5):20140048. doi: 10.1098/rsfs.2014.0048. PMID: 25285201.
Yeung, A., Wang, F., Feng, F., Zhang, J., Cooper, A., Hong, L., . . . Fava, M. (2017). Outcomes of an online computerized cognitive behavioral treatment program for treating chinese patients with depression: A pilot study. Asian journal of psychiatry. doi:10.1016/j.ajp.2017
Ying, Y., Ji, Y., Kong, F., Wang, M., Chen, Q., Wang, L., . . . Ruan, L. (2022). Efficacy of an internet-based cognitive behavioral therapy for subthreshold depression among Chinese adults: a randomized controlled trial. Psychological medicine, 1-11. doi:10.1017/S0033291722000599
Yokomitsu, K., Inoue, K., Kamimura, E., Matsushita, S., & So, R. (2025). Effectiveness of Internet-based personalized normative feedback among individuals experiencing problem gambling: Randomized controlled trial. Journal of Gambling Studies, 1-27.
Yoshinaga, N., Matsuki, S., Niitsu, T., Sato, Y., Tanaka, M., Ibuki, H., Takanashi, R., Ohshiro, K., Ohshima, F., Asano, K., Kobori, O., Yoshimura, K., Hirano, Y., Sawaguchi, K., Koshizaka, M., Hanaoka, H., Nakagawa, A., Nakazato, M., Iyo, M., & Shimizu, E. (2016). Cognitive Behavioral Therapy for Patients with Social Anxiety Disorder Who Remain Symptomatic following Antidepressant Treatment: A Randomized, Assessor-Blinded, Controlled Trial. Psychother Psychosom, 85(4), 208-217. https://doi.org/10.1159/000444221
Yuan J, Yin Y, Tang X, et al. Culturally adapted and lay-delivered cognitive behaviour therapy for older adults with depressive symptoms in rural China: a pilot trial. Behavioural and cognitive psychotherapy. 2020:1-5.
Yurtsever A, Konuk E, Akyüz T, et al. An Eye Movement Desensitization and Reprocessing (EMDR) group intervention for Syrian refugees with post-traumatic stress symptoms: results of a randomized controlled trial. Front Psychol. 2018 Jun 12;9:493. doi: 10.3389/fpsyg.2018.00493. PMID: 29946275.
Zainal, N. H., Chan, W. W., Saxena, A. P., Taylor, C. B., & Newman, M. G. (2021). Pilot randomized trial of self-guided virtual reality exposure therapy for social anxiety disorder. Behav Res Ther, 147, 103984. https://doi.org/10.1016/j.brat.2021.103984
Zang Y, Hunt NC, Cox T. Adapting narrative exposure therapy for Chinese earthquake survivors: a pilot randomised controlled feasibility study. BMC Psychiatry. 2014 Oct 3;14:262. doi: 10.1186/s12888-014-0262-3. PMID: 25927297.
Zargar, F., Farid, A. A. A., Atef-Vahid, M., Afshar, H., Maroofi, M., & Omranifard, V. (2012). Effect of acceptance-based behavior therapy on severity of symptoms, worry and quality of life in women with generalized anxiety disorder. Iranian Journal of Psychiatry and Behavioral Sciences, 6(2), 23-32. Retrieved from http://www.embase.com/search/results?subaction=viewrecord&from=export&id=L368838694
Zargar, F., Farid, A. A. A., Atef-Vahid, M., Afshar, H., Maroofi, M., & Omranifard, V. (2012). Effect of acceptance-based behavior therapy on severity of symptoms, worry and quality of life in women with generalized anxiety disorder. Iranian Journal of Psychiatry and Behavioral Sciences, 6(2), 23-32. Retrieved from http://www.embase.com/search/results?subaction=viewrecord&from=export&id=L368838694
Zejuan FU, Hailing XIE. Effect evaluation of cognitive behavior intervention therapy in improving psychological status and quality of life in patients with obsessive-compulsive disorder[J]. Journal of Clinical Medicine in Practice, 2016, (8): 17-20. doi: 10.7619/jcmp.201608006
Zemestani M, Davoodi I, Honarmand MM, Zargar Y, Ottaviani C. Comparative effects of group metacognitive therapy versus behavioural activation in moderately depressed students. Journal of Mental Health. 2016;25(6):479-85.
Zemestani M, Mohammed AF, Ismail AA, et al. A Pilot Randomized Clinical Trial of a Novel, Culturally Adapted, Trauma-Focused Cognitive-Behavioral Intervention for War-Related PTSD in Iraqi Women. Behav Ther. 2022 Jul;53(4):656-72. doi: 10.1016/j.beth.2022.01.009. PMID: 35697429
Zemestani, M., & Fazeli Nikoo, Z. (2020). Effectiveness of mindfulness-based cognitive therapy for comorbid depression and anxiety in pregnancy: a randomized controlled trial. Archives of Women's Mental Health. doi:10.1007/s00737-019-00962-8

Zhang, T., Lu, L., Didonna, F., Wang, Z., Zhang, H., & Fan, Q. (2021). Mindfulness-Based Cognitive Therapy for Unmedicated Obsessive-Compulsive Disorder: A Randomized Controlled Trial With 6-Month Follow-Up. Frontiers in Psychiatry, 12.

Zhao, Y., Lin, Q., & Wang, J. (2021). An evaluation of a prenatal individualised mixed management intervention addressing breastfeeding outcomes and postpartum depression: A randomised controlled trial. J Clin Nurs, 30(9-10), 1347-1359. doi:10.1111/jocn.15684

Zhao, Y., Munro-Kramer, M. L., Shi, S., Wang, J., & Zhao, Q. (2019). Effects of antenatal depression screening and intervention among Chinese high-risk pregnant women with medically defined complications: A randomized controlled trial. Early intervention in psychiatry, 13(5), 1090-1098. doi:10.1111/eip.12731

Zinbarg, R. E., Lee, J. E., & Yoon, K. L. (2007). Dyadic predictors of outcome in a cognitive-behavioral program for patients with generalized anxiety disorder in committed relationships: a "spoonful of sugar" and a dose of non-hostile criticism may help. Behav Res Ther, 45(4), 699-713. doi:10.1016/j.brat.2006.06.005

Zinbarg, R. E., Lee, J. E., & Yoon, K. L. (2007). Dyadic predictors of outcome in a cognitive-behavioral program for patients with generalized anxiety disorder in committed relationships: a "spoonful of sugar" and a dose of non-hostile criticism may help. Behav Res Ther, 45(4), 699-713. doi:10.1016/j.brat.2006.06.005

Zitrin, C. M., et al. (1978). "Behavior therapy, supportive psychotherapy, imipramine, and phobias." Arch Gen Psychiatry 35(3): 307-316.

Zlotnick C, Shea MT, Rosen KH, et al. An affect-management group for women with posttraumatic stress disorder and histories of childhood sexual abuse. J Trauma Stress. 1997 Jul;10(3):425-36. doi: 10.1023/A:1024841321156. PMID: 9246650.

Zoellner LA, Telch M, Foa EB, et al. Enhancing extinction learning in posttraumatic stress disorder with brief daily imaginal exposure and methylene blue: a randomized controlled trial. J Clin Psychiatry. 2017;78(7):e782-e9. doi: 10.4088/JCP.16m10936. PMID: 28686823.

Zu S, Xiang Y-T, Liu J, Zhang L, Wang G, Ma X, et al. A comparison of cognitive-behavioral therapy, antidepressants, their combination and standard treatment for Chinese patients with moderate–severe major depressive disorders. Journal of Affective Disorders. 2014;152-154:262-7.

### S3. Characteristics of the included studies by indication.

| Indication | *k* | $n_{comp}$ | Sample Size | | | Waitlist Ctrl. | | Low RoB | | Publication Years | | |
|---|---|---|---|---|---|---|---|---|---|---|---|---|
| | | | *M* | Median | Range | *N* | % | *N* | % | *M* | Median | Range |
| Depression | 504 | 933 | 110.7 | 68 | [12; 1140] | 181 | 35.9% | 196 | 38.9% | 2013.2 | 2015 | [1977; 2025] |
| Gen. Anxiety | 50 | 352 | 69.5 | 45 | [12; 756] | 36 | 72.0% | 9 | 18.0% | 2008.0 | 2010 | [1980; 2022] |
| Panic | 61 | 88 | 61.4 | 39 | [14; 419] | 39 | 63.9% | 4 | 6.6% | 2002.7 | 2003 | [1978; 2020] |
| Social Anxiety | 60 | 279 | 56.2 | 51 | [14; 288] | 50 | 83.3% | 4 | 6.7% | 2010.3 | 2011 | [1989; 2023] |
| Phobia | 36 | 61 | 38.8 | 29 | [12; 151] | 29 | 80.6% | 3 | 8.3% | 2002.2 | 2003 | [1973; 2022] |
| PTSD | 102 | 131 | 48.6 | 40 | [8; 159] | 67 | 65.7% | 29 | 28.4% | 2011.2 | 2012 | [1989; 2022] |
| OCD | 67 | 144 | 53.0 | 42 | [12; 179] | 36 | 53.7% | 3 | 4.5% | 2012.6 | 2014 | [1992; 2023] |
| BPD | 27 | 28 | 51.7 | 44 | [19; 134] | 0 | 0.0% | 3 | 11.1% | 2011.9 | 2012 | [2000; 2020] |
| Grief | 45 | 49 | 89.8 | 61 | [12; 455] | 19 | 42.2% | 9 | 20.0% | 2012.6 | 2014 | [1981; 2023] |
| Gambling | 41 | 267 | 89.4 | 65 | [14; 278] | 25 | 61.0% | 0 | 0.0% | 2013.6 | 2015 | [1997; 2025] |
| Psychosis | 175 | 465 | 79.8 | 58 | [11; 744] | 2 | 1.1% | 20 | 11.4% | 2014.6 | 2016 | [1995; 2025] |
| Suicidality | 78 | 84 | 82.5 | 63 | [8; 552] | 16 | 20.5% | 2 | 2.6% | 2014.2 | 2017 | [1981; 2022] |
| **Overall** | 1,246 | 2,881 | 85.4 | 57 | [8; 1140] | 500 | 40.1% | 282 | 22.6% | 2012.0 | 2014 | [1973; 2025] |

*Note*. BPD = borderline personality disorder; PTSD = posttraumatic stress disorder; OCD = obsessive-compulsive disorder; RoB = Risk of Bias; k = number of studies; $n_{comp}$ = number of comparisons/effect sizes.

### S4. Comparative fit of the reference effect size distributions.

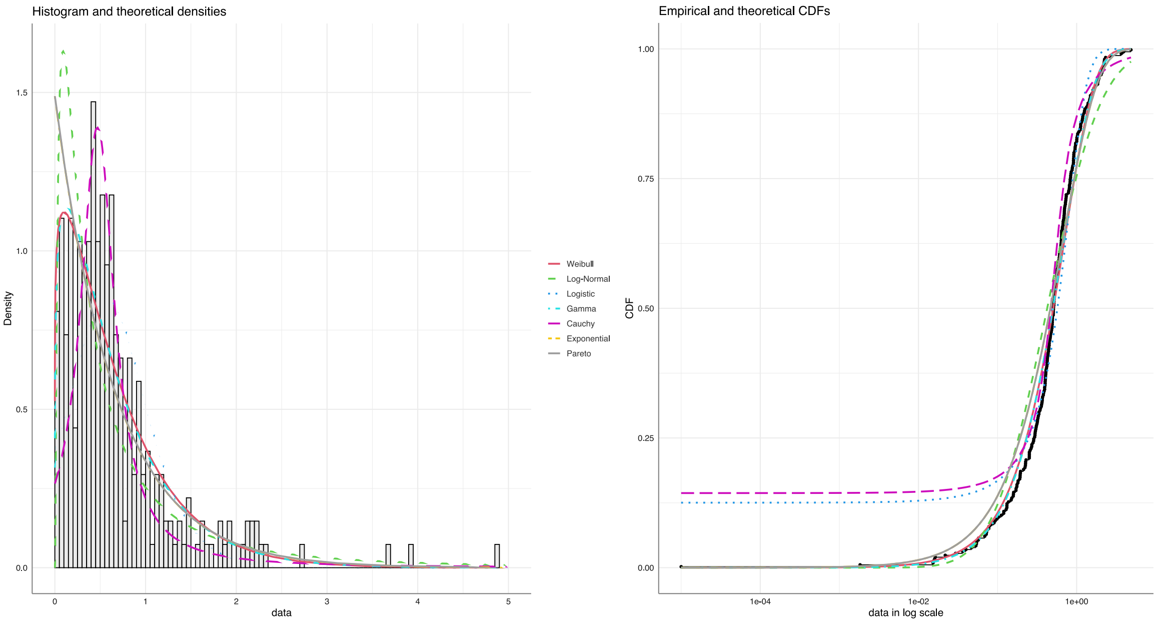

| | Weibull | Log-Normal | Logistic | Gamma | Cauchy | Exponential | Pareto |
|---|---|---|---|---|---|---|---|
| **Goodness-of-fit statistics** | | | | | | | |
| Kolmogorov-Smirnov | 0.0673023 | 0.1358204 | 0.1252800 | 0.06761565 | 0.1437766 | 0.1047671 | 0.1047998 |
| Cramer-von Mises | 0.3727933 | 1.4842402 | 0.8037424 | 0.34579887 | 0.9578037 | 0.8248794 | 0.8252272 |
| Anderson-Darling | 1.9509118 | 8.1032724 | 7.5596091 | 1.79145400 | 7.1798092 | 4.2678049 | 4.2691815 |
| **Goodness-of-fit criteria** | | | | | | | |
| Akaike's Information Crit. | 325.2170 | 426.1861 | 454.1184 | 324.7594 | 428.7423 | 329.5019 | 331.5019 |
| Bayesian Information Crit. | 332.4286 | 433.3977 | 461.3300 | 331.9710 | 435.9539 | 333.1077 | 338.7135 |

### S5. Empirical distributions of the estimated achieved power across trials.

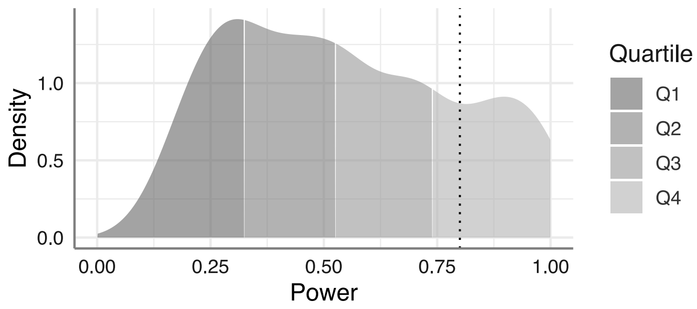

| | *Power* $(1-\beta)$ |
|---|---|
| 1st Quartile | 0.3240959 |
| 2nd Quartile | 0.5261834 |
| 3rd Quartile | 0.7381103 |

*Note*. The dotted line indicated $1-\beta=0.8$ (i.e., the conventional target threshold for sufficient power).

### S6. Recalculated effects of psychological treatment when excluding specific flags.

| | $k$ | $\Delta_k$ | SMD | 95%-CI | $\Delta_{SMD}$ | $\tau^2$ | $\Delta_\tau$ | 95%-CI | NNT |
|---|---|---|---|---|---|---|---|---|---|
| ***Depressive Disorders*** | | | | | | | | | |
| All Effect Sizes Included | 1,246 | *ref.* | 0.73 | [0.67; 0.79] | *ref.* | 0.3917 | *ref.* | [-0.50; 1.96] | 4.04 |
| - Flag 1 Removed (Effect) | 1,140 | 8.5% | 0.58 | [0.54; 0.61] | 20.5% | 0.1159 | 45.6% | [-0.09; 1.24] | 5.35 |
| - Flag 2 Removed (Power) | 540 | 56.7% | 0.54 | [0.49; 0.59] | 26.0% | 0.1546 | 37.2% | [-0.24; 1.31] | 5.79 |
| - Flag 3 Removed (RoB) | 413 | 66.9% | 0.56 | [0.49; 0.64] | 23.3% | 0.2518 | 19.8% | [-0.43; 1.55] | 5.49 |
| ***Posttraumatic Stress Disorder*** | | | | | | | | | |
| All Effect Sizes Included | 131 | *ref.* | 1.18 | [1.01; 1.35] | *ref.* | 0.6177 | *ref.* | [-0.39; 2.74] | 2.79 |
| - Flag 1 Removed (Effect) | 123 | 6.1% | 1.02 | [0.89; 1.15] | 13.6% | 0.2931 | 31.1% | [-0.06; 2.10] | 3.37 |
| - Flag 2 Removed (Power) | 82 | 37.4% | 0.97 | [0.80; 1.14] | 17.8% | 0.3964 | 19.9% | [-0.29; 2.24] | 3.60 |
| - Flag 3 Removed (RoB) | 32 | 75.6% | 1.25 | [0.93; 1.57] | -5.9% | 0.5742 | 3.6% | [-0.33; 2.83] | 2.58 |
| ***Suicidal Ideation*** | | | | | | | | | |
| All Effect Sizes Included | 84 | *ref.* | 0.34 | [0.22; 0.45] | *ref.* | 0.1842 | *ref.* | [-0.52; 1.2] | 9.82 |
| - Flag 1 Removed (Effect) | 81 | 3.6% | 0.27 | [0.18; 0.36] | 20.6% | 0.0783 | 34.8% | [-0.30; 0.83] | 12.74 |
| - Flag 2 Removed (Power) | 54 | 35.7% | 0.22 | [0.12; 0.32] | 35.3% | 0.0683 | 39.1% | [-0.31; 0.75] | 15.73 |
| - Flag 3 Removed (RoB) † | - | - | - | - | - | - | - | - | - |
| ***Psychotic Disorders*** | | | | | | | | | |
| All Effect Sizes Included | 174 | *ref.* | 0.32 | [0.23; 0.40] | *ref.* | 0.0741 | *ref.* | [-0.23; 0.86] | 14.85 |
| - Flag 1 Removed (Effect) | 172 | 1.1% | 0.30 | [0.22; 0.38] | 6.3% | 0.0644 | 6.8% | [-0.21; 0.81] | 15.76 |
| - Flag 2 Removed (Power) | 88 | 49.4% | 0.25 | [0.14; 0.35] | 21.9% | 0.0748 | -0.5% | [-0.30; 0.8] | 19.58 |
| - Flag 3 Removed (RoB) | 36 | 79.3% | 0.29 | [0.11; 0.47] | 9.4% | 0.0529 | 15.5% | [-0.21; 0.78] | 16.65 |
| ***Panic Disorders*** | | | | | | | | | |
| All Effect Sizes Included | 88 | *ref.* | 0.83 | [0.68; 0.98] | *ref.* | 0.2841 | *ref.* | [-0.24; 1.90] | 3.83 |
| - Flag 1 Removed (Effect) | 80 | 9.1% | 0.75 | [0.62; 0.88] | 9.6% | 0.1805 | 20.3% | [-0.10; 1.61] | 4.32 |
| - Flag 2 Removed (Power) | 29 | 67.0% | 0.67 | [0.47; 0.86] | 19.3% | 0.1635 | 24.1% | [-0.18; 1.52] | 5.02 |
| - Flag 3 Removed (RoB) | 6 | 93.2% | 0.44 | [-0.07; 0.94] | 47.0% | 0.1487 | 27.7% | [-0.66; 1.53] | 8.37 |
| ***Social Anxiety Disorder*** | | | | | | | | | |
| All Effect Sizes Included | 279 | *ref.* | 0.95 | [0.80; 1.10] | *ref.* | 0.3460 | *ref.* | [-0.22; 2.12] | 3.56 |
| - Flag 1 Removed (Effect) | 252 | 9.7% | 0.79 | [0.70; 0.88] | 16.8% | 0.0801 | 51.9% | [0.22; 1.35] | 4.53 |
| - Flag 2 Removed (Power) | 122 | 56.3% | 0.73 | [0.63; 0.83] | 23.2% | 0.0508 | 61.7% | [0.27; 1.19] | 5.00 |
| - Flag 3 Removed (RoB) | 11 | 96.1% | 1.63 | [0.59; 2.66] | -71.6% | 0.4839 | -18.3% | [-0.08; 3.33] | 1.83 |
| ***Generalized Anxiety Disorder*** | | | | | | | | | |
| All Effect Sizes Included | 118 | *ref.* | 0.86 | [0.69; 1.03] | *ref.* | 0.2672 | *ref.* | [-0.18; 1.90] | 3.66 |
| - Flag 1 Removed (Effect) | 108 | 8.5% | 0.75 | [0.62; 0.88] | 12.8% | 0.1455 | 26.2% | [-0.02; 1.51] | 4.35 |
| - Flag 2 Removed (Power) | 50 | 57.6% | 0.68 | [0.51; 0.86] | 20.9% | 0.1313 | 29.9% | [-0.06; 1.43] | 4.85 |
| - Flag 3 Removed (RoB) | 17 | 85.6% | 0.75 | [0.41; 1.09] | 12.8% | 0.1518 | 24.6% | [-0.13; 1.63] | 4.34 |

| | $k$ | $\Delta_k$ | SMD | 95%-CI | $\Delta_{SMD}$ | $\tau^2$ | $\Delta_\tau$ | 95%-CI | NNT |
|---|---|---|---|---|---|---|---|---|---|
| ***Specific Phobias*** | | | | | | | | | |
| All Effect Sizes Included | 61 | *ref.* | 1.25 | [1.01; 1.49] | *ref.* | 0.4999 | *ref.* | [-0.18; 2.68] | 2.68 |
| - Flag 1 Removed (Effect) | 48 | 21.3% | 0.94 | [0.75; 1.14] | 24.8% | 0.1623 | 43.0% | [0.11; 1.78] | 3.91 |
| - Flag 2 Removed (Power) | 12 | 80.3% | 1.12 | [0.63; 1.62] | 10.4% | 0.3601 | 15.1% | [-0.28; 2.52] | 3.09 |
| - Flag 3 Removed (RoB) | 3 | 95.1% | 1.31 | [-0.56; 3.19] | -4.8% | 0.4635 | 3.7% | [-2.16; 4.79] | 2.51 |
| ***Obsessive-Compulsive Disorder*** | | | | | | | | | |
| All Effect Sizes Included | 55 | *ref.* | 1.18 | [0.96; 1.39] | *ref.* | 0.2725 | *ref.* | [0.11; 2.24] | 3.71 |
| - Flag 1 Removed (Effect) | 50 | 9.1% | 1.06 | [0.89; 1.22] | 10.2% | 0.1099 | 36.5% | [0.37; 1.74] | 4.38 |
| - Flag 2 Removed (Power) | 13 | 76.4% | 0.96 | [0.64; 1.27] | 18.6% | 0.1707 | 20.9% | [0.01; 1.91] | 5.12 |
| - Flag 3 Removed (RoB) † | - | - | - | - | - | - | - | - | - |
| ***Prolonged Grief Disorder*** | | | | | | | | | |
| All Effect Sizes Included | 32 | *ref.* | 0.49 | [0.24; 0.73] | *ref.* | 0.3341 | *ref.* | [-0.71; 1.69] | 5.33 |
| - Flag 1 Removed (Effect) | 30 | 6.2% | 0.37 | [0.18; 0.57] | 24.5% | 0.1766 | 27.3% | [-0.51; 1.26] | 6.86 |
| - Flag 2 Removed (Power) | 19 | 40.6% | 0.33 | [0.12; 0.54] | 32.7% | 0.1235 | 39.2% | [-0.44; 1.10] | 7.74 |
| - Flag 3 Removed (RoB) | 13 | 59.4% | 0.43 | [0; 0.85] | 12.2% | 0.3977 | -9.1% | [-1.01; 1.86] | 6.07 |
| ***Borderline Personality Disorder*** | | | | | | | | | |
| All Effect Sizes Included | 28 | *ref.* | 0.46 | [0.26; 0.67] | *ref.* | 0.1707 | *ref.* | [-0.41; 1.34] | 7.48 |
| - Flag 1 Removed (Effect) | 27 | 3.6% | 0.42 | [0.23; 0.61] | 8.7% | 0.1239 | 14.8% | [-0.33; 1.17] | 8.48 |
| - Flag 2 Removed (Power) | 13 | 53.6% | 0.30 | [0.05; 0.56] | 34.8% | 0.1140 | 18.3% | [-0.47; 1.08] | 12.19 |
| - Flag 3 Removed (RoB) | 3 | 89.3% | 0.53 | [-0.45; 1.52] | -15% | 0.0860 | 29.0% | [-1.04; 2.11] | 6.34 |
| ***Problem Gambling*** | | | | | | | | | |
| All Effect Sizes Included | 81 | *ref.* | 0.80 | [0.43; 1.17] | *ref.* | 0.8725 | *ref.* | [-1.09; 2.69] | 4.65 |
| - Flag 1 Removed (Effect) | 75 | 7.4% | 0.55 | [0.35; 0.76] | 31.2% | 0.2083 | 51.1% | [-0.38; 1.48] | 7.49 |
| - Flag 2 Removed (Power) | 53 | 34.6% | 0.46 | [0.22; 0.70] | 42.5% | 0.2268 | 49.0% | [-0.52; 1.44] | 9.46 |
| - Flag 3 Removed (RoB) | 8 | 90.1% | 0.09 | [-0.46; 0.65] | 88.8% | 0.0000 | 100.0% | [-0.12; 0.31] | 57.38 |

†Not enough effect sizes available for meta-analysis ($k$=3).

*Notes*. Reanalysis of Harrer et al. (2025); $\Delta_k$ = percent difference in the number of available effect sizes compared to the "All Effect Sizes Included" analysis; $\Delta_{SMD}$ = percent difference in the pooled effect size (SMD) compared to the "All Effect Sizes Included" analysis; $\Delta_\tau$ = percent difference in between-study heterogeneity variance compared to the "All Effect Sizes Included" analysis; SMD = standardized mean difference; $\tau^2$ = between-study heterogeneity variance; PI = prediction interval; NNT = number needed to treat.

**S7.** Recalculation of subgroup-specific effects when flagged studies are excluded.

| Intervention | Control | Analysis | $k$ | $\Delta_k$ | SMD | 95%-CI | $\Delta_{SMD}$ | $I^2$ | 95%-CI | $\Delta_{het}$ | NNT |
|---|---|---|---|---|---|---|---|---|---|---|---|
| ***Borderline Personality Disorder*** | | | | | | | | | | | |
| CBT | Care As Usual | All Effect Sizes Included | 3 | 0.0% | 0.36 | [-1.52; 2.24] | 0.0% | 80.4 | [38.3; 93.8] | 0.0% | 10.11 |
| CBT | Care As Usual | - 3 Flags Removed | 3 | 0.0% | 0.36 | [-1.52; 2.24] | 0.0% | 80.4 | [38.3; 93.8] | 0.0% | 10.11 |
| CBT | Care As Usual | - At Least 2 Flags Removed | 1 | 66.7% | -0.23 | [-0.63; 0.16] | 163.9% | | - | | 16.59 |
| CBT | Care As Usual | - Flag 1 Removed (Effect) | 3 | 0.0% | 0.36 | [-1.52; 2.24] | 0.0% | 80.4 | [38.3; 93.8] | 0.0% | 10.11 |
| CBT | Care As Usual | - Flag 2 Removed (Power) | 1 | 66.7% | -0.23 | [-0.63; 0.16] | 163.9% | | - | | 16.59 |
| DBT | Care As Usual | All Effect Sizes Included | 10 | 0.0% | 0.38 | [0.11; 0.64] | 0.0% | 34.7 | [0; 68.9] | 0.0% | 9.52 |
| DBT | Care As Usual | - 3 Flags Removed | 10 | 0.0% | 0.38 | [0.11; 0.64] | 0.0% | 34.7 | [0; 68.9] | 0.0% | 9.52 |
| DBT | Care As Usual | - At Least 2 Flags Removed | 6 | 40.0% | 0.41 | [0.05; 0.77] | -7.9% | 43.9 | [0; 77.8] | -26.5% | 8.68 |
| DBT | Care As Usual | - Flag 1 Removed (Effect) | 10 | 0.0% | 0.38 | [0.11; 0.64] | 0.0% | 34.7 | [0; 68.9] | 0.0% | 9.52 |
| DBT | Care As Usual | - Flag 2 Removed (Power) | 6 | 40.0% | 0.41 | [0.05; 0.77] | -7.9% | 43.9 | [0; 77.8] | -26.5% | 8.68 |
| DBT | Care As Usual | - Flag 3 Removed (RoB) | 1 | 90.0% | 0.91 | [0.39; 1.43] | -139.5% | | - | | 3.33 |
| IPT | Care As Usual | All Effect Sizes Included | 2 | 0.0% | 0.59 | [-6.53; 7.7] | 0.0% | 84.4 | [36.1; 96.2] | 0.0% | 5.68 |
| IPT | Care As Usual | - 3 Flags Removed | 2 | 0.0% | 0.59 | [-6.53; 7.7] | 0.0% | 84.4 | [36.1; 96.2] | 0.0% | 5.68 |
| IPT | Care As Usual | - At Least 2 Flags Removed | 1 | 50.0% | 0.03 | [-0.56; 0.62] | 94.9% | | - | | 136.16 |
| IPT | Care As Usual | - Flag 1 Removed (Effect) | 2 | 0.0% | 0.59 | [-6.53; 7.7] | 0.0% | 84.4 | [36.1; 96.2] | 0.0% | 5.68 |
| IPT | Care As Usual | - Flag 2 Removed (Power) | 1 | 50.0% | 0.03 | [-0.56; 0.62] | 94.9% | | - | | 136.16 |
| MBT | Care As Usual | All Effect Sizes Included | 3 | 0.0% | 0.49 | [-0.69; 1.66] | 0.0% | 80.5 | [38.8; 93.8] | 0.0% | 7.08 |
| MBT | Care As Usual | - 3 Flags Removed | 3 | 0.0% | 0.49 | [-0.69; 1.66] | 0.0% | 80.5 | [38.8; 93.8] | 0.0% | 7.08 |
| MBT | Care As Usual | - At Least 2 Flags Removed | 2 | 33.3% | 0.51 | [-5.07; 6.1] | -4.1% | 90.1 | [63.9; 97.3] | -11.9% | 6.65 |
| MBT | Care As Usual | - Flag 1 Removed (Effect) | 3 | 0.0% | 0.49 | [-0.69; 1.66] | 0.0% | 80.5 | [38.8; 93.8] | 0.0% | 7.08 |
| MBT | Care As Usual | - Flag 2 Removed (Power) | 2 | 33.3% | 0.51 | [-5.07; 6.1] | -4.1% | 90.1 | [63.9; 97.3] | -11.9% | 6.65 |
| Mixed | Care As Usual | All Effect Sizes Included | 2 | 0.0% | 0 | [-4.45; 4.46] | | 65.8 | [0; 92.2] | 0.0% | 2322.12 |
| Mixed | Care As Usual | - 3 Flags Removed | 2 | 0.0% | 0 | [-4.45; 4.46] | | 65.8 | [0; 92.2] | 0.0% | 2322.12 |
| Mixed | Care As Usual | - At Least 2 Flags Removed | 2 | 0.0% | 0 | [-4.45; 4.46] | | 65.8 | [0; 92.2] | 0.0% | 2322.12 |
| Mixed | Care As Usual | - Flag 1 Removed (Effect) | 2 | 0.0% | 0 | [-4.45; 4.46] | | 65.8 | [0; 92.2] | 0.0% | 2322.12 |
| Mixed | Care As Usual | - Flag 2 Removed (Power) | 2 | 0.0% | 0 | [-4.45; 4.46] | | 65.8 | [0; 92.2] | 0.0% | 2322.12 |
| PDP | Care As Usual | All Effect Sizes Included | 4 | 0.0% | 0.69 | [-0.41; 1.79] | 0.0% | 66.6 | [2.3; 88.6] | 0.0% | 4.64 |
| PDP | Care As Usual | - 3 Flags Removed | 4 | 0.0% | 0.69 | [-0.41; 1.79] | 0.0% | 66.6 | [2.3; 88.6] | 0.0% | 4.64 |
| PDP | Care As Usual | - At Least 2 Flags Removed | 1 | 75.0% | 0 | [-0.68; 0.68] | 100.0% | | - | | |
| PDP | Care As Usual | - Flag 1 Removed (Effect) | 4 | 0.0% | 0.69 | [-0.41; 1.79] | 0.0% | 66.6 | [2.3; 88.6] | 0.0% | 4.64 |
| PDP | Care As Usual | - Flag 3 Removed (RoB) | 1 | 75.0% | 0 | [-0.68; 0.68] | 100.0% | | - | | |
| ST | Care As Usual | All Effect Sizes Included | 2 | 0.0% | 1.35 | [-8.52; 11.22] | 0.0% | 84.8 | [38.2; 96.3] | 0.0% | 2.12 |
| ST | Care As Usual | - 3 Flags Removed | 1 | 50.0% | 0.6 | [-0.11; 1.31] | 55.6% | | - | | 5.51 |

| Intervention | Control | Analysis | *k* | $\Delta_k$ | SMD | 95%-CI | $\Delta_{SMD}$ | $I^2$ | 95%-CI | $\Delta_{het}$ | NNT |
|---|---|---|---|---|---|---|---|---|---|---|---|
| ST | Care As Usual | - Flag 1 Removed (Effect) | 1 | 50.0% | 0.6 | [-0.11; 1.31] | 55.6% | | - | | 5.51 |
| TFP | Care As Usual | All Effect Sizes Included | 1 | 0.0% | 0.55 | [0.16; 0.95] | 0.0% | | - | | 6.07 |
| TFP | Care As Usual | - 3 Flags Removed | 1 | 0.0% | 0.55 | [0.16; 0.95] | 0.0% | | - | | 6.07 |
| TFP | Care As Usual | - At Least 2 Flags Removed | 1 | 0.0% | 0.55 | [0.16; 0.95] | 0.0% | | - | | 6.07 |
| TFP | Care As Usual | - Flag 1 Removed (Effect) | 1 | 0.0% | 0.55 | [0.16; 0.95] | 0.0% | | - | | 6.07 |
| TFP | Care As Usual | - Flag 2 Removed (Power) | 1 | 0.0% | 0.55 | [0.16; 0.95] | 0.0% | | - | | 6.07 |
| TFP | Care As Usual | - Flag 3 Removed (RoB) | 1 | 0.0% | 0.55 | [0.16; 0.95] | 0.0% | | - | | 6.07 |
| ***Depression*** | | | | | | | | | | | |
| 3rd Wave | Care As Usual | All Effect Sizes Included | 12 | 0.0% | 0.82 | [0.47; 1.17] | 0.0% | 75.2 | [56.3; 85.9] | 0.0% | 3.44 |
| 3rd Wave | Care As Usual | - 3 Flags Removed | 11 | 8.3% | 0.73 | [0.4; 1.05] | 11.0% | 68.7 | [41.5; 83.3] | 8.6% | 3.93 |
| 3rd Wave | Care As Usual | - At Least 2 Flags Removed | 8 | 33.3% | 0.53 | [0.24; 0.83] | 35.4% | 47.6 | [0; 76.7] | 36.7% | 5.59 |
| 3rd Wave | Care As Usual | - At Least 1 Flag Removed | 4 | 66.7% | 0.44 | [0.01; 0.86] | 46.3% | 46.4 | [0; 82.2] | 38.3% | 7.03 |
| 3rd Wave | Care As Usual | - Flag 1 Removed (Effect) | 11 | 8.3% | 0.73 | [0.4; 1.05] | 11.0% | 68.7 | [41.5; 83.3] | 8.6% | 3.93 |
| 3rd Wave | Care As Usual | - Flag 2 Removed (Power) | 4 | 66.7% | 0.44 | [0.01; 0.86] | 46.3% | 46.4 | [0; 82.2] | 38.3% | 7.03 |
| 3rd Wave | Care As Usual | - Flag 3 Removed (RoB) | 8 | 33.3% | 0.53 | [0.24; 0.83] | 35.4% | 47.6 | [0; 76.7] | 36.7% | 5.59 |
| BAT | Care As Usual | All Effect Sizes Included | 24 | 0.0% | 0.66 | [0.45; 0.86] | 0.0% | 79.7 | [70.5; 86.1] | 0.0% | 4.42 |
| BAT | Care As Usual | - 3 Flags Removed | 24 | 0.0% | 0.66 | [0.45; 0.86] | 0.0% | 79.7 | [70.5; 86] | 0.0% | 4.42 |
| BAT | Care As Usual | - At Least 2 Flags Removed | 18 | 25.0% | 0.56 | [0.37; 0.75] | 15.2% | 79 | [67.4; 86.4] | 0.9% | 5.25 |
| BAT | Care As Usual | - At Least 1 Flag Removed | 8 | 66.7% | 0.54 | [0.28; 0.8] | 18.2% | 78.3 | [57.2; 88.9] | 1.8% | 5.51 |
| BAT | Care As Usual | - Flag 1 Removed (Effect) | 23 | 4.2% | 0.61 | [0.42; 0.79] | 7.6% | 77.6 | [66.8; 84.9] | 2.6% | 4.81 |
| BAT | Care As Usual | - Flag 2 Removed (Power) | 14 | 41.7% | 0.56 | [0.38; 0.73] | 15.2% | 71.4 | [50.8; 83.3] | 10.4% | 5.33 |
| BAT | Care As Usual | - Flag 3 Removed (RoB) | 13 | 45.8% | 0.63 | [0.3; 0.95] | 4.5% | 85.7 | [77.2; 91] | -7.5% | 4.64 |
| CBT | Care As Usual | All Effect Sizes Included | 128 | 0.0% | 0.68 | [0.55; 0.82] | 0.0% | 87.6 | [85.7; 89.2] | 0.0% | 4.21 |
| CBT | Care As Usual | - 3 Flags Removed | 122 | 4.7% | 0.58 | [0.49; 0.68] | 14.7% | 84.3 | [81.8; 86.6] | 3.8% | 5.05 |
| CBT | Care As Usual | - At Least 2 Flags Removed | 89 | 30.5% | 0.53 | [0.44; 0.61] | 22.1% | 84.5 | [81.5; 87] | 3.5% | 5.67 |
| CBT | Care As Usual | - At Least 1 Flag Removed | 29 | 77.3% | 0.43 | [0.3; 0.57] | 36.8% | 77 | [67.3; 83.8] | 12.1% | 7.06 |
| CBT | Care As Usual | - Flag 1 Removed (Effect) | 119 | 7.0% | 0.53 | [0.45; 0.61] | 22.1% | 81.8 | [78.6; 84.5] | 6.6% | 5.62 |
| CBT | Care As Usual | - Flag 2 Removed (Power) | 72 | 43.8% | 0.51 | [0.41; 0.61] | 25.0% | 87.5 | [84.9; 89.6] | 0.1% | 5.83 |
| CBT | Care As Usual | - Flag 3 Removed (RoB) | 49 | 61.7% | 0.62 | [0.46; 0.79] | 8.8% | 82 | [76.9; 86] | 6.4% | 4.71 |
| Psychodynamic | Care As Usual | All Effect Sizes Included | 8 | 0.0% | 0.33 | [0.05; 0.61] | 0.0% | 70.6 | [39.2; 85.8] | 0.0% | 9.63 |
| Psychodynamic | Care As Usual | - 3 Flags Removed | 8 | 0.0% | 0.33 | [0.05; 0.61] | 0.0% | 70.6 | [39.2; 85.8] | 0.0% | 9.63 |
| Psychodynamic | Care As Usual | - At Least 2 Flags Removed | 7 | 12.5% | 0.36 | [0.05; 0.66] | -9.1% | 72.9 | [41.7; 87.4] | -3.3% | 8.81 |
| Psychodynamic | Care As Usual | - At Least 1 Flag Removed | 4 | 50.0% | 0.34 | [-0.08; 0.77] | -3.0% | 54.9 | [0; 85.1] | 22.2% | 9.18 |
| Psychodynamic | Care As Usual | - Flag 1 Removed (Effect) | 8 | 0.0% | 0.33 | [0.05; 0.61] | 0.0% | 70.6 | [39.2; 85.8] | 0.0% | 9.63 |
| Psychodynamic | Care As Usual | - Flag 2 Removed (Power) | 6 | 25.0% | 0.3 | [0; 0.59] | 9.1% | 70.5 | [31.1; 87.4] | 0.1% | 10.7 |

| Intervention | Control | Analysis | $k$ | $\Delta_k$ | SMD | 95%-CI | $\Delta_{SMD}$ | $I^2$ | 95%-CI | $\Delta_{het}$ | NNT |
|---|---|---|---|---|---|---|---|---|---|---|---|
| Psychodynamic | Care As Usual | - Flag 3 Removed (RoB) | 5 | 37.5% | 0.41 | [0.01; 0.82] | -24.2% | 60.5 | [0; 85.2] | 14.3% | 7.45 |
| IPT | Care As Usual | All Effect Sizes Included | 24 | 0.0% | 0.44 | [0.26; 0.62] | 0.0% | 76 | [64.4; 83.8] | 0.0% | 6.9 |
| IPT | Care As Usual | - 3 Flags Removed | 24 | 0.0% | 0.44 | [0.26; 0.62] | 0.0% | 76 | [64.4; 83.8] | 0.0% | 6.9 |
| IPT | Care As Usual | - At Least 2 Flags Removed | 14 | 41.7% | 0.35 | [0.1; 0.59] | 20.5% | 83 | [72.7; 89.4] | -9.2% | 9.02 |
| IPT | Care As Usual | - At Least 1 Flag Removed | 2 | 91.7% | 0.14 | [-0.96; 1.23] | 68.2% | 0 | - | 100.0% | 24.79 |
| IPT | Care As Usual | - Flag 1 Removed (Effect) | 24 | 0.0% | 0.44 | [0.26; 0.62] | 0.0% | 76 | [64.4; 83.8] | 0.0% | 6.9 |
| IPT | Care As Usual | - Flag 2 Removed (Power) | 11 | 54.2% | 0.42 | [0.15; 0.69] | 4.5% | 84.7 | [74.3; 90.9] | -11.4% | 7.33 |
| IPT | Care As Usual | - Flag 3 Removed (RoB) | 5 | 79.2% | 0.08 | [-0.21; 0.36] | 81.8% | 7 | [0; 80.7] | 90.8% | 45.88 |
| LRT | Care As Usual | All Effect Sizes Included | 7 | 0.0% | 1.7 | [1.11; 2.3] | 0.0% | 71.3 | [37.6; 86.8] | 0.0% | 1.65 |
| LRT | Care As Usual | - 3 Flags Removed | 5 | 28.6% | 1.48 | [0.84; 2.12] | 12.9% | 65.2 | [8.9; 86.7] | 8.6% | 1.86 |
| LRT | Care As Usual | - At Least 2 Flags Removed | 1 | 85.7% | 2 | [1.5; 2.51] | -17.6% |  | - |  | 1.48 |
| LRT | Care As Usual | - Flag 1 Removed (Effect) | 4 | 42.9% | 1.19 | [0.81; 1.58] | 30.0% | 0 | [0; 84.7] | 100.0% | 2.29 |
| LRT | Care As Usual | - Flag 2 Removed (Power) | 1 | 85.7% | 2 | [1.5; 2.51] | -17.6% |  | - |  | 1.48 |
| LRT | Care As Usual | - Flag 3 Removed (RoB) | 1 | 85.7% | 2.2 | [1.73; 2.67] | -29.4% |  | - |  | 1.4 |
| Other | Care As Usual | All Effect Sizes Included | 32 | 0.0% | 0.4 | [0.26; 0.54] | 0.0% | 82.3 | [75.8; 87] | 0.0% | 7.72 |
| Other | Care As Usual | - 3 Flags Removed | 31 | 3.1% | 0.38 | [0.25; 0.51] | 5.0% | 81.8 | [75; 86.8] | 0.6% | 8.21 |
| Other | Care As Usual | - At Least 2 Flags Removed | 27 | 15.6% | 0.37 | [0.23; 0.51] | 7.5% | 83.9 | [77.6; 88.4] | -1.9% | 8.34 |
| Other | Care As Usual | - At Least 1 Flag Removed | 17 | 46.9% | 0.34 | [0.14; 0.53] | 15.0% | 78.5 | [66.1; 86.3] | 4.6% | 9.34 |
| Other | Care As Usual | - Flag 1 Removed (Effect) | 31 | 3.1% | 0.38 | [0.25; 0.51] | 5.0% | 81.8 | [75; 86.8] | 0.6% | 8.21 |
| Other | Care As Usual | - Flag 2 Removed (Power) | 27 | 15.6% | 0.37 | [0.23; 0.51] | 7.5% | 83.8 | [77.5; 88.4] | -1.8% | 8.38 |
| Other | Care As Usual | - Flag 3 Removed (RoB) | 17 | 46.9% | 0.34 | [0.14; 0.54] | 15.0% | 78.7 | [66.4; 86.4] | 4.4% | 9.26 |
| PST | Care As Usual | All Effect Sizes Included | 13 | 0.0% | 0.57 | [0.24; 0.9] | 0.0% | 83.8 | [73.7; 90] | 0.0% | 5.15 |
| PST | Care As Usual | - 3 Flags Removed | 13 | 0.0% | 0.55 | [0.24; 0.86] | 3.5% | 82.2 | [70.8; 89.2] | 1.9% | 5.43 |
| PST | Care As Usual | - At Least 2 Flags Removed | 8 | 38.5% | 0.55 | [0.12; 0.98] | 3.5% | 87.1 | [76.8; 92.9] | -3.9% | 5.39 |
| PST | Care As Usual | - At Least 1 Flag Removed | 2 | 84.6% | 0.33 | [-0.73; 1.38] | 42.1% | 0 | - | 100.0% | 9.64 |
| PST | Care As Usual | - Flag 1 Removed (Effect) | 13 | 0.0% | 0.54 | [0.24; 0.84] | 5.3% | 80.9 | [68.3; 88.5] | 3.5% | 5.49 |
| PST | Care As Usual | - Flag 2 Removed (Power) | 7 | 46.2% | 0.55 | [0.04; 1.07] | 3.5% | 89.8 | [81.5; 94.4] | -7.2% | 5.38 |
| PST | Care As Usual | - Flag 3 Removed (RoB) | 3 | 76.9% | 0.35 | [0.01; 0.68] | 38.6% | 0 | [0; 89.6] | 100.0% | 9.02 |
| SUP | Care As Usual | All Effect Sizes Included | 13 | 0.0% | 0.41 | [0.27; 0.54] | 0.0% | 7.9 | [0; 45.4] | 0.0% | 7.58 |
| SUP | Care As Usual | - 3 Flags Removed | 13 | 0.0% | 0.41 | [0.27; 0.54] | 0.0% | 7.9 | [0; 45.4] | 0.0% | 7.58 |
| SUP | Care As Usual | - At Least 2 Flags Removed | 8 | 38.5% | 0.37 | [0.23; 0.52] | 9.8% | 0 | [0; 67.6] | 100.0% | 8.32 |
| SUP | Care As Usual | - At Least 1 Flag Removed | 3 | 76.9% | 0.28 | [-0.02; 0.57] | 31.7% | 0 | [0; 89.6] | 100.0% | 11.52 |
| SUP | Care As Usual | - Flag 1 Removed (Effect) | 13 | 0.0% | 0.41 | [0.27; 0.54] | 0.0% | 7.9 | [0; 45.4] | 0.0% | 7.58 |
| SUP | Care As Usual | - Flag 2 Removed (Power) | 6 | 53.8% | 0.33 | [0.2; 0.46] | 19.5% | 0 | [0; 74.6] | 100.0% | 9.53 |
| SUP | Care As Usual | - Flag 3 Removed (RoB) | 5 | 61.5% | 0.37 | [0.1; 0.64] | 9.8% | 23.5 | [0; 68.6] | -197.5% | 8.4 |
| 3rd Wave | Waitlist | All Effect Sizes Included | 20 | 0.0% | 0.95 | [0.54; 1.36] | 0.0% | 77.1 | [64.9; 85] | 0.0% | 2.92 |

| Intervention | Control | Analysis | $k$ | $\Delta_k$ | SMD | 95%-CI | $\Delta_{SMD}$ | $I^2$ | 95%-CI | $\Delta_{het}$ | NNT |
|---|---|---|---|---|---|---|---|---|---|---|---|
| 3rd Wave | Waitlist | - 3 Flags Removed | 18 | 10.0% | 0.68 | [0.53; 0.83] | 28.4% | 37.9 | [0; 64.7] | 50.8% | 4.24 |
| 3rd Wave | Waitlist | - At Least 2 Flags Removed | 8 | 60.0% | 0.64 | [0.51; 0.78] | 32.6% | 0 | [0; 67.6] | 100.0% | 4.51 |
| 3rd Wave | Waitlist | - At Least 1 Flag Removed | 4 | 80.0% | 0.59 | [0.45; 0.73] | 37.9% | 0 | [0; 84.7] | 100.0% | 4.97 |
| 3rd Wave | Waitlist | - Flag 1 Removed (Effect) | 18 | 10.0% | 0.68 | [0.53; 0.83] | 28.4% | 37.9 | [0; 64.7] | 50.8% | 4.24 |
| 3rd Wave | Waitlist | - Flag 2 Removed (Power) | 8 | 60.0% | 0.65 | [0.51; 0.78] | 31.6% | 0 | [0; 67.6] | 100.0% | 4.48 |
| 3rd Wave | Waitlist | - Flag 3 Removed (RoB) | 4 | 80.0% | 0.59 | [0.47; 0.7] | 37.9% | 0 | [0; 84.7] | 100.0% | 5.03 |
| BAT | Waitlist | All Effect Sizes Included | 12 | 0.0% | 0.97 | [0.74; 1.21] | 0.0% | 17.9 | [0; 57.2] | 0.0% | 2.84 |
| BAT | Waitlist | - 3 Flags Removed | 10 | 16.7% | 0.87 | [0.72; 1.03] | 10.3% | 0 | [0; 62.4] | 100.0% | 3.2 |
| BAT | Waitlist | - At Least 2 Flags Removed | 3 | 75.0% | 0.82 | [0.37; 1.26] | 15.5% | 0 | [0; 89.6] | 100.0% | 3.45 |
| BAT | Waitlist | - At Least 1 Flag Removed | 2 | 83.3% | 0.81 | [-1.24; 2.87] | 16.5% | 23.8 | - | -33.0% | 3.46 |
| BAT | Waitlist | - Flag 1 Removed (Effect) | 10 | 16.7% | 0.87 | [0.72; 1.03] | 10.3% | 0 | [0; 62.4] | 100.0% | 3.2 |
| BAT | Waitlist | - Flag 2 Removed (Power) | 2 | 83.3% | 0.81 | [-1.24; 2.87] | 16.5% | 23.8 | - | -33.0% | 3.46 |
| BAT | Waitlist | - Flag 3 Removed (RoB) | 3 | 75.0% | 0.82 | [0.37; 1.26] | 15.5% | 0 | [0; 89.6] | 100.0% | 3.45 |
| CBT | Waitlist | All Effect Sizes Included | 129 | 0.0% | 0.96 | [0.83; 1.09] | 0.0% | 80.5 | [77.2; 83.4] | 0.0% | 2.89 |
| CBT | Waitlist | - 3 Flags Removed | 119 | 7.8% | 0.8 | [0.71; 0.89] | 16.7% | 70.6 | [64.6; 75.5] | 12.3% | 3.53 |
| CBT | Waitlist | - At Least 2 Flags Removed | 57 | 55.8% | 0.72 | [0.63; 0.81] | 25.0% | 72.5 | [64.3; 78.8] | 9.9% | 4 |
| CBT | Waitlist | - At Least 1 Flag Removed | 26 | 79.8% | 0.63 | [0.52; 0.74] | 34.4% | 70.2 | [55.7; 80] | 12.8% | 4.63 |
| CBT | Waitlist | - Flag 1 Removed (Effect) | 116 | 10.1% | 0.74 | [0.68; 0.8] | 22.9% | 52.2 | [40.8; 61.3] | 35.2% | 3.85 |
| CBT | Waitlist | - Flag 2 Removed (Power) | 50 | 61.2% | 0.7 | [0.61; 0.8] | 27.1% | 73.8 | [65.4; 80.1] | 8.3% | 4.08 |
| CBT | Waitlist | - Flag 3 Removed (RoB) | 36 | 72.1% | 0.85 | [0.57; 1.12] | 11.5% | 87.1 | [83.2; 90.2] | -8.2% | 3.31 |
| Psychodynamic | Waitlist | All Effect Sizes Included | 1 | 0.0% | 1.09 | [0.14; 2.03] | 0.0% | - | - |  | 2.52 |
| Psychodynamic | Waitlist | - 3 Flags Removed | 1 | 0.0% | 1.09 | [0.14; 2.03] | 0.0% | - | - |  | 2.52 |
| Psychodynamic | Waitlist | - Flag 1 Removed (Effect) | 1 | 0.0% | 1.09 | [0.14; 2.03] | 0.0% | - | - |  | 2.52 |
| IPT | Waitlist | All Effect Sizes Included | 4 | 0.0% | 0.56 | [-0.01; 1.14] | 0.0% | 57.8 | [0; 86] | 0.0% | 5.25 |
| IPT | Waitlist | - 3 Flags Removed | 4 | 0.0% | 0.56 | [-0.01; 1.14] | 0.0% | 57.8 | [0; 86] | 0.0% | 5.25 |
| IPT | Waitlist | - At Least 2 Flags Removed | 2 | 50.0% | 0.57 | [-4.7; 5.85] | -1.8% | 87.3 | [50.2; 96.7] | -51.0% | 5.15 |
| IPT | Waitlist | - Flag 1 Removed (Effect) | 4 | 0.0% | 0.56 | [-0.01; 1.14] | 0.0% | 57.8 | [0; 86] | 0.0% | 5.25 |
| IPT | Waitlist | - Flag 2 Removed (Power) | 2 | 50.0% | 0.57 | [-4.7; 5.85] | -1.8% | 87.3 | [50.2; 96.7] | -51.0% | 5.15 |
| LRT | Waitlist | All Effect Sizes Included | 5 | 0.0% | 0.56 | [0.25; 0.87] | 0.0% | 26.8 | [0; 71] | 0.0% | 5.26 |
| LRT | Waitlist | - 3 Flags Removed | 5 | 0.0% | 0.56 | [0.25; 0.87] | 0.0% | 26.8 | [0; 71] | 0.0% | 5.26 |
| LRT | Waitlist | - At Least 2 Flags Removed | 3 | 40.0% | 0.59 | [-0.18; 1.36] | -5.4% | 59.2 | [0; 88.4] | -120.9% | 4.98 |
| LRT | Waitlist | - At Least 1 Flag Removed | 2 | 60.0% | 0.46 | [-0.62; 1.53] | 17.9% | 0 | - | 100.0% | 6.69 |
| LRT | Waitlist | - Flag 1 Removed (Effect) | 5 | 0.0% | 0.56 | [0.25; 0.87] | 0.0% | 26.8 | [0; 71] | 0.0% | 5.26 |
| LRT | Waitlist | - Flag 2 Removed (Power) | 3 | 40.0% | 0.59 | [-0.18; 1.36] | -5.4% | 59.2 | [0; 88.4] | -120.9% | 4.98 |
| LRT | Waitlist | - Flag 3 Removed (RoB) | 2 | 60.0% | 0.46 | [-0.62; 1.53] | 17.9% | 0 | - | 100.0% | 6.69 |
| Other | Waitlist | All Effect Sizes Included | 27 | 0.0% | 0.46 | [0.36; 0.57] | 0.0% | 10.8 | [0; 43.3] | 0.0% | 6.58 |

| Intervention | Control | Analysis | *k* | $\Delta_k$ | SMD | 95%-CI | $\Delta_{SMD}$ | $I^2$ | 95%-CI | $\Delta_{het}$ | NNT |
|---|---|---|---|---|---|---|---|---|---|---|---|
| Other | Waitlist | - 3 Flags Removed | 27 | 0.0% | 0.46 | [0.36; 0.57] | 0.0% | 10.8 | [0; 43.3] | 0.0% | 6.58 |
| Other | Waitlist | - At Least 2 Flags Removed | 11 | 59.3% | 0.4 | [0.26; 0.53] | 13.0% | 15.7 | [0; 56.1] | -45.4% | 7.81 |
| Other | Waitlist | - At Least 1 Flag Removed | 5 | 81.5% | 0.47 | [0.31; 0.63] | -2.2% | 0 | [0; 79.2] | 100.0% | 6.49 |
| Other | Waitlist | - Flag 1 Removed (Effect) | 27 | 0.0% | 0.47 | [0.37; 0.57] | -2.2% | 10.7 | [0; 43.2] | 0.9% | 6.44 |
| Other | Waitlist | - Flag 2 Removed (Power) | 8 | 70.4% | 0.4 | [0.27; 0.53] | 13.0% | 0 | [0; 67.6] | 100.0% | 7.66 |
| Other | Waitlist | - Flag 3 Removed (RoB) | 8 | 70.4% | 0.44 | [0.28; 0.6] | 4.3% | 16.5 | [0; 59.5] | -52.8% | 6.93 |
| PST | Waitlist | All Effect Sizes Included | 15 | 0.0% | 0.81 | [0.41; 1.2] | 0.0% | 81.1 | [69.9; 88.2] | 0.0% | 3.5 |
| PST | Waitlist | - 3 Flags Removed | 13 | 13.3% | 0.58 | [0.29; 0.86] | 28.4% | 72.6 | [52.3; 84.3] | 10.5% | 5.11 |
| PST | Waitlist | - At Least 2 Flags Removed | 6 | 60.0% | 0.36 | [0.03; 0.69] | 55.6% | 75.1 | [43.7; 89] | 7.4% | 8.66 |
| PST | Waitlist | - At Least 1 Flag Removed | 4 | 73.3% | 0.3 | [-0.25; 0.86] | 63.0% | 83.4 | [57.9; 93.5] | -2.8% | 10.45 |
| PST | Waitlist | - Flag 1 Removed (Effect) | 13 | 13.3% | 0.58 | [0.29; 0.86] | 28.4% | 72.6 | [52.3; 84.3] | 10.5% | 5.11 |
| PST | Waitlist | - Flag 2 Removed (Power) | 5 | 66.7% | 0.33 | [-0.07; 0.73] | 59.3% | 78.8 | [49.3; 91.1] | 2.8% | 9.55 |
| PST | Waitlist | - Flag 3 Removed (RoB) | 5 | 66.7% | 0.35 | [-0.07; 0.76] | 56.8% | 79.5 | [51.3; 91.3] | 2.0% | 9.09 |
| SUP | Waitlist | All Effect Sizes Included | 3 | 0.0% | 1.05 | [-1.25; 3.36] | 0.0% | 80.2 | [37.4; 93.7] | 0.0% | 2.6 |
| SUP | Waitlist | - 3 Flags Removed | 2 | 33.3% | 0.59 | [-6.21; 7.4] | 43.8% | 63.8 | [0; 91.7] | 20.4% | 4.98 |
| SUP | Waitlist | - Flag 1 Removed (Effect) | 2 | 33.3% | 0.59 | [-6.21; 7.4] | 43.8% | 63.8 | [0; 91.7] | 20.4% | 4.98 |
| ***<u>Gambling</u>*** | | | | | | | | | | | |
| CBT | Care As Usual | All Effect Sizes Included | 7 | 0.0% | 0.24 | [-0.08; 0.55] | 0.0% | 68.6 | [30.7; 85.8] | 0.0% | 20.85 |
| CBT | Care As Usual | - 3 Flags Removed | 7 | 0.0% | 0.24 | [-0.08; 0.55] | 0.0% | 68.6 | [30.7; 85.8] | 0.0% | 20.85 |
| CBT | Care As Usual | - At Least 2 Flags Removed | 6 | 14.3% | 0.25 | [-0.12; 0.63] | -4.2% | 73.8 | [40.1; 88.5] | -7.6% | 19.21 |
| CBT | Care As Usual | - Flag 1 Removed (Effect) | 7 | 0.0% | 0.24 | [-0.08; 0.55] | 0.0% | 68.6 | [30.7; 85.8] | 0.0% | 20.85 |
| CBT | Care As Usual | - Flag 2 Removed (Power) | 6 | 14.3% | 0.25 | [-0.12; 0.63] | -4.2% | 73.8 | [40.1; 88.5] | -7.6% | 19.21 |
| CBT | Waitlist | All Effect Sizes Included | 24 | 0.0% | 0.74 | [0.38; 1.1] | 0.0% | 84.8 | [78.5; 89.2] | 0.0% | 5.12 |
| CBT | Waitlist | - 3 Flags Removed | 23 | 4.2% | 0.61 | [0.39; 0.82] | 17.6% | 77.7 | [67; 84.9] | 8.4% | 6.65 |
| CBT | Waitlist | - At Least 2 Flags Removed | 16 | 33.3% | 0.49 | [0.22; 0.76] | 33.8% | 80.8 | [69.7; 87.8] | 4.7% | 8.67 |
| CBT | Waitlist | - Flag 1 Removed (Effect) | 23 | 4.2% | 0.61 | [0.39; 0.82] | 17.6% | 77.7 | [67; 84.9] | 8.4% | 6.65 |
| CBT | Waitlist | - Flag 2 Removed (Power) | 16 | 33.3% | 0.49 | [0.22; 0.76] | 33.8% | 80.8 | [69.7; 87.8] | 4.7% | 8.67 |
| ***<u>Generalized Anxiety Disorder</u>*** | | | | | | | | | | | |
| 3rd Wave | Care As Usual | All Effect Sizes Included | 3 | 0.0% | 0.43 | [0.34; 0.52] | 0.0% | 0 | [0; 89.6] | | 8.4 |
| 3rd Wave | Care As Usual | - 3 Flags Removed | 3 | 0.0% | 0.43 | [0.34; 0.52] | 0.0% | 0 | [0; 89.6] | | 8.4 |
| 3rd Wave | Care As Usual | - At Least 2 Flags Removed | 2 | 33.3% | 0.43 | [0.06; 0.8] | 0.0% | 0 | - | | 8.47 |
| 3rd Wave | Care As Usual | - Flag 1 Removed (Effect) | 3 | 0.0% | 0.43 | [0.34; 0.52] | 0.0% | 0 | [0; 89.6] | | 8.4 |
| 3rd Wave | Care As Usual | - Flag 2 Removed (Power) | 2 | 33.3% | 0.43 | [0.06; 0.8] | 0.0% | 0 | - | | 8.47 |
| AR | Care As Usual | All Effect Sizes Included | 1 | 0.0% | 0.76 | [-0.33; 1.85] | 0.0% | | - | | 4.27 |

| Intervention | Control | Analysis | *k* | $\Delta_k$ | SMD | 95%-CI | $\Delta_{SMD}$ | $I^2$ | 95%-CI | $\Delta_{het}$ | NNT |
|---|---|---|---|---|---|---|---|---|---|---|---|
| AR | Care As Usual | - 3 Flags Removed | 1 | 0.0% | 0.76 | [-0.33; 1.85] | 0.0% | | - | | 4.27 |
| AR | Care As Usual | - Flag 1 Removed (Effect) | 1 | 0.0% | 0.76 | [-0.33; 1.85] | 0.0% | | - | | 4.27 |
| CBT | Care As Usual | All Effect Sizes Included | 6 | 0.0% | 0.96 | [0.09; 1.84] | 0.0% | 86.8 | [73.6; 93.4] | 0.0% | 3.19 |
| CBT | Care As Usual | - 3 Flags Removed | 5 | 16.7% | 0.72 | [0.01; 1.44] | 25.0% | 83.7 | [63.1; 92.8] | 3.6% | 4.55 |
| CBT | Care As Usual | - At Least 2 Flags Removed | 4 | 33.3% | 0.7 | [-0.3; 1.7] | 27.1% | 87.4 | [69.9; 94.7] | -0.7% | 4.72 |
| CBT | Care As Usual | - Flag 1 Removed (Effect) | 5 | 16.7% | 0.72 | [0.01; 1.44] | 25.0% | 83.7 | [63.1; 92.8] | 3.6% | 4.55 |
| CBT | Care As Usual | - Flag 2 Removed (Power) | 4 | 33.3% | 0.7 | [-0.3; 1.7] | 27.1% | 87.4 | [69.9; 94.7] | -0.7% | 4.72 |
| CR | Care As Usual | All Effect Sizes Included | 1 | 0.0% | 0.13 | [-0.92; 1.18] | 0.0% | | - | | 31.76 |
| CR | Care As Usual | - 3 Flags Removed | 1 | 0.0% | 0.13 | [-0.92; 1.18] | 0.0% | | - | | 31.76 |
| CR | Care As Usual | - Flag 1 Removed (Effect) | 1 | 0.0% | 0.13 | [-0.92; 1.18] | 0.0% | | - | | 31.76 |
| Other | Care As Usual | All Effect Sizes Included | 2 | 0.0% | 1.56 | [-17.95; 21.06] | 0.0% | 95.2 | [85.7; 98.4] | 0.0% | 1.84 |
| Other | Care As Usual | - 3 Flags Removed | 1 | 50.0% | 0.07 | [-0.43; 0.58] | 95.5% | | - | | 59.62 |
| Other | Care As Usual | - At Least 2 Flags Removed | 1 | 50.0% | 0.07 | [-0.43; 0.58] | 95.5% | | - | | 59.62 |
| Other | Care As Usual | - Flag 1 Removed (Effect) | 1 | 50.0% | 0.07 | [-0.43; 0.58] | 95.5% | | - | | 59.62 |
| Other | Care As Usual | - Flag 2 Removed (Power) | 1 | 50.0% | 0.07 | [-0.43; 0.58] | 95.5% | | - | | 59.62 |
| PE | Care As Usual | All Effect Sizes Included | 1 | 0.0% | 0.62 | [0.29; 0.96] | 0.0% | | - | | 5.44 |
| PE | Care As Usual | - 3 Flags Removed | 1 | 0.0% | 0.62 | [0.29; 0.96] | 0.0% | | - | | 5.44 |
| PE | Care As Usual | - At Least 2 Flags Removed | 1 | 0.0% | 0.62 | [0.29; 0.96] | 0.0% | | - | | 5.44 |
| PE | Care As Usual | - Flag 1 Removed (Effect) | 1 | 0.0% | 0.62 | [0.29; 0.96] | 0.0% | | - | | 5.44 |
| PE | Care As Usual | - Flag 2 Removed (Power) | 1 | 0.0% | 0.62 | [0.29; 0.96] | 0.0% | | - | | 5.44 |
| 3rd Wave | Waitlist | All Effect Sizes Included | 5 | 0.0% | 1.38 | [0.55; 2.21] | 0.0% | 83.5 | [62.6; 92.7] | 0.0% | 2.09 |
| 3rd Wave | Waitlist | - 3 Flags Removed | 4 | 20.0% | 1.13 | [0.27; 1.99] | 18.1% | 70.3 | [14.9; 89.7] | 15.8% | 2.64 |
| 3rd Wave | Waitlist | - At Least 2 Flags Removed | 3 | 40.0% | 1.01 | [-0.33; 2.34] | 26.8% | 72.4 | [6.9; 91.8] | 13.3% | 3.02 |
| 3rd Wave | Waitlist | - Flag 1 Removed (Effect) | 4 | 20.0% | 1.13 | [0.27; 1.99] | 18.1% | 70.3 | [14.9; 89.7] | 15.8% | 2.64 |
| 3rd Wave | Waitlist | - Flag 2 Removed (Power) | 2 | 60.0% | 0.74 | [0.14; 1.33] | 46.4% | 0 | - | 100.0% | 4.43 |
| 3rd Wave | Waitlist | - Flag 3 Removed (RoB) | 1 | 80.0% | 1.7 | [1.06; 2.34] | -23.2% | | - | | 1.69 |
| AR | Waitlist | All Effect Sizes Included | 5 | 0.0% | 0.77 | [0.31; 1.23] | 0.0% | 24.2 | [0; 69.2] | 0.0% | 4.18 |
| AR | Waitlist | - 3 Flags Removed | 5 | 0.0% | 0.77 | [0.31; 1.23] | 0.0% | 24.2 | [0; 69.2] | 0.0% | 4.18 |
| AR | Waitlist | - At Least 2 Flags Removed | 1 | 80.0% | 1.22 | [0.72; 1.73] | -58.4% | | - | | 2.4 |
| AR | Waitlist | - Flag 1 Removed (Effect) | 5 | 0.0% | 0.77 | [0.31; 1.23] | 0.0% | 24.2 | [0; 69.2] | 0.0% | 4.18 |
| AR | Waitlist | - Flag 2 Removed (Power) | 1 | 80.0% | 1.22 | [0.72; 1.73] | -58.4% | | - | | 2.4 |
| BT | Waitlist | All Effect Sizes Included | 3 | 0.0% | 0.37 | [-0.45; 1.19] | 0.0% | 6.1 | [0; 90.2] | 0.0% | 10.1 |
| BT | Waitlist | - 3 Flags Removed | 3 | 0.0% | 0.37 | [-0.45; 1.19] | 0.0% | 6.1 | [0; 90.2] | 0.0% | 10.1 |
| BT | Waitlist | - At Least 2 Flags Removed | 1 | 66.7% | 0.63 | [0.16; 1.11] | -70.3% | | - | | 5.32 |
| BT | Waitlist | - Flag 1 Removed (Effect) | 3 | 0.0% | 0.37 | [-0.45; 1.19] | 0.0% | 6.1 | [0; 90.2] | 0.0% | 10.1 |
| BT | Waitlist | - Flag 2 Removed (Power) | 1 | 66.7% | 0.63 | [0.16; 1.11] | -70.3% | | - | | 5.32 |

| Intervention | Control | Analysis | *k* | $\Delta_k$ | SMD | 95%-CI | $\Delta_{SMD}$ | $I^2$ | 95%-CI | $\Delta_{het}$ | NNT |
|---|---|---|---|---|---|---|---|---|---|---|---|
| CBT | Waitlist | All Effect Sizes Included | 27 | 0.0% | 0.87 | [0.67; 1.06] | 0.0% | 68.4 | [53.1; 78.8] | 0.0% | 3.64 |
| CBT | Waitlist | - 3 Flags Removed | 26 | 3.7% | 0.82 | [0.64; 1] | 5.7% | 62.1 | [42; 75.2] | 9.2% | 3.89 |
| CBT | Waitlist | - At Least 2 Flags Removed | 15 | 44.4% | 0.75 | [0.53; 0.98] | 13.8% | 67.5 | [44.3; 81] | 1.3% | 4.31 |
| CBT | Waitlist | - Flag 1 Removed (Effect) | 26 | 3.7% | 0.82 | [0.64; 1] | 5.7% | 62.1 | [42; 75.2] | 9.2% | 3.89 |
| CBT | Waitlist | - Flag 2 Removed (Power) | 12 | 55.6% | 0.74 | [0.5; 0.98] | 14.9% | 68.7 | [43; 82.8] | -0.4% | 4.39 |
| CBT | Waitlist | - Flag 3 Removed (RoB) | 8 | 70.4% | 0.75 | [0.41; 1.09] | 13.8% | 71.8 | [42.1; 86.3] | -5.0% | 4.34 |
| CT | Waitlist | All Effect Sizes Included | 1 | 0.0% | 1.17 | [0.38; 1.96] | 0.0% | | - | | 2.53 |
| CT | Waitlist | - 3 Flags Removed | 1 | 0.0% | 1.02 | [0.22; 1.82] | 12.8% | | - | | 2.97 |
| CT | Waitlist | - Flag 1 Removed (Effect) | 1 | 0.0% | 1.02 | [0.22; 1.82] | 12.8% | | - | | 2.97 |
| Psychodynamic | Waitlist | All Effect Sizes Included | 1 | 0.0% | 0.18 | [-0.29; 0.64] | 0.0% | | - | | 23.37 |
| Psychodynamic | Waitlist | - 3 Flags Removed | 1 | 0.0% | 0.18 | [-0.29; 0.64] | 0.0% | | - | | 23.37 |
| Psychodynamic | Waitlist | - At Least 2 Flags Removed | 1 | 0.0% | 0.18 | [-0.29; 0.64] | 0.0% | | - | | 23.37 |
| Psychodynamic | Waitlist | - Flag 1 Removed (Effect) | 1 | 0.0% | 0.18 | [-0.29; 0.64] | 0.0% | | - | | 23.37 |
| Psychodynamic | Waitlist | - Flag 2 Removed (Power) | 1 | 0.0% | 0.18 | [-0.29; 0.64] | 0.0% | | - | | 23.37 |
| Other | Waitlist | All Effect Sizes Included | 1 | 0.0% | 0.76 | [-0.02; 1.54] | 0.0% | | - | | 4.28 |
| Other | Waitlist | - 3 Flags Removed | 1 | 0.0% | 0.76 | [-0.02; 1.54] | 0.0% | | - | | 4.28 |
| Other | Waitlist | - Flag 1 Removed (Effect) | 1 | 0.0% | 0.76 | [-0.02; 1.54] | 0.0% | | - | | 4.28 |
| SUP | Waitlist | All Effect Sizes Included | 1 | 0.0% | 0.49 | [-0.09; 1.06] | 0.0% | | - | | 7.35 |
| SUP | Waitlist | - 3 Flags Removed | 1 | 0.0% | 0.49 | [-0.09; 1.06] | 0.0% | | - | | 7.35 |
| SUP | Waitlist | - Flag 1 Removed (Effect) | 1 | 0.0% | 0.49 | [-0.09; 1.06] | 0.0% | | - | | 7.35 |
| ***Complicated Grief*** | | | | | | | | | | | |
| CBT | Care As Usual | All Effect Sizes Included | 2 | 0.0% | 0.11 | [-2.89; 3.11] | 0.0% | 77.1 | [0; 94.8] | 0.0% | 23.14 |
| CBT | Care As Usual | - 3 Flags Removed | 2 | 0.0% | 0.11 | [-2.89; 3.11] | 0.0% | 77.1 | [0; 94.8] | 0.0% | 23.14 |
| CBT | Care As Usual | - At Least 2 Flags Removed | 2 | 0.0% | 0.11 | [-2.89; 3.11] | 0.0% | 77.1 | [0; 94.8] | 0.0% | 23.14 |
| CBT | Care As Usual | - Flag 1 Removed (Effect) | 2 | 0.0% | 0.11 | [-2.89; 3.11] | 0.0% | 77.1 | [0; 94.8] | 0.0% | 23.14 |
| CBT | Care As Usual | - Flag 2 Removed (Power) | 2 | 0.0% | 0.11 | [-2.89; 3.11] | 0.0% | 77.1 | [0; 94.8] | 0.0% | 23.14 |
| Other | Care As Usual | All Effect Sizes Included | 5 | 0.0% | 0.08 | [-0.2; 0.37] | 0.0% | 0 | [0; 79.2] | | 30.48 |
| Other | Care As Usual | - 3 Flags Removed | 5 | 0.0% | 0.08 | [-0.2; 0.37] | 0.0% | 0 | [0; 79.2] | | 30.48 |
| Other | Care As Usual | - At Least 2 Flags Removed | 4 | 20.0% | 0.08 | [-0.33; 0.49] | 0.0% | 22.5 | [0; 88.1] | -Inf | 30.81 |
| Other | Care As Usual | - Flag 1 Removed (Effect) | 5 | 0.0% | 0.08 | [-0.2; 0.37] | 0.0% | 0 | [0; 79.2] | | 30.48 |
| Other | Care As Usual | - Flag 2 Removed (Power) | 3 | 40.0% | 0.07 | [-0.67; 0.8] | 12.5% | 47.3 | [0; 84.5] | -Inf | 37.93 |
| CBT | Waitlist | All Effect Sizes Included | 6 | 0.0% | 0.85 | [-0.08; 1.78] | 0.0% | 87.1 | [74.1; 93.5] | 0.0% | 3.32 |
| CBT | Waitlist | - 3 Flags Removed | 6 | 0.0% | 0.85 | [-0.08; 1.78] | 0.0% | 87.1 | [74.1; 93.5] | 0.0% | 3.32 |
| CBT | Waitlist | - At Least 2 Flags Removed | 2 | 66.7% | 0.44 | [-5.39; 6.27] | 48.2% | 89.3 | [59.9; 97.1] | -2.5% | 5.87 |
| CBT | Waitlist | - Flag 1 Removed (Effect) | 5 | 16.7% | 0.56 | [-0.14; 1.27] | 34.1% | 78.8 | [49.3; 91.1] | 9.5% | 4.68 |

| Intervention | Control | Analysis | *k* | $\Delta_k$ | SMD | 95%-CI | $\Delta_{SMD}$ | $I^2$ | 95%-CI | $\Delta_{het}$ | NNT |
|---|---|---|---|---|---|---|---|---|---|---|---|
| CBT | Waitlist | - Flag 2 Removed (Power) | 2 | 66.7% | 0.44 | [-5.39; 6.27] | 48.2% | 89.3 | [59.9; 97.1] | -2.5% | 5.87 |
| CBT | Waitlist | - Flag 3 Removed (RoB) | 1 | 83.3% | 2.41 | [1.57; 3.24] | -183.5% | | - | | 2.03 |
| Other | Waitlist | All Effect Sizes Included | 4 | 0.0% | 0.59 | [-0.86; 2.03] | 0.0% | 87 | [68.9; 94.6] | 0.0% | 4.5 |
| Other | Waitlist | - 3 Flags Removed | 3 | 25.0% | 0.09 | [-0.23; 0.42] | 84.7% | 0 | [0; 89.6] | 100.0% | 26.59 |
| Other | Waitlist | - At Least 2 Flags Removed | 2 | 50.0% | 0.1 | [-1.53; 1.74] | 83.1% | 0 | - | 100.0% | 24.52 |
| Other | Waitlist | - Flag 1 Removed (Effect) | 3 | 25.0% | 0.09 | [-0.23; 0.42] | 84.7% | 0 | [0; 89.6] | 100.0% | 26.59 |
| Other | Waitlist | - Flag 2 Removed (Power) | 1 | 75.0% | 0.03 | [-0.48; 0.53] | 94.9% | | - | | 85.12 |
| Other | Waitlist | - Flag 3 Removed (RoB) | 1 | 75.0% | 0.33 | [-0.56; 1.22] | 44.1% | | - | | 7.74 |
| Writ | Waitlist | All Effect Sizes Included | 5 | 0.0% | 0.59 | [-0.1; 1.27] | 0.0% | 80.9 | [55.3; 91.8] | 0.0% | 4.52 |
| Writ | Waitlist | - 3 Flags Removed | 5 | 0.0% | 0.59 | [-0.1; 1.27] | 0.0% | 80.9 | [55.3; 91.8] | 0.0% | 4.52 |
| Writ | Waitlist | - At Least 2 Flags Removed | 5 | 0.0% | 0.59 | [-0.1; 1.27] | 0.0% | 80.9 | [55.3; 91.8] | 0.0% | 4.52 |
| Writ | Waitlist | - At Least 1 Flag Removed | 4 | 20.0% | 0.54 | [-0.46; 1.54] | 8.5% | 82.8 | [56.1; 93.3] | -2.3% | 4.87 |
| Writ | Waitlist | - Flag 1 Removed (Effect) | 5 | 0.0% | 0.59 | [-0.1; 1.27] | 0.0% | 80.9 | [55.3; 91.8] | 0.0% | 4.52 |
| Writ | Waitlist | - Flag 2 Removed (Power) | 5 | 0.0% | 0.59 | [-0.1; 1.27] | 0.0% | 80.9 | [55.3; 91.8] | 0.0% | 4.52 |
| Writ | Waitlist | - Flag 3 Removed (RoB) | 4 | 20.0% | 0.54 | [-0.46; 1.54] | 8.5% | 82.8 | [56.1; 93.3] | -2.3% | 4.87 |
| ***Obsessive-Compulsive Disorder*** | | | | | | | | | | | |
| CBT | Care As Usual | All Effect Sizes Included | 2 | 0.0% | 1.35 | [-3.8; 6.49] | 0.0% | 75.3 | [0; 94.4] | 0.0% | 3 |
| CBT | Care As Usual | - 3 Flags Removed | 2 | 0.0% | 1.35 | [-3.8; 6.49] | 0.0% | 75.3 | [0; 94.4] | 0.0% | 3 |
| CBT | Care As Usual | - Flag 1 Removed (Effect) | 2 | 0.0% | 1.35 | [-3.8; 6.49] | 0.0% | 75.3 | [0; 94.4] | 0.0% | 3 |
| ERP | Care As Usual | All Effect Sizes Included | 2 | 0.0% | 1.08 | [-4.92; 7.07] | 0.0% | 52.5 | [0; 88.1] | 0.0% | 4.26 |
| ERP | Care As Usual | - 3 Flags Removed | 2 | 0.0% | 1.08 | [-4.92; 7.07] | 0.0% | 52.5 | [0; 88.1] | 0.0% | 4.26 |
| ERP | Care As Usual | - Flag 1 Removed (Effect) | 2 | 0.0% | 1.08 | [-4.92; 7.07] | 0.0% | 52.5 | [0; 88.1] | 0.0% | 4.26 |
| CBT | Waitlist | All Effect Sizes Included | 9 | 0.0% | 1.11 | [0.95; 1.27] | 0.0% | 0 | [0; 64.8] | | 4.08 |
| CBT | Waitlist | - 3 Flags Removed | 9 | 0.0% | 1.11 | [0.95; 1.27] | 0.0% | 0 | [0; 64.8] | | 4.08 |
| CBT | Waitlist | - Flag 1 Removed (Effect) | 9 | 0.0% | 1.11 | [0.95; 1.27] | 0.0% | 0 | [0; 64.8] | | 4.08 |
| CBT | Waitlist | - Flag 2 Removed (Power) | 2 | 77.8% | 1.3 | [0.31; 2.28] | -17.1% | 0 | - | | 3.18 |
| CT | Waitlist | All Effect Sizes Included | 3 | 0.0% | 1.22 | [-0.81; 3.26] | 0.0% | 78.6 | [31.2; 93.3] | 0.0% | 3.48 |
| CT | Waitlist | - 3 Flags Removed | 3 | 0.0% | 1.22 | [-0.81; 3.26] | 0.0% | 78.6 | [31.2; 93.3] | 0.0% | 3.48 |
| CT | Waitlist | - Flag 1 Removed (Effect) | 3 | 0.0% | 1.22 | [-0.81; 3.26] | 0.0% | 78.6 | [31.2; 93.3] | 0.0% | 3.48 |
| ERP | Waitlist | All Effect Sizes Included | 5 | 0.0% | 1.52 | [0.38; 2.66] | 0.0% | 81.5 | [57.1; 92] | 0.0% | 2.49 |
| ERP | Waitlist | - 3 Flags Removed | 4 | 20.0% | 1.09 | [0.26; 1.92] | 28.3% | 49.1 | [0; 83.2] | 39.8% | 4.17 |
| ERP | Waitlist | - Flag 1 Removed (Effect) | 4 | 20.0% | 1.13 | [0.24; 2.02] | 25.7% | 56.1 | [0; 85.4] | 31.2% | 3.96 |
| Other | Waitlist | All Effect Sizes Included | 2 | 0.0% | 1.8 | [-13.47; 17.07] | 0.0% | 94.4 | [82.5; 98.2] | 0.0% | 1.95 |
| Other | Waitlist | - 3 Flags Removed | 1 | 50.0% | 0.62 | [-0.01; 1.26] | 65.6% | | - | | 9.68 |
| Other | Waitlist | - Flag 1 Removed (Effect) | 1 | 50.0% | 0.62 | [-0.01; 1.26] | 65.6% | | - | | 9.68 |

| Intervention | Control | Analysis | *k* | $\Delta_k$ | SMD | 95%-CI | $\Delta_{SMD}$ | $I^2$ | 95%-CI | $\Delta_{het}$ | NNT |
|---|---|---|---|---|---|---|---|---|---|---|---|
| ***Panic Disorder*** | | | | | | | | | | | |
| BT | Care As Usual | All Effect Sizes Included | 2 | 0.0% | 0.58 | [0.16; 1] | 0.0% | 0 | - | | 5.93 |
| BT | Care As Usual | - 3 Flags Removed | 2 | 0.0% | 0.58 | [0.16; 1] | 0.0% | 0 | - | | 5.93 |
| BT | Care As Usual | - At Least 2 Flags Removed | 1 | 50.0% | 0.61 | [0.08; 1.13] | -5.2% | | - | | 5.64 |
| BT | Care As Usual | - Flag 1 Removed (Effect) | 2 | 0.0% | 0.58 | [0.16; 1] | 0.0% | 0 | - | | 5.93 |
| BT | Care As Usual | - Flag 2 Removed (Power) | 1 | 50.0% | 0.61 | [0.08; 1.13] | -5.2% | | - | | 5.64 |
| CBT | Care As Usual | All Effect Sizes Included | 11 | 0.0% | 0.7 | [0.23; 1.17] | 0.0% | 80.4 | [65.9; 88.8] | 0.0% | 4.7 |
| CBT | Care As Usual | - 3 Flags Removed | 10 | 9.1% | 0.5 | [0.17; 0.82] | 28.6% | 68.1 | [38.2; 83.5] | 15.3% | 7.17 |
| CBT | Care As Usual | - At Least 2 Flags Removed | 7 | 36.4% | 0.31 | [0.14; 0.48] | 55.7% | 32.4 | [0; 71.3] | 59.7% | 12.43 |
| CBT | Care As Usual | - At Least 1 Flag Removed | 3 | 72.7% | 0.2 | [0; 0.39] | 71.4% | 0 | [0; 89.6] | 100.0% | 20.45 |
| CBT | Care As Usual | - Flag 1 Removed (Effect) | 10 | 9.1% | 0.5 | [0.17; 0.82] | 28.6% | 68.1 | [38.2; 83.5] | 15.3% | 7.17 |
| CBT | Care As Usual | - Flag 2 Removed (Power) | 7 | 36.4% | 0.31 | [0.14; 0.48] | 55.7% | 32.4 | [0; 71.3] | 59.7% | 12.43 |
| CBT | Care As Usual | - Flag 3 Removed (RoB) | 3 | 72.7% | 0.2 | [0; 0.39] | 71.4% | 0 | [0; 89.6] | 100.0% | 20.45 |
| CT | Care As Usual | All Effect Sizes Included | 2 | 0.0% | 0.38 | [-0.51; 1.28] | 0.0% | 0 | - | | 9.68 |
| CT | Care As Usual | - 3 Flags Removed | 2 | 0.0% | 0.38 | [-0.51; 1.28] | 0.0% | 0 | - | | 9.68 |
| CT | Care As Usual | - At Least 2 Flags Removed | 1 | 50.0% | 0.33 | [-0.18; 0.84] | 13.2% | | - | | 11.6 |
| CT | Care As Usual | - Flag 1 Removed (Effect) | 2 | 0.0% | 0.38 | [-0.51; 1.28] | 0.0% | 0 | - | | 9.68 |
| CT | Care As Usual | - Flag 2 Removed (Power) | 1 | 50.0% | 0.33 | [-0.18; 0.84] | 13.2% | | - | | 11.6 |
| PD | Care As Usual | All Effect Sizes Included | 1 | 0.0% | 1.32 | [0.63; 2] | 0.0% | | - | | 2.2 |
| PD | Care As Usual | - 3 Flags Removed | 1 | 0.0% | 1.32 | [0.63; 2] | 0.0% | | - | | 2.2 |
| PD | Care As Usual | - Flag 1 Removed (Effect) | 1 | 0.0% | 1.32 | [0.63; 2] | 0.0% | | - | | 2.2 |
| PT | Care As Usual | All Effect Sizes Included | 1 | 0.0% | 0.38 | [-0.24; 1] | 0.0% | | - | | 9.88 |
| PT | Care As Usual | - 3 Flags Removed | 1 | 0.0% | 0.38 | [-0.24; 1] | 0.0% | | - | | 9.88 |
| PT | Care As Usual | - Flag 1 Removed (Effect) | 1 | 0.0% | 0.38 | [-0.24; 1] | 0.0% | | - | | 9.88 |
| 3W | Waitlist | All Effect Sizes Included | 2 | 0.0% | 0.57 | [-0.36; 1.51] | 0.0% | 0 | - | | 6.03 |
| 3W | Waitlist | - 3 Flags Removed | 2 | 0.0% | 0.57 | [-0.36; 1.51] | 0.0% | 0 | - | | 6.03 |
| 3W | Waitlist | - Flag 1 Removed (Effect) | 2 | 0.0% | 0.57 | [-0.36; 1.51] | 0.0% | 0 | - | | 6.03 |
| BT | Waitlist | All Effect Sizes Included | 10 | 0.0% | 1.41 | [0.96; 1.85] | 0.0% | 57 | [13.1; 78.8] | 0.0% | 2.05 |
| BT | Waitlist | - 3 Flags Removed | 8 | 20.0% | 1.2 | [0.77; 1.63] | 14.9% | 37.4 | [0; 72.3] | 34.4% | 2.46 |
| BT | Waitlist | - Flag 1 Removed (Effect) | 8 | 20.0% | 1.2 | [0.77; 1.63] | 14.9% | 37.4 | [0; 72.3] | 34.4% | 2.46 |
| CBT | Waitlist | All Effect Sizes Included | 37 | 0.0% | 1.03 | [0.81; 1.24] | 0.0% | 70.7 | [59.1; 79] | 0.0% | 2.96 |
| CBT | Waitlist | - 3 Flags Removed | 34 | 8.1% | 0.94 | [0.75; 1.13] | 8.7% | 65.8 | [50.9; 76.1] | 6.9% | 3.29 |
| CBT | Waitlist | - At Least 2 Flags Removed | 13 | 64.9% | 1.02 | [0.75; 1.29] | 1.0% | 73.5 | [54; 84.7] | -4.0% | 2.97 |
| CBT | Waitlist | - At Least 1 Flag Removed | 3 | 91.9% | 0.89 | [0.32; 1.46] | 13.6% | 58.7 | [0; 88.2] | 17.0% | 3.52 |

| Intervention | Control | Analysis | *k* | $\Delta_k$ | SMD | 95%-CI | $\Delta_{SMD}$ | $I^2$ | 95%-CI | $\Delta_{het}$ | NNT |
|---|---|---|---|---|---|---|---|---|---|---|---|
| CBT | Waitlist | - Flag 1 Removed (Effect) | 34 | 8.1% | 0.94 | [0.75; 1.13] | 8.7% | 65.8 | [50.9; 76.1] | 6.9% | 3.29 |
| CBT | Waitlist | - Flag 2 Removed (Power) | 13 | 64.9% | 1.02 | [0.75; 1.29] | 1.0% | 73.5 | [54; 84.7] | -4.0% | 2.97 |
| CBT | Waitlist | - Flag 3 Removed (RoB) | 3 | 91.9% | 0.89 | [0.32; 1.46] | 13.6% | 58.7 | [0; 88.2] | 17.0% | 3.52 |
| CT | Waitlist | All Effect Sizes Included | 1 | 0.0% | 0.28 | [-0.57; 1.14] | 0.0% | | - | | 13.7 |
| CT | Waitlist | - 3 Flags Removed | 1 | 0.0% | 0.28 | [-0.57; 1.14] | 0.0% | | - | | 13.7 |
| CT | Waitlist | - Flag 1 Removed (Effect) | 1 | 0.0% | 0.28 | [-0.57; 1.14] | 0.0% | | - | | 13.7 |
| EMDR | Waitlist | All Effect Sizes Included | 2 | 0.0% | 0.57 | [-1.08; 2.23] | 0.0% | 0 | - | | 6.05 |
| EMDR | Waitlist | - 3 Flags Removed | 2 | 0.0% | 0.57 | [-1.08; 2.23] | 0.0% | 0 | - | | 6.05 |
| EMDR | Waitlist | - Flag 1 Removed (Effect) | 2 | 0.0% | 0.57 | [-1.08; 2.23] | 0.0% | 0 | - | | 6.05 |
| PT | Waitlist | All Effect Sizes Included | 6 | 0.0% | 0.89 | [0.2; 1.58] | 0.0% | 64.6 | [14.7; 85.3] | 0.0% | 3.51 |
| PT | Waitlist | - 3 Flags Removed | 5 | 16.7% | 0.66 | [0.49; 0.82] | 25.8% | 0 | [0; 79.2] | 100.0% | 5.09 |
| PT | Waitlist | - At Least 2 Flags Removed | 1 | 83.3% | 0.76 | [0.21; 1.31] | 14.6% | | - | | 4.28 |
| PT | Waitlist | - Flag 1 Removed (Effect) | 5 | 16.7% | 0.66 | [0.49; 0.82] | 25.8% | 0 | [0; 79.2] | 100.0% | 5.09 |
| PT | Waitlist | - Flag 2 Removed (Power) | 1 | 83.3% | 0.76 | [0.21; 1.31] | 14.6% | | - | | 4.28 |
| ***Phobia*** | | | | | | | | | | | |
| CBT | Care As Usual | All Effect Sizes Included | 3 | 0.0% | 1.57 | [0.31; 2.83] | 0.0% | 56 | [0; 87.5] | 0.0% | 2 |
| CBT | Care As Usual | - 3 Flags Removed | 3 | 0.0% | 1.57 | [0.31; 2.83] | 0.0% | 56 | [0; 87.5] | 0.0% | 2 |
| CBT | Care As Usual | - Flag 1 Removed (Effect) | 2 | 33.3% | 1.2 | [0.57; 1.84] | 23.6% | 0 | - | 100.0% | 2.81 |
| Cognitive | Care As Usual | All Effect Sizes Included | 1 | 0.0% | 1.25 | [0.45; 2.05] | 0.0% | | - | | 2.67 |
| Cognitive | Care As Usual | - 3 Flags Removed | 1 | 0.0% | 1.25 | [0.45; 2.05] | 0.0% | | - | | 2.67 |
| Cognitive | Care As Usual | - Flag 1 Removed (Effect) | 1 | 0.0% | 1.25 | [0.45; 2.05] | 0.0% | | - | | 2.67 |
| Exposure | Care As Usual | All Effect Sizes Included | 5 | 0.0% | 1.47 | [0.12; 2.81] | 0.0% | 80.8 | [55.2; 91.8] | 0.0% | 2.17 |
| Exposure | Care As Usual | - 3 Flags Removed | 3 | 40.0% | 0.8 | [-0.73; 2.34] | 45.6% | 62.3 | [0; 89.2] | 22.9% | 4.89 |
| Exposure | Care As Usual | - Flag 1 Removed (Effect) | 3 | 40.0% | 0.8 | [-0.73; 2.34] | 45.6% | 62.3 | [0; 89.2] | 22.9% | 4.89 |
| Other | Care As Usual | All Effect Sizes Included | 2 | 0.0% | -0.06 | [-13.82; 13.7] | 0.0% | 88 | [53.7; 96.9] | 0.0% | 102.93 |
| Other | Care As Usual | - 3 Flags Removed | 2 | 0.0% | -0.06 | [-13.82; 13.7] | 0.0% | 88 | [53.7; 96.9] | 0.0% | 102.93 |
| Other | Care As Usual | - Flag 1 Removed (Effect) | 2 | 0.0% | -0.06 | [-13.82; 13.7] | 0.0% | 88 | [53.7; 96.9] | 0.0% | 102.93 |
| CBT | Waitlist | All Effect Sizes Included | 6 | 0.0% | 1.14 | [0.03; 2.25] | 0.0% | 90 | [80.8; 94.7] | 0.0% | 3.03 |
| CBT | Waitlist | - 3 Flags Removed | 5 | 16.7% | 0.65 | [0.25; 1.04] | 43.0% | 0 | [0; 79.2] | 100.0% | 6.5 |
| CBT | Waitlist | - At Least 2 Flags Removed | 2 | 66.7% | 0.52 | [-0.14; 1.19] | 54.4% | 0 | - | 100.0% | 8.58 |
| CBT | Waitlist | - Flag 1 Removed (Effect) | 5 | 16.7% | 0.65 | [0.25; 1.04] | 43.0% | 0 | [0; 79.2] | 100.0% | 6.5 |
| CBT | Waitlist | - Flag 2 Removed (Power) | 2 | 66.7% | 0.52 | [-0.14; 1.19] | 54.4% | 0 | - | 100.0% | 8.58 |
| Cognitive | Waitlist | All Effect Sizes Included | 1 | 0.0% | 0.14 | [-0.66; 0.94] | 0.0% | | - | | 41.23 |
| Cognitive | Waitlist | - 3 Flags Removed | 1 | 0.0% | 0.14 | [-0.66; 0.94] | 0.0% | | - | | 41.23 |
| Cognitive | Waitlist | - Flag 1 Removed (Effect) | 1 | 0.0% | 0.14 | [-0.66; 0.94] | 0.0% | | - | | 41.23 |

| Intervention | Control | Analysis | $k$ | $\Delta_k$ | SMD | 95%-CI | $\Delta_{SMD}$ | $I^2$ | 95%-CI | $\Delta_{het}$ | NNT |
|---|---|---|---|---|---|---|---|---|---|---|---|
| EMDR | Waitlist | All Effect Sizes Included | 2 | 0.0% | 1.22 | [-3.34; 5.78] | 0.0% | 17.9 | - | 0.0% | 2.77 |
| EMDR | Waitlist | - 3 Flags Removed | 2 | 0.0% | 1.22 | [-3.34; 5.78] | 0.0% | 17.9 | - | 0.0% | 2.77 |
| EMDR | Waitlist | - Flag 1 Removed (Effect) | 2 | 0.0% | 1.22 | [-3.34; 5.78] | 0.0% | 17.9 | - | 0.0% | 2.77 |
| Exposure | Waitlist | All Effect Sizes Included | 35 | 0.0% | 1.28 | [1.01; 1.54] | 0.0% | 74.3 | [64.2; 81.5] | 0.0% | 2.59 |
| Exposure | Waitlist | - 3 Flags Removed | 27 | 22.9% | 1 | [0.78; 1.21] | 21.9% | 62.1 | [42.5; 75] | 16.4% | 3.64 |
| Exposure | Waitlist | - At Least 2 Flags Removed | 8 | 77.1% | 0.88 | [0.56; 1.2] | 31.2% | 61.9 | [17.7; 82.4] | 16.7% | 4.29 |
| Exposure | Waitlist | - Flag 1 Removed (Effect) | 26 | 25.7% | 0.93 | [0.74; 1.12] | 27.3% | 48 | [17.8; 67.1] | 35.4% | 3.99 |
| Exposure | Waitlist | - Flag 2 Removed (Power) | 7 | 80.0% | 1.07 | [0.51; 1.64] | 16.4% | 84.2 | [69.2; 91.9] | -13.3% | 3.28 |
| Other | Waitlist | All Effect Sizes Included | 5 | 0.0% | 1.32 | [0.89; 1.74] | 0.0% | 0 | [0; 79.2] | | 2.5 |
| Other | Waitlist | - 3 Flags Removed | 5 | 0.0% | 1.32 | [0.89; 1.74] | 0.0% | 0 | [0; 79.2] | | 2.5 |
| Other | Waitlist | - At Least 2 Flags Removed | 1 | 80.0% | 1.32 | [0.85; 1.79] | 0.0% | | - | | 2.49 |
| Other | Waitlist | - Flag 1 Removed (Effect) | 5 | 0.0% | 1.32 | [0.89; 1.74] | 0.0% | 0 | [0; 79.2] | | 2.5 |
| Other | Waitlist | - Flag 2 Removed (Power) | 1 | 80.0% | 1.32 | [0.85; 1.79] | 0.0% | | - | | 2.49 |
| Relaxation | Waitlist | All Effect Sizes Included | 1 | 0.0% | 0.74 | [0.19; 1.28] | 0.0% | | - | | 5.46 |
| Relaxation | Waitlist | - 3 Flags Removed | 1 | 0.0% | 0.74 | [0.19; 1.28] | 0.0% | | - | | 5.46 |
| Relaxation | Waitlist | - At Least 2 Flags Removed | 1 | 0.0% | 0.74 | [0.19; 1.28] | 0.0% | | - | | 5.46 |
| Relaxation | Waitlist | - Flag 1 Removed (Effect) | 1 | 0.0% | 0.74 | [0.19; 1.28] | 0.0% | | - | | 5.46 |
| Relaxation | Waitlist | - Flag 2 Removed (Power) | 1 | 0.0% | 0.74 | [0.19; 1.28] | 0.0% | | - | | 5.46 |
| ***Psychosis*** | | | | | | | | | | | |
| ALL | Care As Usual | All Effect Sizes Included | 1 | 0.0% | -0.29 | [-0.44; -0.15] | 0.0% | | - | | 16.31 |
| ALL | Care As Usual | - 3 Flags Removed | 1 | 0.0% | -0.29 | [-0.44; -0.15] | 0.0% | | - | | 16.31 |
| ALL | Care As Usual | - At Least 2 Flags Removed | 1 | 0.0% | -0.29 | [-0.44; -0.15] | 0.0% | | - | | 16.31 |
| ALL | Care As Usual | - Flag 1 Removed (Effect) | 1 | 0.0% | -0.29 | [-0.44; -0.15] | 0.0% | | - | | 16.31 |
| ALL | Care As Usual | - Flag 2 Removed (Power) | 1 | 0.0% | -0.29 | [-0.44; -0.15] | 0.0% | | - | | 16.31 |
| CBT | Care As Usual | All Effect Sizes Included | 23 | 0.0% | 0.29 | [0.15; 0.42] | 0.0% | 49 | [17.3; 68.5] | 0.0% | 16.65 |
| CBT | Care As Usual | - 3 Flags Removed | 22 | 4.3% | 0.25 | [0.15; 0.35] | 13.8% | 22.4 | [0; 53.9] | 54.3% | 19.51 |
| CBT | Care As Usual | - At Least 2 Flags Removed | 15 | 34.8% | 0.23 | [0.11; 0.35] | 20.7% | 27.5 | [0; 61] | 43.9% | 21.48 |
| CBT | Care As Usual | - At Least 1 Flag Removed | 4 | 82.6% | 0.36 | [-0.15; 0.88] | -24.1% | 73.6 | [26; 90.6] | -50.2% | 12.59 |
| CBT | Care As Usual | - Flag 1 Removed (Effect) | 22 | 4.3% | 0.25 | [0.15; 0.35] | 13.8% | 22.4 | [0; 53.9] | 54.3% | 19.51 |
| CBT | Care As Usual | - Flag 2 Removed (Power) | 10 | 56.5% | 0.25 | [0.08; 0.42] | 13.8% | 51.2 | [0; 76.3] | -4.5% | 19.72 |
| CBT | Care As Usual | - Flag 3 Removed (RoB) | 9 | 60.9% | 0.27 | [0.08; 0.47] | 6.9% | 39.6 | [0; 72.2] | 19.2% | 17.62 |
| CR | Care As Usual | All Effect Sizes Included | 23 | 0.0% | 0.3 | [0.14; 0.47] | 0.0% | 53.6 | [25.5; 71.1] | 0.0% | 15.61 |
| CR | Care As Usual | - 3 Flags Removed | 23 | 0.0% | 0.29 | [0.13; 0.46] | 3.3% | 50.3 | [19.7; 69.3] | 6.2% | 16.2 |
| CR | Care As Usual | - At Least 2 Flags Removed | 9 | 60.9% | 0.02 | [-0.08; 0.11] | 93.3% | 0 | [0; 64.8] | 100.0% | 340.36 |
| CR | Care As Usual | - At Least 1 Flag Removed | 2 | 91.3% | -0.01 | [-0.38; 0.35] | 103.3% | 0 | - | 100.0% | 407.32 |

| Intervention | Control | Analysis | $k$ | $\Delta_k$ | SMD | 95%-CI | $\Delta_{SMD}$ | $I^2$ | 95%-CI | $\Delta_{het}$ | NNT |
|---|---|---|---|---|---|---|---|---|---|---|---|
| CR | Care As Usual | - Flag 1 Removed (Effect) | 23 | 0.0% | 0.29 | [0.13; 0.46] | 3.3% | 50.3 | [19.7; 69.3] | 6.2% | 16.2 |
| CR | Care As Usual | - Flag 2 Removed (Power) | 8 | 65.2% | -0.01 | [-0.09; 0.08] | 103.3% | 0 | [0; 67.6] | 100.0% | 829.81 |
| CR | Care As Usual | - Flag 3 Removed (RoB) | 3 | 87.0% | 0.07 | [-0.36; 0.5] | 76.7% | 0 | [0; 89.6] | 100.0% | 78.97 |
| EMDR | Care As Usual | All Effect Sizes Included | 1 | 0.0% | 0.4 | [-0.26; 1.07] | 0.0% | | - | | 11.07 |
| EMDR | Care As Usual | - 3 Flags Removed | 1 | 0.0% | 0.4 | [-0.26; 1.07] | 0.0% | | - | | 11.07 |
| EMDR | Care As Usual | - Flag 1 Removed (Effect) | 1 | 0.0% | 0.4 | [-0.26; 1.07] | 0.0% | | - | | 11.07 |
| FPE | Care As Usual | All Effect Sizes Included | 2 | 0.0% | 0.38 | [0.1; 0.65] | 0.0% | 0 | - | | 12.04 |
| FPE | Care As Usual | - 3 Flags Removed | 2 | 0.0% | 0.38 | [0.1; 0.65] | 0.0% | 0 | - | | 12.04 |
| FPE | Care As Usual | - At Least 2 Flags Removed | 2 | 0.0% | 0.38 | [0.1; 0.65] | 0.0% | 0 | - | | 12.04 |
| FPE | Care As Usual | - Flag 1 Removed (Effect) | 2 | 0.0% | 0.38 | [0.1; 0.65] | 0.0% | 0 | - | | 12.04 |
| FPE | Care As Usual | - Flag 2 Removed (Power) | 2 | 0.0% | 0.38 | [0.1; 0.65] | 0.0% | 0 | - | | 12.04 |
| FSG | Care As Usual | All Effect Sizes Included | 2 | 0.0% | 0.03 | [-1.39; 1.46] | 0.0% | 0 | - | | 161.13 |
| FSG | Care As Usual | - 3 Flags Removed | 2 | 0.0% | 0.03 | [-1.39; 1.46] | 0.0% | 0 | - | | 161.13 |
| FSG | Care As Usual | - At Least 2 Flags Removed | 1 | 50.0% | 0.14 | [-0.33; 0.61] | -366.7% | | - | | 37.4 |
| FSG | Care As Usual | - Flag 1 Removed (Effect) | 2 | 0.0% | 0.03 | [-1.39; 1.46] | 0.0% | 0 | - | | 161.13 |
| FSG | Care As Usual | - Flag 2 Removed (Power) | 1 | 50.0% | 0.14 | [-0.33; 0.61] | -366.7% | | - | | 37.4 |
| FSIT | Care As Usual | All Effect Sizes Included | 1 | 0.0% | 0.72 | [0.19; 1.24] | 0.0% | | - | | 5.36 |
| FSIT | Care As Usual | - 3 Flags Removed | 1 | 0.0% | 0.72 | [0.19; 1.24] | 0.0% | | - | | 5.36 |
| FSIT | Care As Usual | - Flag 1 Removed (Effect) | 1 | 0.0% | 0.72 | [0.19; 1.24] | 0.0% | | - | | 5.36 |
| FT | Care As Usual | All Effect Sizes Included | 1 | 0.0% | 0.06 | [-0.43; 0.55] | 0.0% | | - | | 84.91 |
| FT | Care As Usual | - 3 Flags Removed | 1 | 0.0% | 0.06 | [-0.43; 0.55] | 0.0% | | - | | 84.91 |
| FT | Care As Usual | - At Least 2 Flags Removed | 1 | 0.0% | 0.06 | [-0.43; 0.55] | 0.0% | | - | | 84.91 |
| FT | Care As Usual | - Flag 1 Removed (Effect) | 1 | 0.0% | 0.06 | [-0.43; 0.55] | 0.0% | | - | | 84.91 |
| FT | Care As Usual | - Flag 3 Removed (RoB) | 1 | 0.0% | 0.06 | [-0.43; 0.55] | 0.0% | | - | | 84.91 |
| HIT | Care As Usual | All Effect Sizes Included | 1 | 0.0% | 0.44 | [0.03; 0.85] | 0.0% | | - | | 9.92 |
| HIT | Care As Usual | - 3 Flags Removed | 1 | 0.0% | 0.44 | [0.03; 0.85] | 0.0% | | - | | 9.92 |
| HIT | Care As Usual | - At Least 2 Flags Removed | 1 | 0.0% | 0.44 | [0.03; 0.85] | 0.0% | | - | | 9.92 |
| HIT | Care As Usual | - Flag 1 Removed (Effect) | 1 | 0.0% | 0.44 | [0.03; 0.85] | 0.0% | | - | | 9.92 |
| HIT | Care As Usual | - Flag 2 Removed (Power) | 1 | 0.0% | 0.44 | [0.03; 0.85] | 0.0% | | - | | 9.92 |
| MCT | Care As Usual | All Effect Sizes Included | 2 | 0.0% | 0.4 | [-0.95; 1.75] | 0.0% | 0 | - | | 11.35 |
| MCT | Care As Usual | - 3 Flags Removed | 2 | 0.0% | 0.4 | [-0.95; 1.75] | 0.0% | 0 | - | | 11.35 |
| MCT | Care As Usual | - Flag 1 Removed (Effect) | 2 | 0.0% | 0.4 | [-0.95; 1.75] | 0.0% | 0 | - | | 11.35 |
| MPE | Care As Usual | All Effect Sizes Included | 3 | 0.0% | 0.64 | [-0.06; 1.35] | 0.0% | 41.5 | [0; 82.2] | 0.0% | 6.21 |
| MPE | Care As Usual | - 3 Flags Removed | 3 | 0.0% | 0.64 | [-0.06; 1.35] | 0.0% | 41.5 | [0; 82.2] | 0.0% | 6.21 |
| MPE | Care As Usual | - At Least 2 Flags Removed | 3 | 0.0% | 0.64 | [-0.06; 1.35] | 0.0% | 41.5 | [0; 82.2] | 0.0% | 6.21 |
| MPE | Care As Usual | - At Least 1 Flag Removed | 2 | 33.3% | 0.78 | [-0.77; 2.33] | -21.9% | 0 | - | 100.0% | 4.8 |

| Intervention | Control | Analysis | *k* | $\Delta_k$ | SMD | 95%-CI | $\Delta_{SMD}$ | $I^2$ | 95%-CI | $\Delta_{het}$ | NNT |
|---|---|---|---|---|---|---|---|---|---|---|---|
| MPE | Care As Usual | - Flag 1 Removed (Effect) | 3 | 0.0% | 0.64 | [-0.06; 1.35] | 0.0% | 41.5 | [0; 82.2] | 0.0% | 6.21 |
| MPE | Care As Usual | - Flag 2 Removed (Power) | 3 | 0.0% | 0.64 | [-0.06; 1.35] | 0.0% | 41.5 | [0; 82.2] | 0.0% | 6.21 |
| MPE | Care As Usual | - Flag 3 Removed (RoB) | 2 | 33.3% | 0.78 | [-0.77; 2.33] | -21.9% | 0 | - | 100.0% | 4.8 |
| PE | Care As Usual | All Effect Sizes Included | 4 | 0.0% | 0.41 | [-0.03; 0.84] | 0.0% | 27.3 | [0; 72.7] | 0.0% | 11.05 |
| PE | Care As Usual | - 3 Flags Removed | 4 | 0.0% | 0.41 | [-0.03; 0.84] | 0.0% | 27.3 | [0; 72.7] | 0.0% | 11.05 |
| PE | Care As Usual | - At Least 2 Flags Removed | 3 | 25.0% | 0.31 | [0; 0.62] | 24.4% | 0 | [0; 89.6] | 100.0% | 15.3 |
| PE | Care As Usual | - At Least 1 Flag Removed | 2 | 50.0% | 0.37 | [-0.02; 0.76] | 9.8% | 0 | - | 100.0% | 12.37 |
| PE | Care As Usual | - Flag 1 Removed (Effect) | 4 | 0.0% | 0.41 | [-0.03; 0.84] | 0.0% | 27.3 | [0; 72.7] | 0.0% | 11.05 |
| PE | Care As Usual | - Flag 2 Removed (Power) | 3 | 25.0% | 0.31 | [0; 0.62] | 24.4% | 0 | [0; 89.6] | 100.0% | 15.3 |
| PE | Care As Usual | - Flag 3 Removed (RoB) | 2 | 50.0% | 0.37 | [-0.02; 0.76] | 9.8% | 0 | - | 100.0% | 12.37 |
| PMR | Care As Usual | All Effect Sizes Included | 1 | 0.0% | 0.27 | [-0.38; 0.92] | 0.0% | | - | | 17.96 |
| PMR | Care As Usual | - 3 Flags Removed | 1 | 0.0% | 0.27 | [-0.38; 0.92] | 0.0% | | - | | 17.96 |
| PMR | Care As Usual | - Flag 1 Removed (Effect) | 1 | 0.0% | 0.27 | [-0.38; 0.92] | 0.0% | | - | | 17.96 |
| PST | Care As Usual | All Effect Sizes Included | 1 | 0.0% | 0.09 | [-0.27; 0.45] | 0.0% | | - | | 57.85 |
| PST | Care As Usual | - 3 Flags Removed | 1 | 0.0% | 0.09 | [-0.27; 0.45] | 0.0% | | - | | 57.85 |
| PST | Care As Usual | - At Least 2 Flags Removed | 1 | 0.0% | 0.09 | [-0.27; 0.45] | 0.0% | | - | | 57.85 |
| PST | Care As Usual | - Flag 1 Removed (Effect) | 1 | 0.0% | 0.09 | [-0.27; 0.45] | 0.0% | | - | | 57.85 |
| PST | Care As Usual | - Flag 2 Removed (Power) | 1 | 0.0% | 0.09 | [-0.27; 0.45] | 0.0% | | - | | 57.85 |
| SST | Care As Usual | All Effect Sizes Included | 4 | 0.0% | 0.37 | [0; 0.74] | 0.0% | 40.6 | [0; 79.9] | 0.0% | 12.37 |
| SST | Care As Usual | - 3 Flags Removed | 4 | 0.0% | 0.37 | [0; 0.74] | 0.0% | 40.6 | [0; 79.9] | 0.0% | 12.37 |
| SST | Care As Usual | - At Least 2 Flags Removed | 3 | 25.0% | 0.4 | [-0.22; 1.02] | -8.1% | 56.3 | [0; 87.5] | -38.7% | 11.36 |
| SST | Care As Usual | - Flag 1 Removed (Effect) | 4 | 0.0% | 0.37 | [0; 0.74] | 0.0% | 40.6 | [0; 79.9] | 0.0% | 12.37 |
| SST | Care As Usual | - Flag 2 Removed (Power) | 3 | 25.0% | 0.4 | [-0.22; 1.02] | -8.1% | 56.3 | [0; 87.5] | -38.7% | 11.36 |
| SST-FPE | Care As Usual | All Effect Sizes Included | 4 | 0.0% | 0.49 | [-0.27; 1.24] | 0.0% | 90.7 | [79.3; 95.8] | 0.0% | 8.81 |
| SST-FPE | Care As Usual | - 3 Flags Removed | 4 | 0.0% | 0.49 | [-0.27; 1.24] | 0.0% | 90.7 | [79.3; 95.8] | 0.0% | 8.81 |
| SST-FPE | Care As Usual | - At Least 2 Flags Removed | 4 | 0.0% | 0.49 | [-0.27; 1.24] | 0.0% | 90.7 | [79.3; 95.8] | 0.0% | 8.81 |
| SST-FPE | Care As Usual | - Flag 1 Removed (Effect) | 4 | 0.0% | 0.49 | [-0.27; 1.24] | 0.0% | 90.7 | [79.3; 95.8] | 0.0% | 8.81 |
| SST-FPE | Care As Usual | - Flag 2 Removed (Power) | 4 | 0.0% | 0.49 | [-0.27; 1.24] | 0.0% | 90.7 | [79.3; 95.8] | 0.0% | 8.81 |
| WB | Care As Usual | All Effect Sizes Included | 1 | 0.0% | 0.45 | [0.02; 0.88] | 0.0% | | - | | 9.73 |
| WB | Care As Usual | - 3 Flags Removed | 1 | 0.0% | 0.45 | [0.02; 0.88] | 0.0% | | - | | 9.73 |
| WB | Care As Usual | - At Least 2 Flags Removed | 1 | 0.0% | 0.45 | [0.02; 0.88] | 0.0% | | - | | 9.73 |
| WB | Care As Usual | - Flag 1 Removed (Effect) | 1 | 0.0% | 0.45 | [0.02; 0.88] | 0.0% | | - | | 9.73 |
| WB | Care As Usual | - Flag 2 Removed (Power) | 1 | 0.0% | 0.45 | [0.02; 0.88] | 0.0% | | - | | 9.73 |
| ***Posttraumatic Stress Disorder*** | | | | | | | | | | | |
| EMDR | Care As Usual | All Effect Sizes Included | 3 | 0.0% | 0.67 | [-0.02; 1.36] | 0.0% | 0 | [0; 89.6] | | 5.85 |

| Intervention | Control | Analysis | *k* | $\Delta_k$ | SMD | 95%-CI | $\Delta_{SMD}$ | $I^2$ | 95%-CI | $\Delta_{het}$ | NNT |
|---|---|---|---|---|---|---|---|---|---|---|---|
| EMDR | Care As Usual | - 3 Flags Removed | 3 | 0.0% | 0.67 | [-0.02; 1.36] | 0.0% | 0 | [0; 89.6] | | 5.85 |
| EMDR | Care As Usual | - At Least 2 Flags Removed | 1 | 66.7% | 0.4 | [-0.39; 1.19] | 40.3% | | - | | 11.21 |
| EMDR | Care As Usual | - Flag 1 Removed (Effect) | 3 | 0.0% | 0.67 | [-0.02; 1.36] | 0.0% | 0 | [0; 89.6] | | 5.85 |
| EMDR | Care As Usual | - Flag 2 Removed (Power) | 1 | 66.7% | 0.4 | [-0.39; 1.19] | 40.3% | | - | | 11.21 |
| Mixed | Care As Usual | All Effect Sizes Included | 1 | 0.0% | 1.03 | [0.53; 1.54] | 0.0% | | - | | 3.32 |
| Mixed | Care As Usual | - 3 Flags Removed | 1 | 0.0% | 1.03 | [0.53; 1.54] | 0.0% | | - | | 3.32 |
| Mixed | Care As Usual | - At Least 2 Flags Removed | 1 | 0.0% | 1.03 | [0.53; 1.54] | 0.0% | | - | | 3.32 |
| Mixed | Care As Usual | - At Least 1 Flag Removed | 1 | 0.0% | 1.03 | [0.53; 1.54] | 0.0% | | - | | 3.32 |
| Mixed | Care As Usual | - Flag 1 Removed (Effect) | 1 | 0.0% | 1.03 | [0.53; 1.54] | 0.0% | | - | | 3.32 |
| Mixed | Care As Usual | - Flag 2 Removed (Power) | 1 | 0.0% | 1.03 | [0.53; 1.54] | 0.0% | | - | | 3.32 |
| Mixed | Care As Usual | - Flag 3 Removed (RoB) | 1 | 0.0% | 1.03 | [0.53; 1.54] | 0.0% | | - | | 3.32 |
| Non-trauma-focused | Care As Usual | All Effect Sizes Included | 8 | 0.0% | 0.44 | [0.17; 0.72] | 0.0% | 17.3 | [0; 60.2] | 0.0% | 9.87 |
| Non-trauma-focused | Care As Usual | - 3 Flags Removed | 8 | 0.0% | 0.44 | [0.17; 0.72] | 0.0% | 17.3 | [0; 60.2] | 0.0% | 9.87 |
| Non-trauma-focused | Care As Usual | - At Least 2 Flags Removed | 6 | 25.0% | 0.41 | [0.07; 0.74] | 6.8% | 32 | [0; 72.5] | -85.0% | 10.94 |
| Non-trauma-focused | Care As Usual | - At Least 1 Flag Removed | 3 | 62.5% | 0.41 | [-0.76; 1.57] | 6.8% | 68.5 | [0; 90.9] | -296.0% | 11.02 |
| Non-trauma-focused | Care As Usual | - Flag 1 Removed (Effect) | 8 | 0.0% | 0.44 | [0.17; 0.72] | 0.0% | 17.3 | [0; 60.2] | 0.0% | 9.87 |
| Non-trauma-focused | Care As Usual | - Flag 2 Removed (Power) | 6 | 25.0% | 0.41 | [0.07; 0.74] | 6.8% | 32 | [0; 72.5] | -85.0% | 10.94 |
| Non-trauma-focused | Care As Usual | - Flag 3 Removed (RoB) | 3 | 62.5% | 0.41 | [-0.76; 1.57] | 6.8% | 68.5 | [0; 90.9] | -296.0% | 11.02 |
| TF-CBT | Care As Usual | All Effect Sizes Included | 8 | 0.0% | 0.76 | [0.44; 1.08] | 0.0% | 1.4 | [0; 68] | 0.0% | 4.98 |
| TF-CBT | Care As Usual | - 3 Flags Removed | 8 | 0.0% | 0.76 | [0.44; 1.08] | 0.0% | 1.4 | [0; 68] | 0.0% | 4.98 |
| TF-CBT | Care As Usual | - At Least 2 Flags Removed | 4 | 50.0% | 0.72 | [0.11; 1.33] | 5.3% | 36 | [0; 77.8] | -2 471.4% | 5.33 |
| TF-CBT | Care As Usual | - At Least 1 Flag Removed | 2 | 75.0% | 0.64 | [-2.83; 4.1] | 15.8% | 43.4 | - | -3 000.0% | 6.29 |
| TF-CBT | Care As Usual | - Flag 1 Removed (Effect) | 8 | 0.0% | 0.76 | [0.44; 1.08] | 0.0% | 1.4 | [0; 68] | 0.0% | 4.98 |
| TF-CBT | Care As Usual | - Flag 2 Removed (Power) | 3 | 62.5% | 0.58 | [-0.01; 1.17] | 23.7% | 0 | [0; 89.6] | 100.0% | 7.07 |
| TF-CBT | Care As Usual | - Flag 3 Removed (RoB) | 3 | 62.5% | 0.84 | [-0.37; 2.05] | -10.5% | 57.2 | [0; 87.8] | -3 985.7% | 4.34 |
| EMDR | Waitlist | All Effect Sizes Included | 11 | 0.0% | 1.9 | [1.05; 2.74] | 0.0% | 88.9 | [82.2; 93.1] | 0.0% | 1.58 |
| EMDR | Waitlist | - 3 Flags Removed | 10 | 9.1% | 1.57 | [1; 2.13] | 17.4% | 79.6 | [63.2; 88.7] | 10.5% | 1.95 |
| EMDR | Waitlist | - At Least 2 Flags Removed | 6 | 45.5% | 1.47 | [0.65; 2.28] | 22.6% | 85.1 | [69.3; 92.7] | 4.3% | 2.11 |
| EMDR | Waitlist | - At Least 1 Flag Removed | 1 | 90.9% | 2.22 | [1.62; 2.82] | -16.8% | | - | | 1.38 |
| EMDR | Waitlist | - Flag 1 Removed (Effect) | 10 | 9.1% | 1.57 | [1; 2.13] | 17.4% | 79.6 | [63.2; 88.7] | 10.5% | 1.95 |
| EMDR | Waitlist | - Flag 2 Removed (Power) | 5 | 54.5% | 1.49 | [0.44; 2.54] | 21.6% | 88 | [74.6; 94.4] | 1.0% | 2.07 |
| EMDR | Waitlist | - Flag 3 Removed (RoB) | 2 | 81.8% | 1.89 | [-3.52; 7.31] | 0.5% | 50.1 | [0; 87.2] | 43.6% | 1.59 |
| Mixed | Waitlist | All Effect Sizes Included | 6 | 0.0% | 0.32 | [-0.35; 0.99] | 0.0% | 70.2 | [30.2; 87.3] | 0.0% | 14.76 |
| Mixed | Waitlist | - 3 Flags Removed | 6 | 0.0% | 0.32 | [-0.35; 0.99] | 0.0% | 70.2 | [30.2; 87.3] | 0.0% | 14.76 |
| Mixed | Waitlist | - At Least 2 Flags Removed | 5 | 16.7% | 0.41 | [-0.39; 1.21] | -28.1% | 74.2 | [35.9; 89.6] | -5.7% | 11 |
| Mixed | Waitlist | - At Least 1 Flag Removed | 2 | 66.7% | 0.82 | [-6.8; 8.44] | -156.2% | 86.3 | [45.7; 96.6] | -22.9% | 4.53 |

| Intervention | Control | Analysis | *k* | $\Delta_k$ | SMD | 95%-CI | $\Delta_{SMD}$ | $I^2$ | 95%-CI | $\Delta_{het}$ | NNT |
|---|---|---|---|---|---|---|---|---|---|---|---|
| Mixed | Waitlist | - Flag 1 Removed (Effect) | 6 | 0.0% | 0.32 | [-0.35; 0.99] | 0.0% | 70.2 | [30.2; 87.3] | 0.0% | 14.76 |
| Mixed | Waitlist | - Flag 2 Removed (Power) | 5 | 16.7% | 0.41 | [-0.39; 1.21] | -28.1% | 74.2 | [35.9; 89.6] | -5.7% | 11 |
| Mixed | Waitlist | - Flag 3 Removed (RoB) | 2 | 66.7% | 0.82 | [-6.8; 8.44] | -156.2% | 86.3 | [45.7; 96.6] | -22.9% | 4.53 |
| Non-trauma-focused | Waitlist | All Effect Sizes Included | 16 | 0.0% | 1.14 | [0.68; 1.59] | 0.0% | 79.8 | [67.9; 87.2] | 0.0% | 2.92 |
| Non-trauma-focused | Waitlist | - 3 Flags Removed | 15 | 6.2% | 1.02 | [0.64; 1.41] | 10.5% | 76.6 | [61.6; 85.8] | 4.0% | 3.36 |
| Non-trauma-focused | Waitlist | - At Least 2 Flags Removed | 11 | 31.2% | 1.02 | [0.52; 1.52] | 10.5% | 81.7 | [68.4; 89.4] | -2.4% | 3.36 |
| Non-trauma-focused | Waitlist | - At Least 1 Flag Removed | 1 | 93.8% | 1.57 | [0.79; 2.34] | -37.7% | | - | | 1.95 |
| Non-trauma-focused | Waitlist | - Flag 1 Removed (Effect) | 15 | 6.2% | 1.02 | [0.64; 1.41] | 10.5% | 76.6 | [61.6; 85.8] | 4.0% | 3.36 |
| Non-trauma-focused | Waitlist | - Flag 2 Removed (Power) | 10 | 37.5% | 0.95 | [0.43; 1.47] | 16.7% | 82.3 | [68.7; 90] | -3.1% | 3.69 |
| Non-trauma-focused | Waitlist | - Flag 3 Removed (RoB) | 2 | 87.5% | 1.7 | [-0.43; 3.83] | -49.1% | 0 | - | 100.0% | 1.78 |
| TF-CBT | Waitlist | All Effect Sizes Included | 48 | 0.0% | 1.19 | [0.95; 1.42] | 0.0% | 76.9 | [69.6; 82.4] | 0.0% | 2.76 |
| TF-CBT | Waitlist | - 3 Flags Removed | 45 | 6.2% | 1.05 | [0.87; 1.23] | 11.8% | 69 | [57.9; 77.1] | 10.3% | 3.24 |
| TF-CBT | Waitlist | - At Least 2 Flags Removed | 36 | 25.0% | 0.99 | [0.81; 1.17] | 16.8% | 68.9 | [56.2; 77.9] | 10.4% | 3.51 |
| TF-CBT | Waitlist | - At Least 1 Flag Removed | 6 | 87.5% | 1.15 | [0.63; 1.68] | 3.4% | 61.5 | [6; 84.2] | 20.0% | 2.86 |
| TF-CBT | Waitlist | - Flag 1 Removed (Effect) | 45 | 6.2% | 1.05 | [0.87; 1.23] | 11.8% | 69 | [57.9; 77.1] | 10.3% | 3.24 |
| TF-CBT | Waitlist | - Flag 2 Removed (Power) | 33 | 31.2% | 0.94 | [0.77; 1.11] | 21.0% | 67.5 | [53.4; 77.4] | 12.2% | 3.77 |
| TF-CBT | Waitlist | - Flag 3 Removed (RoB) | 9 | 81.2% | 1.36 | [0.89; 1.83] | -14.3% | 63 | [24; 82] | 18.1% | 2.32 |
| TF-Other | Waitlist | All Effect Sizes Included | 6 | 0.0% | 1.8 | [0.59; 3.01] | 0.0% | 76.1 | [46.3; 89.3] | 0.0% | 1.68 |
| TF-Other | Waitlist | - 3 Flags Removed | 5 | 16.7% | 1.46 | [0.36; 2.56] | 18.9% | 66.4 | [12.5; 87.1] | 12.7% | 2.12 |
| TF-Other | Waitlist | - At Least 2 Flags Removed | 3 | 50.0% | 1.61 | [0.23; 2.99] | 10.6% | 52.2 | [0; 86.2] | 31.4% | 1.89 |
| TF-Other | Waitlist | - At Least 1 Flag Removed | 1 | 83.3% | 2.13 | [1.56; 2.7] | -18.3% | | - | | 1.42 |
| TF-Other | Waitlist | - Flag 1 Removed (Effect) | 5 | 16.7% | 1.46 | [0.36; 2.56] | 18.9% | 66.4 | [12.5; 87.1] | 12.7% | 2.12 |
| TF-Other | Waitlist | - Flag 2 Removed (Power) | 1 | 83.3% | 2.13 | [1.56; 2.7] | -18.3% | | - | | 1.42 |
| TF-Other | Waitlist | - Flag 3 Removed (RoB) | 3 | 50.0% | 1.61 | [0.23; 2.99] | 10.6% | 52.2 | [0; 86.2] | 31.4% | 1.89 |
| ***Social Anxiety Disorder*** | | | | | | | | | | | |
| 3rd Wave | Care As Usual | All Effect Sizes Included | 1 | 0.0% | 0.84 | [0.13; 1.55] | 0.0% | | - | | 4.18 |
| 3rd Wave | Care As Usual | - 3 Flags Removed | 1 | 0.0% | 0.84 | [0.13; 1.55] | 0.0% | | - | | 4.18 |
| 3rd Wave | Care As Usual | - Flag 1 Removed (Effect) | 1 | 0.0% | 0.84 | [0.13; 1.55] | 0.0% | | - | | 4.18 |
| CBT | Care As Usual | All Effect Sizes Included | 4 | 0.0% | 0.65 | [-0.21; 1.51] | 0.0% | 76.8 | [36.6; 91.5] | 0.0% | 5.82 |
| CBT | Care As Usual | - 3 Flags Removed | 4 | 0.0% | 0.56 | [-0.09; 1.22] | 13.8% | 62.5 | [0; 87.4] | 18.6% | 6.96 |
| CBT | Care As Usual | - At Least 2 Flags Removed | 3 | 25.0% | 0.42 | [-0.24; 1.07] | 35.4% | 37.2 | [0; 80.2] | 51.6% | 10.11 |
| CBT | Care As Usual | - Flag 1 Removed (Effect) | 4 | 0.0% | 0.56 | [-0.09; 1.22] | 13.8% | 62.5 | [0; 87.4] | 18.6% | 6.96 |
| CBT | Care As Usual | - Flag 2 Removed (Power) | 3 | 25.0% | 0.42 | [-0.24; 1.07] | 35.4% | 37.2 | [0; 80.2] | 51.6% | 10.11 |
| 3rd Wave | Waitlist | All Effect Sizes Included | 6 | 0.0% | 1.08 | [0.44; 1.73] | 0.0% | 75.2 | [43.9; 89] | 0.0% | 3 |
| 3rd Wave | Waitlist | - 3 Flags Removed | 6 | 0.0% | 1.08 | [0.44; 1.73] | 0.0% | 75.2 | [43.9; 89] | 0.0% | 3 |

| Intervention | Control | Analysis | *k* | $\Delta_k$ | SMD | 95%-CI | $\Delta_{SMD}$ | $I^2$ | 95%-CI | $\Delta_{het}$ | NNT |
|---|---|---|---|---|---|---|---|---|---|---|---|
| 3rd Wave | Waitlist | - At Least 2 Flags Removed | 5 | 16.7% | 1.17 | [0.49; 1.85] | -8.3% | 74.2 | [36; 89.6] | 1.3% | 2.72 |
| 3rd Wave | Waitlist | - Flag 1 Removed (Effect) | 6 | 0.0% | 1.06 | [0.45; 1.68] | 1.9% | 73 | [37.9; 88.2] | 2.9% | 3.08 |
| 3rd Wave | Waitlist | - Flag 2 Removed (Power) | 4 | 33.3% | 1.06 | [0.23; 1.89] | 1.9% | 76.2 | [34.8; 91.3] | -1.3% | 3.09 |
| CBT | Waitlist | All Effect Sizes Included | 47 | 0.0% | 0.96 | [0.79; 1.13] | 0.0% | 71.2 | [61.5; 78.5] | 0.0% | 3.52 |
| CBT | Waitlist | - 3 Flags Removed | 45 | 4.3% | 0.87 | [0.76; 0.99] | 9.4% | 52.9 | [33.9; 66.5] | 25.7% | 3.97 |
| CBT | Waitlist | - At Least 2 Flags Removed | 27 | 42.6% | 0.78 | [0.7; 0.85] | 18.7% | 0 | [0; 42.5] | 100.0% | 4.63 |
| CBT | Waitlist | - Flag 1 Removed (Effect) | 43 | 8.5% | 0.8 | [0.72; 0.88] | 16.7% | 16.1 | [0; 42.8] | 77.4% | 4.44 |
| CBT | Waitlist | - Flag 2 Removed (Power) | 26 | 44.7% | 0.77 | [0.69; 0.85] | 19.8% | 0 | [0; 43.2] | 100.0% | 4.66 |
| Exposure | Waitlist | All Effect Sizes Included | 11 | 0.0% | 0.78 | [0.47; 1.08] | 0.0% | 61.4 | [25.5; 80] | 0.0% | 4.62 |
| Exposure | Waitlist | - 3 Flags Removed | 11 | 0.0% | 0.76 | [0.47; 1.05] | 2.6% | 55.6 | [12.7; 77.4] | 9.4% | 4.75 |
| Exposure | Waitlist | - At Least 2 Flags Removed | 3 | 72.7% | 0.9 | [-0.28; 2.07] | -15.4% | 71 | [1.4; 91.5] | -15.6% | 3.84 |
| Exposure | Waitlist | - Flag 1 Removed (Effect) | 11 | 0.0% | 0.76 | [0.47; 1.05] | 2.6% | 55.6 | [12.7; 77.4] | 9.4% | 4.75 |
| Exposure | Waitlist | - Flag 2 Removed (Power) | 3 | 72.7% | 0.9 | [-0.28; 2.07] | -15.4% | 71 | [1.4; 91.5] | -15.6% | 3.84 |
| Other | Waitlist | All Effect Sizes Included | 10 | 0.0% | 1.03 | [0.36; 1.71] | 0.0% | 85.2 | [74.6; 91.4] | 0.0% | 3.19 |
| Other | Waitlist | - 3 Flags Removed | 9 | 10.0% | 0.64 | [0.4; 0.88] | 37.9% | 25.2 | [0; 64.9] | 70.4% | 5.88 |
| Other | Waitlist | - At Least 2 Flags Removed | 3 | 70.0% | 0.48 | [0.38; 0.59] | 53.4% | 0 | [0; 89.6] | 100.0% | 8.41 |
| Other | Waitlist | - Flag 1 Removed (Effect) | 9 | 10.0% | 0.64 | [0.4; 0.88] | 37.9% | 25.2 | [0; 64.9] | 70.4% | 5.88 |
| Other | Waitlist | - Flag 2 Removed (Power) | 3 | 70.0% | 0.48 | [0.38; 0.59] | 53.4% | 0 | [0; 89.6] | 100.0% | 8.41 |
| ***Suicide*** | | | | | | | | | | | |
| ASSIP | Care As Usual | All Effect Sizes Included | 1 | 0.0% | 0.73 | [-0.06; 1.53] | 0.0% | | - | | 3.88 |
| ASSIP | Care As Usual | - 3 Flags Removed | 1 | 0.0% | 0.73 | [-0.06; 1.53] | 0.0% | | - | | 3.88 |
| ASSIP | Care As Usual | - Flag 1 Removed (Effect) | 1 | 0.0% | 0.73 | [-0.06; 1.53] | 0.0% | | - | | 3.88 |
| CAMS | Care As Usual | All Effect Sizes Included | 3 | 0.0% | 0.46 | [0.08; 0.85] | 0.0% | 0 | [0; 89.6] | | 6.53 |
| CAMS | Care As Usual | - 3 Flags Removed | 3 | 0.0% | 0.46 | [0.08; 0.85] | 0.0% | 0 | [0; 89.6] | | 6.53 |
| CAMS | Care As Usual | - At Least 2 Flags Removed | 3 | 0.0% | 0.46 | [0.08; 0.85] | 0.0% | 0 | [0; 89.6] | | 6.53 |
| CAMS | Care As Usual | - Flag 1 Removed (Effect) | 3 | 0.0% | 0.46 | [0.08; 0.85] | 0.0% | 0 | [0; 89.6] | | 6.53 |
| CAMS | Care As Usual | - Flag 2 Removed (Power) | 3 | 0.0% | 0.46 | [0.08; 0.85] | 0.0% | 0 | [0; 89.6] | | 6.53 |
| CBT | Care As Usual | All Effect Sizes Included | 20 | 0.0% | 0.42 | [0.11; 0.73] | 0.0% | 74.1 | [59.8; 83.3] | 0.0% | 7.3 |
| CBT | Care As Usual | - 3 Flags Removed | 18 | 10.0% | 0.3 | [0.07; 0.53] | 28.6% | 62.8 | [38.1; 77.6] | 15.2% | 10.57 |
| CBT | Care As Usual | - At Least 2 Flags Removed | 12 | 40.0% | 0.34 | [0.12; 0.57] | 19.0% | 59.6 | [23.8; 78.6] | 19.6% | 9.21 |
| CBT | Care As Usual | - Flag 1 Removed (Effect) | 18 | 10.0% | 0.3 | [0.07; 0.53] | 28.6% | 62.8 | [38.1; 77.6] | 15.2% | 10.57 |
| CBT | Care As Usual | - Flag 2 Removed (Power) | 12 | 40.0% | 0.34 | [0.12; 0.57] | 19.0% | 59.6 | [23.8; 78.6] | 19.6% | 9.21 |
| DBT | Care As Usual | All Effect Sizes Included | 3 | 0.0% | 0.29 | [-1.43; 2.01] | 0.0% | 72.8 | [8.3; 91.9] | 0.0% | 11.03 |
| DBT | Care As Usual | - 3 Flags Removed | 3 | 0.0% | 0.29 | [-1.43; 2.01] | 0.0% | 72.8 | [8.3; 91.9] | 0.0% | 11.03 |
| DBT | Care As Usual | - At Least 2 Flags Removed | 2 | 33.3% | -0.04 | [-1.8; 1.73] | 113.8% | 0 | - | 100.0% | 97.89 |

| Intervention | Control | Analysis | $k$ | $\Delta_k$ | SMD | 95%-CI | $\Delta_{SMD}$ | $I^2$ | 95%-CI | $\Delta_{het}$ | NNT |
|---|---|---|---|---|---|---|---|---|---|---|---|
| DBT | Care As Usual | - Flag 1 Removed (Effect) | 3 | 0.0% | 0.29 | [-1.43; 2.01] | 0.0% | 72.8 | [8.3; 91.9] | 0.0% | 11.03 |
| DBT | Care As Usual | - Flag 2 Removed (Power) | 2 | 33.3% | -0.04 | [-1.8; 1.73] | 113.8% | 0 | - | 100.0% | 97.89 |
| Psychodynamic | Care As Usual | All Effect Sizes Included | 3 | 0.0% | -0.02 | [-0.76; 0.72] | 0.0% | 45.8 | [0; 84] | 0.0% | 180.41 |
| Psychodynamic | Care As Usual | - 3 Flags Removed | 3 | 0.0% | -0.02 | [-0.76; 0.72] | 0.0% | 45.8 | [0; 84] | 0.0% | 180.41 |
| Psychodynamic | Care As Usual | - At Least 2 Flags Removed | 3 | 0.0% | -0.02 | [-0.76; 0.72] | 0.0% | 45.8 | [0; 84] | 0.0% | 180.41 |
| Psychodynamic | Care As Usual | - Flag 1 Removed (Effect) | 3 | 0.0% | -0.02 | [-0.76; 0.72] | 0.0% | 45.8 | [0; 84] | 0.0% | 180.41 |
| Psychodynamic | Care As Usual | - Flag 2 Removed (Power) | 3 | 0.0% | -0.02 | [-0.76; 0.72] | 0.0% | 45.8 | [0; 84] | 0.0% | 180.41 |
| Family | Care As Usual | All Effect Sizes Included | 1 | 0.0% | 1.15 | [0.37; 1.92] | 0.0% | | - | | 2.38 |
| Family | Care As Usual | - 3 Flags Removed | 1 | 0.0% | 1.15 | [0.37; 1.92] | 0.0% | | - | | 2.38 |
| Family | Care As Usual | - Flag 1 Removed (Effect) | 1 | 0.0% | 1.15 | [0.37; 1.92] | 0.0% | | - | | 2.38 |
| IMG | Care As Usual | All Effect Sizes Included | 1 | 0.0% | 3.19 | [2.48; 3.9] | 0.0% | | - | | 1.26 |
| MBCT | Care As Usual | All Effect Sizes Included | 1 | 0.0% | 0 | [-0.33; 0.33] | | | - | | |
| MBCT | Care As Usual | - 3 Flags Removed | 1 | 0.0% | 0 | [-0.33; 0.33] | | | - | | |
| MBCT | Care As Usual | - At Least 2 Flags Removed | 1 | 0.0% | 0 | [-0.33; 0.33] | | | - | | |
| MBCT | Care As Usual | - Flag 1 Removed (Effect) | 1 | 0.0% | 0 | [-0.33; 0.33] | | | - | | |
| MBCT | Care As Usual | - Flag 2 Removed (Power) | 1 | 0.0% | 0 | [-0.33; 0.33] | | | - | | |
| Mixed | Care As Usual | All Effect Sizes Included | 1 | 0.0% | -0.1 | [-0.45; 0.24] | 0.0% | | - | | 32.73 |
| Mixed | Care As Usual | - 3 Flags Removed | 1 | 0.0% | -0.1 | [-0.45; 0.24] | 0.0% | | - | | 32.73 |
| Mixed | Care As Usual | - At Least 2 Flags Removed | 1 | 0.0% | -0.1 | [-0.45; 0.24] | 0.0% | | - | | 32.73 |
| Mixed | Care As Usual | - Flag 1 Removed (Effect) | 1 | 0.0% | -0.1 | [-0.45; 0.24] | 0.0% | | - | | 32.73 |
| Mixed | Care As Usual | - Flag 2 Removed (Power) | 1 | 0.0% | -0.1 | [-0.45; 0.24] | 0.0% | | - | | 32.73 |
| Other | Care As Usual | All Effect Sizes Included | 13 | 0.0% | 0.14 | [-0.13; 0.4] | 0.0% | 78.7 | [64.2; 87.4] | 0.0% | 24.3 |
| Other | Care As Usual | - 3 Flags Removed | 13 | 0.0% | 0.14 | [-0.13; 0.4] | 0.0% | 78.7 | [64.2; 87.4] | 0.0% | 24.3 |
| Other | Care As Usual | - At Least 2 Flags Removed | 11 | 15.4% | 0.16 | [-0.15; 0.46] | -14.3% | 82.1 | [69.3; 89.6] | -4.3% | 21.3 |
| Other | Care As Usual | - Flag 1 Removed (Effect) | 13 | 0.0% | 0.14 | [-0.13; 0.4] | 0.0% | 78.7 | [64.2; 87.4] | 0.0% | 24.3 |
| Other | Care As Usual | - Flag 2 Removed (Power) | 11 | 15.4% | 0.16 | [-0.15; 0.46] | -14.3% | 82.1 | [69.3; 89.6] | -4.3% | 21.3 |
| PST | Care As Usual | All Effect Sizes Included | 4 | 0.0% | 0.43 | [0.29; 0.57] | 0.0% | 0 | [0; 84.7] | | 7.12 |
| PST | Care As Usual | - 3 Flags Removed | 4 | 0.0% | 0.43 | [0.29; 0.57] | 0.0% | 0 | [0; 84.7] | | 7.12 |
| PST | Care As Usual | - At Least 2 Flags Removed | 3 | 25.0% | 0.42 | [0.24; 0.61] | 2.3% | 0 | [0; 89.6] | | 7.25 |
| PST | Care As Usual | - Flag 1 Removed (Effect) | 4 | 0.0% | 0.43 | [0.29; 0.57] | 0.0% | 0 | [0; 84.7] | | 7.12 |
| PST | Care As Usual | - Flag 2 Removed (Power) | 3 | 25.0% | 0.42 | [0.24; 0.61] | 2.3% | 0 | [0; 89.6] | | 7.25 |
| SPI | Care As Usual | All Effect Sizes Included | 2 | 0.0% | 0.41 | [-0.98; 1.81] | 0.0% | 0 | - | | 7.42 |
| SPI | Care As Usual | - 3 Flags Removed | 2 | 0.0% | 0.41 | [-0.98; 1.81] | 0.0% | 0 | - | | 7.42 |
| SPI | Care As Usual | - At Least 2 Flags Removed | 2 | 0.0% | 0.41 | [-0.98; 1.81] | 0.0% | 0 | - | | 7.42 |
| SPI | Care As Usual | - Flag 1 Removed (Effect) | 2 | 0.0% | 0.41 | [-0.98; 1.81] | 0.0% | 0 | - | | 7.42 |
| SPI | Care As Usual | - Flag 2 Removed (Power) | 2 | 0.0% | 0.41 | [-0.98; 1.81] | 0.0% | 0 | - | | 7.42 |

| Intervention | Control | Analysis | *k* | $\Delta_k$ | SMD | 95%-CI | $\Delta_{SMD}$ | $I^2$ | 95%-CI | $\Delta_{het}$ | NNT |
|---|---|---|---|---|---|---|---|---|---|---|---|
| CBT | Waitlist | All Effect Sizes Included | 4 | 0.0% | 0.23 | [-0.32; 0.77] | 0.0% | 8.8 | [0; 86] | 0.0% | 14.4 |
| CBT | Waitlist | - 3 Flags Removed | 4 | 0.0% | 0.23 | [-0.32; 0.77] | 0.0% | 8.8 | [0; 86] | 0.0% | 14.4 |
| CBT | Waitlist | - At Least 2 Flags Removed | 1 | 75.0% | -0.05 | [-0.46; 0.36] | 121.7% | | - | | 69.32 |
| CBT | Waitlist | - Flag 1 Removed (Effect) | 4 | 0.0% | 0.23 | [-0.32; 0.77] | 0.0% | 8.8 | [0; 86] | 0.0% | 14.4 |
| CBT | Waitlist | - Flag 2 Removed (Power) | 1 | 75.0% | -0.05 | [-0.46; 0.36] | 121.7% | | - | | 69.32 |
| IMG | Waitlist | All Effect Sizes Included | 2 | 0.0% | 0.13 | [-5.89; 6.15] | 0.0% | 76.5 | [0; 94.7] | 0.0% | 25.96 |
| IMG | Waitlist | - 3 Flags Removed | 2 | 0.0% | 0.13 | [-5.89; 6.15] | 0.0% | 76.5 | [0; 94.7] | 0.0% | 25.96 |
| IMG | Waitlist | - At Least 2 Flags Removed | 1 | 50.0% | -0.31 | [-0.83; 0.21] | 338.5% | | - | | 10.34 |
| IMG | Waitlist | - Flag 1 Removed (Effect) | 2 | 0.0% | 0.13 | [-5.89; 6.15] | 0.0% | 76.5 | [0; 94.7] | 0.0% | 25.96 |
| IMG | Waitlist | - Flag 2 Removed (Power) | 1 | 50.0% | -0.31 | [-0.83; 0.21] | 338.5% | | - | | 10.34 |
| MBCT | Waitlist | All Effect Sizes Included | 1 | 0.0% | 0.57 | [-0.15; 1.29] | 0.0% | | - | | 5.16 |
| MBCT | Waitlist | - 3 Flags Removed | 1 | 0.0% | 0.57 | [-0.15; 1.29] | 0.0% | | - | | 5.16 |
| MBCT | Waitlist | - Flag 1 Removed (Effect) | 1 | 0.0% | 0.57 | [-0.15; 1.29] | 0.0% | | - | | 5.16 |
| Mixed | Waitlist | All Effect Sizes Included | 1 | 0.0% | 1.11 | [0.38; 1.83] | 0.0% | | - | | 2.47 |
| Mixed | Waitlist | - 3 Flags Removed | 1 | 0.0% | 1.11 | [0.38; 1.83] | 0.0% | | - | | 2.47 |
| Mixed | Waitlist | - Flag 1 Removed (Effect) | 1 | 0.0% | 1.11 | [0.38; 1.83] | 0.0% | | - | | 2.47 |
| Other | Waitlist | All Effect Sizes Included | 8 | 0.0% | 0.24 | [-0.07; 0.54] | 0.0% | 33.4 | [0; 70.5] | 0.0% | 13.73 |
| Other | Waitlist | - 3 Flags Removed | 8 | 0.0% | 0.24 | [-0.07; 0.54] | 0.0% | 33.4 | [0; 70.5] | 0.0% | 13.73 |
| Other | Waitlist | - At Least 2 Flags Removed | 5 | 37.5% | 0.16 | [-0.27; 0.59] | 33.3% | 38.6 | [0; 77.2] | -15.6% | 20.64 |
| Other | Waitlist | - Flag 1 Removed (Effect) | 8 | 0.0% | 0.24 | [-0.07; 0.54] | 0.0% | 33.4 | [0; 70.5] | 0.0% | 13.73 |
| Other | Waitlist | - Flag 2 Removed (Power) | 5 | 37.5% | 0.16 | [-0.27; 0.59] | 33.3% | 38.6 | [0; 77.2] | -15.6% | 20.64 |
| PST | Waitlist | All Effect Sizes Included | 1 | 0.0% | 0.21 | [-0.38; 0.8] | 0.0% | | - | | 15.47 |
| PST | Waitlist | - 3 Flags Removed | 1 | 0.0% | 0.21 | [-0.38; 0.8] | 0.0% | | - | | 15.47 |
| PST | Waitlist | - At Least 2 Flags Removed | 1 | 0.0% | 0.21 | [-0.38; 0.8] | 0.0% | | - | | 15.47 |
| PST | Waitlist | - Flag 1 Removed (Effect) | 1 | 0.0% | 0.21 | [-0.38; 0.8] | 0.0% | | - | | 15.47 |
| PST | Waitlist | - Flag 2 Removed (Power) | 1 | 0.0% | 0.21 | [-0.38; 0.8] | 0.0% | | - | | 15.47 |

*Note*. ALL = Protocol with 4 psychotherapies combined; AR = Applied relaxation; BAT = Behavioural activation; BT = Behaviour therapy; CAU = Care as usual; CBT = Cognitive behaviour therapy; CR = Cognitive restructuring; CRSS = Cognitive remediation focussed on social cognition; DBT = Dialectical behavior therapy; DYN = Psychodynamic therapy; EMDR = Eye movement desensitisation and reprocessing; FT = Family therapy; HIT = Hallucinations focused integrative therapy; IPT = Interpersonal psychotherapy; LRT = Life review therapy; MBT = Mentalisation based therapy; MCT = Metacognitive therapy; MPE = Mindfulness-based psychoeducation; Mixed = Mixed approaches/therapeutic techniques; Other ctr = Other type of inactive control group; Other psy = Other type of psychotherapy; PE = Psychoeducation; PDP = Psychodynamic psychotherapy; PHA = Pharmacological treatment; PST = Problem-solving therapy; SST = Social skills training; ST = Schema therapy; SUP = Supportive counseling; TFP = Transferrence-focused therapy; WB = Wellbeing; WL = Waitlist; 3rd = Third wave therapies.

### S8. Change of effect size and heterogeneity when excluding flagged studies.

| | Effect ↓<br>Heterogeneity ↓ | Effect ↓<br>Heterogeneity ↑ | Effect ↑<br>Heterogeneity ↓ | Effect ↑<br>Heterogeneity ↑ |
|---|---|---|---|---|
| - 3 Flags Removed | 39 (100.0%) | 0 (0.0%) | 0 (0.0%) | 0 (0.0%) |
| - At Least 2 Flags Removed | 36 (60.0%) | 12 (20.0%) | 1 (1.7%) | 11 (18.3%) |
| - At Least 1 Flag Removed | 26 (70.3%) | 5 (13.5%) | 3 (8.1%) | 3 (8.1%) |
| - Flag 1 Removed (Effect) | 43 (97.7%) | 0 (0.0%) | 1 (2.3%) | 0 (0.0%) |
| - Flag 2 Removed (Power) | 29 (51.8%) | 14 (25.0%) | 2 (3.6%) | 11 (19.6%) |
| - Flag 3 Removed (RoB) | 21 (58.3%) | 8 (22.2%) | 5 (13.9%) | 2 (5.6%) |

*Note*. Based on subgroup-specific reanalyses of Harrer et al. (2025) across indications (see S7 above). Only displays reanalysis leading to changed in both the pooled effect and heterogeneity.

### S9. Characteristics of flagged studies.

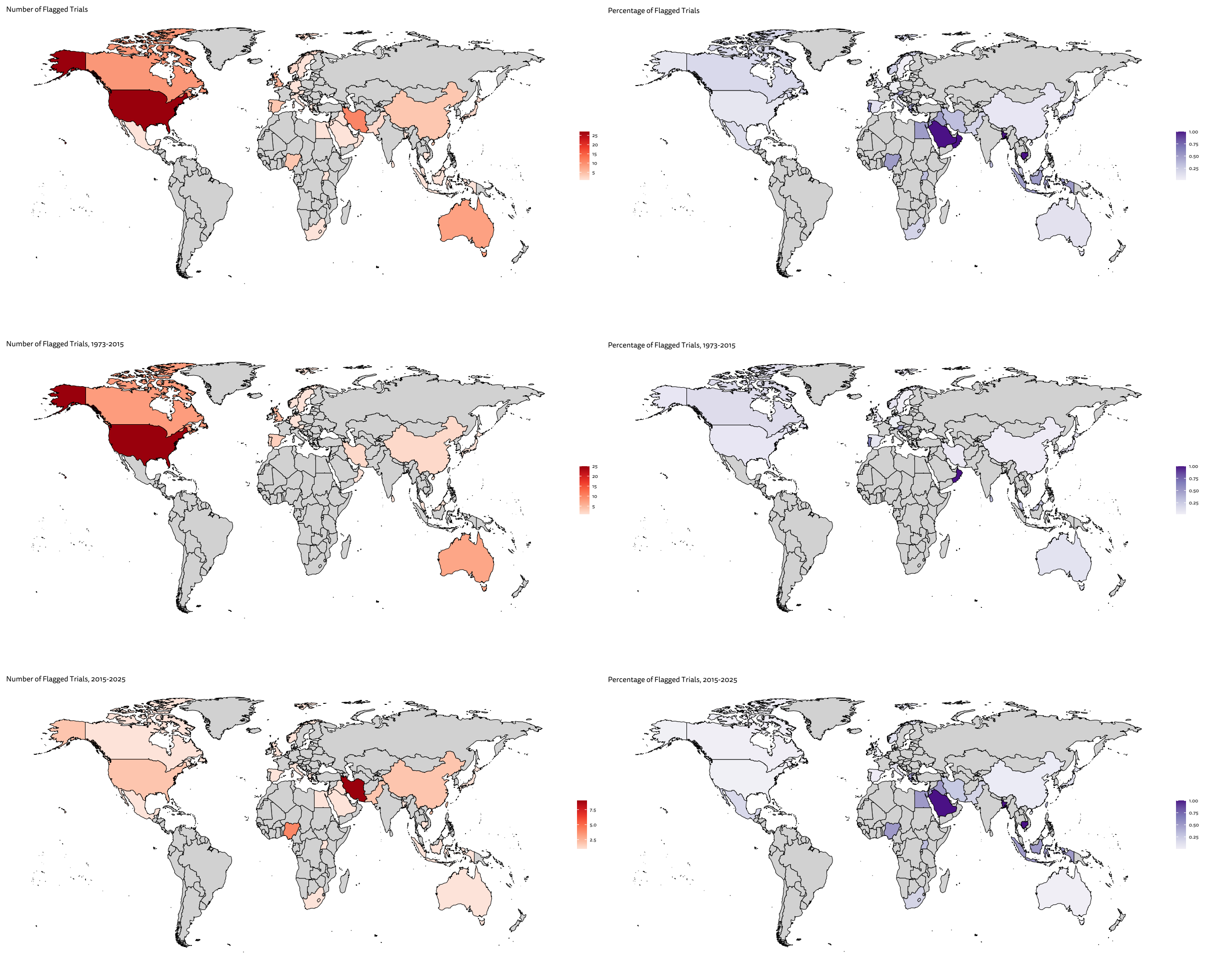

Number of flagged studies by country

| Country | Total | % (Flagged) | % (Country) |
|---|---|---|---|
| Australia | 8 | 7.84 | 11.94 |
| Austria | 1 | 0.98 | 50.00 |
| Bangladesh | 1 | 0.98 | 100.00 |
| Cambodia | 1 | 0.98 | 100.00 |
| Canada | 9 | 8.82 | 17.31 |

Number of flagged studies by country (since 2015)

| Country | Total | % (Flagged) | % (Country) |
|---|---|---|---|
| Australia | 1 | 2.56 | 1.49 |
| Bangladesh | 1 | 2.56 | 100.00 |
| Cambodia | 1 | 2.56 | 100.00 |
| Canada | 1 | 2.56 | 1.92 |
| China | 2 | 5.13 | 3.70 |

| | | | |
|---|---|---|---|
| China | 4 | 3.92 | 7.41 |
| Egypt | 1 | 0.98 | 50.00 |
| Germany | 1 | 0.98 | 1.72 |
| Greece | 1 | 0.98 | 50.00 |
| Indonesia | 1 | 0.98 | 50.00 |
| Iran | 11 | 10.78 | 32.35 |
| Iraq | 1 | 0.98 | 50.00 |
| Italy | 1 | 0.98 | 11.11 |
| Japan | 2 | 1.96 | 20.00 |
| Jordan | 1 | 0.98 | 50.00 |
| Malaysia | 1 | 0.98 | 25.00 |
| Mexico | 1 | 0.98 | 16.67 |
| Netherlands | 1 | 0.98 | 1.52 |
| Nigeria | 4 | 3.92 | 50.00 |
| Norway | 2 | 1.96 | 20.00 |
| Oman | 1 | 0.98 | 100.00 |
| Pakistan | 2 | 1.96 | 20.00 |
| Portugal | 2 | 1.96 | 66.67 |
| Saudi Arabia | 1 | 0.98 | 100.00 |
| South Africa | 1 | 0.98 | 20.00 |
| Spain | 4 | 3.92 | 11.76 |
| Sri Lanka | 1 | 0.98 | 33.33 |
| Sweden | 1 | 0.98 | 2.33 |
| Taiwan | 2 | 1.96 | 33.33 |
| Uganda | 1 | 0.98 | 33.33 |
| UK | 6 | 5.88 | 5.41 |
| USA | 27 | 26.47 | 8.13 |

| | | | |
|---|---|---|---|
| Egypt | 1 | 2.56 | 50.00 |
| Greece | 1 | 2.56 | 50.00 |
| Indonesia | 1 | 2.56 | 50.00 |
| Iran | 9 | 23.08 | 26.47 |
| Iraq | 1 | 2.56 | 50.00 |
| Italy | 1 | 2.56 | 11.11 |
| Japan | 1 | 2.56 | 10.00 |
| Jordan | 1 | 2.56 | 50.00 |
| Mexico | 1 | 2.56 | 16.67 |
| Nigeria | 4 | 10.26 | 50.00 |
| Norway | 1 | 2.56 | 10.00 |
| Pakistan | 2 | 5.13 | 20.00 |
| Saudi Arabia | 1 | 2.56 | 100.00 |
| South Africa | 1 | 2.56 | 20.00 |
| Spain | 1 | 2.56 | 2.94 |
| Taiwan | 2 | 5.13 | 33.33 |
| Uganda | 1 | 2.56 | 33.33 |
| UK | 1 | 2.56 | 0.90 |
| USA | 2 | 5.13 | 0.60 |

Studies by World Bank income region

| | Total | % |
|---|---|---|
| High Income | 70 | 68.63 |
| Low Income | 1 | 0.98 |
| Lower-Middle Income | 18 | 17.65 |
| Upper-Middle Income | 13 | 12.75 |

Studies by world bank income region (since 2015)

| | Total | % |
|---|---|---|
| High Income | 13 | 33.33 |
| Low Income | 1 | 2.56 |
| Lower-Middle Income | 16 | 41.03 |
| Upper-Middle Income | 9 | 23.08 |

### **S10**. Number of newly published trial by flagging status.

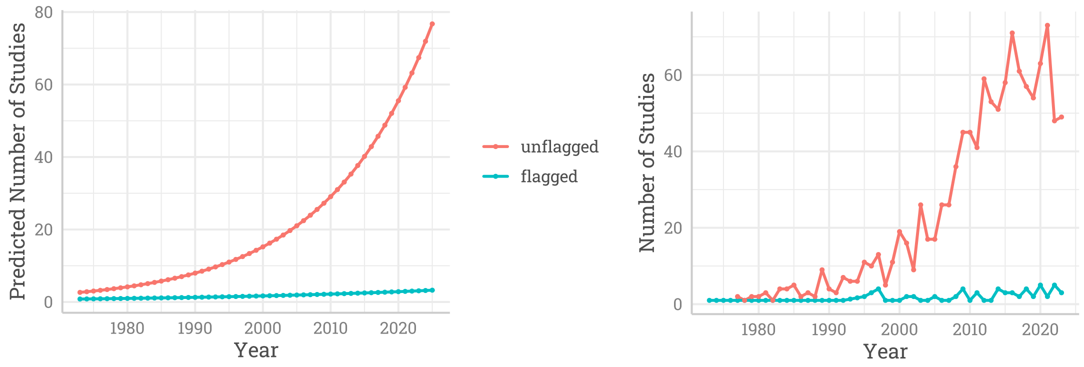

| | **Estimate** | **S.E.** | **z value** | **Pr(>\|z\|)** | **Signif.** |
|---|---|---|---|---|---|
| (Intercept) | 0.5903 | 0.1399 | 4.219 | <0.0001 | *** |
| Publication Year | 0.3704 | 0.1489 | 2.487 | 0.01288 | * |
| Not Flagged | 2.3178 | 0.1449 | 15.993 | <0.0001 | *** |
| Publication Year × Not Flagged | 0.5310 | 0.1532 | 3.466 | 0.00052 | *** |

Signif. codes: 0 ‘***’ 0.001 ‘**’ 0.01 ‘*’ 0.05 ‘.’ 0.1 ‘ ’ 1
